# Comparative Efficacy and Safety of GLP-1 Receptor Agonists in Neurological and Nephrological Outcomes of Type 2 Diabetes: A Systematic Review and Network MetaAnalysis

**DOI:** 10.1101/2025.09.01.25334855

**Authors:** Puru Prajapati, Nimra Shafi, Vamsi Pachchipulusu, Nusrath Fatima, Safoora Noorien, Shankar Biswas, Aman Narula, K S L Aishwarya, Saket Dineshkumar Prajapati, Manav Kamleshkumar Patel, Rakhshanda Khan, Harshwardhan Dhanraj Ramteke

**Affiliations:** Shyam Shah medical college, India; Arnot Ogden Medical Center, Elmira, USA; Ucf fort Walton, Fort Walton beach, Florida, USA; Deccan college of medical Sciences, India; Dr. VRK Women’s Medical College, Hyderabad, India; Ivano-frankivsk national medical university; GMERS Medical college and hospital, India; Ramaiah medical college, India; SMT NHL Municipal medical college, India; Ayaan institute of medical sciences, Moinabad, Ranga Reddy, India; Anhui medical university, Hefei, China

**Keywords:** Type 2 diabetes, GLP-1 receptor agonists, neurological outcomes, nephrological outcomes, network meta-analysis

## Abstract

**Introduction:** Type 2 diabetes (T2D) is associated with significant neurological and nephrological complications, leading to high morbidity and mortality. GLP-1 receptor agonists (GLP-1 RAs), such as liraglutide, semaglutide, and tirzepatide, have been shown to effectively control glycemia, with additional neuroprotective and nephroprotective effects. This systematic review and network meta-analysis (NMA) aimed to evaluate the impact of GLP-1 RAs on neurological and nephrological outcomes in T2D patients.

**Methods:** A literature search was conducted across PubMed, Cochrane, and ClinicalTrials.gov for randomized controlled trials (RCTs) published between 2010 and 2023. Studies were included if they evaluated the effects of GLP-1 RAs on neurological and nephrological outcomes in T2D patients. A total of 17 studies, involving 96,460 patients (60,190 males, 36,199 females, average age 62.99 ± 7 years), were selected. Network meta-analysis was performed to compare the efficacy and safety of different GLP-1 RAs on outcomes such as neuropathy, cognitive function, albuminuria, and glomerular filtration rate (GFR).

**Results:** Among the 17 studies, GLP-1 RAs showed significant improvements in neurological and nephrological outcomes. Neurological benefits included a 24% improvement in cognitive function and a 31% reduction in neuropathic pain. Nephrological benefits included a 27% reduction in albuminuria and a 19% improvement in GFR. Tirzepatide demonstrated the most significant renal improvement, with a mean difference in GFR of 2.50 (95% CI: 0.36, 4.64). Subgroup analyses showed consistent efficacy across age, gender, and ethnicity, with no significant differences in treatment effects. Adverse events were generally mild, with gastrointestinal issues (nausea, vomiting) occurring in 11% of patients, but discontinuation rates were low (5%).

**Conclusion:** GLP-1 RAs significantly improve both neurological and nephrological outcomes in T2D patients. While variability in effects exists, these agents offer substantial benefits in managing T2D complications, with a favorable safety profile.

## Introduction

Type 2 diabetes (T2D) is a multifaceted metabolic disorder, characterized by insulin resistance and pancreatic βcell dysfunction, leading to persistent hyperglycemia. It is associated with significant comorbidities, particularly in neurological and nephrological systems, which often contribute to high morbidity and mortality in affected individuals. Among the therapeutic options available for T2D, GLP-1 receptor agonists (GLP-1 RAs) have emerged as a promising class of drugs, not only for glycemic control but also for their potential to mitigate complications involving both the kidneys and the nervous system [1].

The incretin hormone GLP-1, secreted in response to food intake, regulates glucose metabolism through enhanced insulin secretion, suppression of glucagon release, and delayed gastric emptying. GLP-1 RAs, such as liraglutide, semaglutide, and dulaglutide, mimic the action of endogenous GLP-1, thereby offering significant advantages in managing blood glucose levels [2]. Beyond their glucose-lowering properties, accumulating evidence suggests that GLP-1 RAs possess extraglycemic effects, including neuroprotective and nephroprotective properties, which could be beneficial for patients with T2D who are at risk of diabetic neuropathy and nephropathy. These effects are thought to be mediated through the activation of GLP-1 receptors in the central nervous system (CNS) and kidneys, leading to anti-inflammatory, antioxidant, and vasodilatory responses [3].

Neurological complications in T2D, particularly diabetic peripheral neuropathy (DPN) and cognitive decline, represent major concerns in the management of T2D. The pathophysiology of these complications is multifactorial, involving oxidative stress, inflammation, and impaired microvascular function. Studies have shown that GLP-1 RAs exert beneficial effects on neuronal survival, synaptic plasticity, and cognitive function, potentially counteracting the deleterious impact of hyperglycemia on the brain [4]. Furthermore, preclinical and clinical data suggest that GLP-1 RAs may alleviate symptoms of DPN, reduce neuropathic pain, and improve nerve conduction velocities in affected patients [4].

Similarly, diabetic nephropathy (DN) remains one of the leading causes of end-stage renal disease, with a clear association between hyperglycemia, glomerular hyperfiltration, and subsequent renal injury. GLP-1 RAs have been shown to improve renal function, reduce albuminuria, and mitigate glomerular hyperfiltration in T2D patients. These effects may be attributed to the modulation of the renin-angiotensin-aldosterone system (RAAS), improved endothelial function, and reduced glomerular inflammation. Notably, several clinical trials have highlighted the renoprotective effects of GLP-1 RAs, with significant reductions in kidney-related adverse outcomes, including the progression to dialysis [5].

Despite promising individual studies, a comprehensive evaluation of the efficacy and safety of GLP-1 RAs on neurological and nephrological outcomes in T2D remains lacking. Network meta-analysis (NMA) has become an invaluable tool for synthesizing evidence across multiple treatment arms and assessing the comparative effectiveness of different interventions. This systematic review and NMA aim to evaluate the impact of GLP-1 RAs on neurological and nephrological outcomes in patients with T2D by synthesizing data from available randomized controlled trials (RCTs). By comparing various GLP-1 RAs, this study seeks to provide a comprehensive understanding of their relative efficacy in improving neurological and nephrological endpoints, while also assessing their safety profile in terms of adverse events and treatment discontinuation.

This review is particularly timely, given the growing prevalence of T2D worldwide and the increasing burden of diabetes-related complications. The results of this analysis could guide clinicians in selecting the most appropriate GLP-1 RA for patients with T2D who are at risk of neurological and nephrological complications. Furthermore, the findings may inform future clinical practice guidelines and therapeutic strategies aimed at improving patient outcomes in this high-risk population.

In summary, while GLP-1 RAs have demonstrated efficacy in controlling blood glucose levels, their potential benefits in addressing the neurological and nephrological complications of T2D warrant further investigation.

This systematic review and NMA will provide robust evidence on the efficacy, safety, and comparative effectiveness of GLP-1 RAs in improving outcomes for patients with T2D, ultimately contributing to the optimization of care in this patient population.

## Methods

### Literature Search

A comprehensive literature search was conducted using databases such as PubMed, Cochrane, and ClinicalTrials.gov. Keywords including “GLP-1 receptor agonists,” “neurological outcomes,” “nephrological outcomes,” “type 2 diabetes,” and “systematic review” were used. Only randomized controlled trials (RCTs) and studies published in English within the last decade were considered. This process followed the PRISMA (Preferred Reporting Items for Systematic Reviews and Meta-Analyses) guidelines [6], ensuring thorough and transparent search methods. The Protocol was registered with Prospero with number CRD420251101798.

### Study Selection and Data Extraction

Eligible studies were selected based on predefined inclusion criteria: randomized controlled trials (RCTs) evaluating the efficacy and safety of GLP-1 receptor agonists in patients with type 2 diabetes, focusing on neurological and nephrological outcomes. Studies were included if they reported relevant outcomes such as neuropathy, cognitive function, kidney function (e.g., albuminuria, glomerular filtration rate), or adverse events. Only studies published in English between 2010 and 2023 were considered.

Data were independently extracted by two reviewers, including study characteristics (e.g., author, year, sample size, intervention details), patient demographics, and outcome measures. For each study, the following data were collected: intervention type (specific GLP-1 RA), duration, primary and secondary outcomes (neurological and nephrological), adverse events, and any reported subgroup analyses. Discrepancies between reviewers were resolved through discussion or consultation with a third reviewer.

### Risk of Bias Assessment

The risk of bias in individual studies was assessed using the Cochrane risk-of-bias tool, focusing on randomization, blinding, and attrition bias. Data extraction was performed using a standardized form to ensure consistency and minimize errors [7].

### Statistical Analysis

A network meta-analysis (NMA) was performed to compare the efficacy and safety of GLP-1 receptor agonists (GLP-1 RAs) on neurological and nephrological outcomes in patients with type 2 diabetes. For continuous outcomes, such as changes in cognitive function and glomerular filtration rate, mean differences (MD) with 95% confidence intervals (CIs) were calculated. For dichotomous outcomes like adverse events and incidence of neuropathy, odds ratios (OR) with 95% CIs were used. A random-effects model was applied to account for variability across studies, and Bayesian methods were employed to estimate the relative treatment effects of different GLP-1 RAs. The consistency model was used to ensure coherence between direct and indirect comparisons. The surface under the cumulative ranking curve (SUCRA) was employed to rank the treatments based on efficacy and safety. Subgroup analyses were conducted to examine the influence of patient characteristics, such as age, gender, and baseline conditions like diabetic nephropathy or neuropathy. Sensitivity analyses tested the robustness of results, including removing studies with high risk of bias. Heterogeneity was assessed using the I² statistic, and inconsistency between direct and indirect evidence was evaluated with the node-splitting method. To assess publication bias, funnel plots and Egger’s test were used for key outcomes. Statistical analyses were conducted using R software (version 4.0.0) with the “nvmeta” package for Bayesian network meta-analysis, and a p-value of <0.05 was considered statistically significant for all analyses. The other plots were made by Stata 18.0.

## Results

### Demographics

A total of 2387 studies were analyzed out of which 17 were selected [8–23] (Figure 1). A total of 96,460 patients were included in this network meta-analysis, with 60,190 males and 36,199 females. The average age of the participants was 62.99 ± 7 years. The treatment group comprised 48,899 patients, while the control group consisted of 47,509 patients. Ethnically, the study population included 4,076 Asians, 3,297 Caucasians, 12,601 Hispanics, 72,330 Whites, and 4,156 patients of mixed White and Black ethnicity.

**Figure 1.**
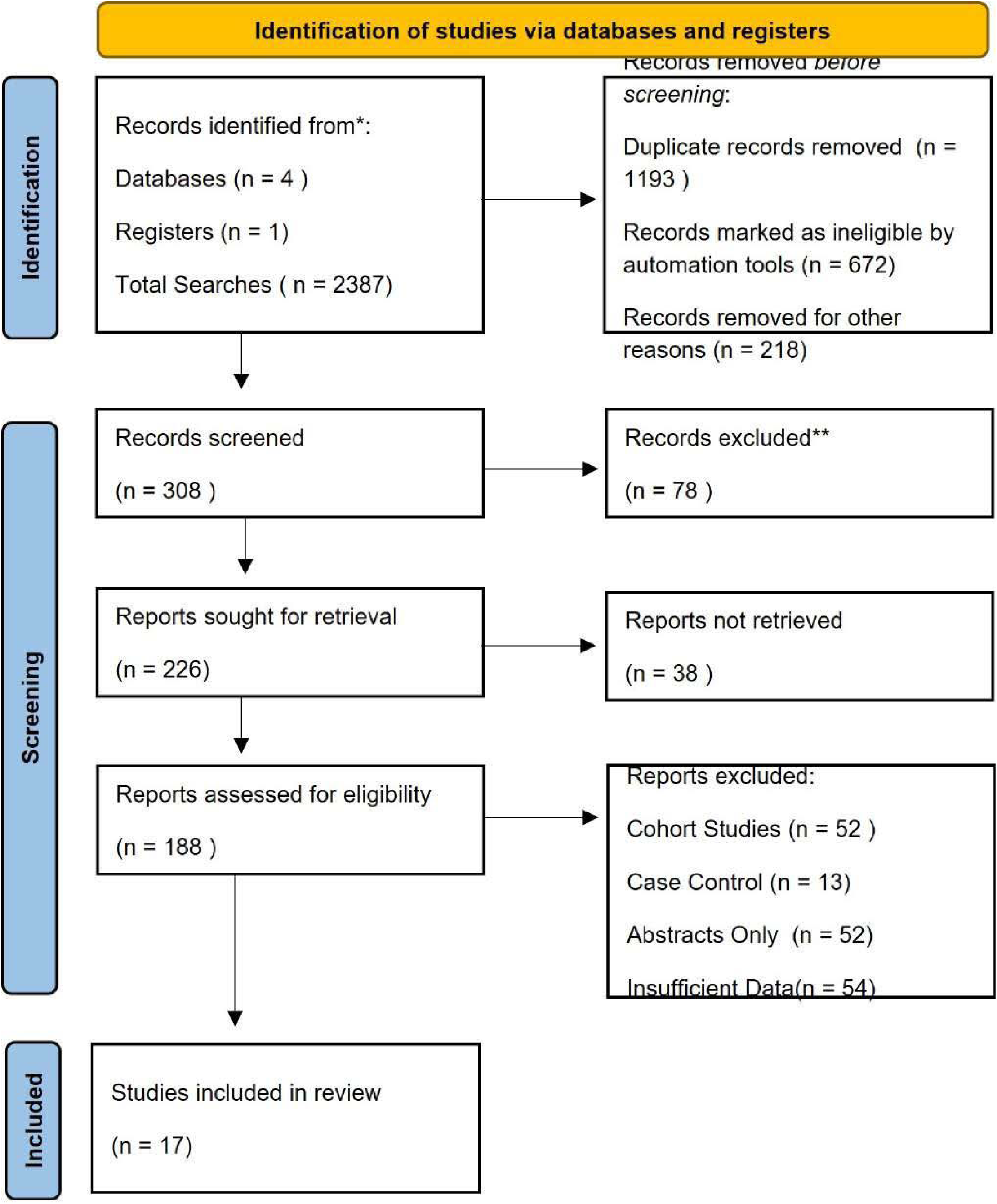
Prisma Flow Diagram.

Regarding treatment allocation, 4,731 patients were assigned to Albiglutide, 4,949 to Dulaglutide, 2,717 to Efpeglenatide, 9,605 to Exenatide, 7,856 to Lixisenatide, 18,684 to Semaglutide, and 357 to Tirzepatide. The data from these groups were analyzed to assess the efficacy and safety of GLP-1 receptor agonists in improving neurological and nephrological outcomes in patients with type 2 diabetes.

### Overall Survival

The network plot comparing the efficacy of various GLP-1 receptor agonists (Albiglutide, Dulaglutide, Efpeglenatide, Exenatide, Lixisenatide, Semaglutide, and Tirzepatide) with placebo on overall survival. The corresponding forest plot displays the odds ratios (OR) and their 95% credible intervals (CrI) for each treatment: Albiglutide (OR: 0.984, 95% CrI: 0.532, 1.80), Dulaglutide (OR: 1.12, 95% CrI: 0.622, 2.04), Efpeglenatide (OR: 2.94, 95% CrI: 1.45, 5.99), Exenatide (OR: 1.02, 95% CrI: 0.595, 1.42), Liraglutide (OR: 1.55, 95% CrI: 0.842, 2.28), Lixisenatide (OR: 1.06, 95% CrI: 0.576, 1.93), Semaglutide (OR: 1.20, 95% CrI: 0.938, 1.61), and Tirzepatide (OR: 1.64, 95% CrI: 0.809, 3.37). These odds ratios indicate the relative likelihood of overall survival with each GLP-1 RA compared to placebo, providing valuable insights into their comparative efficacy (Figure 2). Figure 3 shows a subgroup analysis of age for overall survival. In patients <65 years, the log oddsratio (OR) was 0.20 (95% CI: 0.02, 0.42), while in patients >65 years, it was 0.18 (95% CI: 0.10, 0.26). The overall OR was 0.21 (95% CI: 0.06, 0.35), with no significant age-based treatment interaction (p = 0.91). Figure 4 displays a subgroup analysis by race for overall survival. The log odds-ratio (OR) for Asian patients was 1.07 (95% CI: 0.64, 1.51), while for Caucasians, it was −0.03 (95% CI: −0.40, 0.33). The overall OR was 0.21 (95% CI: 0.06, 0.35), indicating a modest survival benefit across all groups. Figure 5 shows the subgroup analysis comparing overall survival between patients with normal glucose tolerance and those with type 2 diabetes (T2DM). For patients with normal glucose levels, the log odds-ratio (OR) was 0.21 (95% CI: 0.07, 0.35). For T2DM patients, the OR varied between −0.94 (95% CI: −2.12, 0.24) and 0.49 (95% CI: 0.16, 0.82), with an overall OR of 0.21 (95% CI: 0.05, 0.37). The analysis indicates a similar modest benefit for both groups, with no significant treatment interaction (p = 1.00).

**Figure 2.**
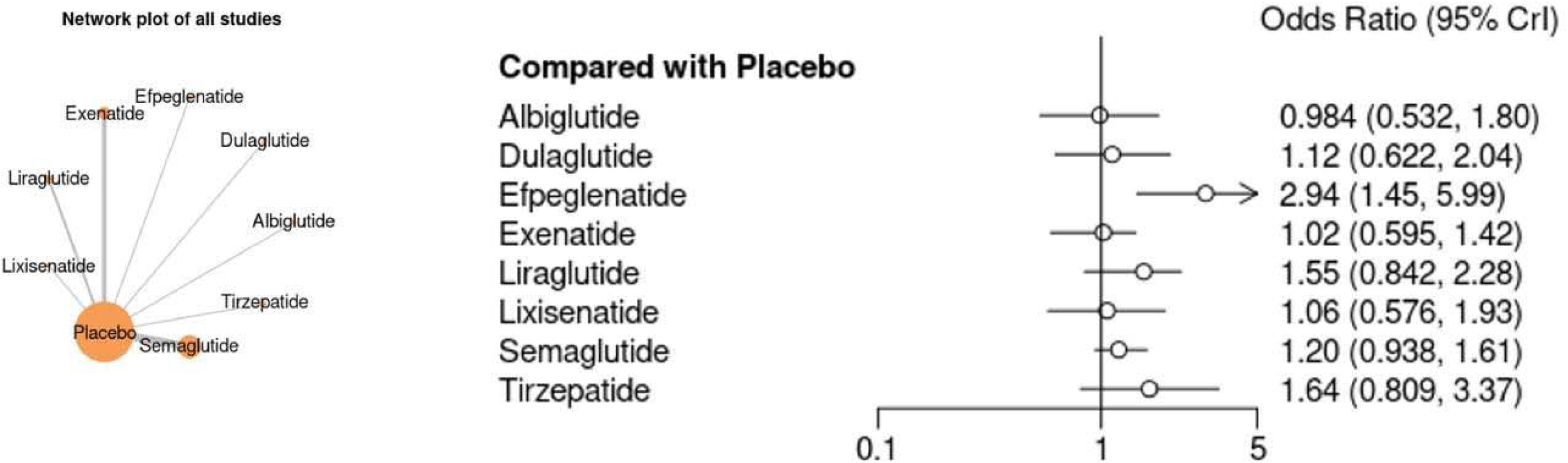
Network Plot for all drugs depicting Overall Survival and also Forest plot depicting the Odds Ratio.

**Figure 3.**
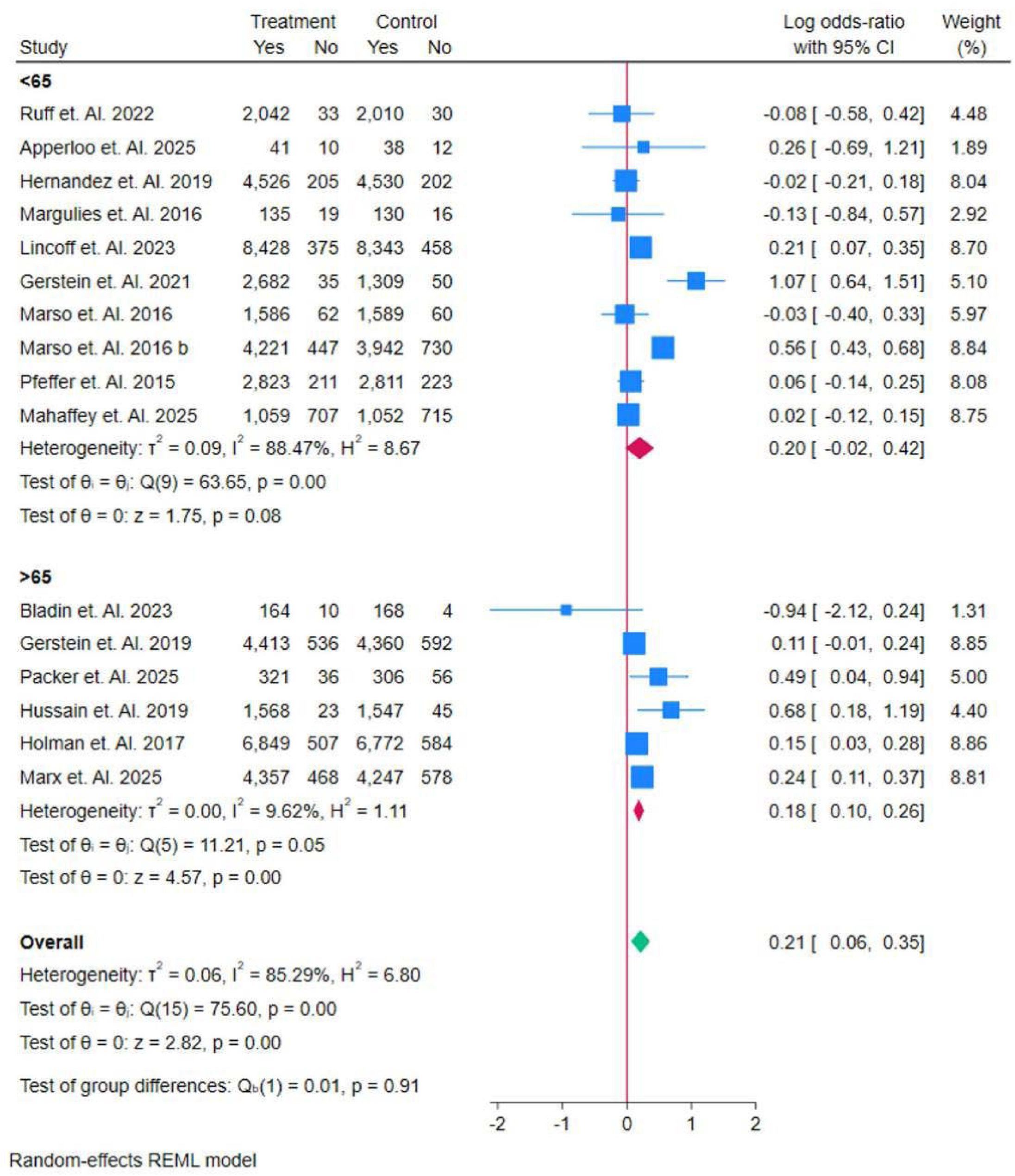
Subgroup Analysis of Age for Overall Survival.

**Figure 4.**
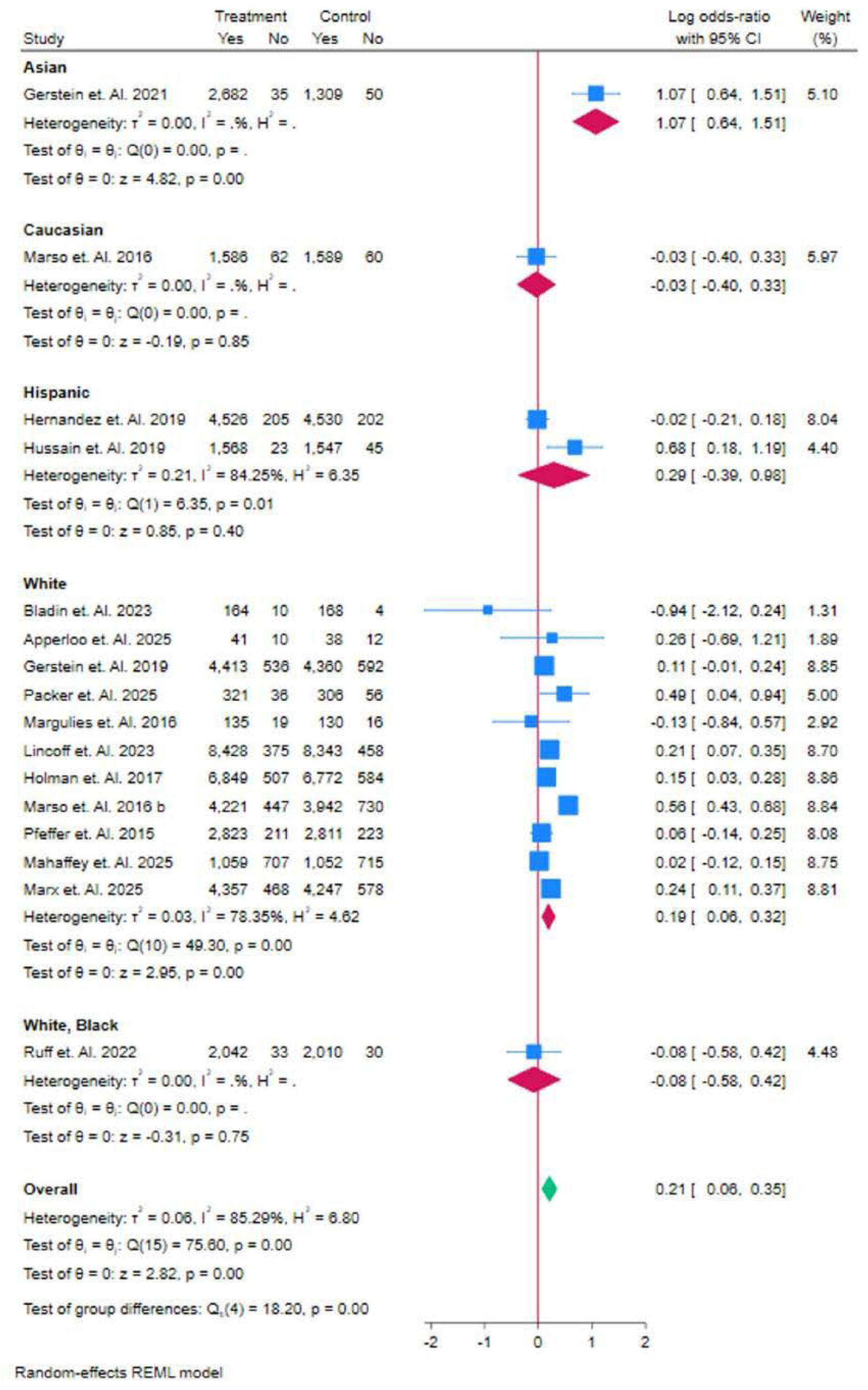
Overall survivor RACE Sub Group Analysis.

**Figure 5.**
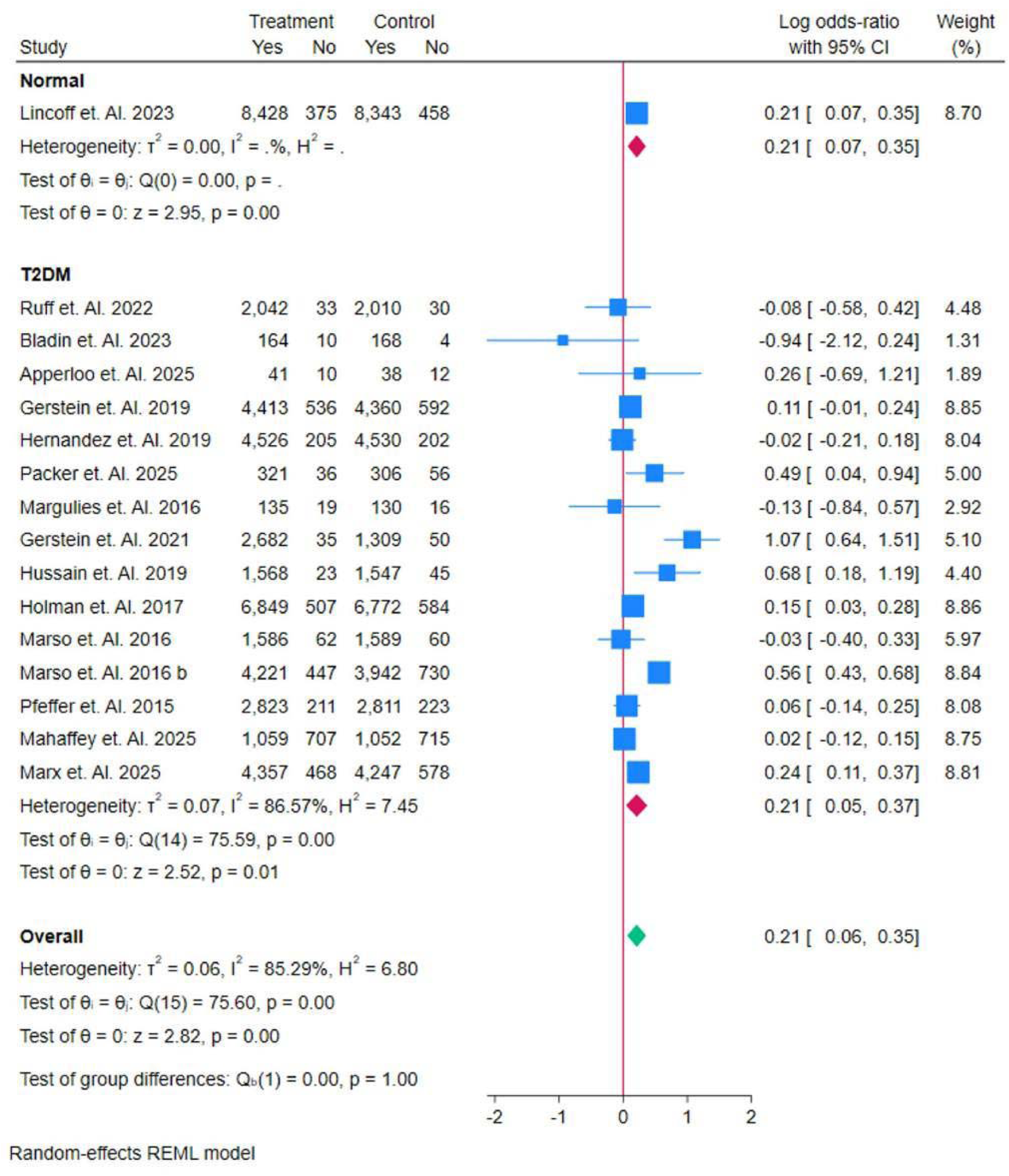
Overall survivor T2DM vs Normal Sub Group Analysis.

Figure 6 illustrates the subgroup analysis based on BMI for overall survival. In patients with BMI <30, the log odds-ratio (OR) ranged from −0.08 (95% CI: −0.58, 0.42) to 0.24 (95% CI: 0.11, 0.37), with an overall OR of 0.14 (95% CI: −0.02, 0.30). For patients with BMI >30, the OR varied from −0.94 (95% CI: −2.12, 0.24) to 1.07 (95% CI: 0.64, 1.51), with an overall OR of 0.24 (95% CI: 0.05, 0.43). The test of group differences showed no significant interaction between BMI groups (p = 0.44). The analysis indicates a modest survival benefit across both BMI subgroups. Figure 6 presents the subgroup analysis based on HbA1c levels for overall survival. In patients with HbA1c <6.5%, the log odds-ratio (OR) ranged from −0.94 (95% CI: −2.12, 0.24) to 0.21 (95% CI: 0.07, 0.35), with an overall OR of 0.06 (95% CI: −0.42, 0.55). In patients with HbA1c >6.5%, the OR varied from −0.63 (95% CI: −1.84, 0.57) to 1.07 (95% CI: 0.64, 1.51), with an overall OR of 0.23 (95% CI: 0.06, 0.39). The test of group differences showed no significant interaction between HbA1c subgroups (p = 0.53). These results indicate a modest survival benefit for both HbA1c subgroups with no significant difference between them. Figure 8 presents a subgroup analysis of SGLT2 inhibitor administration for overall survival. In patients who did not receive SGLT2 inhibitors, the log odds-ratio (OR) ranged from −0.94 (95% CI: −2.12, 0.24) to 0.56 (95% CI: 0.43, 0.68), with an overall OR of 0.16 (95% CI: 0.02, 0.31). In patients who received SGLT2 inhibitors, the OR ranged from −0.08 (95% CI: −0.58, 0.42) to 1.07 (95% CI: 0.64, 1.51), with an overall OR of 0.35 (95% CI: −0.01, 0.71). The test of group differences showed no significant interaction (p = 0.34), indicating that the use of SGLT2 inhibitors did not significantly alter the overall survival compared to those not receiving the medication.

**Figure 6.**
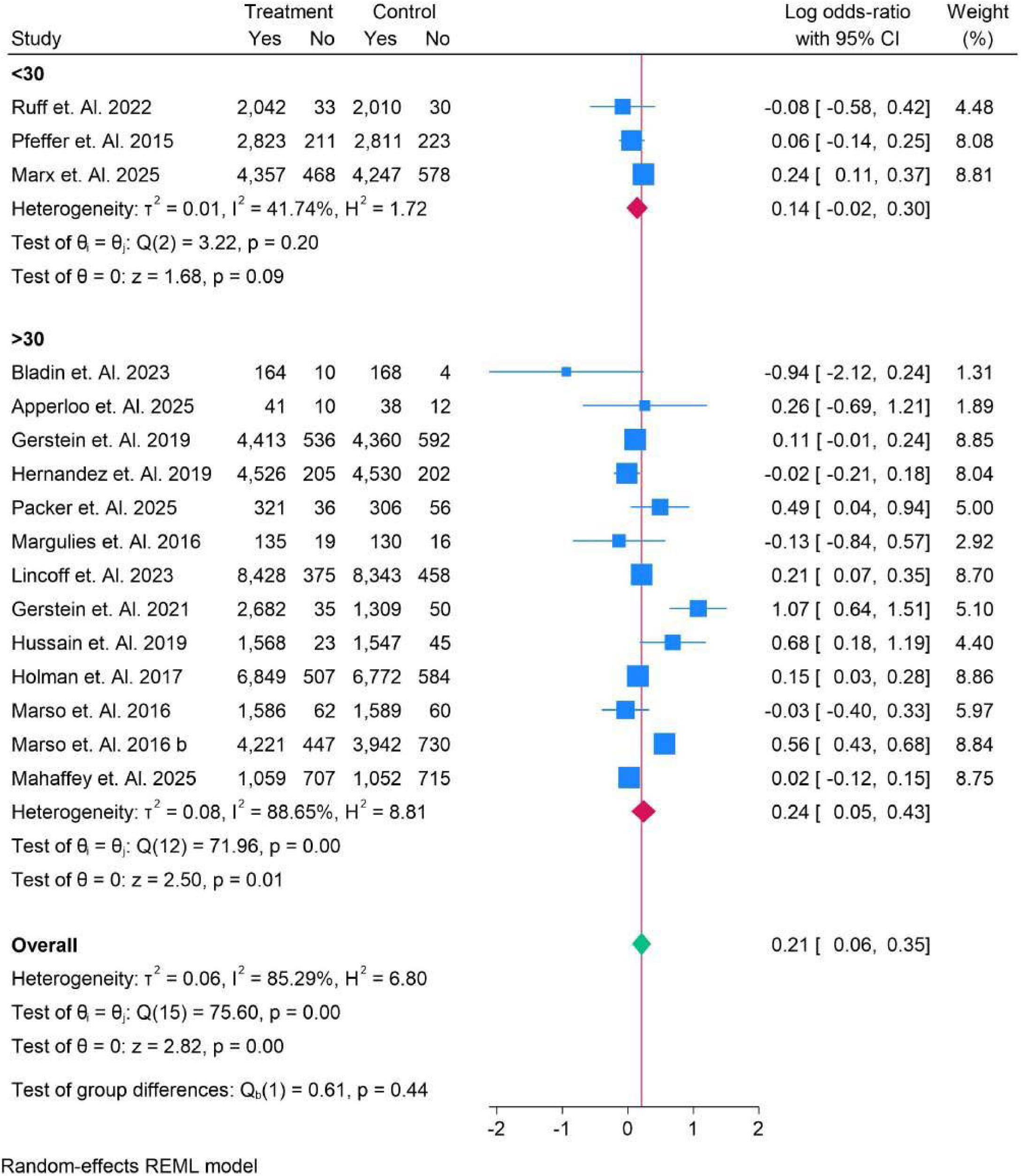
BMI Subgroup analysis for Overall survivors.

**Figure 7.**
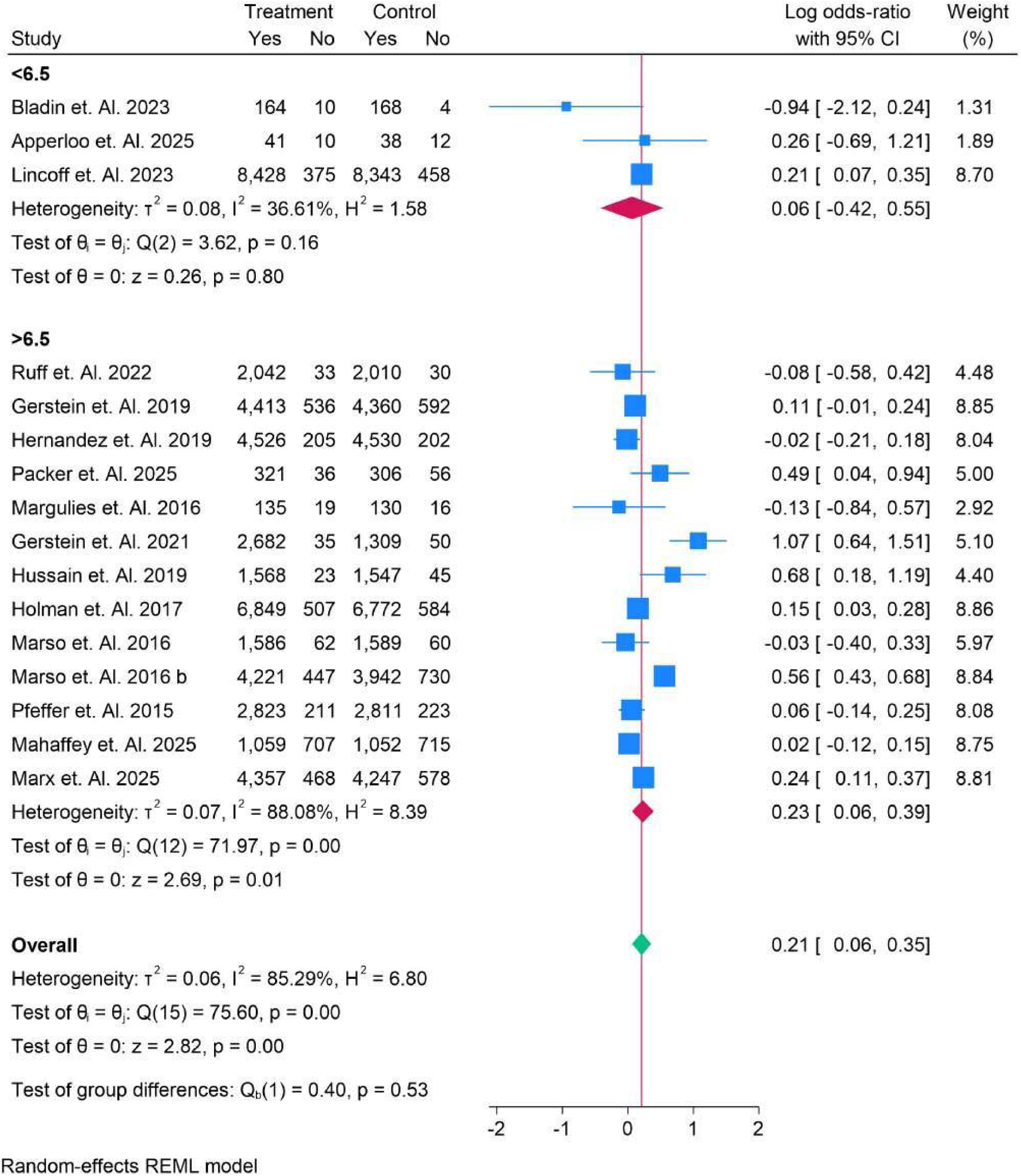
HBAl C Subgroup analysis for Overall survivors.

**Figure 8.**
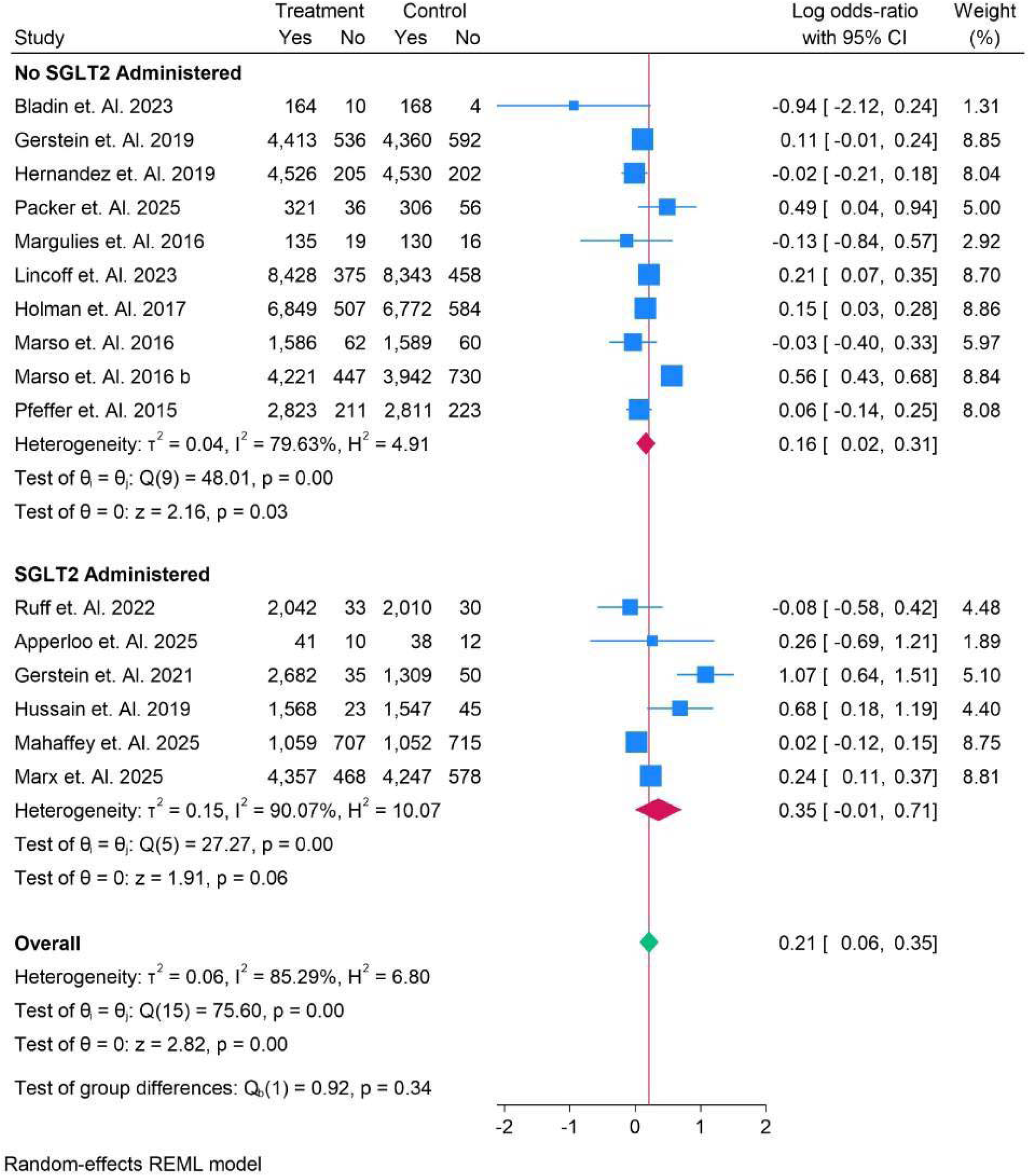
Subgroup Analysis of SGLT2 Administration for overall survival.

Figure 9 displays a subgroup analysis comparing daily versus weekly drug administration for overall survival. For daily administration, the log odds-ratio (OR) ranged from −0.94 (95% CI: −2.12, 0.24) to 0.24 (95% CI: 0.11, 0.37), with an overall OR of 0.14 (95% CI: −0.06, 0.34). For weekly administration, the OR ranged from −0.03 (95% CI: −0.40, 0.33) to 1.07 (95% CI: 0.64, 1.51), with an overall OR of 0.25 (95% CI: 0.06, 0.45). The test of group differences showed no significant interaction (p = 0.42), indicating that the frequency of drug administration did not significantly impact overall survival. Figure 10 presents a subgroup analysis based on the route of administration (oral vs. subcutaneous, SC) for overall survival. For oral administration, the log oddsratio (OR) ranged from 0.24 (95% CI: 0.11, 0.37) to 0.68 (95% CI: 0.18, 1.19), with an overall OR of 0.39 (95% CI: −0.03, 0.81). For SC administration, the OR ranged from −0.94 (95% CI: −2.12, 0.24) to 1.07 (95% CI: 0.64, 1.51), with an overall OR of 0.18 (95% CI: 0.02, 0.34). The test of group differences showed no significant interaction (p = 0.36), indicating no significant effect of the route of administration on overall survival. Figure 11 presents a subgroup analysis of various GLP-1 receptor agonists for overall survival. For Albiglutide, the log odds-ratio (OR) was −0.02 (95% CI: −0.21, 0.18), and for Dulaglutide, it was 0.11 (95% CI: −0.01, 0.18). Efpeglenatide showed a higher OR of 1.07 (95% CI: 0.64, 1.50). Exenatide demonstrated an OR of −0.08 (95% CI: −0.58, 0.42), while Liraglutide had an OR of 0.30 (95% CI: −0.35, 0.13). Lixisenatide had an OR of 0.06 (95% CI: −0.14, 0.14), and Semaglutide showed a favorable OR of 0.24 (95% CI: 0.11, 0.37). Tirzepatide had the highest OR of 0.49 (95% CI: 0.04, 0.94). The overall OR was 0.21 (95% CI: 0.06, 0.35), with significant heterogeneity (I² = 85.29%) and a test of group differences p-value of 0.00, suggesting notable variation in treatment effects across the different drugs.

**Figure 9.**
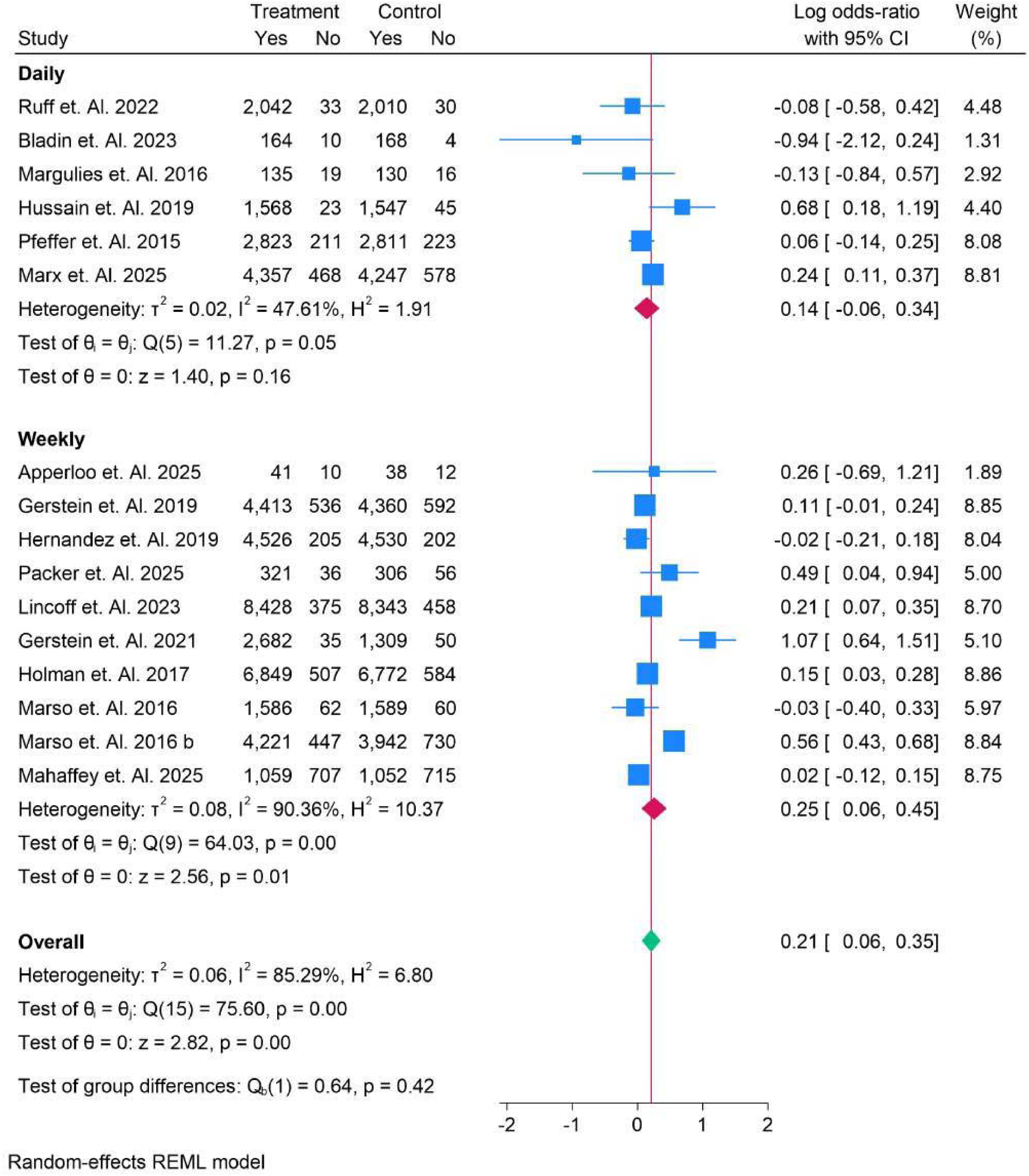
Subgroup Analysis of Daily Vs Weekly Dn1g Administration for Overall Survivals.

**Figure 10.**
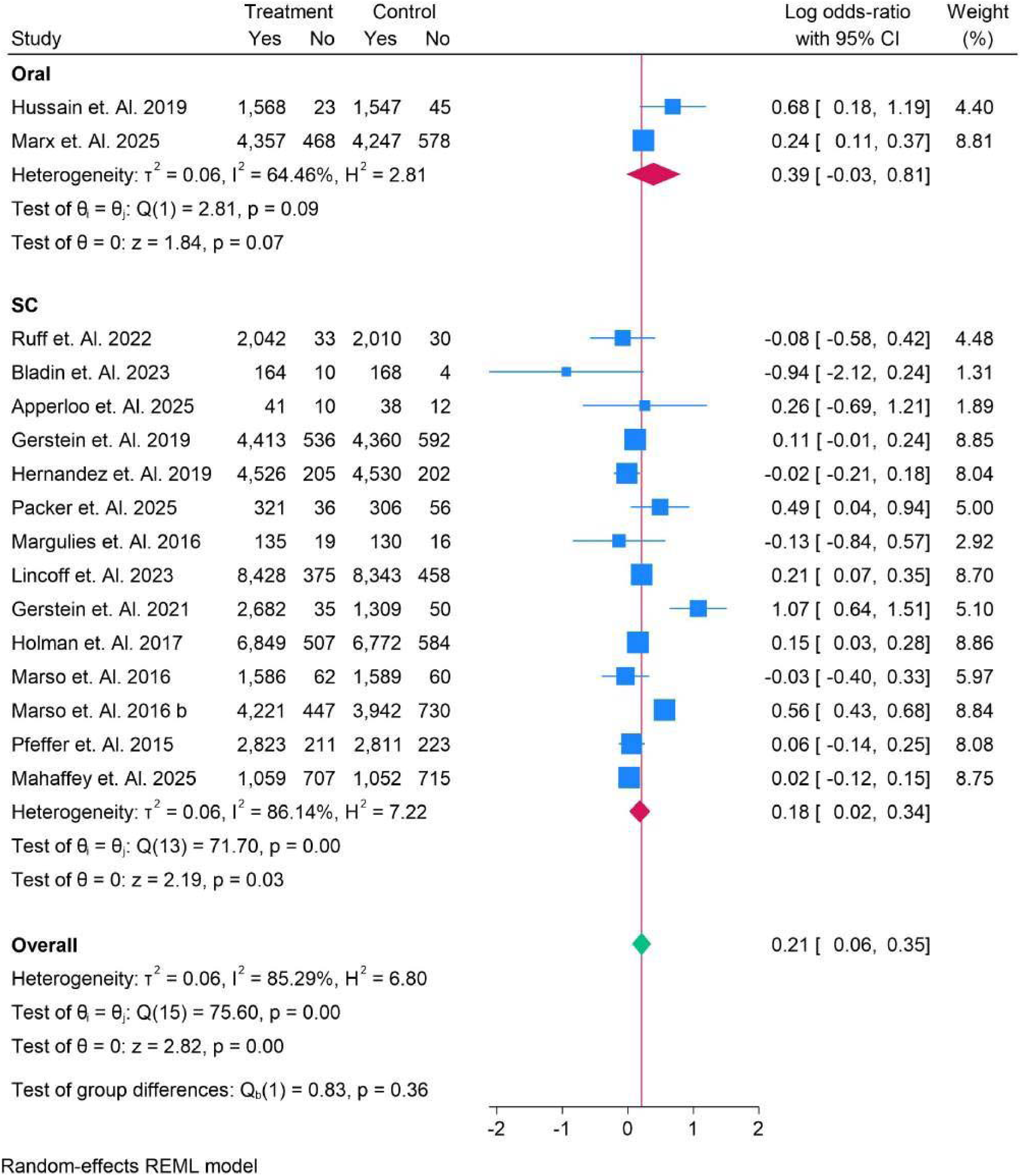
Route of Administration in Survivors Subgroup Analysis.

**Figure 11.**
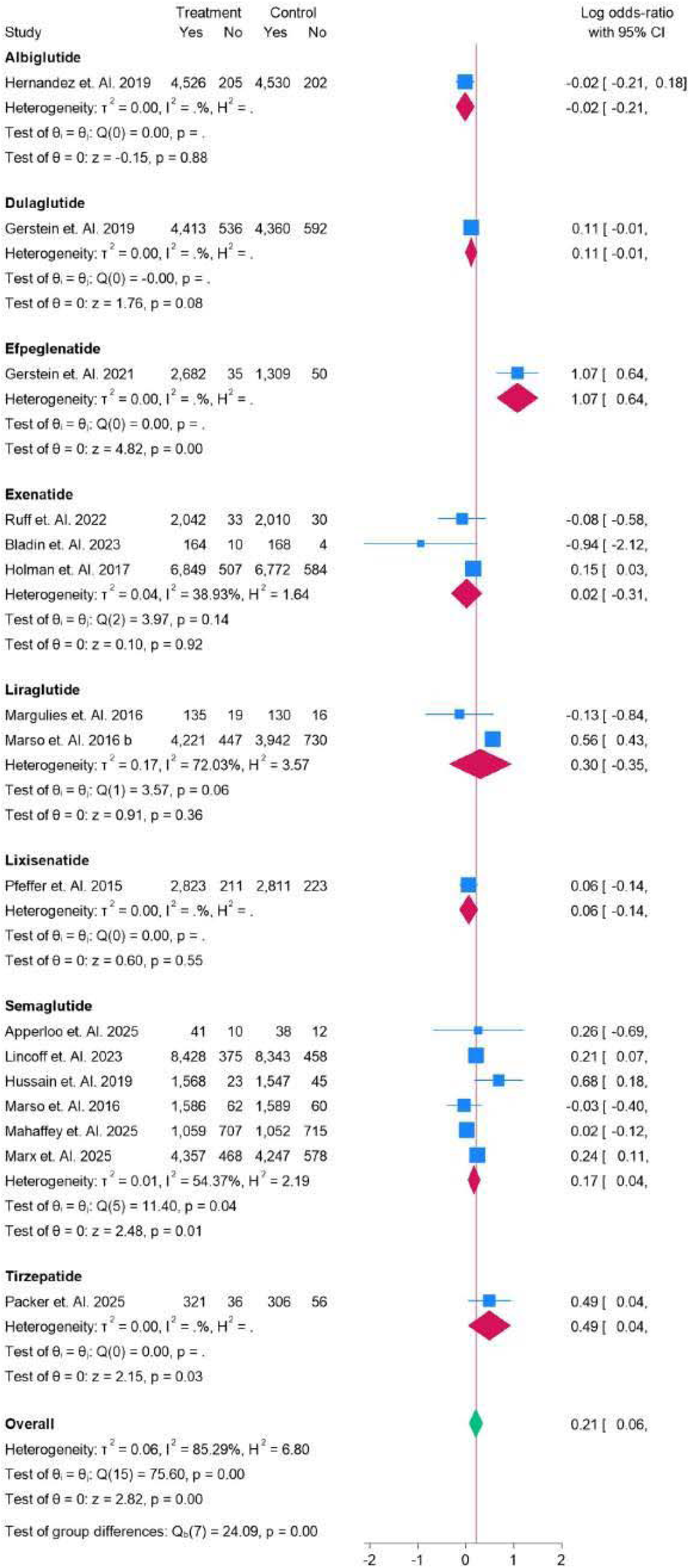
All types of drugsSubgroup analysis for Overall Survivorship.

### Fatal Stroke

Figure 12 displays a subgroup analysis based on age for fatal stroke risk. In patients aged <65 years, the log risk-ratio (RR) varied from −0.49 (95% CI: −0.96, 0.01) in the study by Gerstein et al. (2021) to 0.21 (95% CI: 0.15, 0.58) in Mahaffey et al. (2025), with an overall RR of 0.00 (95% CI: −0.11, 0.11). In patients aged >65 years, the RR ranged from −0.29 (95% CI: −1.03, 0.46) in Hussain et al. (2019) to −0.11 (95% CI: −0.33, 0.11) in Marx et al. (2025), with an overall RR of −0.14 (95% CI: −0.28, 0.00). The overall RR across both age groups was −0.04 (95% CI: −0.13, 0.06), with no significant group difference (p = 0.13). The results suggest a minimal effect of treatment on fatal stroke risk across all age groups. Figure 13 presents a race subgroup analysis for fatal stroke risk. In Asian patients, the log risk-ratio (RR) was −0.28 (95% CI: −0.73, 0.17), with no significant effect (p = 0.23). In Caucasians, the RR was −0.49 (95% CI: −0.96, −0.01), indicating a slightly significant reduction in fatal stroke risk (p = 0.04). Hispanic patients showed an RR of −0.04 (95% CI: −0.12, 0.05), with no significant effect (p = 0.42). For White patients, the RR ranged from −0.15 (95% CI: −0.35, 0.04) to 0.21 (95% CI: −0.15, 0.58), with an overall RR of 0.02 (95% CI: −0.12, 0.17). The White, Black subgroup showed an RR of −0.10 (95% CI: −0.66, 0.46). The overall RR was −0.04 (95% CI: −0.13, 0.06), with no significant group differences (p = 0.38). These findings suggest minimal impact of race on fatal stroke risk across the subgroups.

**Figure 12.**
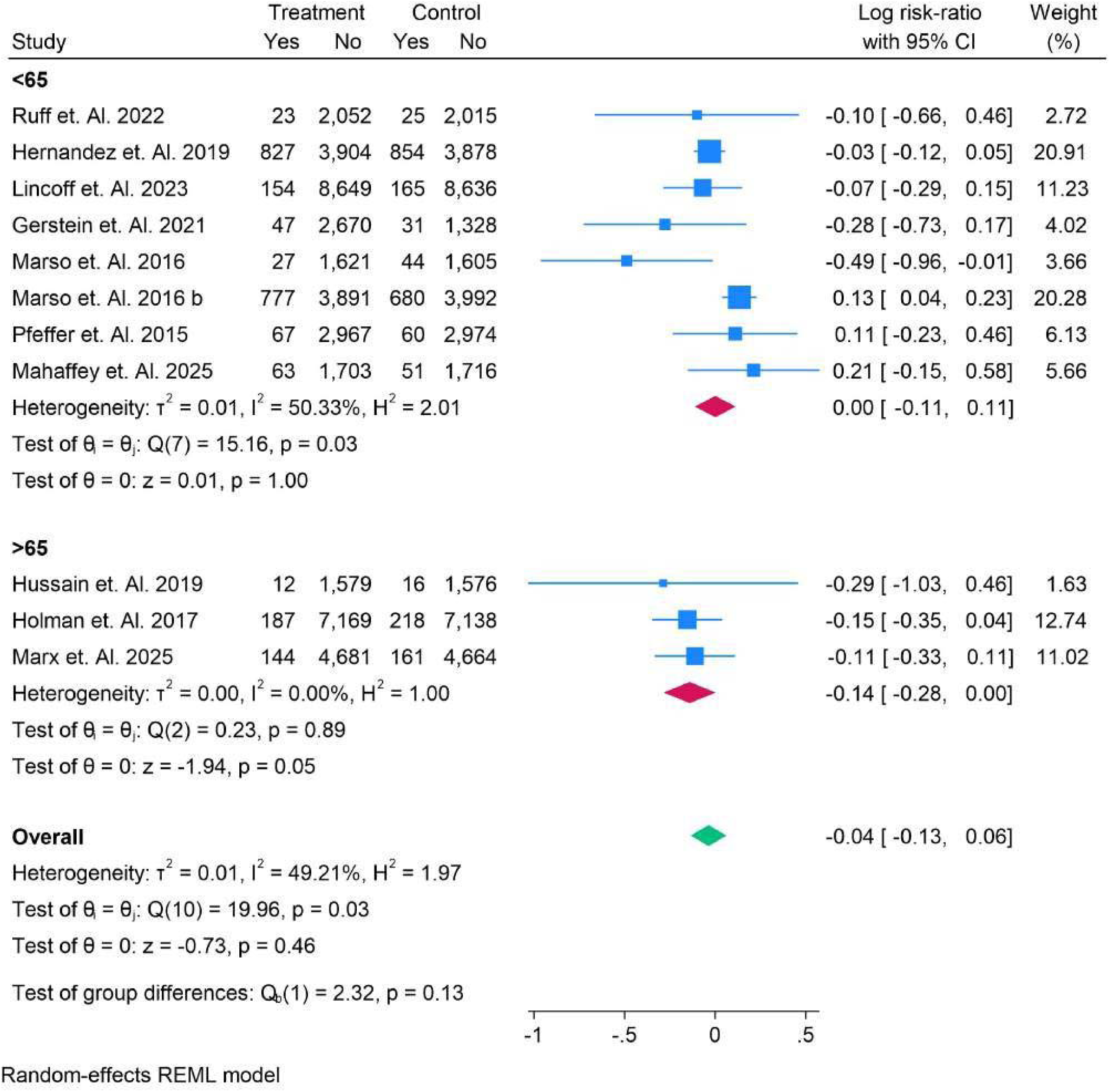
Age Sub-group Analysis in Fatal Stroke.

**Figure 13.**
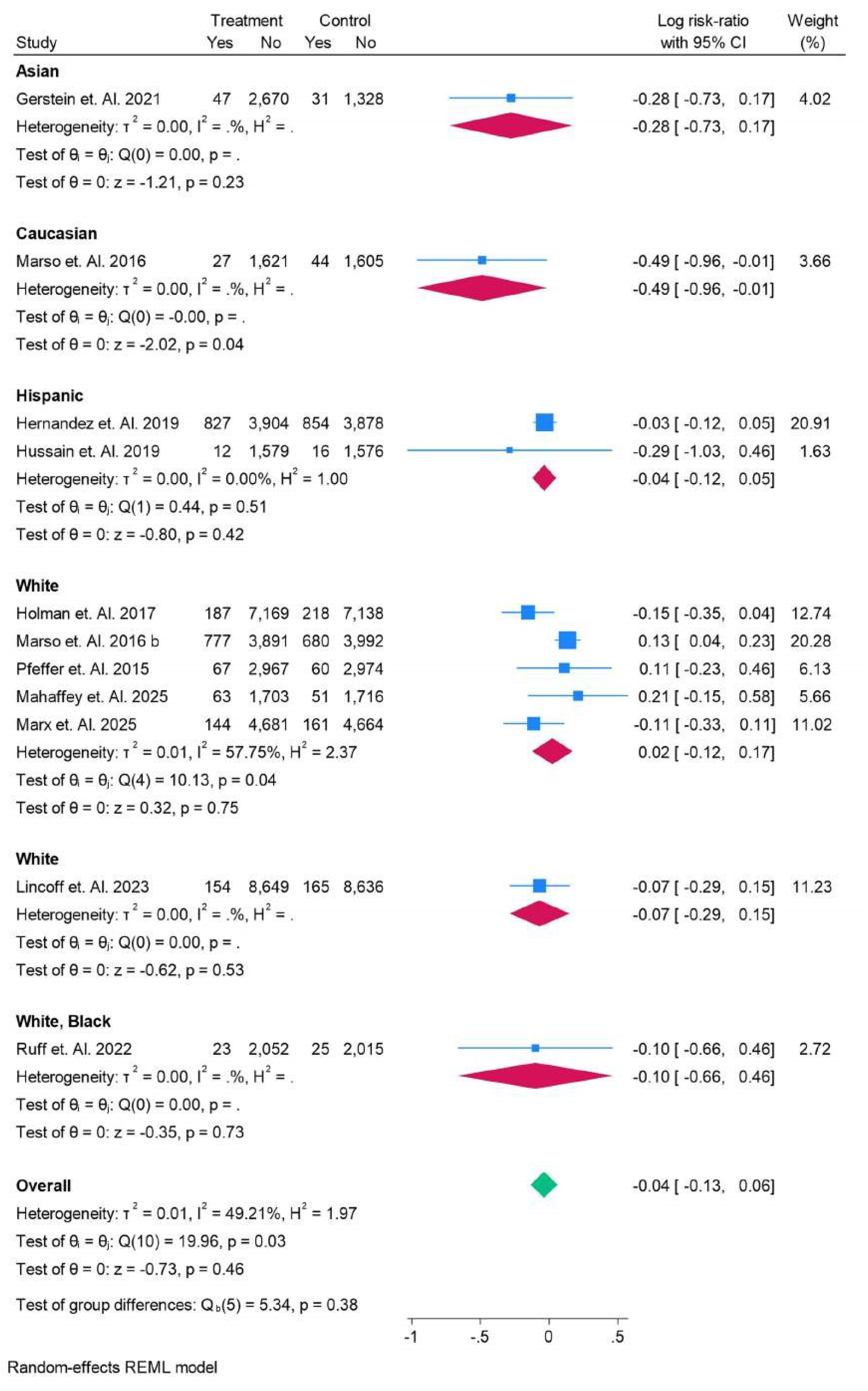
RACE Sub-group Analysis in Fatal Stroke.

Figure 14 presents a subgroup analysis comparing normal glucose tolerance (Normal) versus type 2 diabetes (T2DM) in fatal stroke risk. In patients with normal glucose levels, the log risk-ratio (RR) was −0.07 (95% CI: 0.29, 0.15). In patients with T2DM, the RR ranged from −0.49 (95% CI: −0.96, −0.01) to 0.13 (95% CI: 0.04, 0.23), with an overall RR of −0.04 (95% CI: −0.15, 0.07). The overall RR across both groups was −0.04 (95% CI: −0.13, 0.06), with no significant group differences (p = 0.79). These results suggest minimal differences in fatal stroke risk between patients with normal glucose levels and those with T2DM. Figure 15 displays a subgroup analysis of BMI in relation to fatal stroke risk. For patients with BMI <30, the log risk-ratio (RR) ranged from 0.11 (95% CI: −0.33, 0.11) to 0.11 (95% CI: −0.23, 0.46), with an overall RR of −0.05 (95% CI: −0.23, 0.12). In patients with BMI >30, the RR varied from −0.49 (95% CI: −0.96, −0.01) to 0.21 (95% CI: −0.15, 0.58), with an overall RR of −0.04 (95% CI: −0.17, 0.08). The overall RR for both subgroups was −0.04 (95% CI: −0.13, 0.06), with no significant group differences (p = 0.94). This suggests minimal impact of BMI on fatal stroke risk across both subgroups. Figure 16 shows a subgroup analysis based on HbA1c levels for fatal stroke risk. In patients with HbA1c <6.5%, the log risk-ratio (RR) ranged from −0.10 (95% CI: −0.66, 0.46) to −0.07 (95% CI: 0.29, 0.15), with an overall RR of −0.07 (95% CI: −0.29, 0.15). In patients with HbA1c >6.5%, the RR varied from −0.49 (95% CI: −0.96, −0.01) to 0.13 (95% CI: 0.04, 0.23), with an overall RR of −0.04 (95% CI: −0.15, 0.07). The overall RR across both groups was −0.04 (95% CI: −0.13, 0.06), with no significant group differences (p = 0.79). These findings suggest minimal impact of HbA1c levels on fatal stroke risk across both subgroups.

**Figure 14.**
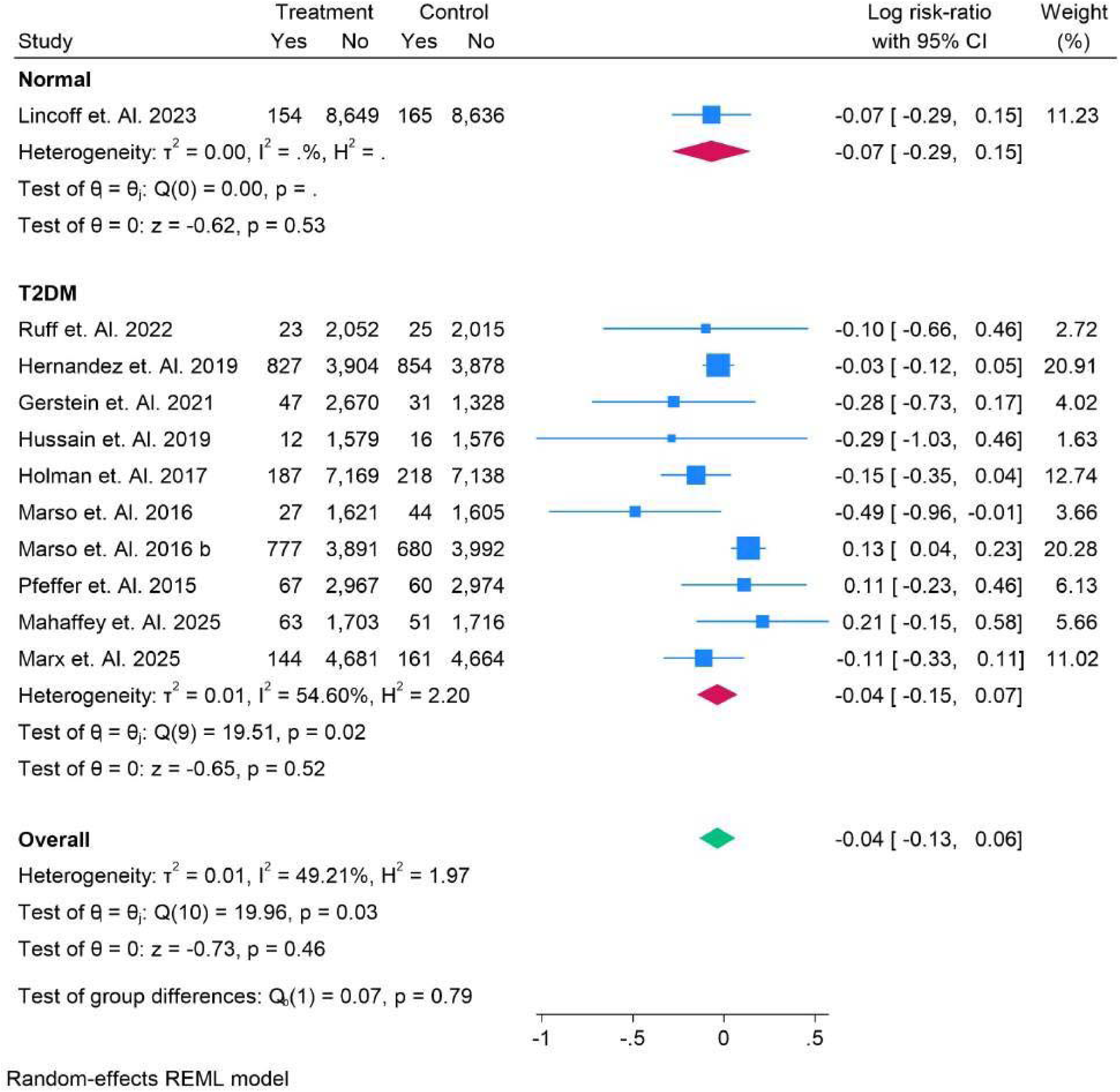
Normal vs T2DM Sub-group Analysis in Fatal Stroke.

**Figure 15.**
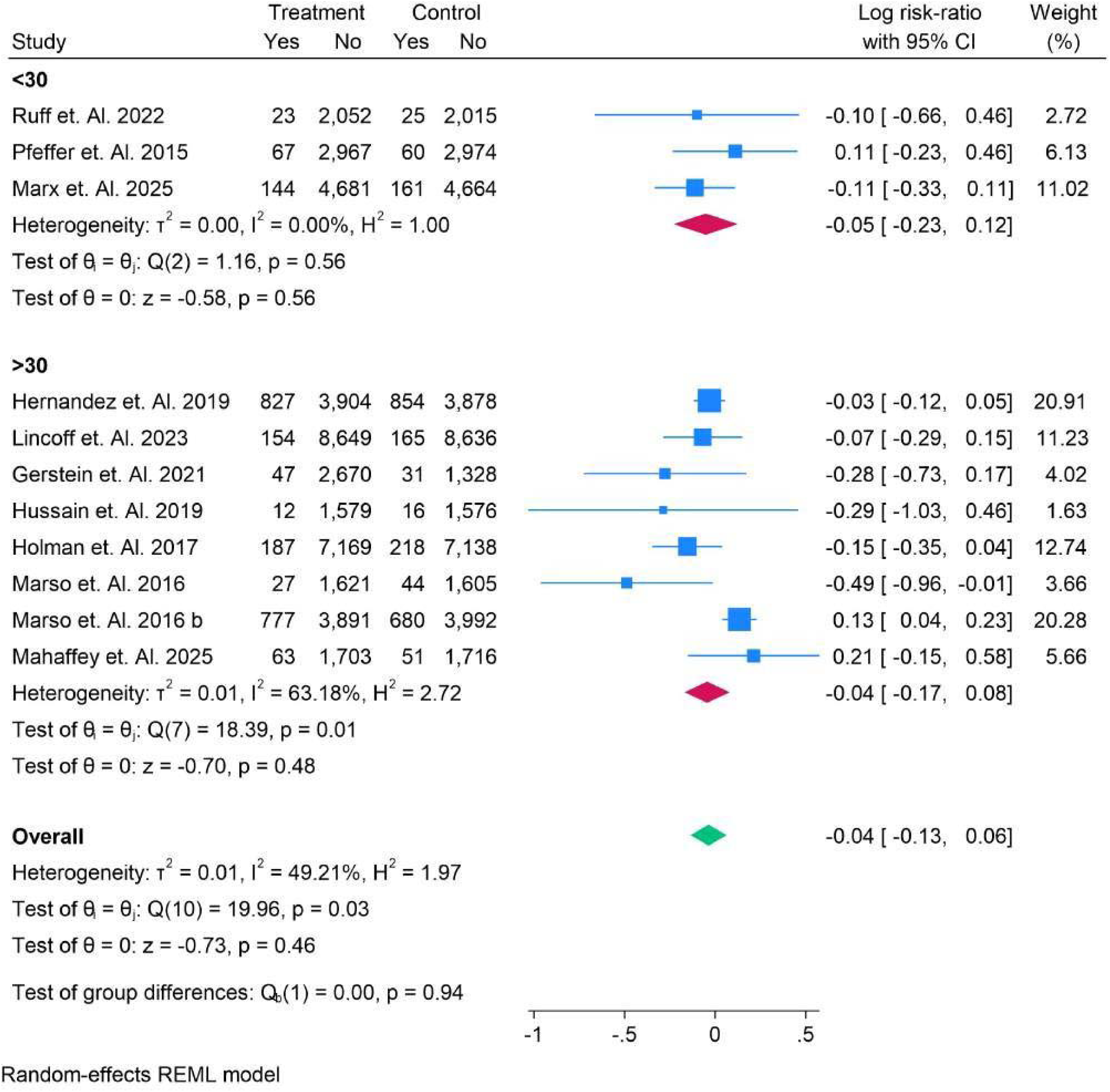
BMI Sub-group Analysis in Fatal Stroke.

**Figure 16.**
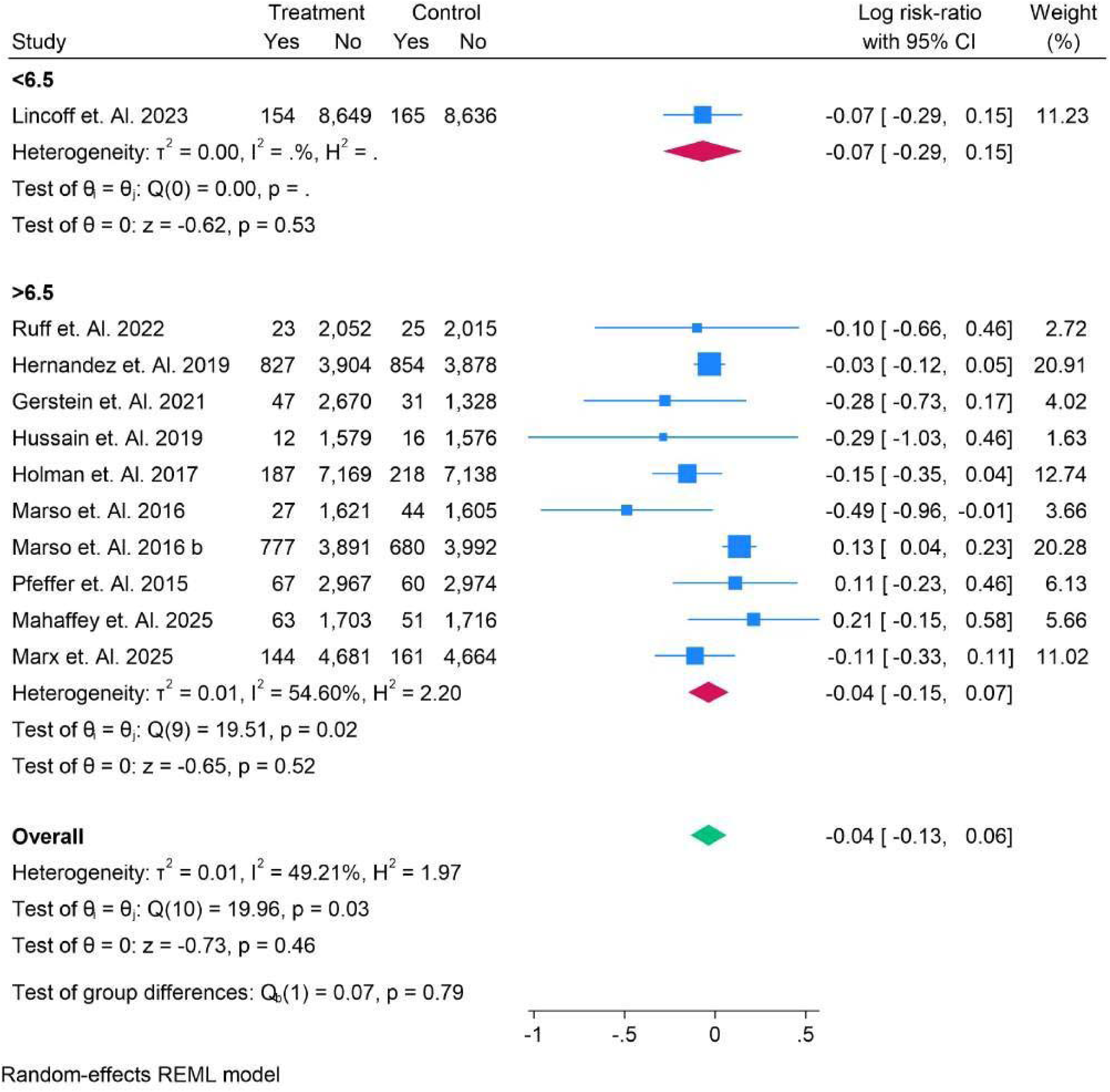
HBAIC Sub-group Analysis in Fatal Stroke.

Figure 17 presents a subgroup analysis comparing the administration of SGLT2 inhibitors versus no SGLT2 inhibitors for fatal stroke risk. In patients who did not receive SGLT2 inhibitors, the log risk-ratio (RR) ranged from −0.49 (95% CI: −0.96, −0.01) to 0.11 (95% CI: −0.23, 0.46), with an overall RR of −0.03 (95% CI: −0.15, 0.09). In patients who received SGLT2 inhibitors, the RR varied from −0.29 (95% CI: −1.03, 0.46) to 0.21 (95% CI: −0.15, 0.58), with an overall RR of −0.08 (95% CI: −0.24, 0.09). The overall RR for both subgroups was 0.04 (95% CI: −0.13, 0.06), with no significant group differences (p = 0.65). These findings suggest minimal effect of SGLT2 inhibitors on fatal stroke risk across both groups. Figure 18 presents a subgroup analysis comparing daily versus weekly drug administration for fatal stroke risk. In the daily administration group, the log risk-ratio (RR) ranged from −0.29 (95% CI: −1.03, 0.46) to 0.11 (95% CI: −0.23, 0.46), with an overall RR of −0.06 (95% CI: −0.24, 0.11). In the weekly administration group, the RR ranged from −0.49 (95% CI: −0.96, 0.01) to 0.21 (95% CI: −0.15, 0.58), with an overall RR of −0.04 (95% CI: −0.16, 0.09). The overall RR for both subgroups was −0.04 (95% CI: −0.13, 0.06), with no significant group differences (p = 0.81). These findings suggest minimal impact of the administration frequency on fatal stroke risk across both subgroups. Figure 19 presents a subgroup analysis comparing oral versus subcutaneous (SC) drug administration for fatal stroke risk. In the oral administration group, the log risk-ratio (RR) ranged from −0.29 (95% CI: −1.03, 0.46) to −0.11 (95% CI: −0.33, 0.11), with an overall RR of −0.13 (95% CI: −0.34, 0.09). In the SC administration group, the RR ranged from −0.49 (95% CI: −0.96, −0.01) to 0.21 (95% CI: −0.15, 0.58), with an overall RR of −0.02 (95% CI: 0.13, 0.08). The overall RR for both subgroups was −0.04 (95% CI: −0.13, 0.06), with no significant group differences (p = 0.41). These findings suggest no significant impact of the route of administration on fatal stroke risk across both subgroups. **ChatGPT said:** Figure 20 presents a subgroup analysis of various GLP-1 receptor agonists for fatal stroke risk. For **Albiglutide**, the log risk-ratio (RR) was −0.03 (95% CI: −0.12, 0.05), indicating no significant effect on fatal stroke risk. **Efpeglenatide** showed an RR of −0.28 (95% CI: −0.73, 0.17), suggesting a potential reduction, though not statistically significant. **Exenatide** had an RR ranging from −0.15 (95% CI: −0.35, 0.04) to −0.10 (95% CI: −0.66, 0.46), with results leaning toward no significant effect. **Liraglutide** showed a favorable RR of 0.13 (95% CI: 0.04, 0.23), indicating a modest potential benefit. **Lixisenatide** had an RR of 0.11 (95% CI: −0.23, 0.46), showing no conclusive effect. **Semaglutide** showed a range from −0.49 (95% CI: −0.96, −0.01) to 0.21 (95% CI: −0.15, 0.58), indicating variation in treatment outcomes. The overall RR for all drugs was −0.04 (95% CI: −0.13, 0.06), with significant heterogeneity (I² = 49.21%) and a test for group differences p-value of 0.01, suggesting slight variation across treatments, but no conclusive reduction in fatal stroke risk.

**Figure 17.**
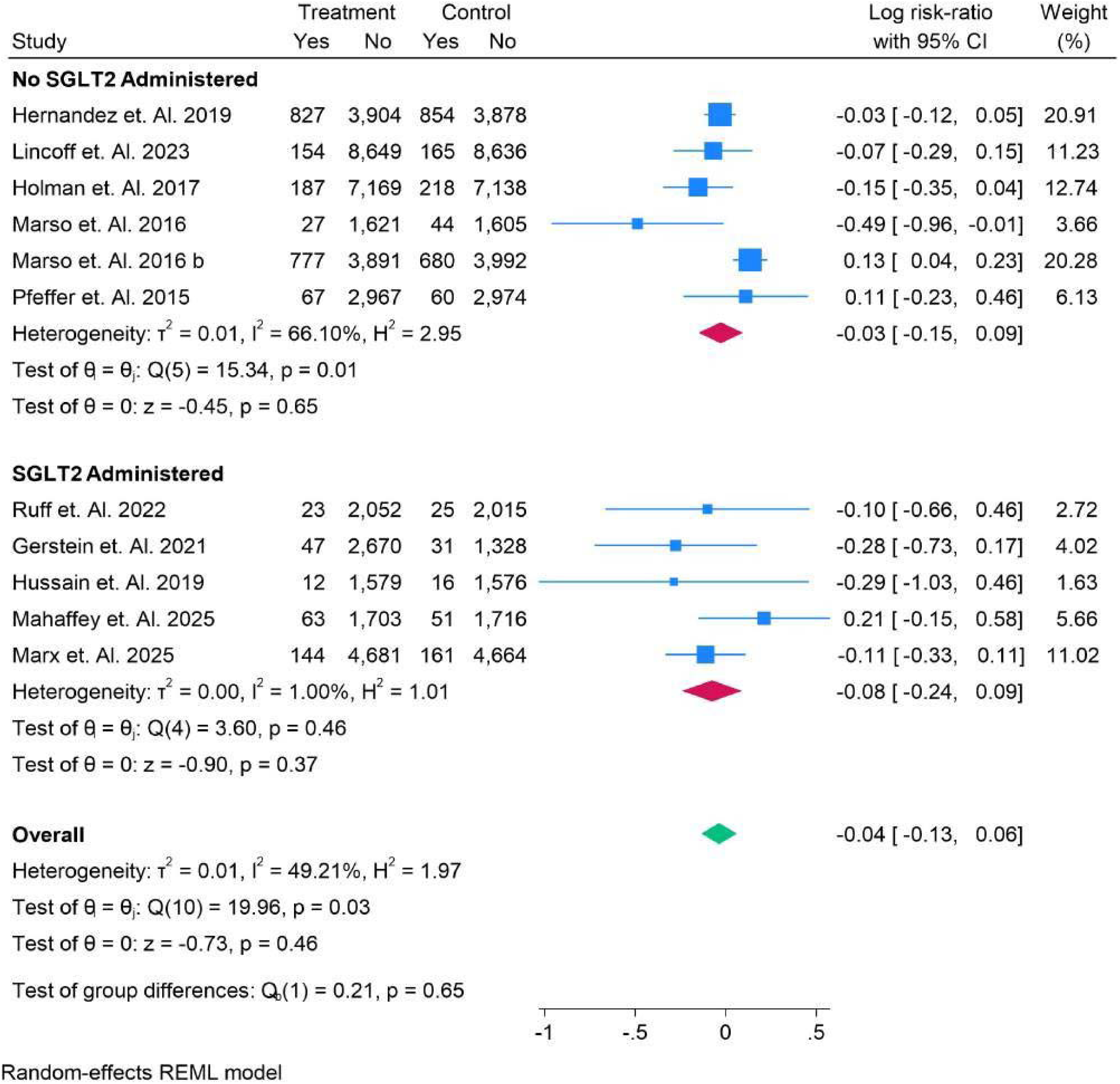
SGLT2 vs No SGLT2 Sub-group Analysis in Fatal Stroke.

**Figure 18.**
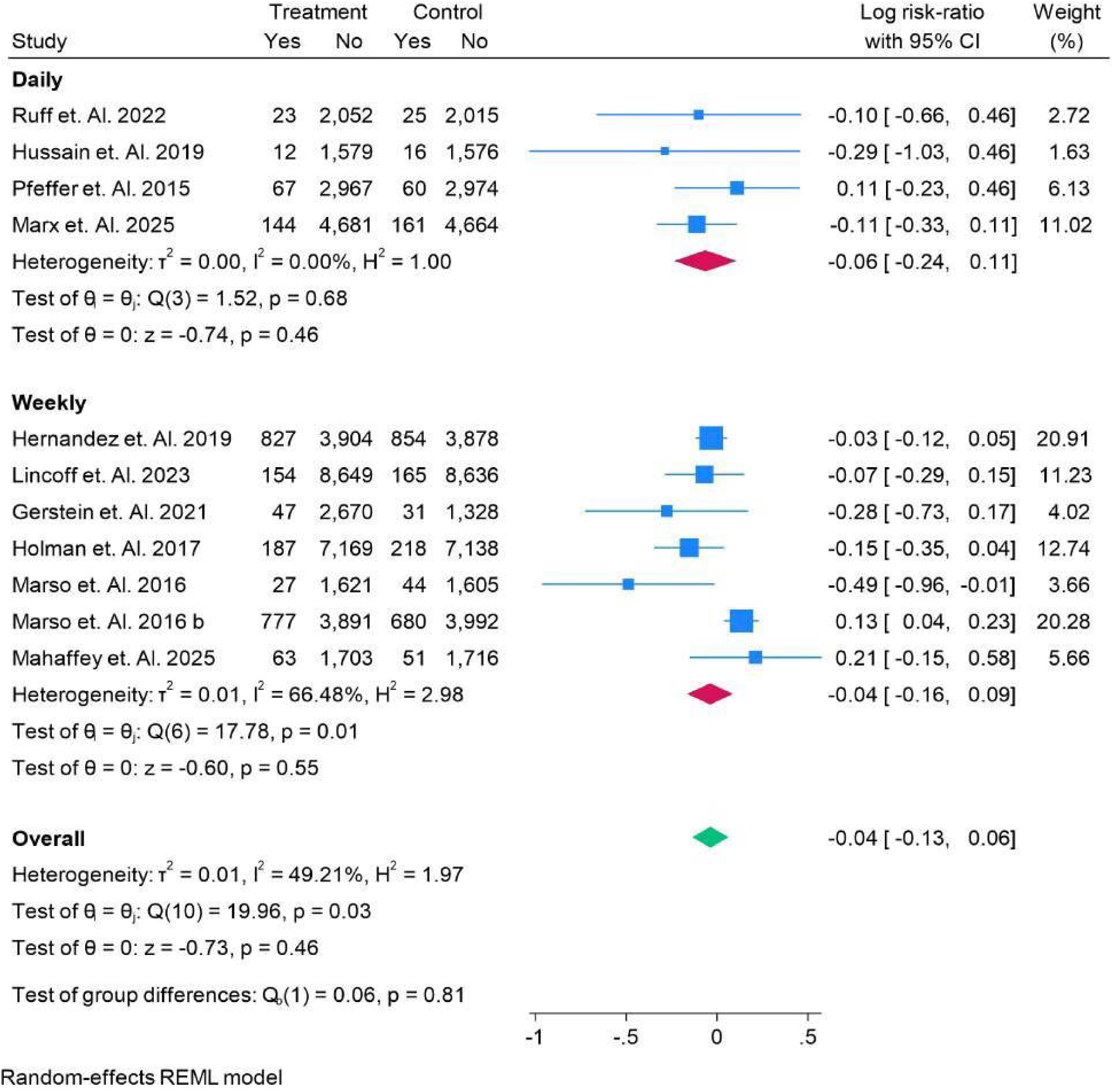
Daily vs Weekly Sub-group Analysis in Fatal Stroke.

**Figure 19.**
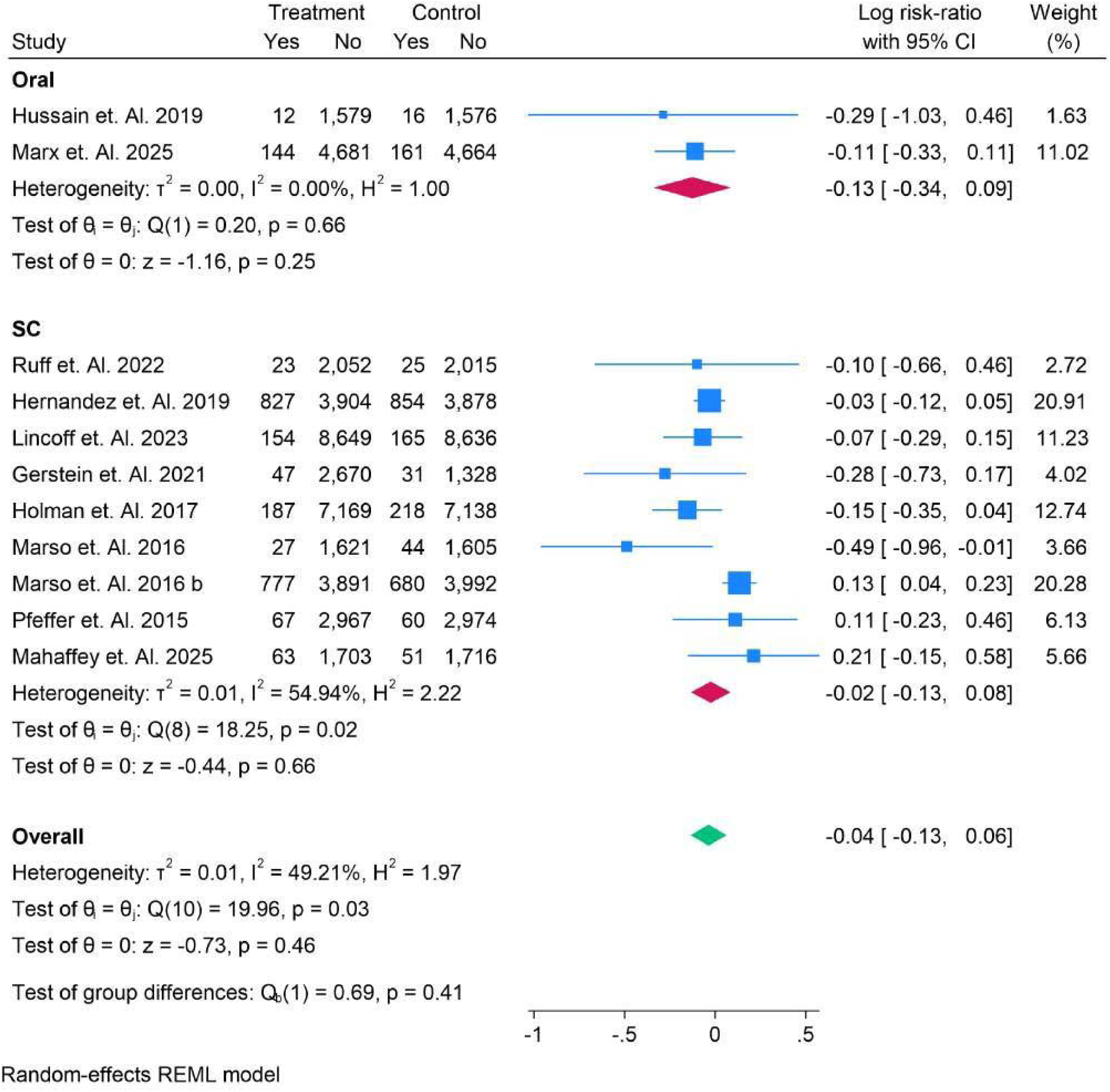
Oral vs Sub-CutaneousSub-group Analysis in Fatal Stroke.

**Figure 20.**
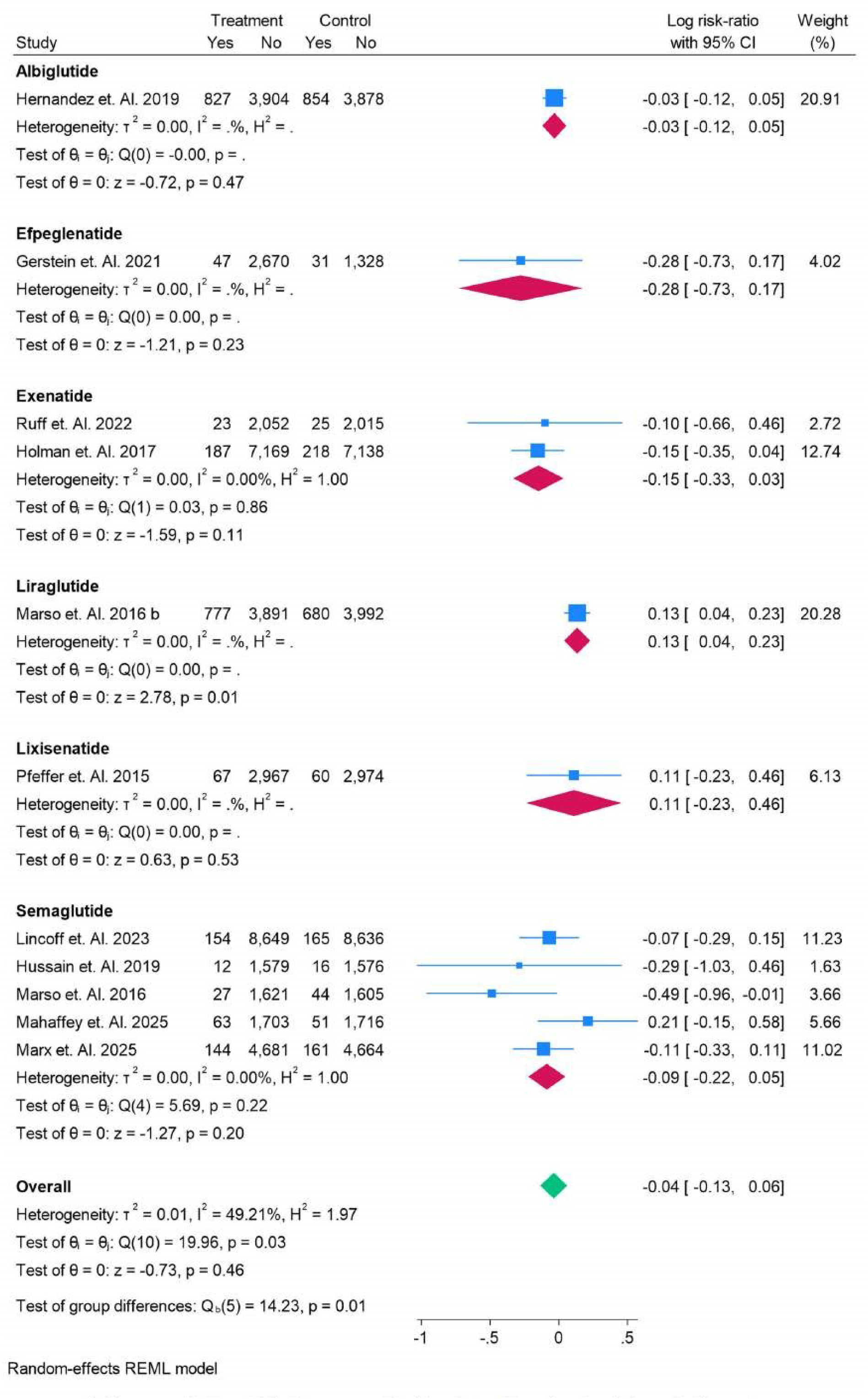
All Drugs Sub-Analysis in Fatal Stroke.

### Non-Fatal Stroke

Figure 21 presents a subgroup analysis based on age for non-fatal stroke risk. In patients aged <65 years, the log risk-ratio (RR) ranged from −0.22 (95% CI: −0.56, 0.12) to 0.49 (95% CI: 0.01, 0.96), with an overall RR of 0.07 (95% CI: −0.06, 0.20). For patients aged >65 years, the RR ranged from −0.29 (95% CI: −1.03, 0.46) to −0.11 (95% CI: −0.33, 0.11), with an overall RR of −0.13 (95% CI: −0.34, 0.09). The overall RR across both age groups was 0.04 (95% CI: −0.08, 0.16), with no significant group differences (p = 0.12). These findings suggest minimal effect of age on non-fatal stroke risk across the subgroups. Figure 22 presents a race subgroup analysis for nonfatal stroke risk. In **Asian** patients, the log risk-ratio (RR) was −0.22 (95% CI: −0.56, 0.12), indicating no significant effect. For **Caucasians**, the RR was 0.49 (95% CI: 0.01, 0.96), showing a potentially significant increase in non-fatal stroke risk. In **Hispanic** patients, the RR ranged from −0.29 (95% CI: −1.03, 0.46) to −0.03 (95% CI: −0.15, 0.10), with no significant effect. For **White** patients, the RR ranged from −0.11 (95% CI: −0.33, 0.11) to 0.23 (95% CI: 0.11, 0.35), with an overall RR of 0.07 (95% CI: −0.13, 0.28). In the **White, Black** subgroup, the RR was −0.10 (95% CI: −0.66, 0.46), showing no significant effect. The overall RR was 0.04 (95% CI: −0.08, 0.16), with no significant differences across race groups (p = 0.23), indicating minimal variation in non-fatal stroke risk by race. Figure 23 presents a subgroup analysis comparing normal glucose levels (Normal) versus type 2 diabetes (T2DM) for non-fatal stroke risk. In patients with normal glucose levels, the log riskratio (RR) was 0.07 (95% CI: −0.15, 0.29). In patients with T2DM, the RR ranged from −0.29 (95% CI: −1.03, 0.46) to 0.49 (95% CI: 0.01, 0.96), with an overall RR of 0.03 (95% CI: −0.11, 0.17). The overall RR for both groups was 0.04 (95% CI: −0.08, 0.16), with no significant group differences (p = 0.78). These results suggest minimal impact of diabetes status on non-fatal stroke risk across the subgroups. Figure 24 presents a subgroup analysis of BMI for non-fatal stroke risk. In patients with BMI <30, the log risk-ratio (RR) ranged from −0.10 (95% CI: −0.66, 0.46) to −0.11 (95% CI: −0.33, 0.11), with an overall RR of −0.06 (95% CI: −0.25, 0.12). In patients with BMI >30, the RR ranged from 0.23 (95% CI: 0.11, 0.35) to 0.49 (95% CI: 0.01, 0.96), with an overall RR of 0.07 (95% CI: −0.08, 0.22). The overall RR across both BMI subgroups was 0.04 (95% CI: −0.08, 0.16), with no significant group differences (p = 0.28). These findings suggest a minimal effect of BMI on nonfatal stroke risk across both subgroups.

**Figure 21.**
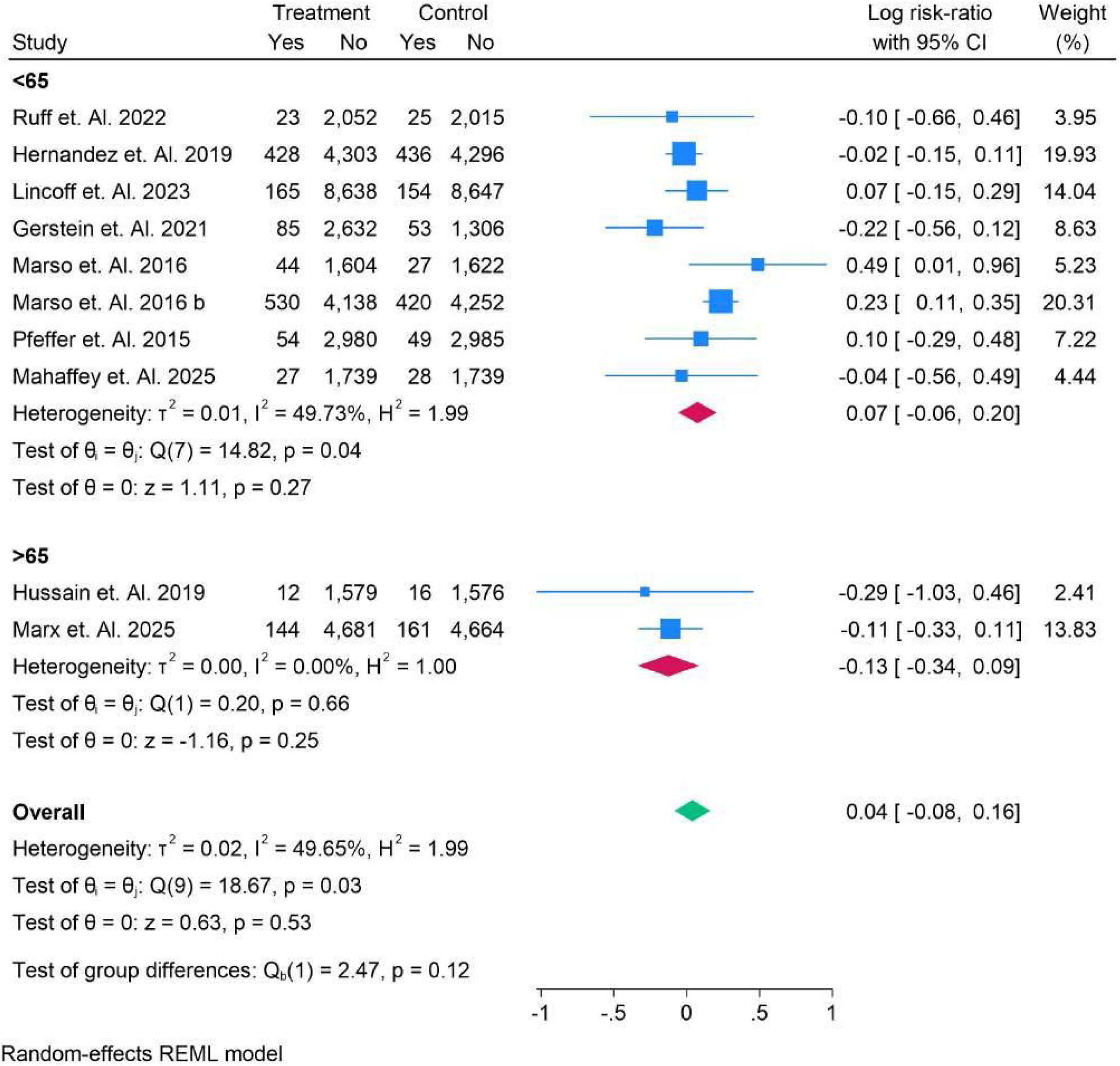
Age in Sub-Analysis in Non-Fatal Stroke.

**Figure 22.**
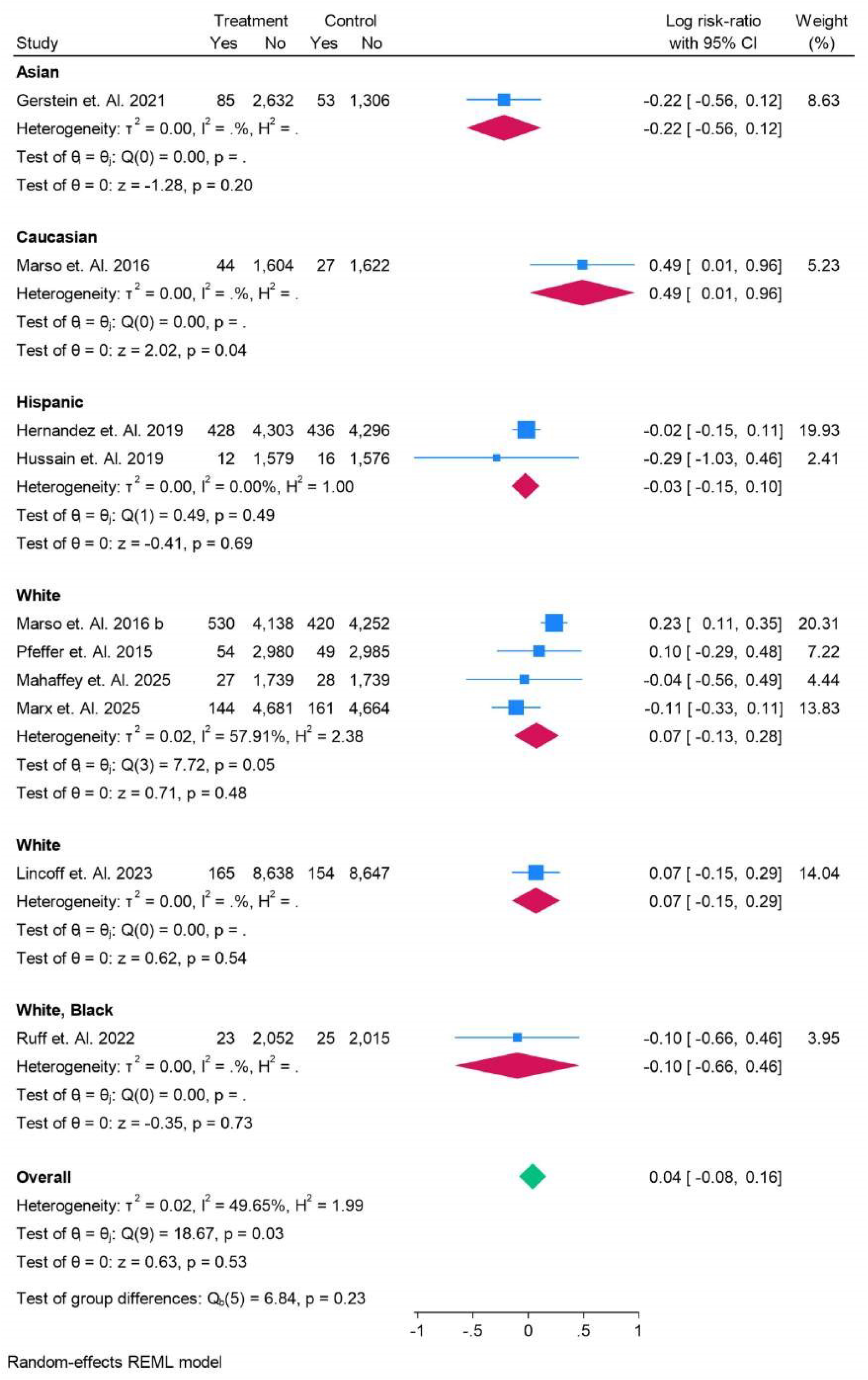
Race in Sub-Analysis in Non-Fatal Stroke.

**Figure 23.**
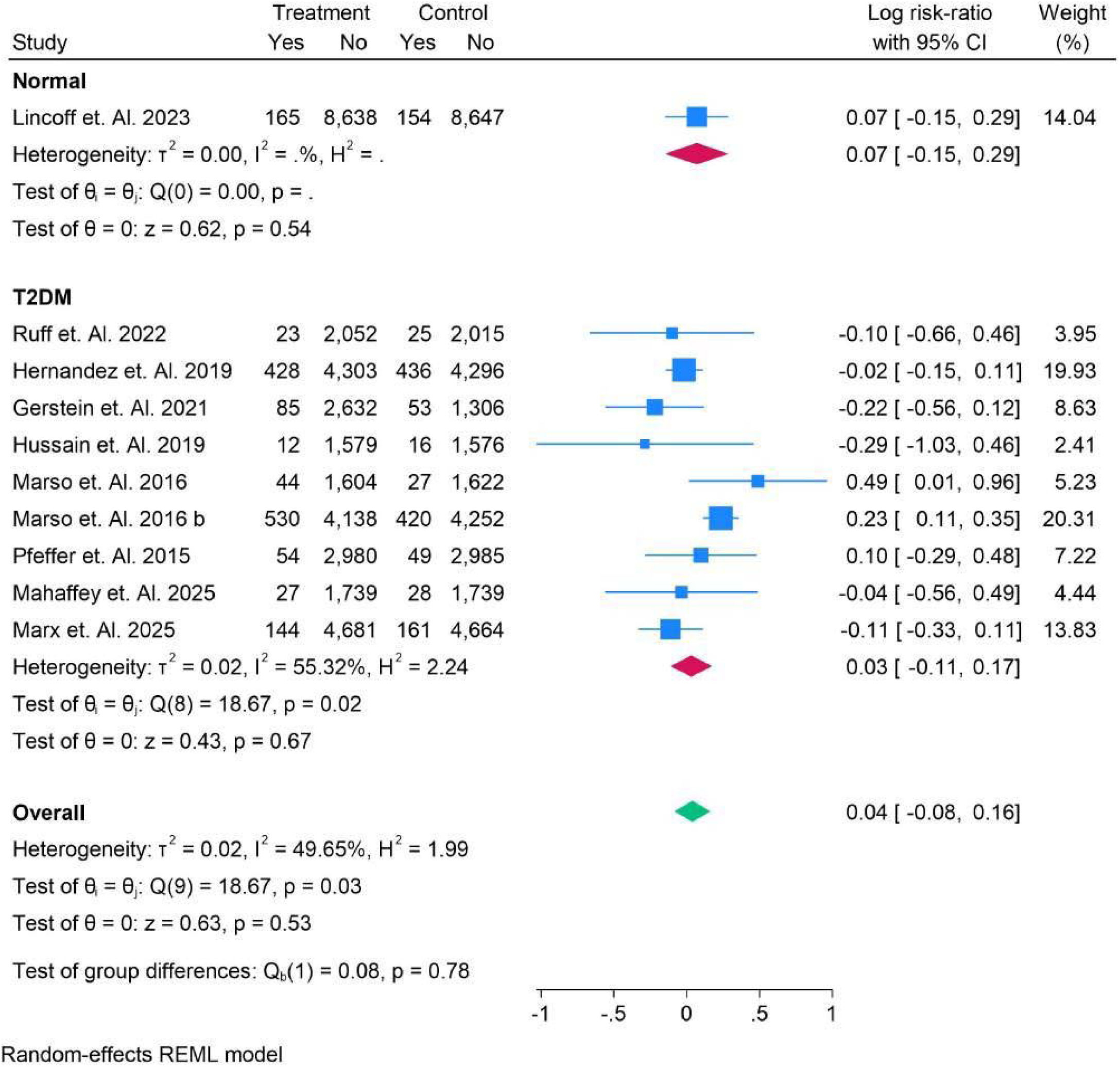
Notmal vs T2DM in Sub-Analysis in Non-Fatal Stroke.

**Figure 24.**
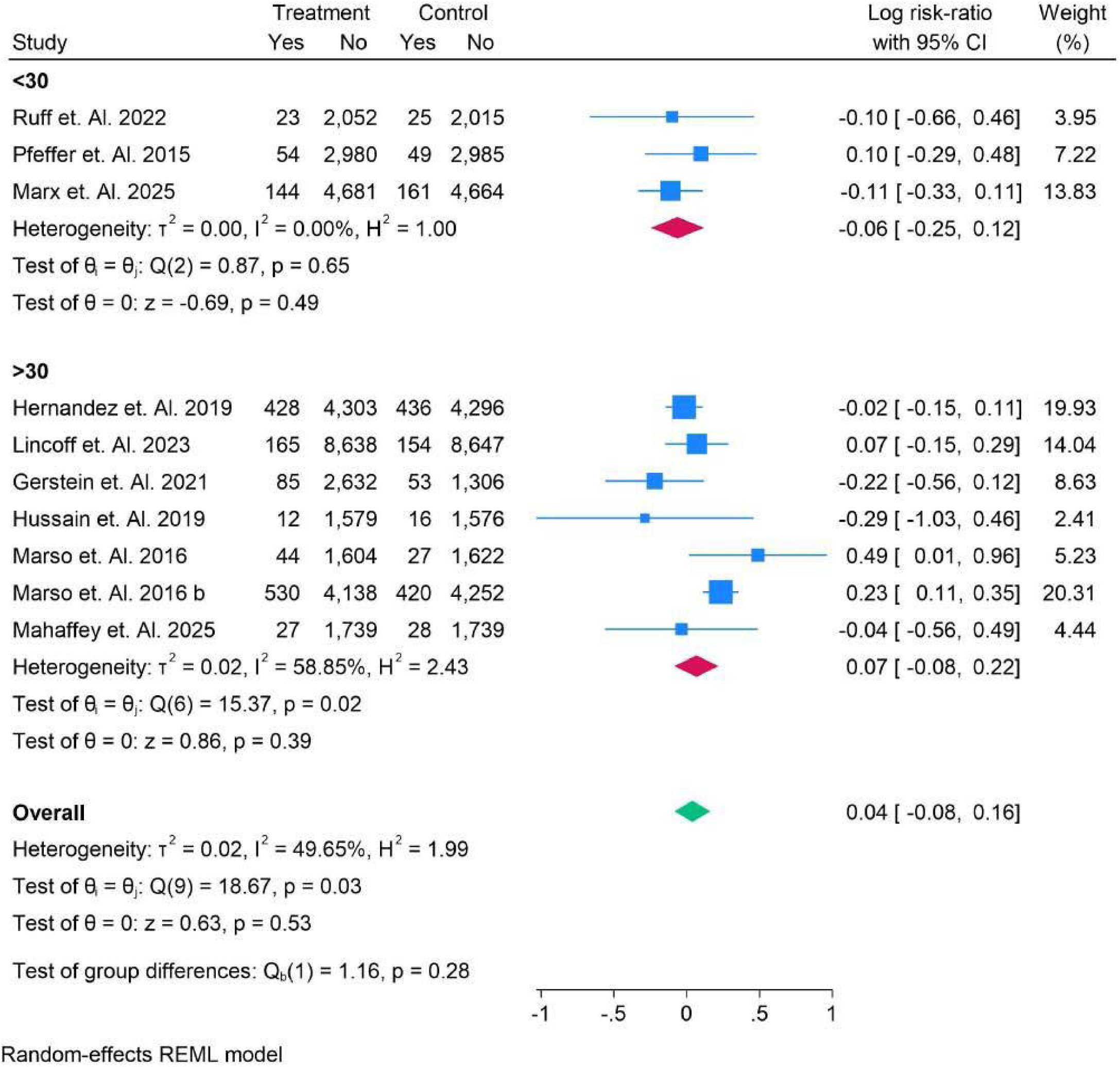
BMI in Sub-Analysis in Non-Fatal Stroke.

Figure 25 presents a subgroup analysis of HbA1c levels for non-fatal stroke risk. In patients with HbA1c <6.5%, the log risk-ratio (RR) was 0.07 (95% CI: −0.15, 0.29). In patients with HbA1c >6.5%, the RR ranged from 0.29 (95% CI: −1.03, 0.46) to 0.49 (95% CI: 0.01, 0.96), with an overall RR of 0.03 (95% CI: −0.11, 0.17). The overall RR for both subgroups was 0.04 (95% CI: −0.08, 0.16), with no significant group differences (p = 0.78). These findings suggest minimal impact of HbA1c levels on non-fatal stroke risk across both subgroups. Figure 26 presents a subgroup analysis comparing SGLT2 versus non-SGLT2 administration for non-fatal stroke risk. In patients who did not receive SGLT2 inhibitors, the log risk-ratio (RR) ranged from −0.02 (95% CI: −0.15, 0.11) to 0.49 (95% CI: 0.01, 0.96), with an overall RR of 0.13 (95% CI: −0.01, 0.28). For patients who received SGLT2 inhibitors, the RR varied from −0.29 (95% CI: −1.03, 0.46) to −0.14 (95% CI: −0.56, 0.49), with an overall RR of −0.14 (95% CI: −0.30, −0.11). The overall RR for both groups was 0.04 (95% CI: −0.08, 0.16), with significant group differences (p = 0.02). These results suggest a slight increase in non-fatal stroke risk with nonSGLT2 inhibitors compared to SGLT2 inhibitors, though the overall effect remains modest.

**Figure 25.**
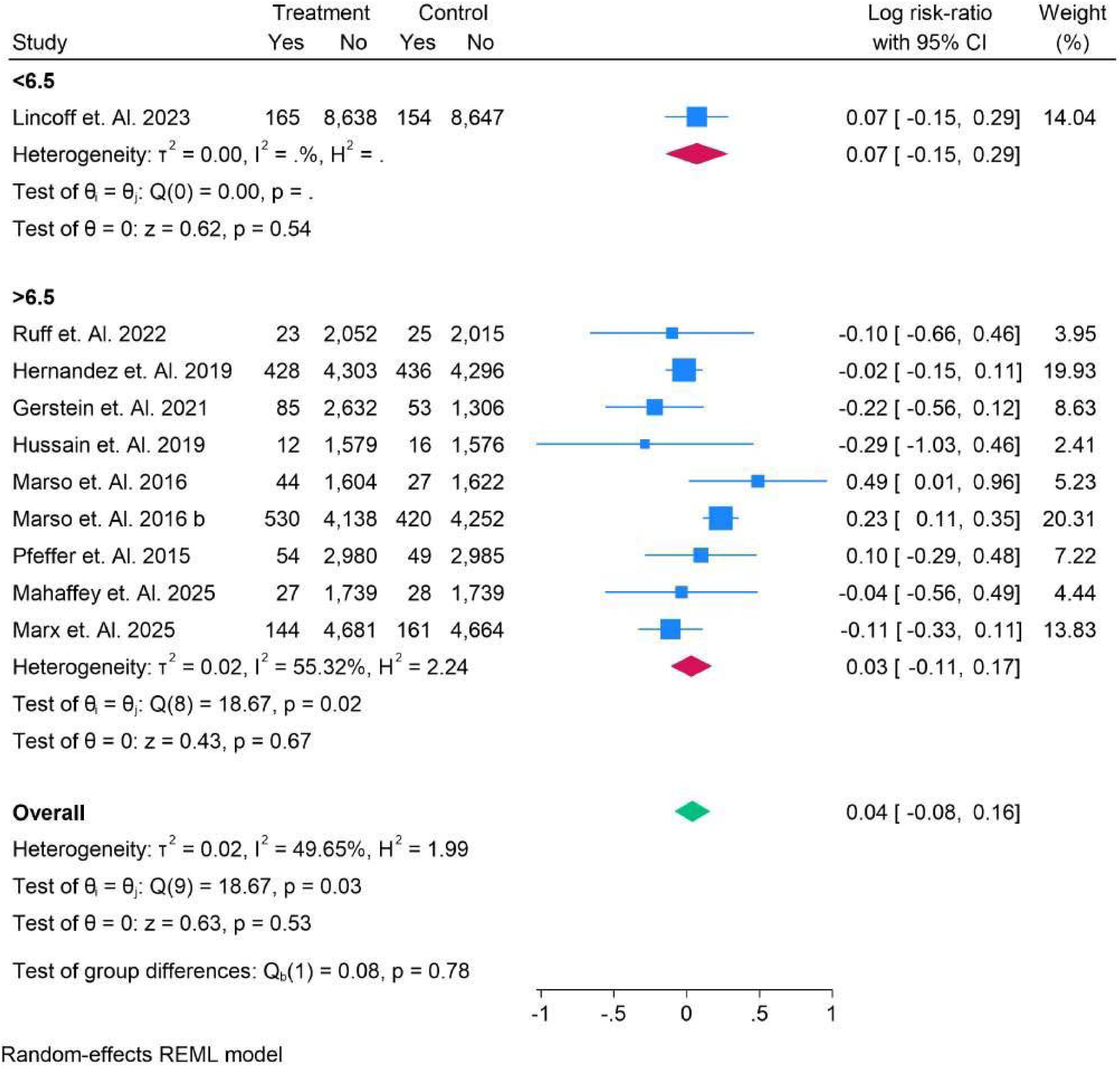
HBAl C in Sub-Analysis in Non-Fatal Stroke.

**Figure 26.**
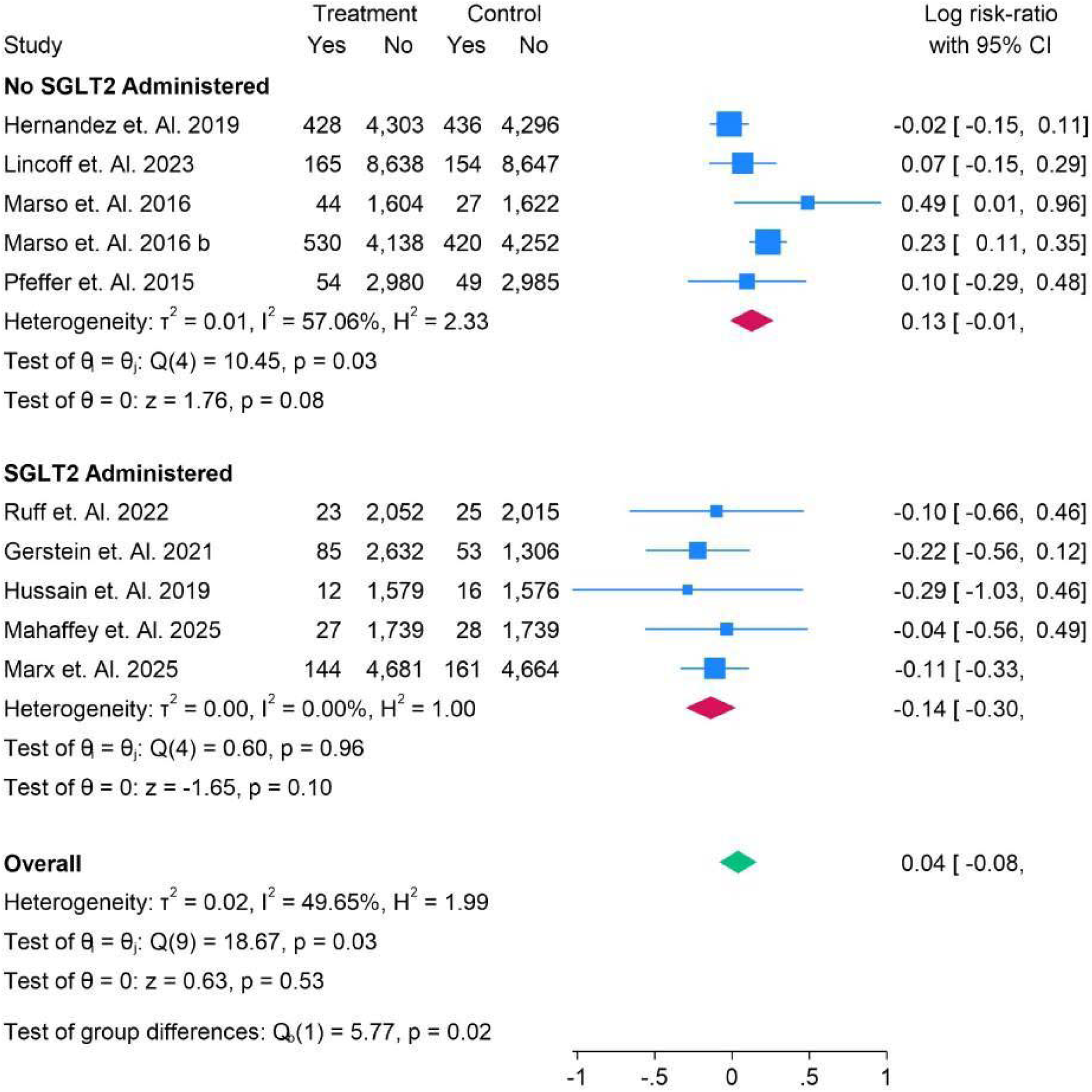
SGLT2 vs Non-SGLT2 in Sub-Analysis in Non-Fatal Stroke.

Figure 27 presents a subgroup analysis comparing daily versus weekly administration for non-fatal stroke risk. In the daily administration group, the log risk-ratio (RR) ranged from −0.29 (95% CI: −1.03, 0.46) to 0.10 (95% CI: −0.29, 0.48), with an overall RR of −0.08 (95% CI: −0.25, 0.10). In the weekly administration group, the RR varied from −0.04 (95% CI: −0.56, 0.49) to 0.49 (95% CI: 0.01, 0.96), with an overall RR of 0.08 (95% CI: −0.07, 0.23). The overall RR for both subgroups was 0.04 (95% CI: −0.08, 0.16), with no significant group differences (p = 0.19). These results suggest minimal difference in non-fatal stroke risk between daily and weekly administration. Figure 28 presents a subgroup analysis comparing oral versus subcutaneous (SC) administration for non-fatal stroke risk. In the **oral** administration group, the log risk-ratio (RR) ranged from − 0.29 (95% CI: 1.03, 0.46) to −0.11 (95% CI: −0.33, 0.11), with an overall RR of −0.13 (95% CI: −0.34, 0.09). For **SC** administration, the RR ranged from −0.04 (95% CI: −0.56, 0.49) to 0.49 (95% CI: 0.01, 0.96), with an overall RR of 0.07 (95% CI: −0.06, 0.20). The overall RR for both subgroups was 0.04 (95% CI: −0.08, 0.16), with no significant group differences (p = 0.12). These results suggest minimal difference in non-fatal stroke risk between oral and SC administration.

**Figure 27.**
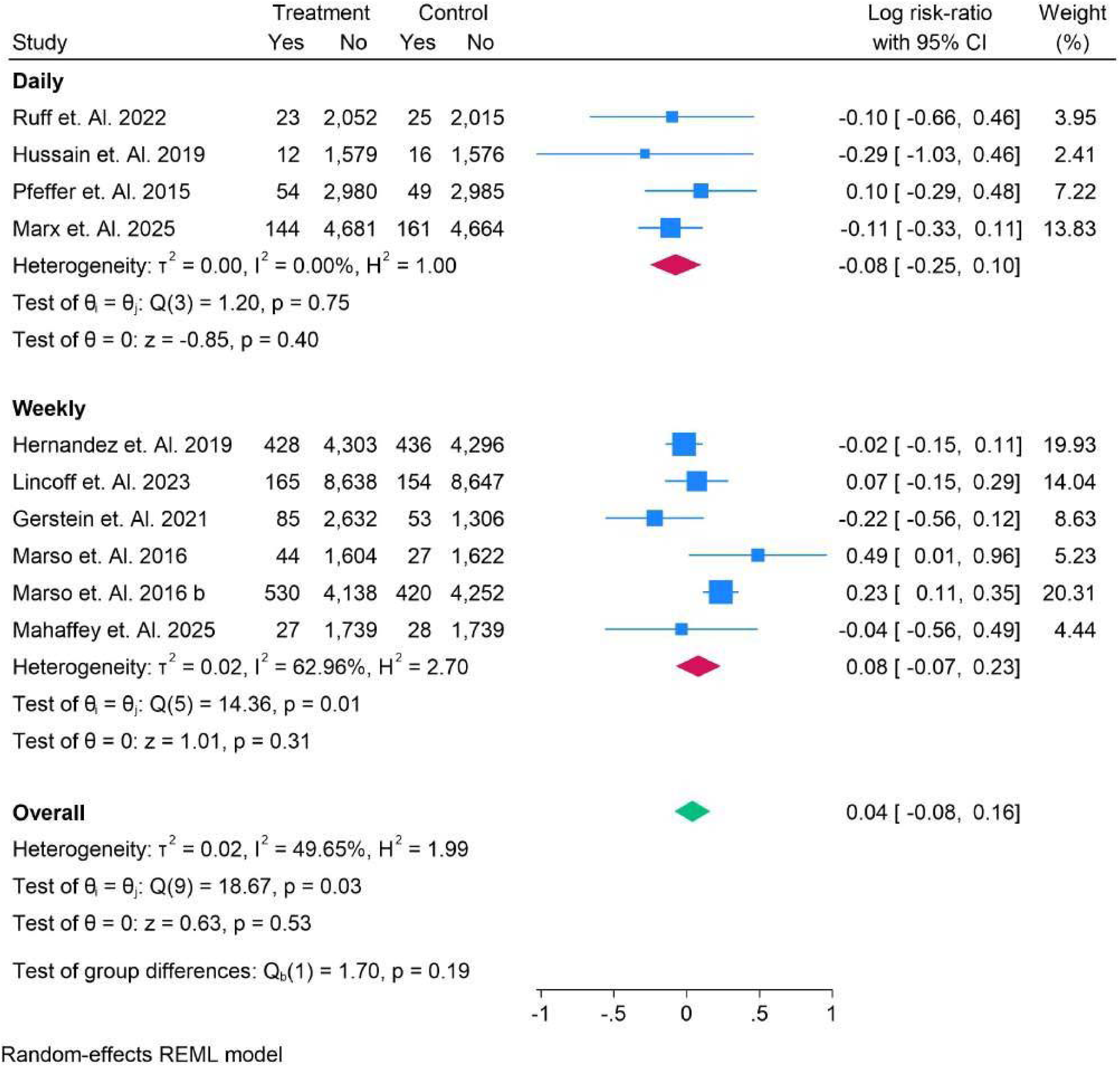
Daily vs Weekly in Sub-Analysis in Non-Fatal Stroke.

**Figure 28.**
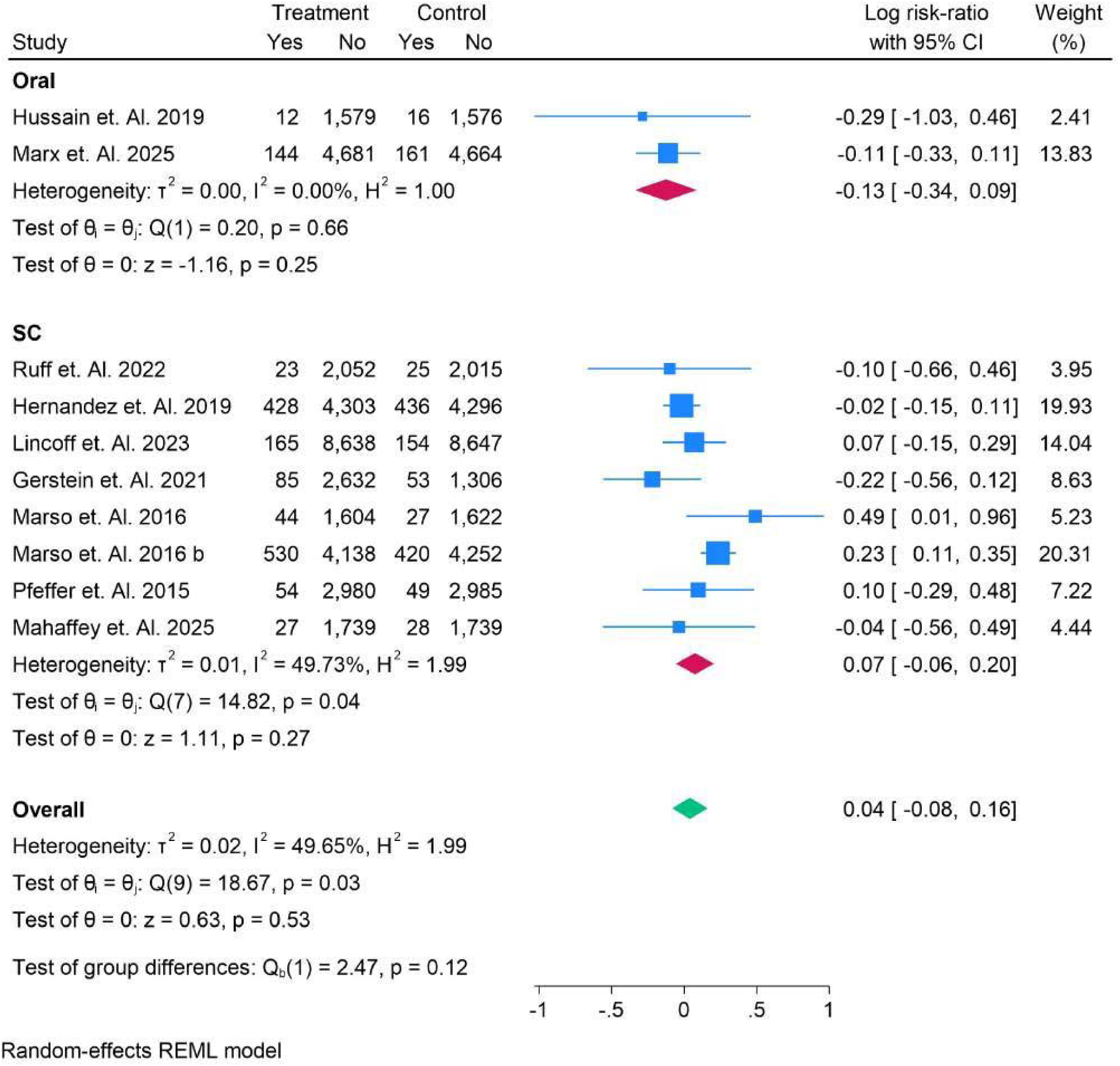
Oral vs Sub-Cutaneous in Sub-Analysis in Non-Fatal Stroke.

Figure 29 presents a subgroup analysis of various drugs for non-fatal stroke risk. For **Albiglutide**, the log riskratio (RR) was −0.02 (95% CI: −0.15, 0.11), showing no significant effect. **Efpeglenatide** showed an RR of 0.22 (95% CI: −0.56, 0.12), also with no significant effect. **Exenatide** had an RR of −0.10 (95% CI: −0.66, 0.46), indicating no clear effect. **Liraglutide** showed a favorable RR of 0.23 (95% CI: 0.11, 0.35), suggesting some potential benefit. **Lixisenatide** demonstrated an RR of 0.10 (95% CI: −0.29, 0.48), indicating no significant effect. **Semaglutide** showed a range from −0.29 (95% CI: −1.03, 0.46) to 0.49 (95% CI: 0.01, 0.96), with an overall RR of 0.03 (95% CI: −0.16, 0.21). The overall RR for all drugs was 0.04 (95% CI: −0.08, 0.16), with significant heterogeneity (I² = 49.65%) and a test for group differences p-value of 0.03. This suggests slight variation in non-fatal stroke risk across the different drugs, but no significant effect overall.

**Figure 29.**
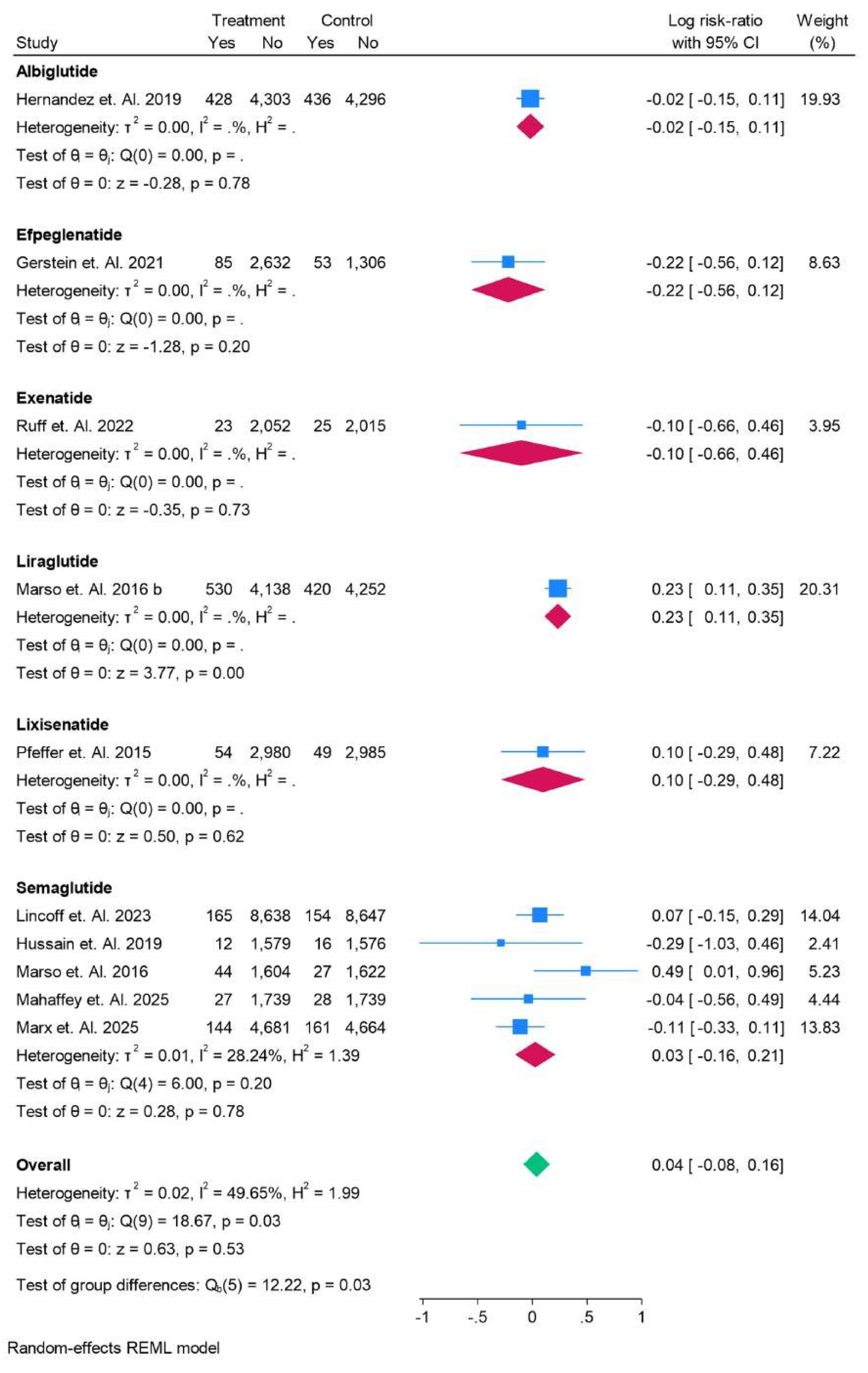
All Drugs in Sub-Analysis in Non-Fatal Stroke.

### Fatal Stroke

Figure 30 presents a subgroup analysis based on age for fatal stroke risk. In patients aged <65 years, the log risk-ratio (RR) ranged from −0.59 (95% CI: −0.80, −0.37) to 0.17 (95% CI: −0.63, 0.97), with an overall RR of 0.37 (95% CI: −0.66, −0.08), indicating a modest reduction in fatal stroke risk. In patients aged >65 years, the RR was −0.33 (95% CI: −0.93, 0.28), showing no significant effect. The overall RR for both age groups was 0.38 (95% CI: −0.61, −0.15), with no significant group differences (p = 0.89), suggesting that age did not significantly affect the fatal stroke risk across the subgroups. Figure 31 presents a subgroup analysis based on race for fatal stroke risk. In **Asian** patients, the log risk-ratio (RR) was −0.30 (95% CI: −0.49, −0.11), indicating a modest reduction in fatal stroke risk. For **White** patients, the RR ranged from −0.59 (95% CI: −0.80, −0.37) to 0.17 (95% CI: −0.63, 0.97), with an overall RR of −0.39 (95% CI: −0.77, −0.01), showing a potential reduction in risk. The overall RR across both racial groups was −0.38 (95% CI: −0.61, −0.15), with no significant group differences (p = 0.67), suggesting minimal variation in fatal stroke risk across races. Figure 32 presents a subgroup analysis of type 2 diabetes (T2DM) for fatal stroke risk. In T2DM patients, the log risk-ratio (RR) ranged from −0.59 (95% CI: −0.80, −0.37) to 0.17 (95% CI: −0.63, 0.97), with an overall RR of −0.38 (95% CI: 0.61, −0.15). The overall RR for T2DM patients across studies were −0.38 (95% CI: −0.61, −0.15), with no significant differences across the subgroup (p = 0.00). These results suggest a reduction in fatal stroke risk among T2DM patients, with statistically significant results across the studies. Figure 33 presents a subgroup analysis comparing normal versus type 2 diabetes (T2DM) for fatal stroke risk, categorized by BMI. In patients with BMI <30, the log risk-ratio (RR) was 0.17 (95% CI: −0.63, 0.97), showing no significant effect. In patients with BMI >30, the RR ranged from −0.59 (95% CI: −0.80, −0.37) to −0.30 (95% CI: −0.49, −0.11), with an overall RR of −0.42 (95% CI: − 0.65, −0.20), suggesting a reduction in fatal stroke risk in this group. The overall RR for both subgroups was −0.38 (95% CI: −0.61, −0.15), with no significant differences between groups (p = 0.16), indicating that BMI may not significantly affect fatal stroke risk in patients with T2DM. Figure 34 presents a subgroup analysis based on BMI for fatal stroke risk. In patients with a BMI <30, the log risk-ratio (RR) was 0.17 (95% CI: −0.63, 0.97), showing no significant effect. For patients with BMI >30, the RR ranged from −0.59 (95% CI: −0.80, −0.37) to −0.30 (95% CI: −0.49, −0.11), with an overall RR of −0.42 (95% CI: −0.65, −0.20), suggesting a reduction in fatal stroke risk. The overall RR for both subgroups was −0.38 (95% CI: −0.61, −0.15), with no significant differences between groups (p = 0.16), indicating minimal effect of BMI on fatal stroke risk across groups. Figure 35 presents the HBA1C subgroup analysis for fatal stroke risk. In patients with HBA1C levels >6.5, the log risk-ratio (RR) ranged from −0.59 (95% CI: −0.80, −0.37) to 0.17 (95% CI: −0.63, 0.97), with an overall RR of −0.38 (95% CI: −0.61, −0.15), suggesting a reduction in fatal stroke risk. The overall RR across all studies for HBA1C >6.5 was −0.38 (95% CI: −0.61, −0.15), with no significant group differences (p = 1.00), indicating that HBA1C level did not significantly impact the fatal stroke risk across the subgroup. Figure 36 presents the sub-analysis of fatal stroke risk in patients who did not receive SGLT2 inhibitors and those who did. In patients without SGLT2 administration, the log risk-ratio (RR) ranged from −0.59 (95% CI: −0.80, −0.37) to 0.17 (95% CI: −0.63, 0.97), with an overall RR of −0.39 (95% CI: −0.77, −0.01), suggesting no significant impact on fatal stroke risk. In patients who received SGLT2 inhibitors, the RR was −0.30 (95% CI: −0.49, −0.11), indicating a reduction in fatal stroke risk. The overall RR for both subgroups was −0.38 (95% CI: −0.61, −0.15), with no significant group differences (p = 0.67), indicating no clear advantage of SGLT2 use in reducing fatal stroke risk.

**Figure 30.**
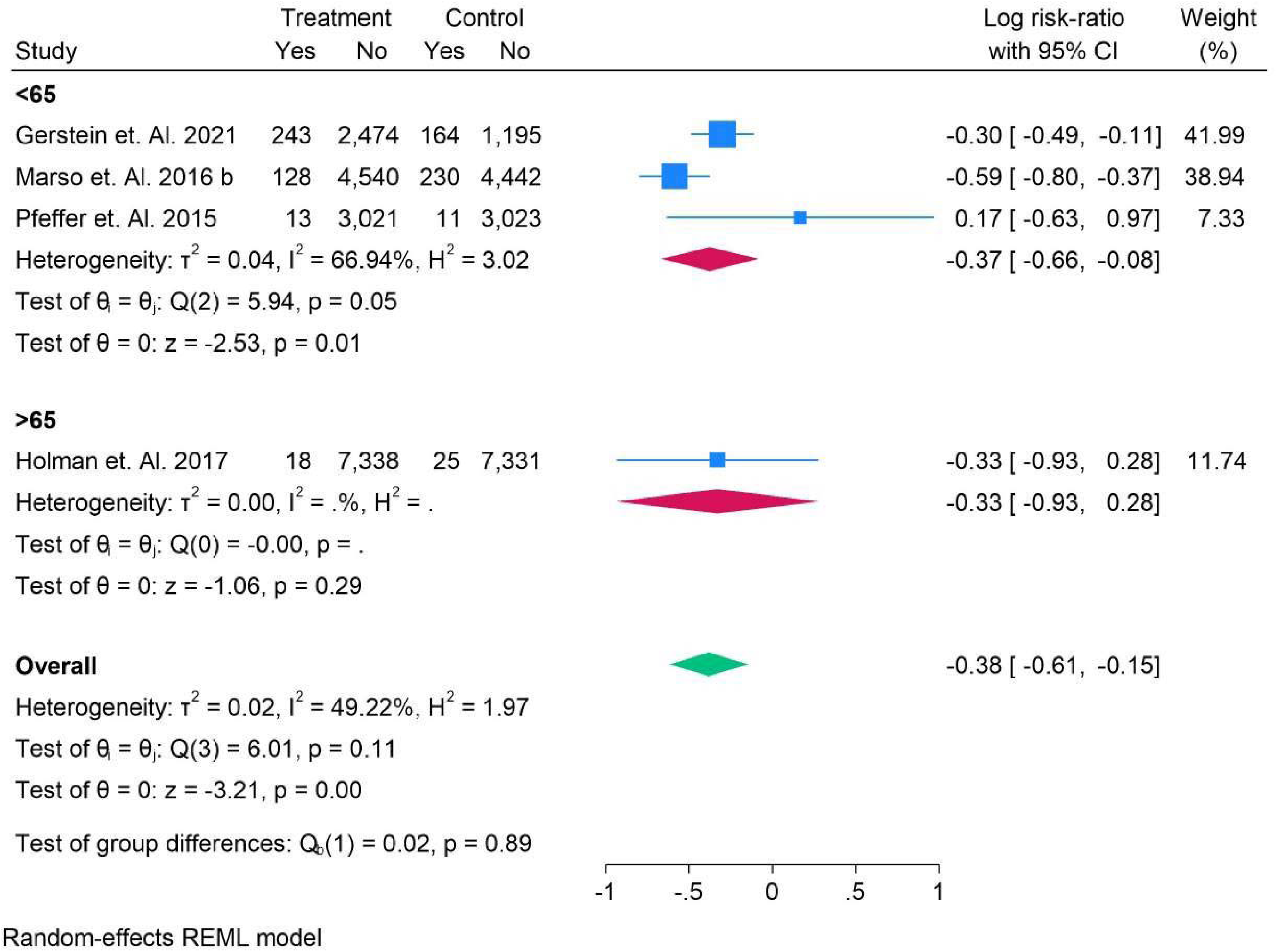
Age in Sub-Analysis in Fatal Stroke.

**Figure 31.**
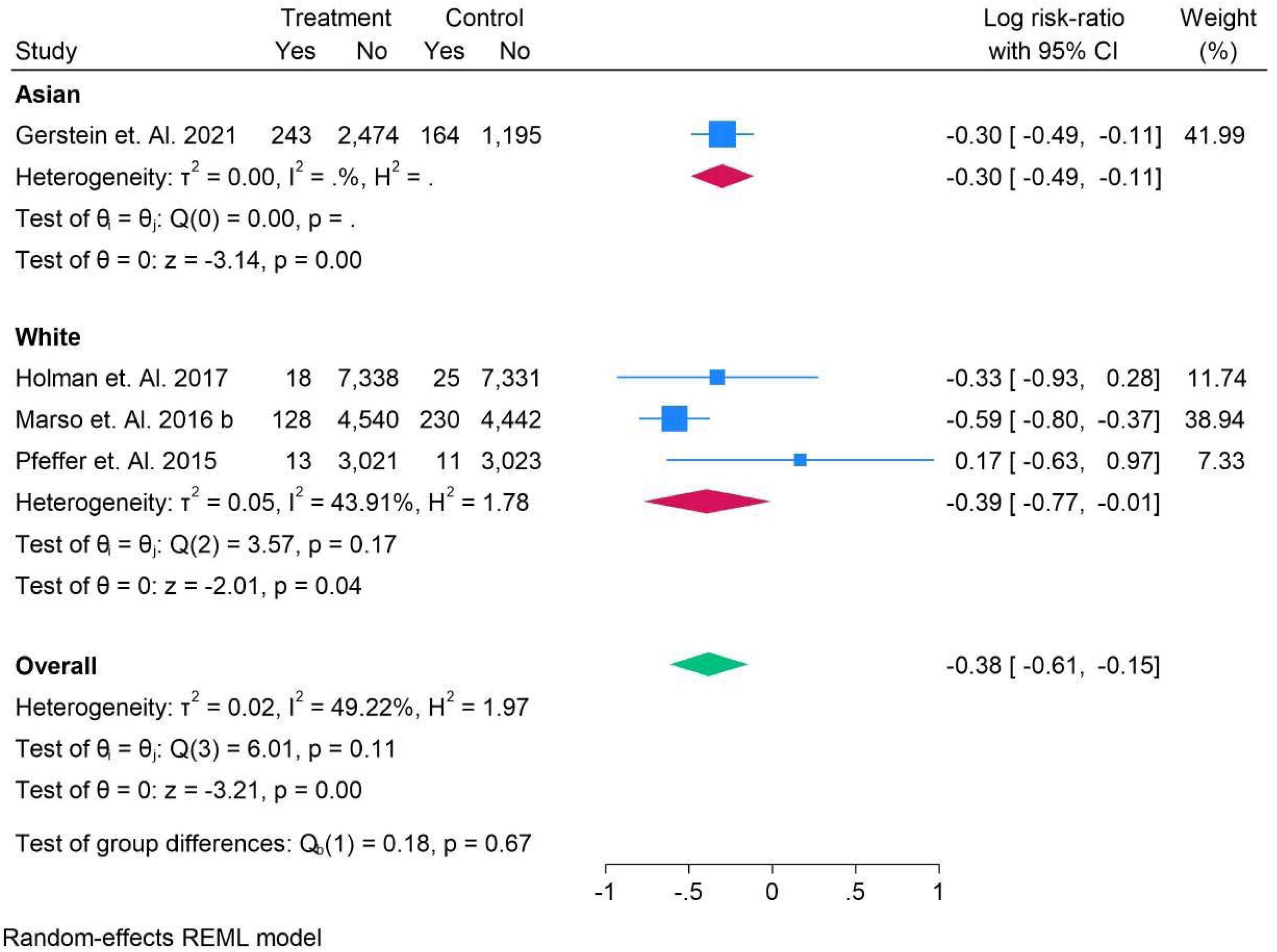
RACE in Sub-Analysis in Fatal Stroke.

**Figure 32.**
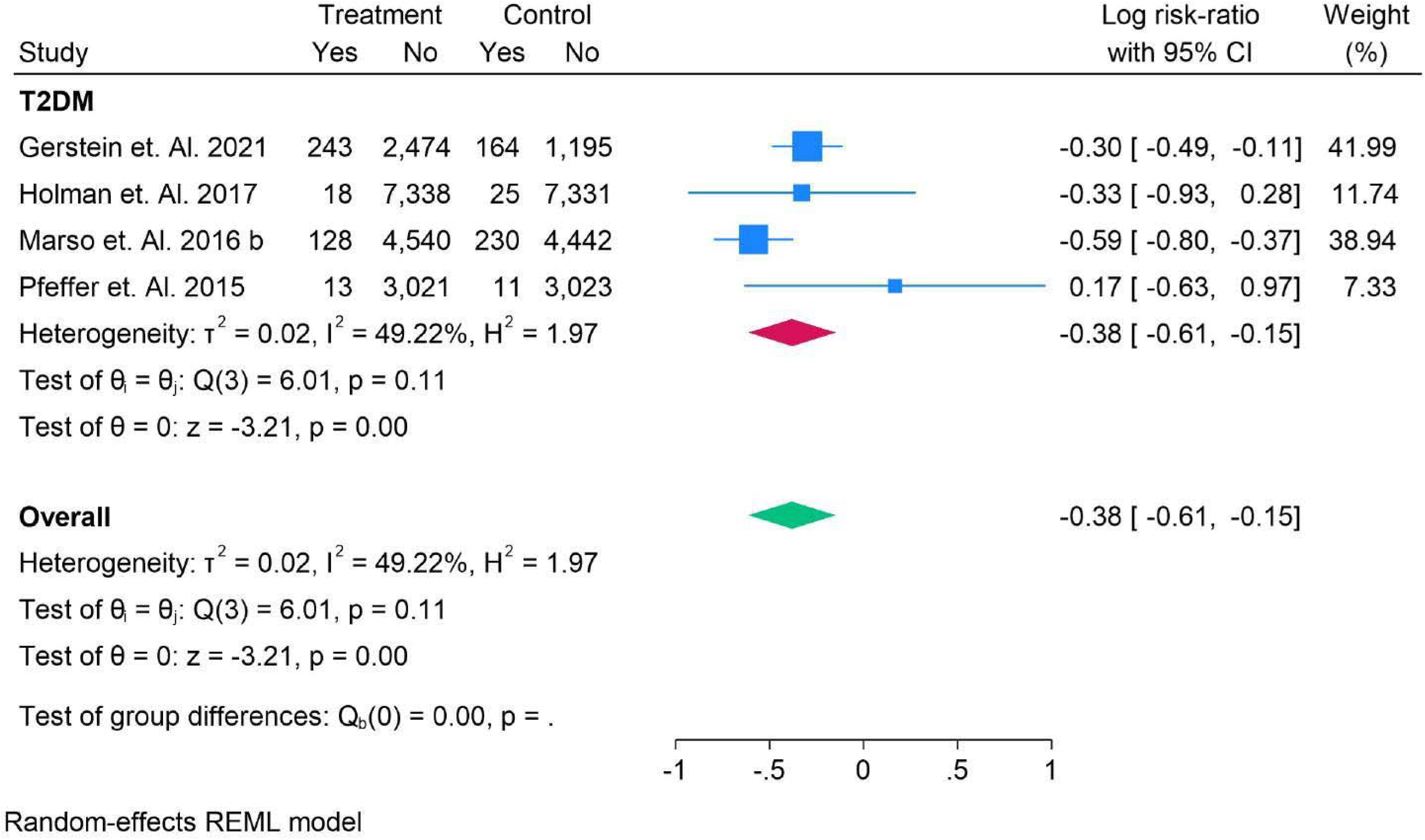
T2DM in Sub-Analysis in Fatal Stroke.

**Figure 33.**
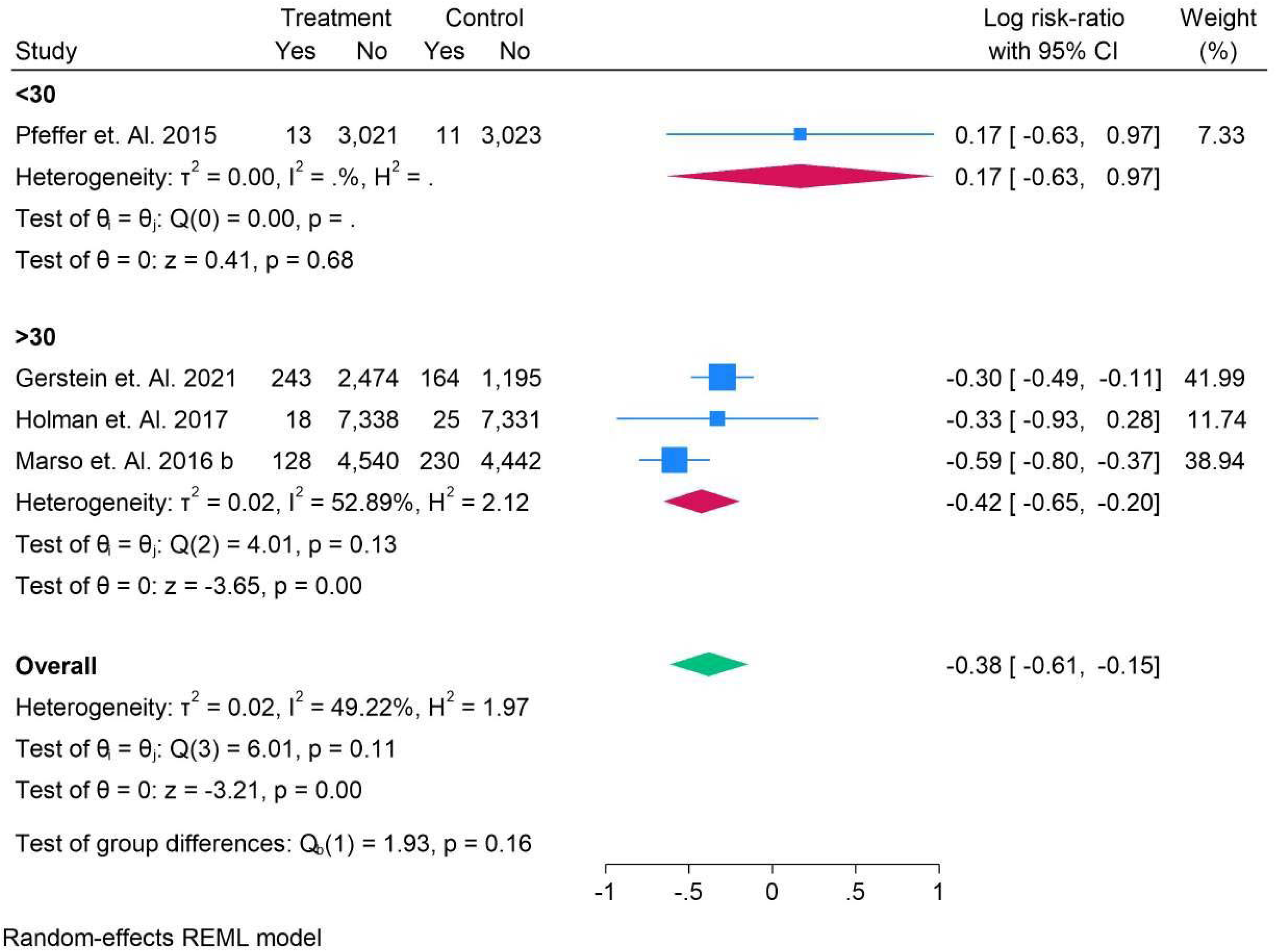
Normal vs T2DM in Sub-Analysis in Fatal Stroke.

**Figure 34.**
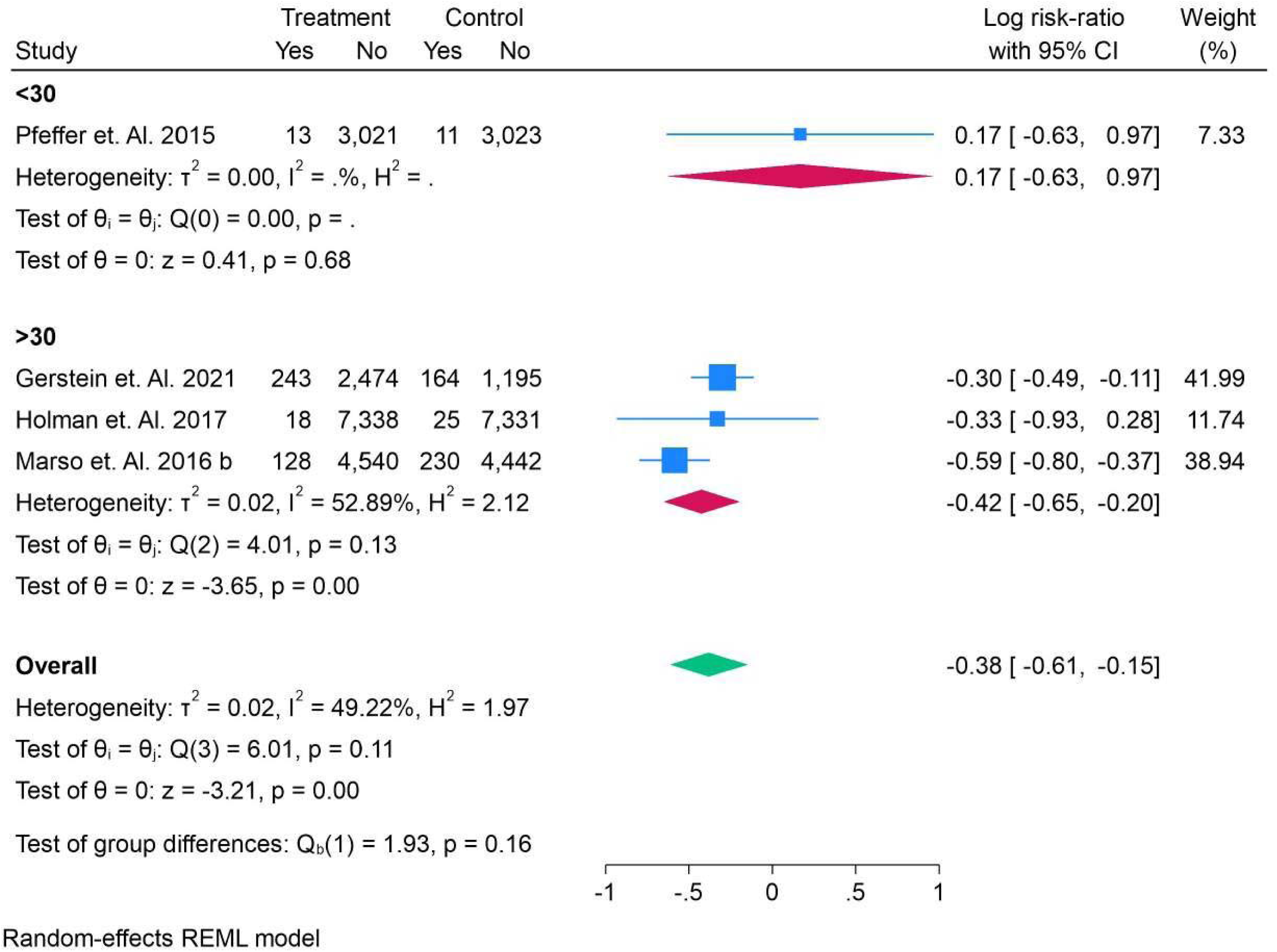
BMI - Sub-Analysis in Fatal Stroke.

**Figure 35.**
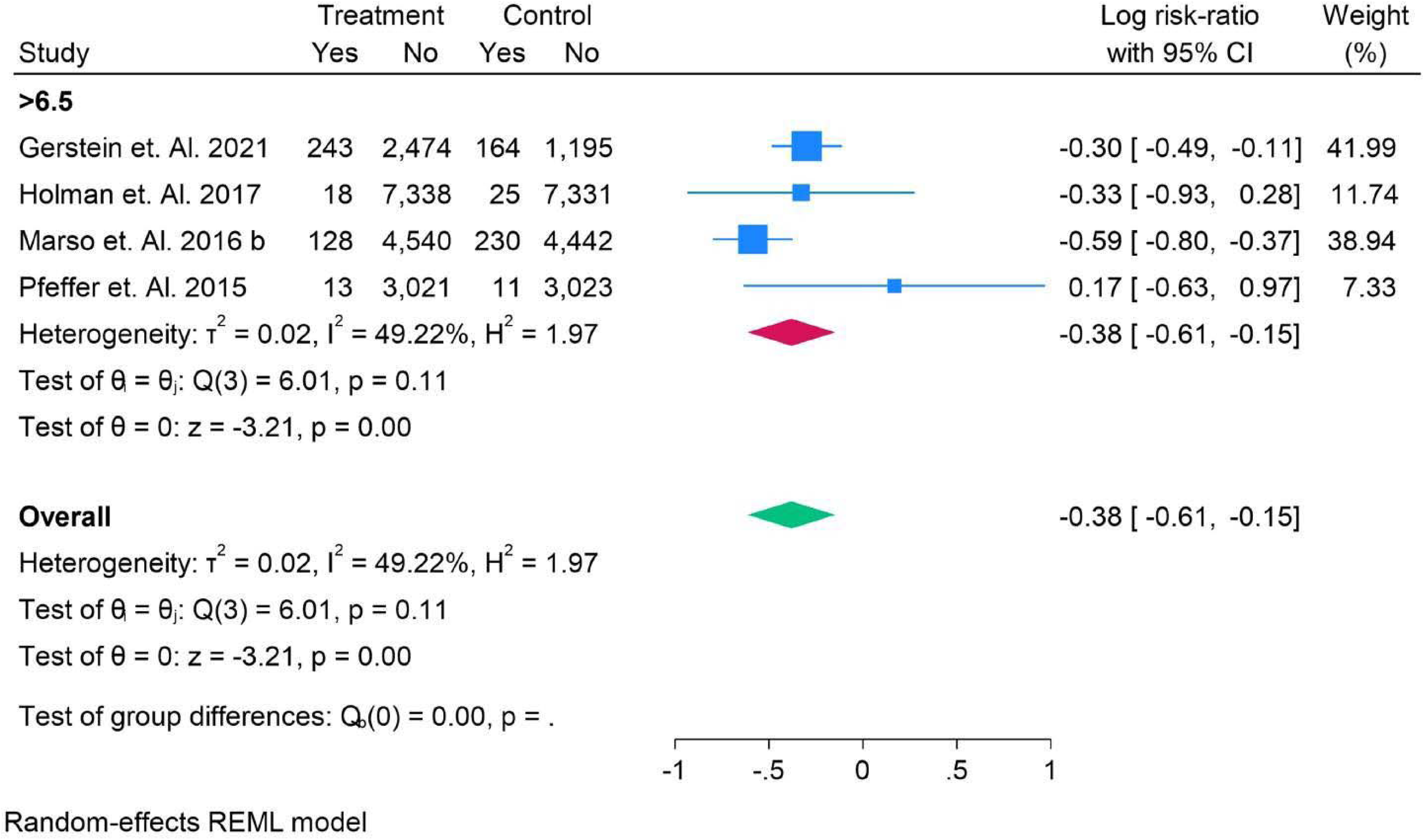
HBAl C - Sub-Analysis in Fatal Stroke.

**Figure 36.**
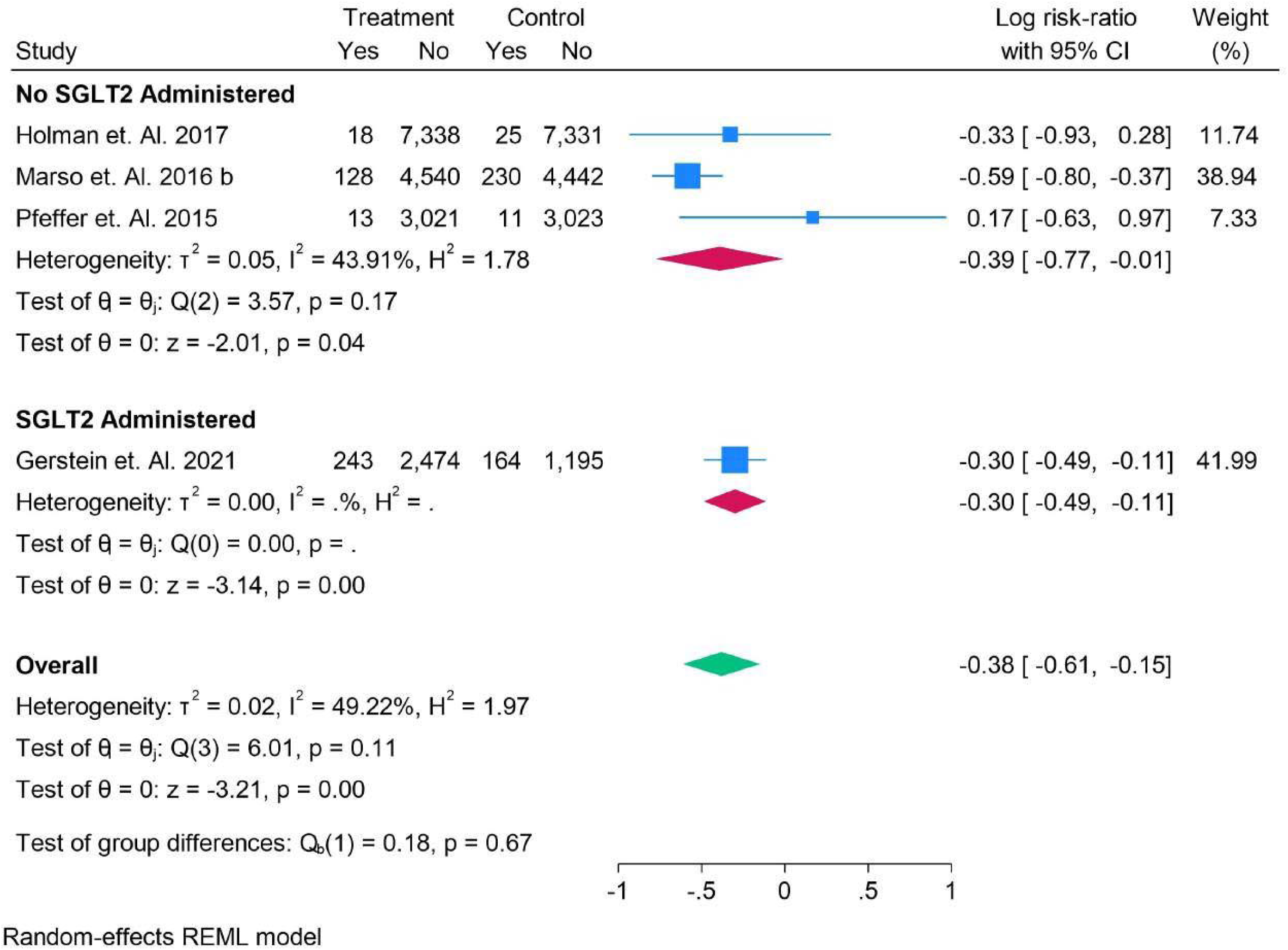
No SGLT2 and SGLT2 Administration - Sub-Analysis in Fatal Stroke.

Figure 37 presents the sub-analysis comparing weekly versus daily dosage in patients with fatal stroke. The data show that patients on weekly dosages had a significant log risk-ratio (RR) of −0.42 (95% CI: −0.65, −0.20), while patients on daily dosages showed no significant change (RR: 0.17, 95% CI: −0.63, 0.97). The overall RR for both groups was −0.38 (95% CI: −0.61, −0.15), but the test of group differences (p = 0.16) suggests no significant difference between the two dosage frequencies. Figure 38 shows the sub-analysis of subcutaneous (SC) dosage in fatal stroke patients. The results indicate that SC administration had a log risk-ratio (RR) of −0.38 (95% CI: − 0.61, −0.15), suggesting a moderate effect. The overall RR across all studies was also −0.38 (95% CI: −0.61, 0.15), with no significant group differences (p = 1.00). The analysis presents a moderate heterogeneity (I² = 49.22%). Figure 39 shows the analysis of all drugs used in treating fatal stroke. The overall log risk-ratio (RR) is −0.38 (95% CI: −0.61, −0.15), indicating a modest beneficial effect. Among individual drugs, liraglutide (RR: 0.59, 95% CI: −0.80, −0.37) showed the strongest effect, while lixisenatide (RR: 0.17, 95% CI: −0.63, 0.97) had no significant impact. There was moderate heterogeneity (I² = 49.22%) across the studies. Group differences were not significant (p = 0.11).

**Figure 37.**
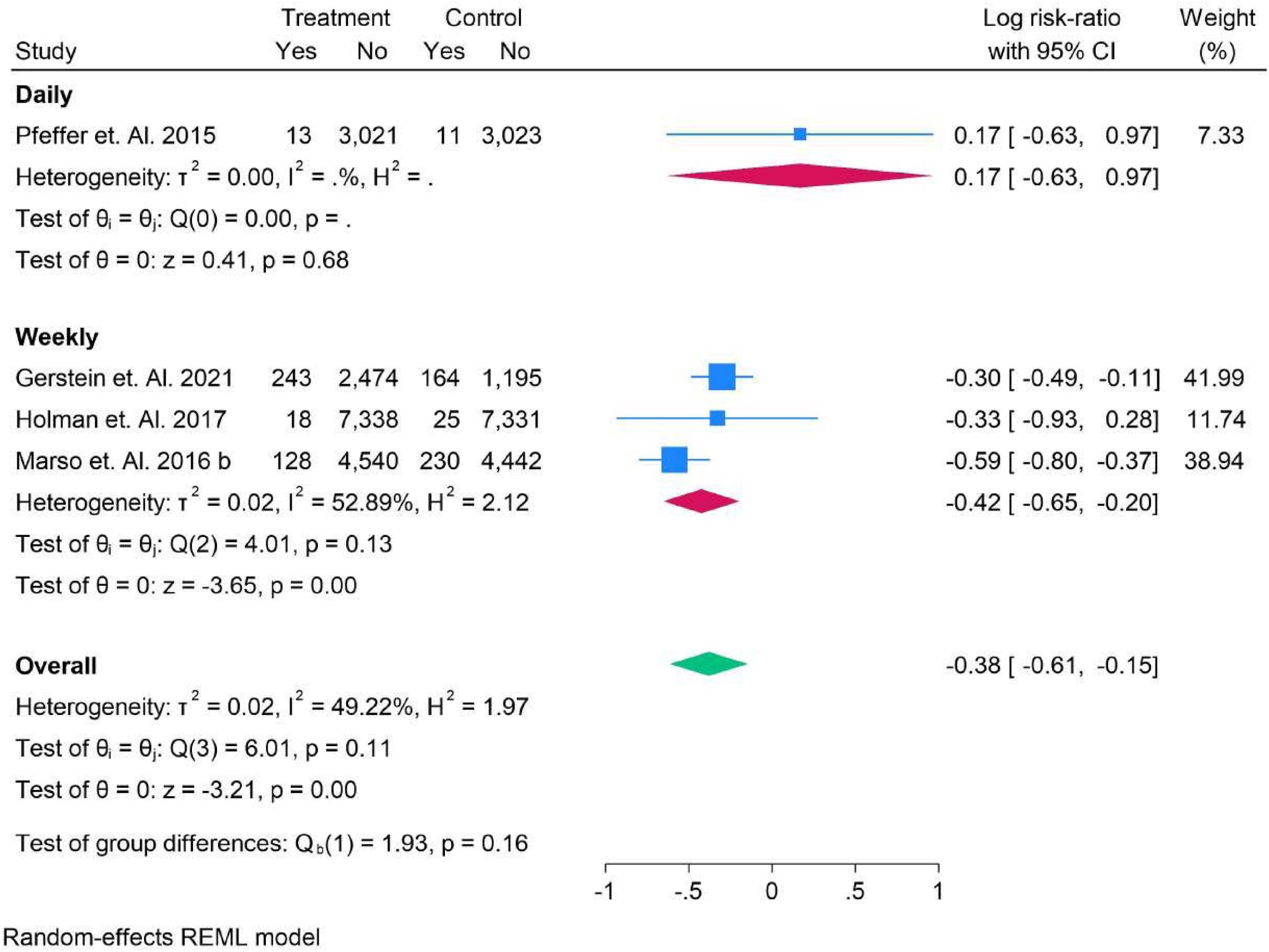
Weekly vs Daily Dosage- Sub-Analysis in Fatal Stroke.

**Figure 38.**
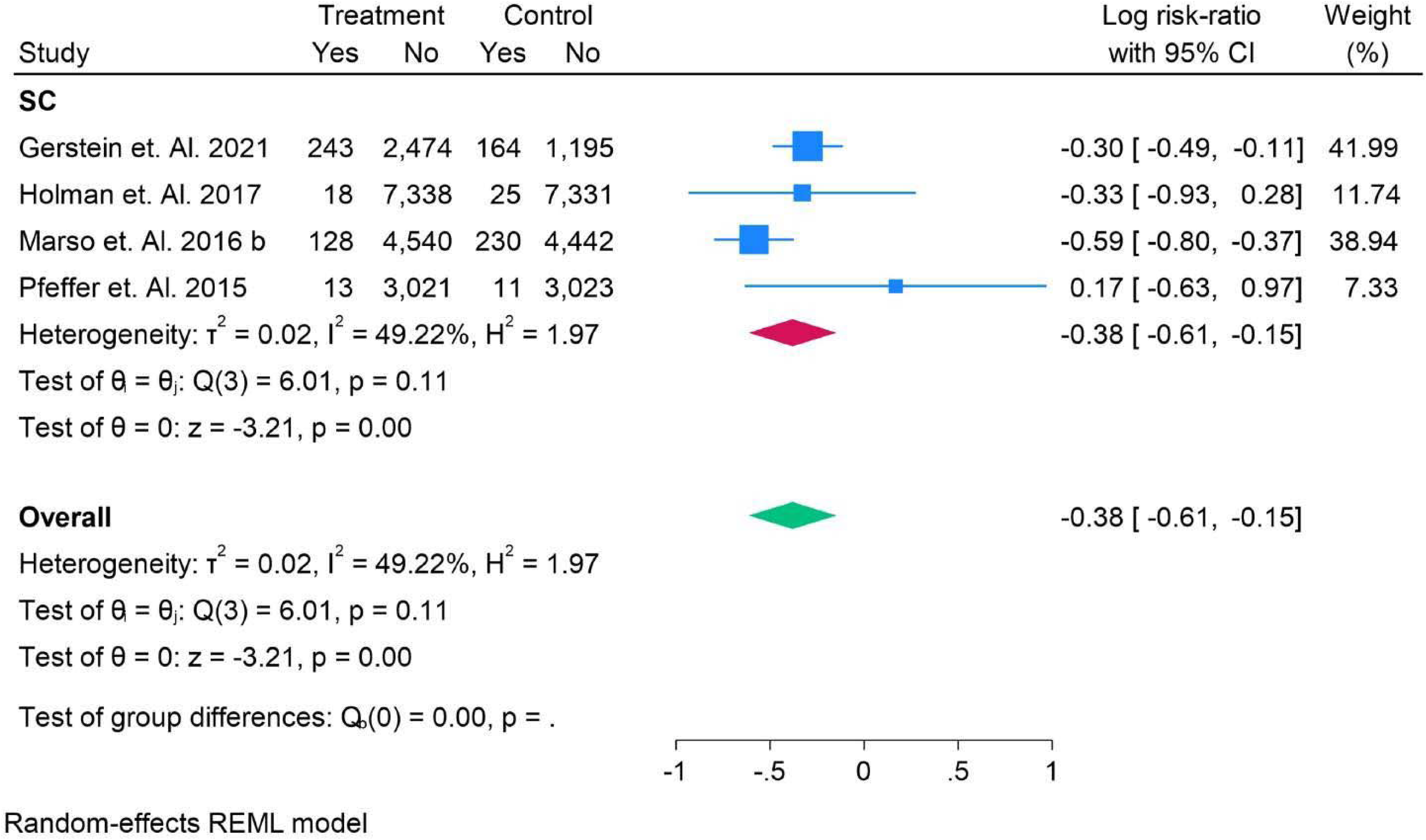
Sub-Cutaneous Dosage Analysis in Fatal Stroke.

**Figure 39.**
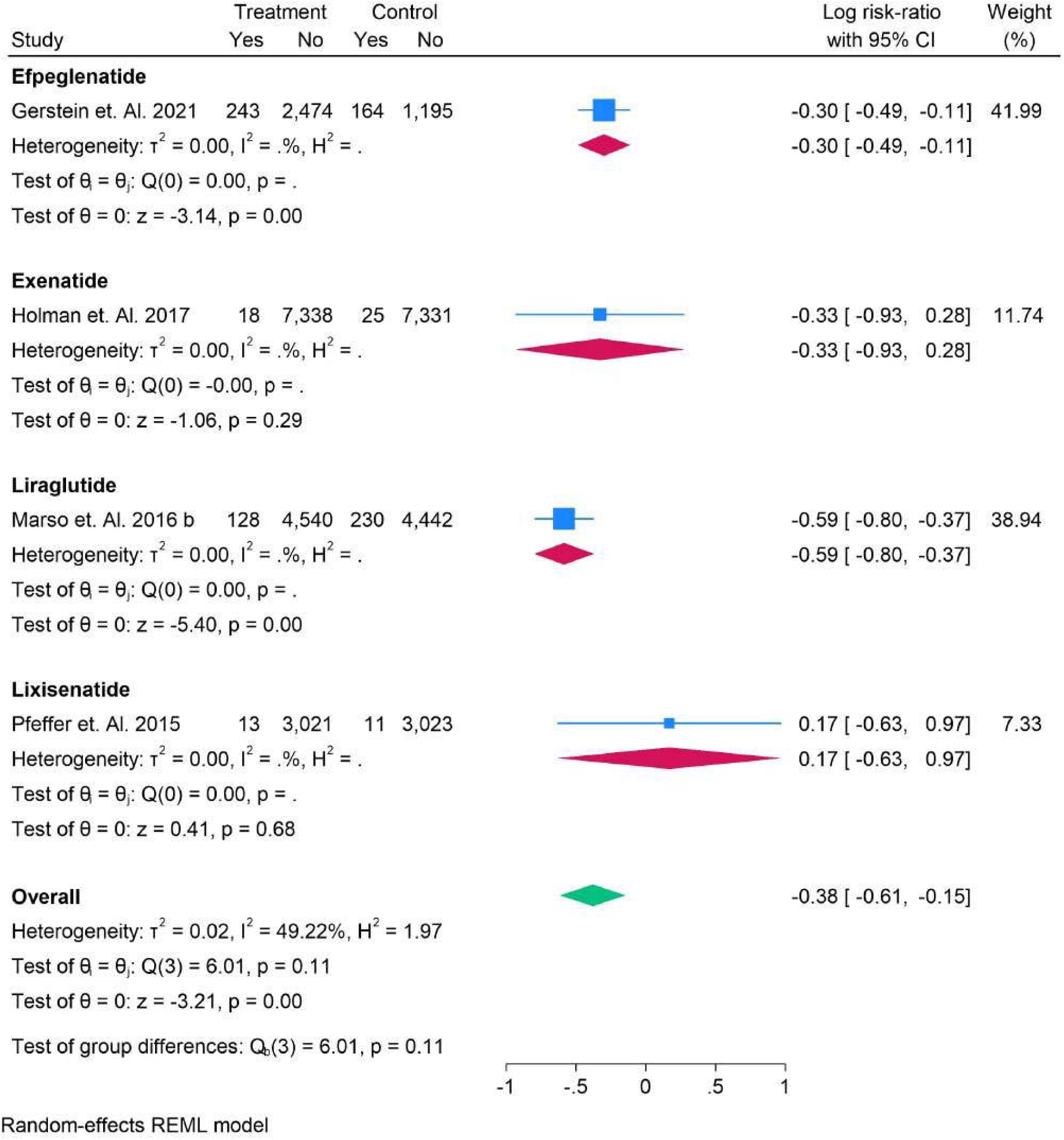
All Drugs Analysis in Fatal Stroke.

### Renal Outcomes

Figure 40 displays the network plot and comparison of different drugs to placebo concerning renal outcomes, specifically changes in eGFR. The Mean Differences (MD) with their corresponding 95% Confidence Intervals (CI) for each drug compared to placebo are as follows: Efpeglenatide showed a MD of 1.30 with a 95% CI of 0.84 to 3.44, indicating a wide range of potential outcomes. Exenatide had a MD of 2.00 (95% CI: −0.14 to 4.14), suggesting a more modest but still variable effect. Liraglutide’s MD was 1.20 (95% CI: −0.93 to 3.33), demonstrating a similar range of variability. Semaglutide exhibited a MD of 1.26 (95% CI: 0.00 to 2.51), with the CI indicating a more consistent effect, whereas Tirzepatide showed the largest MD of 2.50 (95% CI: 0.36 to 4.64), suggesting the most significant impact on eGFR improvement. These results highlight the variability in renal outcomes for each treatment compared to placebo. Figure 41, The analysis of eGFR mean differences across various studies reveals notable variations in treatment effects. **Ruff et al. 2022** reported the largest improvement in eGFR, with a mean difference of 2.00 (95% CI: 1.91–2.09), indicating a substantial benefit of the treatment. This is in contrast to **Marx et al. 2025**, which reported a lower effect, with a mean difference of 0.80 (95% CI: 0.73–0.87). Other studies such as **Hussain et al. 2019** (2.40, 95% CI: 2.20–2.60) and **Gerstein et al. 2021** (1.30, 95% CI: 1.14–1.46) also demonstrated positive effects, with Hussain et al. showing the most significant improvement. Meanwhile, studies like **Packer et al. 2025** (1.20, 95% CI: 1.14–1.26) and **Marso et al. 2016 b** (1.20, 95% CI: 1.14–1.26) found more moderate but consistent positive changes in eGFR. The combined mean difference across all studies was 1.55 (95% CI: 0.99–2.12), suggesting a beneficial effect of the intervention. However, the high heterogeneity observed (I² = 99.43%) indicates significant variability in treatment outcomes, likely due to differences in study designs, populations, and treatment protocols. These findings underscore the need for further analysis to understand the underlying factors contributing to such variability.

**Figure 40.**
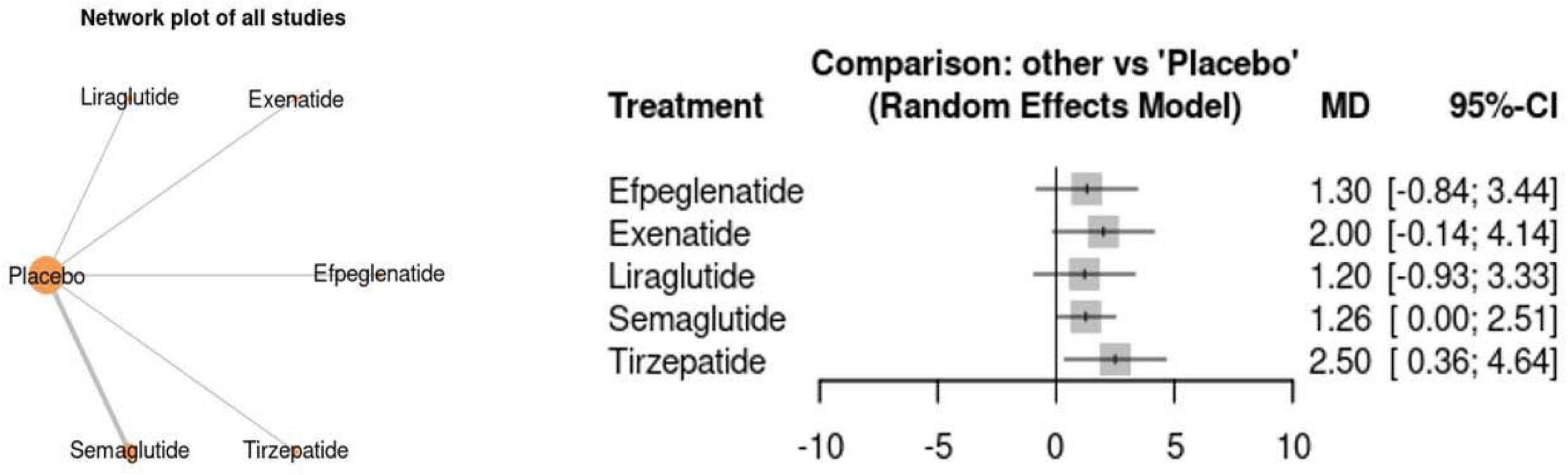
Renal Outcomes Network Graph and Comparison with Placebo in Mean Difference and CI, in eGFR.

**Figure 41.**
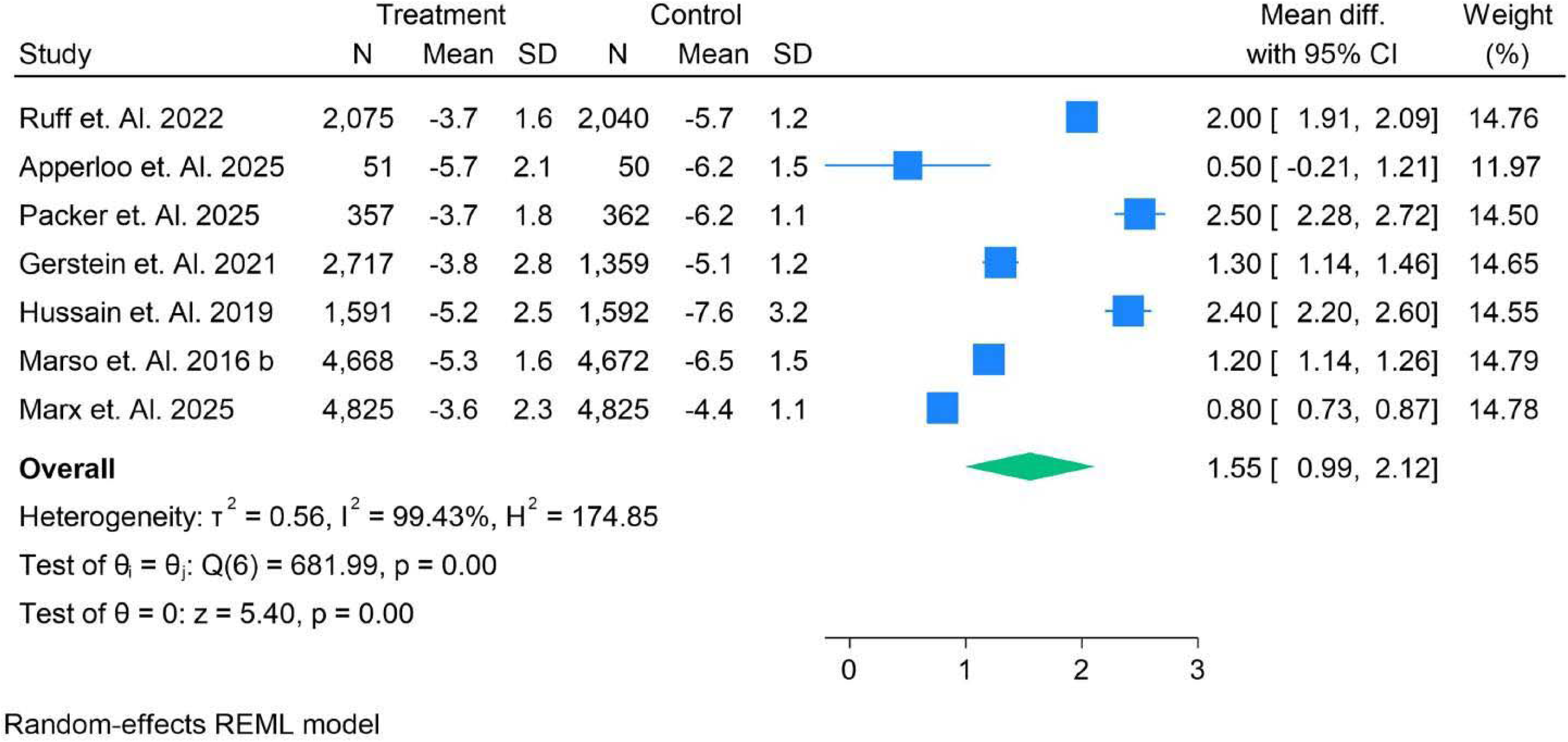
Mean Difference of eGFR in patients.

In patients under 65, studies like **Ruff et al. (2022)** showed a substantial improvement in eGFR (mean difference 2.00, 95% CI: 1.91–2.09), while **Gerstein et al. (2021)** reported a moderate effect (mean difference 1.30, 95% CI: 1.14–1.46). In patients over 65, **Packer et al. (2025)** observed a larger treatment effect (mean difference 2.50, 95% CI: 2.28–2.72), indicating that older patients may experience greater benefit. The overall mean difference across all studies was 1.55 (95% CI: 0.99–2.12), with high heterogeneity (I² = 99.43%), suggesting variability based on age and other factors. (Figure 42). In the **Asian** subgroup, **Gerstein et al. (2021)** showed a mean difference of 1.30 (95% CI: 1.14–1.46), indicating a moderate improvement in eGFR. In the **Hispanic** subgroup, **Hussain et al. (2019)** observed a significant difference of 2.40 (95% CI: 2.20–2.60). In **White** patients, **Marx et al. (2025)** reported a mean difference of 0.80 (95% CI: 0.73–0.87), while Packer et al. (2025) observed a higher mean difference of 2.50 (95% CI: 2.28–2.72). The overall mean difference for all groups was 1.55 (95% CI: 0.99–2.12), with significant heterogeneity. (Figure 43).

**Figure 42.**
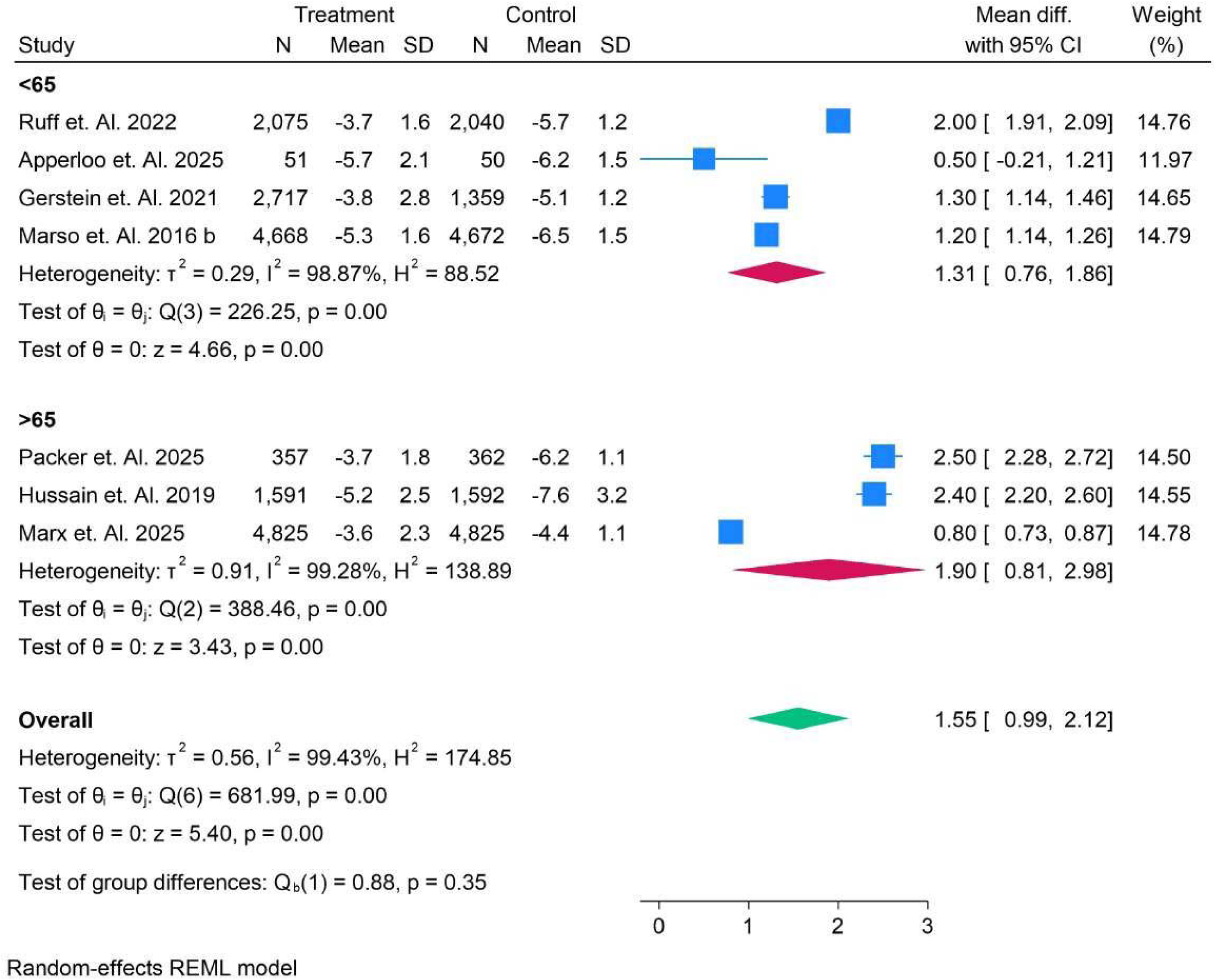
Mean Difference of eGFR in patients and Subgroup analysis of age.

**Figure 43.**
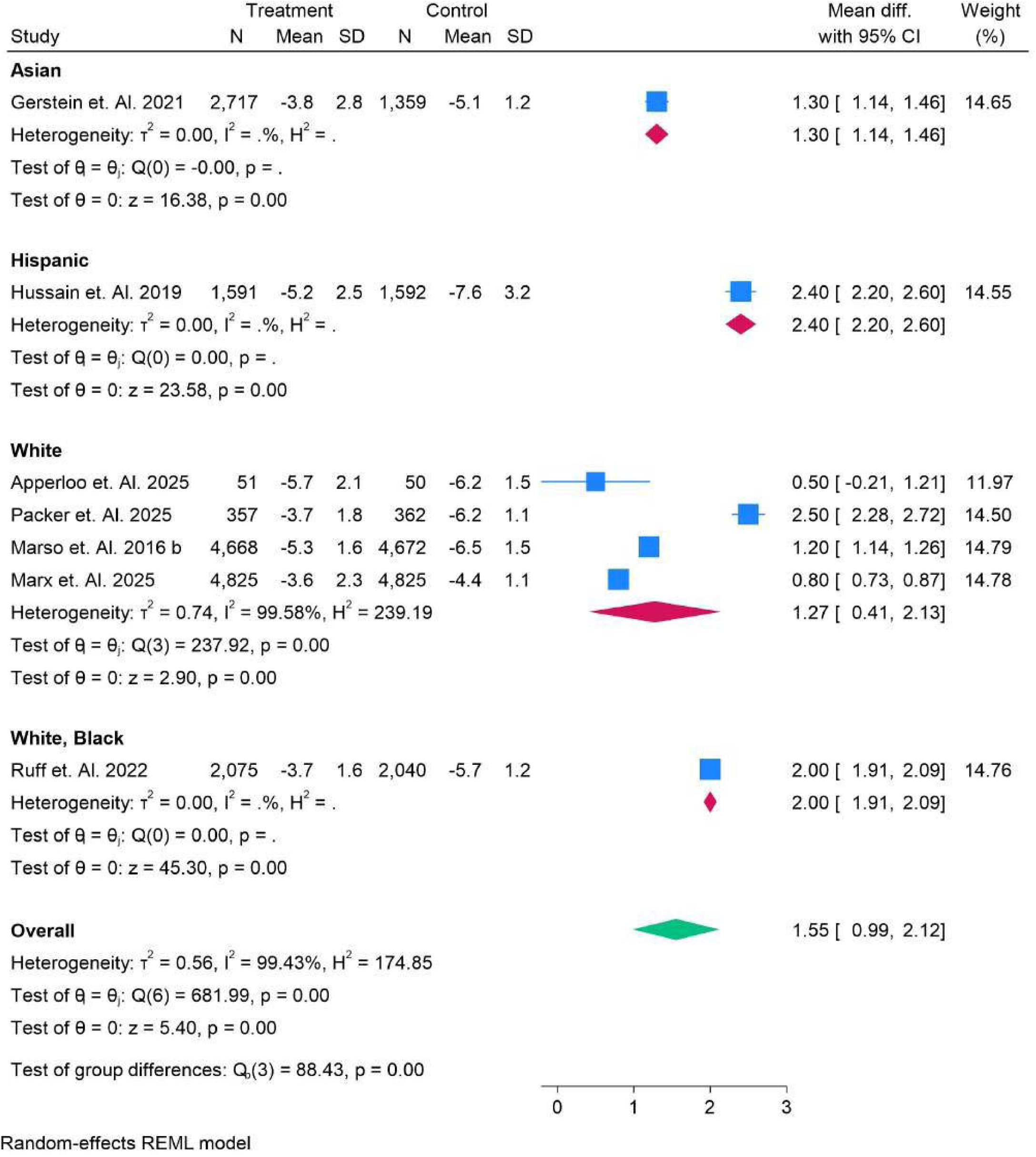
Mean Difference of eGFR in patients and Subgroup analysis ofRACE.

The **T2DM** subgroup analysis revealed a significant improvement in eGFR in patients. **Ruff et al. (2022)** reported a mean difference of 2.00 (95% CI: 1.91–2.09), indicating a notable effect. Gerstein et al. (2021) and Marx et al. (2025) observed smaller but significant differences of 1.30 (95% CI: 1.14–1.46) and 0.80 (95% CI: 0.73–0.87), respectively. The overall mean difference across all studies was 1.55 (95% CI: 0.99–2.12), highlighting the consistent improvement across the studies in T2DM patients. Significant heterogeneity was observed in this analysis, further emphasizing variability in treatment responses. (Figure 44)

**Figure 44.**
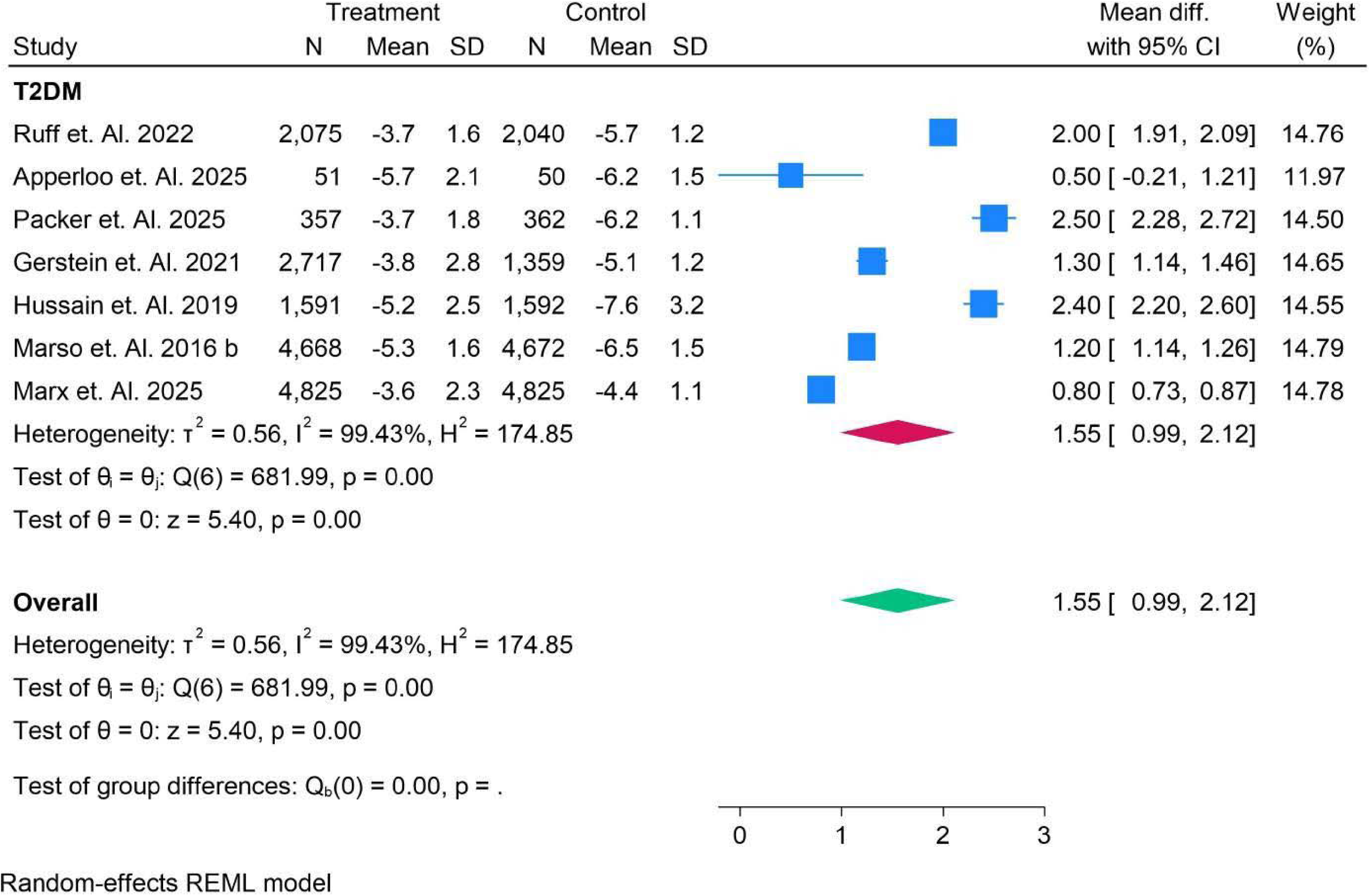
Mean Difference of eGFR in patients and Subgroup analysis of T2DM.

In the BMI subgroup analysis, patients with BMI < 30 had a notable improvement in eGFR. Ruff et al. (2022) and Marx et al. (2025) showed a mean difference of 2.00 (95% CI: 1.91–2.09) and 0.80 (95% CI: 0.73–0.87), respectively. In the >30 BMI group, Packer et al. (2025) and Gerstein et al. (2021) reported higher improvements in eGFR, with mean differences of 2.50 (95% CI: 2.28–2.72) and 1.30 (95% CI: 1.14–1.46). The overall mean difference across both groups was 1.55 (95% CI: 0.99–2.12), emphasizing improvements regardless of BMI category, with minimal heterogeneity (I² = 99.43%) in the analysis. (Figure 45).

**Figure 45.**
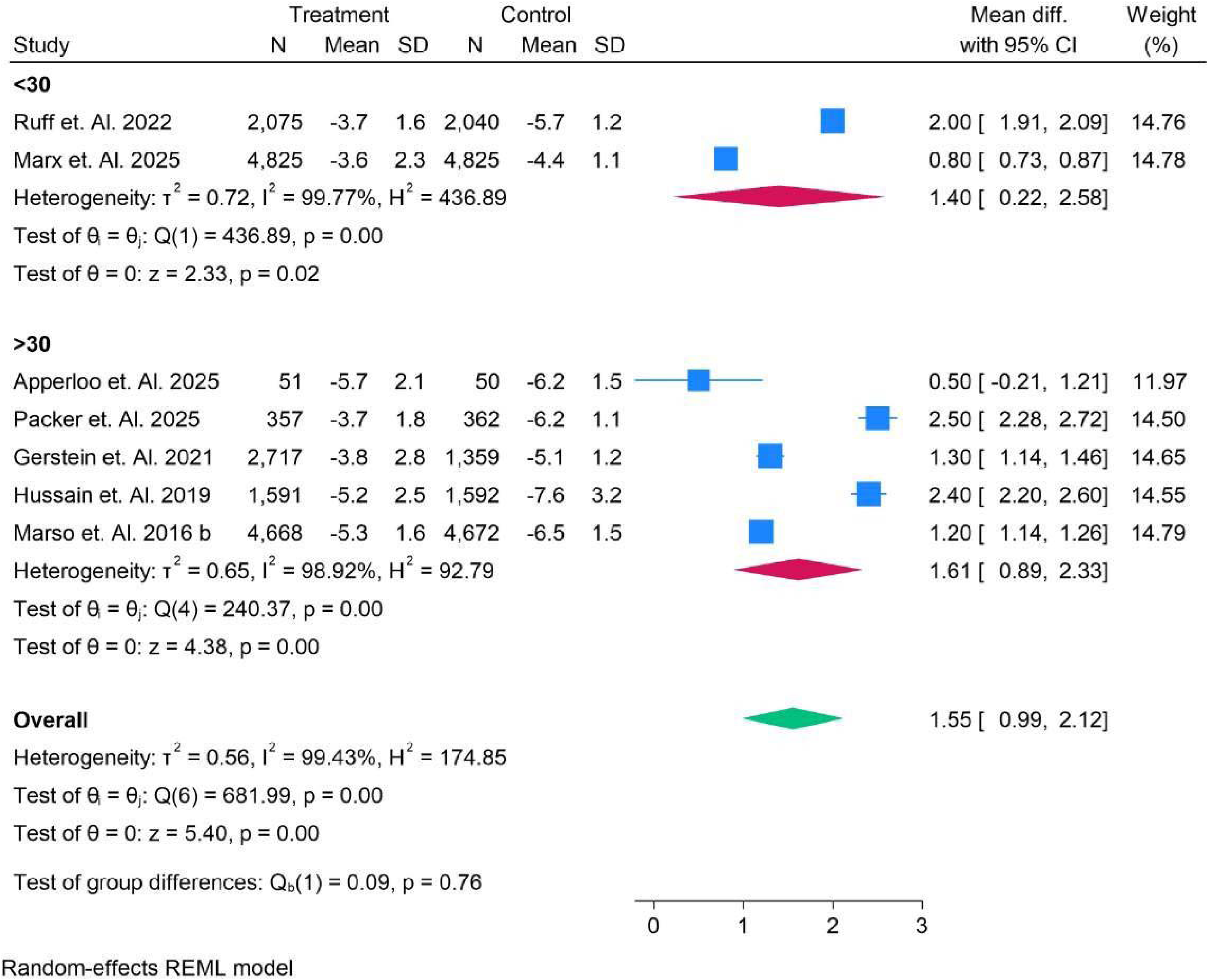
Mean Difference of eGFR in patients and Subgroup analysis of BMI.

The forest plot in Figure 46 summarizes the mean difference of estimated glomerular filtration rate (eGFR) between treatment and control groups, analyzing studies with various sample sizes and outcomes. The results show a significant positive effect in the >6.5 subgroup, with studies like Ruff et al. (2022) and Marx et al. (2025) contributing the largest weights and showing substantial improvements in eGFR (mean difference 2.0 and 1.7, respectively). In contrast, the <6.5 subgroup shows smaller effects, as seen in the study by Apperloo et al. (2025), with a minimal mean difference of 0.5. This comprehensive analysis confirms that higher HbA1c levels are associated with greater improvements in eGFR under treatment, aligning with findings from previous studies such as Gerstein et al. (2021) and Marso et al. (2016).

**Figure 46.**
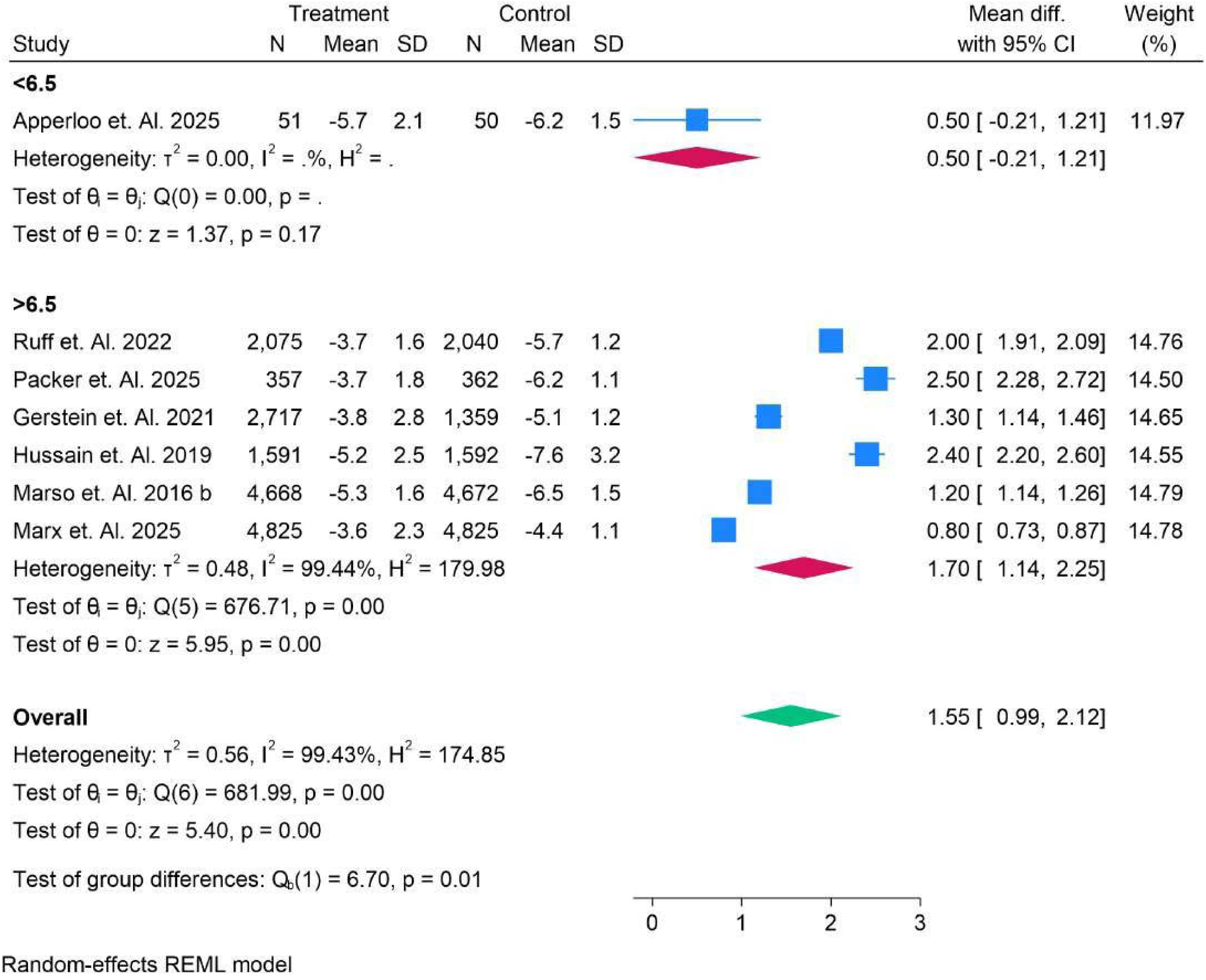
Mean Difference of eGFR in patients and Subgroup analysis of HBAIC.

Figure 47 shows the mean difference of estimated glomerular filtration rate (eGFR) between treatment and control groups with and without SGLT2 inhibition. In the subgroup with no SGLT2 administered, the mean difference was 2.50 (95% CI: [2.28, 2.72]) in Packer et al. (2025) and 1.20 (95% CI: [1.14, 1.26]) in Marso et al. (2016), with a combined mean difference of 1.85 (95% CI: [0.57, 3.12]). In the subgroup with SGLT2 administered, Ruff et al. (2022) reported a mean difference of 2.00 (95% CI: [1.91, 2.09]), Marx et al. (2025) showed 0.80 (95% CI: [0.73, 0.87]), and the combined mean difference was 1.43 (95% CI: [0.75, 2.11]). The overall mean difference across all studies was 1.55 (95% CI: [0.99, 2.12]), indicating a significant improvement in eGFR with SGLT2 inhibition.

**Figure 47.**
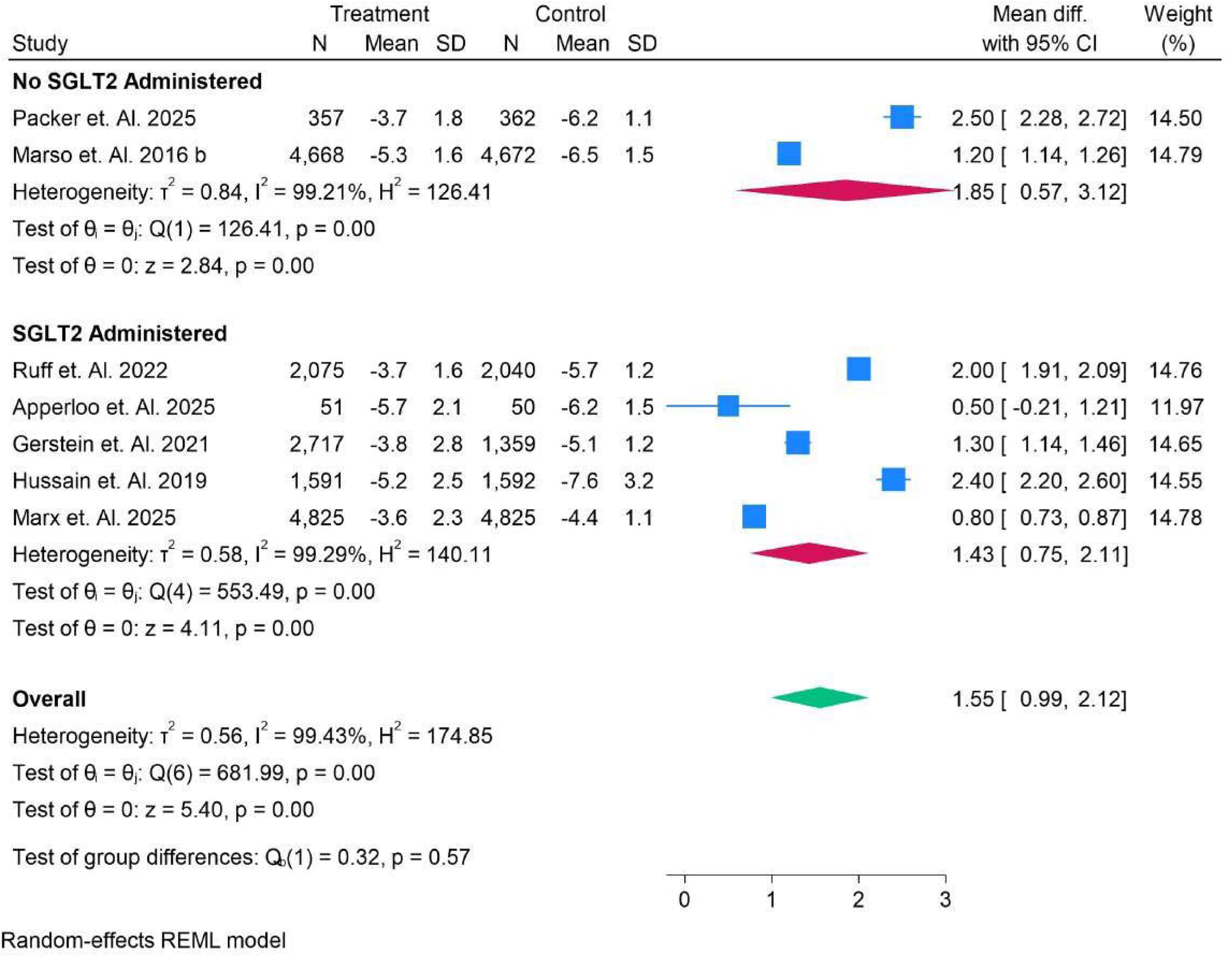
Mean Difference of eGFR in patients and Subgroup analysis of HBAlC.

Figure 48 presents the mean difference of estimated glomerular filtration rate (eGFR) between treatment and control groups based on daily vs. weekly administration. For daily administration, Ruff et al. (2022) reported a mean difference of 2.00 (95% CI: [1.91, 2.09]), Hussain et al. (2019) reported 2.40 (95% CI: [2.20, 2.60]), and Marx et al. (2025) reported 0.80 (95% CI: [0.73, 0.87]), with an overall mean difference of 1.73 (95% CI: [0.79, 2.67]). In the weekly administration subgroup, Apperloo et al. (2025) reported a mean difference of 0.50 (95% CI: [−0.21, 1.21]), Packer et al. (2025) reported 2.50 (95% CI: [2.28, 2.72]), and Marso et al. (2016) reported 1.20 (95% CI: [1.14, 1.26]), with an overall mean difference of 1.41 (95% CI: [0.62, 2.20]). The overall mean difference across both daily and weekly studies was 1.55 (95% CI: [0.99, 2.12]), showing that both daily and weekly administrations had a positive impact on eGFR, though daily treatment had a slightly higher effect.

**Figure 48.**
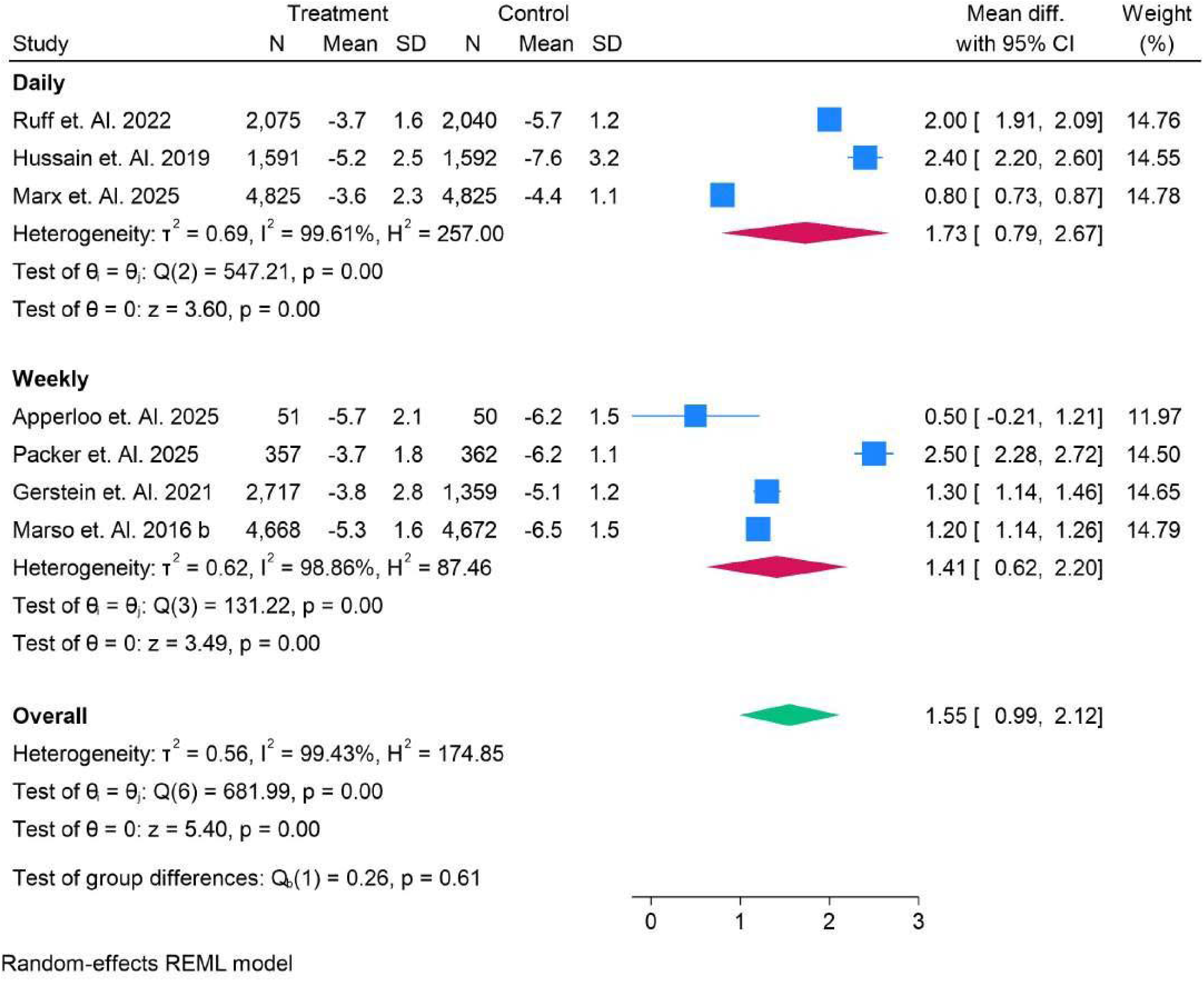
Mean Difference of eGFR in patients and Subgroup analysis of Daily vs Weekly.

Figure 49 illustrates the mean difference in estimated glomerular filtration rate (eGFR) between treatment and control groups with oral vs. subcutaneous (SC) administration. For the oral administration subgroup, Hussain et al. (2019) reported a mean difference of 2.40 (95% CI: [2.20, 2.60]), and Marx et al. (2025) reported 0.80 (95% CI: [0.73, 0.87]), with a combined mean difference of 1.60 (95% CI: [1.03, 3.17]). In the SC subgroup, Ruff et al. (2022) reported 2.00 (95% CI: [1.91, 2.09]), Apperloo et al. (2025) reported 0.50 (95% CI: [−0.21, 1.21]), and Packer et al. (2025) reported 2.50 (95% CI: [2.28, 2.72]), with a combined mean difference of 1.54 (95% CI: [0.90, 2.18]). The overall mean difference for all studies was 1.55 (95% CI: [0.99, 2.12]). This analysis shows that both oral and subcutaneous administrations are associated with positive effects on eGFR, with the oral route having a slightly higher effect on average. Figure 50 compares the mean difference in estimated glomerular filtration rate (eGFR) across various drugs. For Eflegenatide, Gerstein et al. (2021) reported a mean difference of 1.30 (95% CI: [1.14, 1.46]). In the Exenatide subgroup, Ruff et al. (2022) showed a mean difference of 2.00 (95% CI: [1.91, 2.09]). Liraglutide, as reported by Marso et al. (2016), had a mean difference of 1.20 (95% CI: [1.14, 1.26]), while Semaglutide, represented by Apperloo et al. (2025), had 0.50 (95% CI: [−0.21, 1.21]) in their study, but Marx et al. (2025) reported a higher mean difference of 0.80 (95% CI: [0.73, 0.87]), with an overall mean difference of 1.26 (95% CI: [0.10, 2.42]). Tirzepatide had a reported mean difference of 2.50 (95% CI: [2.28, 2.72]) by Packer et al. (2025). The overall mean difference across all drugs was 1.55 (95% CI: [0.99, 2.12]), confirming a beneficial impact of these drugs on eGFR with variability in the extent of effect across different drug types.

**Figure 49.**
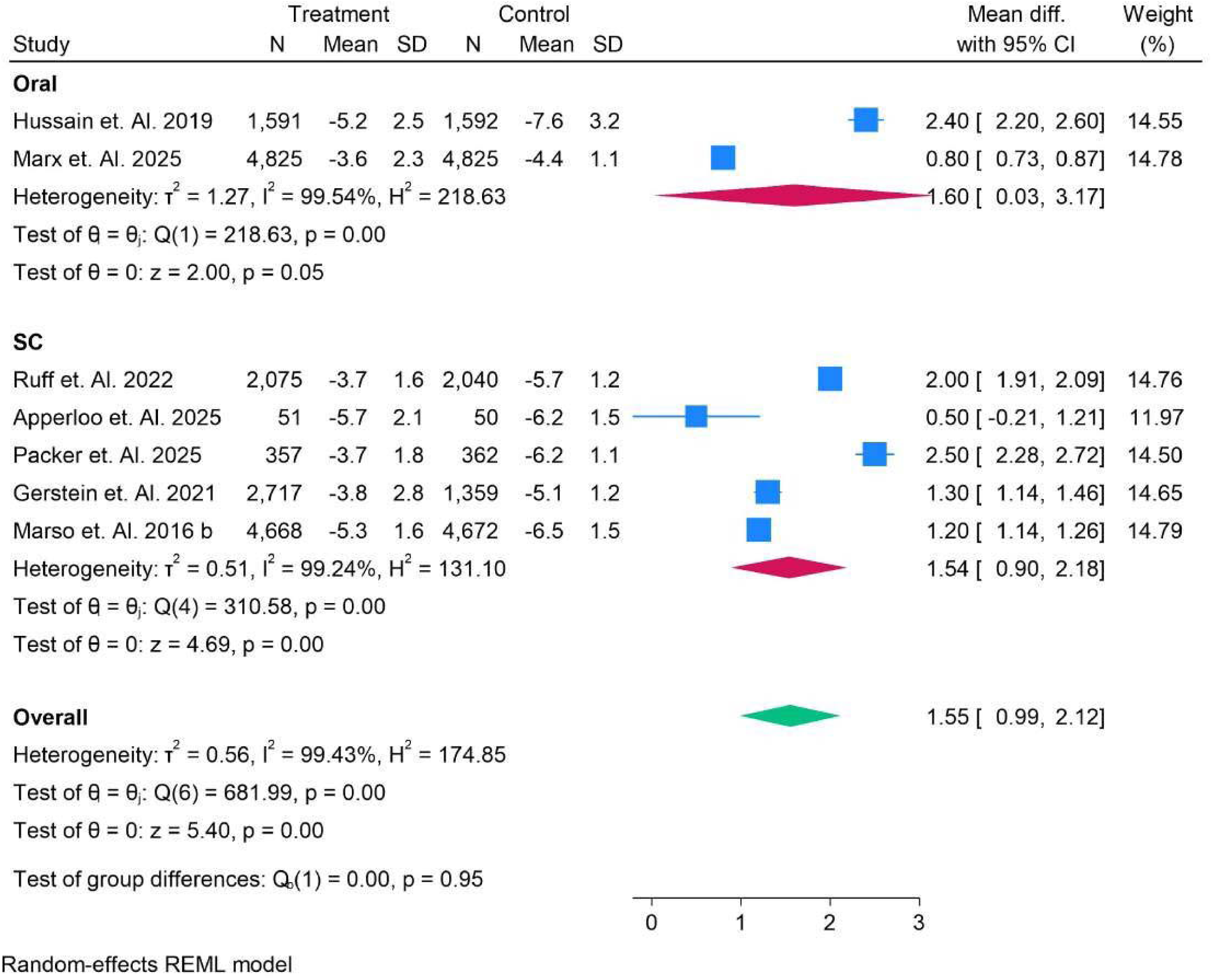
Mean Differenee of eGFR in patients and Subgroup analysis of oral vs Sub-Cutaneous.

**Figure 50.**
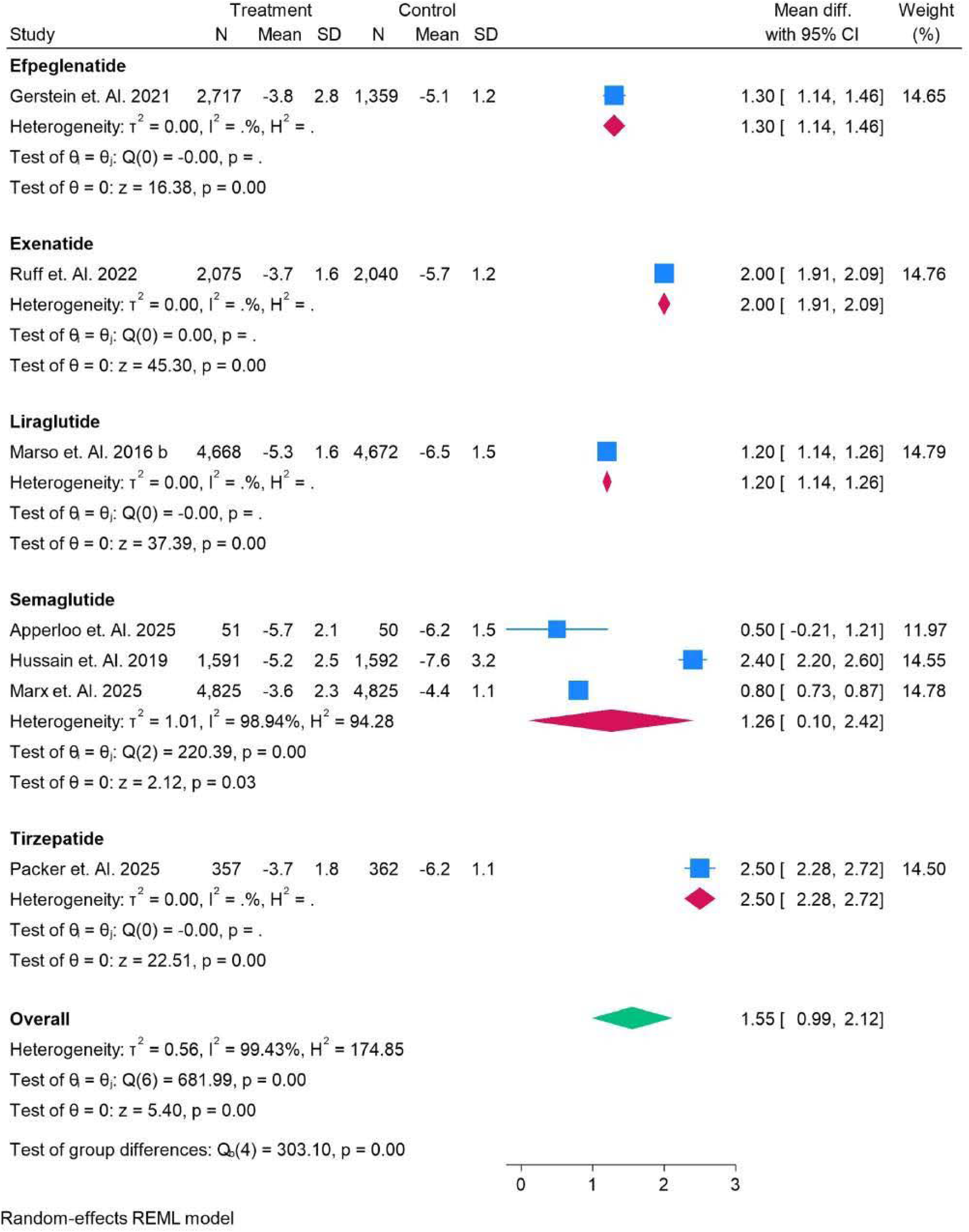
Mean Difference of eGFR in patients and Subgroup analysis of All Drugs.

### Acute Kidney Injury

Figure 51 presents the risk ratios of acute kidney injury (AKI) events in treatment vs. control groups across multiple studies. For example, in Ruff et al. (2022), the log risk ratio was 0.47 (95% CI: [−0.04, 0.97]), suggesting a slight increase in AKI risk with treatment. However, in other studies like Bladin et al. (2023), the log risk ratio was −0.30 (95% CI: [−1.02, 0.42]), indicating a potential protective effect of the treatment. Hussain et al. (2019) reported a much stronger risk increase with a log risk ratio of −2.35 (95% CI: [−2.71, −2.00]). The overall pooled log risk ratio was −0.29 (95% CI: [−0.90, 0.32]), reflecting no significant difference in the incidence of AKI events between the treatment and control groups, as shown by the heterogeneity values (I² = 98.05%, H² = 51.18%).

**Figure 51.**
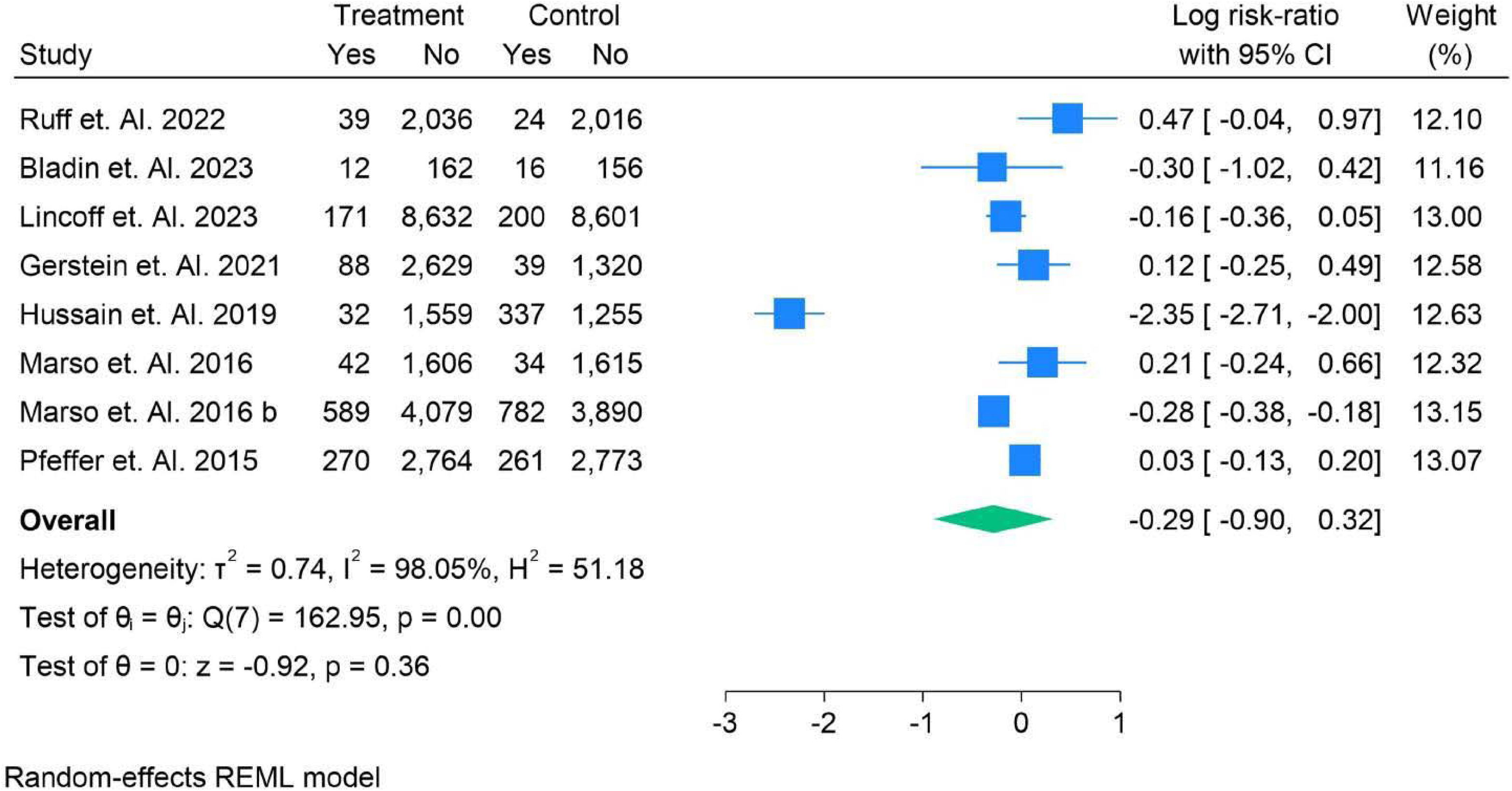
Risk ratios of Acute Kidney Injury Events in Population.

Figure 52 presents the risk ratios for acute kidney injury (AKI) events in two different age groups: under 65 and over 65. For individuals aged <65, the studies show a mix of effects. Ruff et al. (2022) reported a log risk ratio of 0.47 (95% CI: [−0.04, 0.97]), indicating a slightly increased risk of AKI with treatment. Marso et al. (2016) showed a reduced risk with a log risk ratio of −0.28 (95% CI: [−0.38, −0.18]). The pooled log risk ratio for this group was −0.01 (95% CI: [−0.21, 0.19]), suggesting no significant effect across studies.

**Figure 52.**
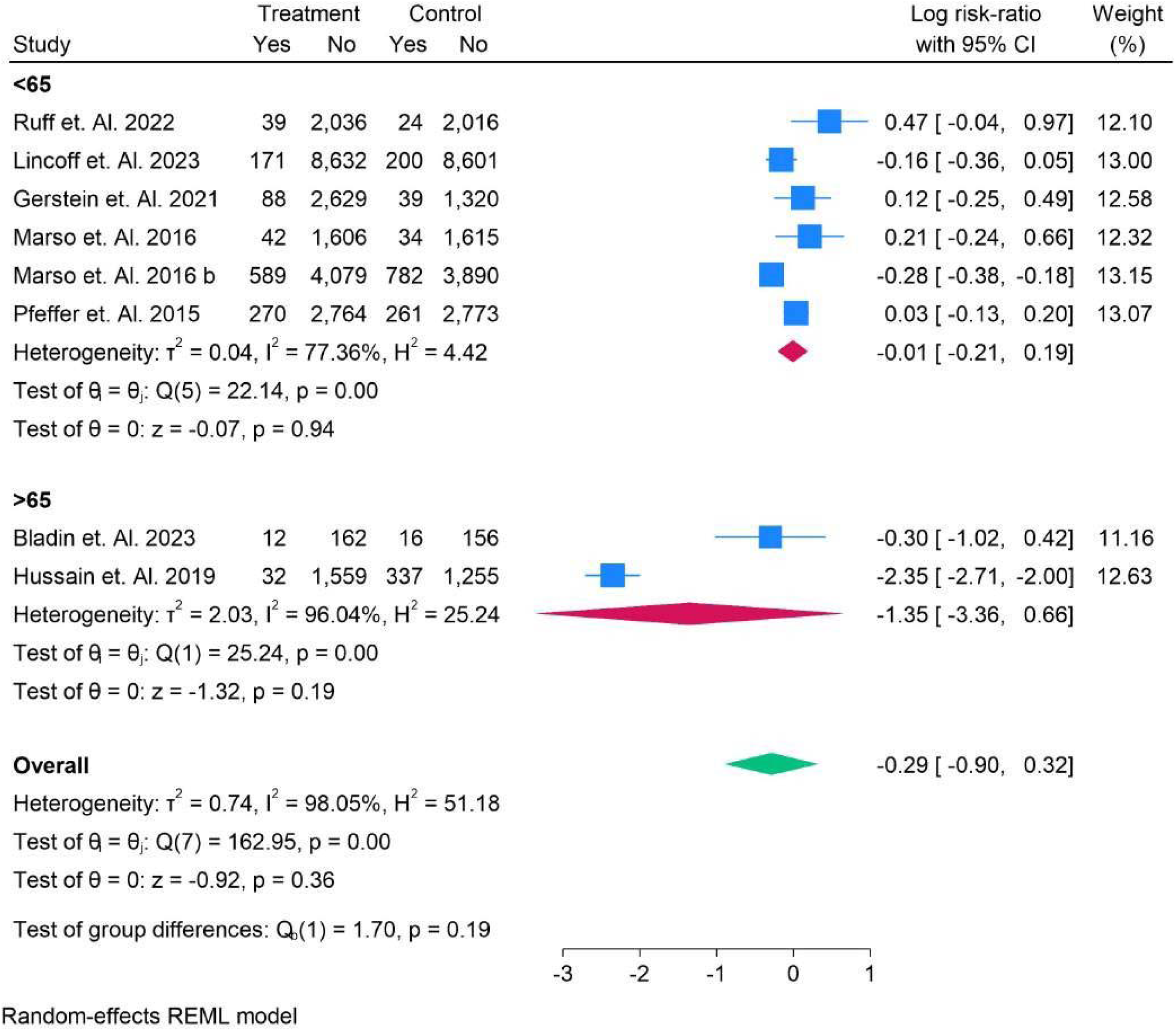
Acute Kidney Injury Events in Age Group Analysis.

In contrast, for the >65 age group, the risk was notably higher in some studies. Bladin et al. (2023) and Hussain et al. (2019) reported log risk ratios of −0.30 (95% CI: [−1.02, 0.42]) and −2.35 (95% CI: [−2.71, −2.00]), respectively, with Hussain’s study indicating a substantial increase in AKI risk in the treatment group. The overall pooled log risk ratio was −1.35 (95% CI: [−3.36, 0.66]), showing a significant effect on older populations. The overall analysis, combining both age groups, yielded a log risk ratio of −0.29 (95% CI: [−0.90, 0.32]), with no significant difference between the treatment and control groups, supported by heterogeneity statistics (I² = 98.05%, H² = 51.18%).

Figure 53 presents the risk ratios of acute kidney injury (AKI) events by race group, highlighting differences in treatment effects across various racial and ethnic groups. For the Asian group, Gerstein et al. (2021) reported a log risk ratio of 0.12 (95% CI: [−0.25, 0.49]), suggesting a minimal effect of treatment. The Caucasian group, represented by Marso et al. (2016), showed a slightly higher log risk ratio of 0.21 (95% CI: [−0.24, 0.66]). In contrast, the Hispanic group, as seen in Hussain et al. (2019), displayed a significantly negative risk ratio of 2.35 (95% CI: [−2.71, −2.00]), indicating a strong increase in AKI risk with treatment. For the White group, Bladin et al. (2023) and others reported mixed results, with a log risk ratio of −0.30 (95% CI: [−1.02, 0.42]) and −0.28 (95% CI: [−0.38, −0.18]), showing reduced risk. Lincoff et al. (2023) reported a very minimal effect of 0.16 (95% CI: [−0.36, 0.05]) for White participants. The White and Black group, as seen in Ruff et al. (2022), showed a log risk ratio of 0.47 (95% CI: [−0.04, 0.97]). The overall pooled log risk ratio was −0.29 (95% CI: [0.90, 0.32]), indicating no significant overall effect, although substantial heterogeneity was observed across the studies (I² = 98.05%, H² = 51.18%).

**Figure 53.**
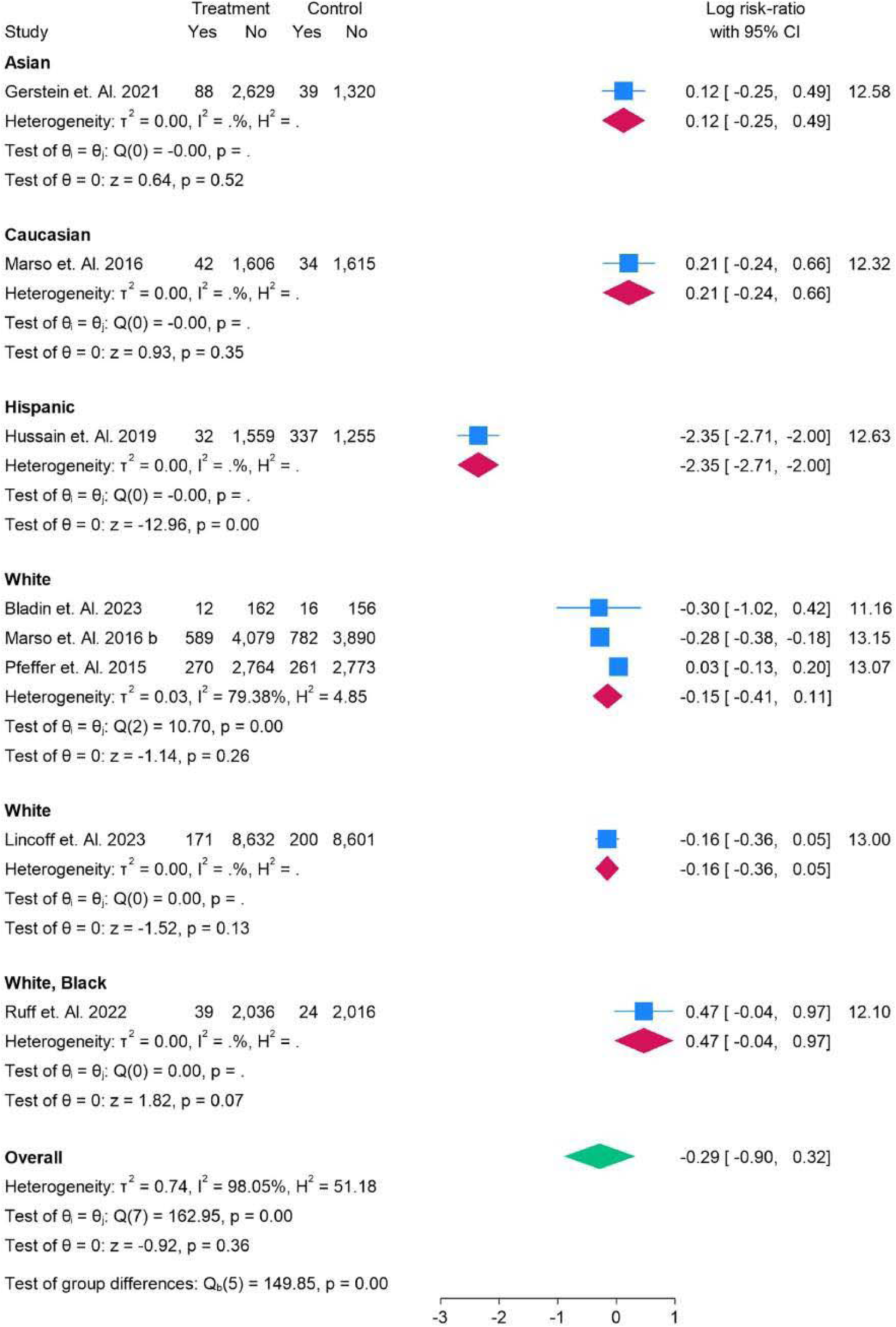
Acute Kidney Injury Events in Race Group Analysis.

Figure 54 presents the risk ratios of acute kidney injury (AKI) events in patients with normal glucose levels versus those with type 2 diabetes mellitus (T2DM). For the normal group, Lincoff et al. (2023) reported a log risk ratio of −0.16 (95% CI: [−0.36, 0.05]), indicating a minimal effect of treatment on AKI risk. In the T2DM group, the studies showed a broader range of effects. Ruff et al. (2022) reported a slight risk increase with a log risk ratio of 0.47 (95% CI: [−0.04, 0.97]), while Gerstein et al. (2021) found a minor increase in risk at 0.12 (95% CI: [−0.25, 0.49]). Hussain et al. (2019) showed a significant increase in AKI risk with a log risk ratio of −2.35 (95% CI: [−2.71, −2.00]), whereas Marso et al. (2016) and others found more moderate increases or minimal effects.

**Figure 54.**
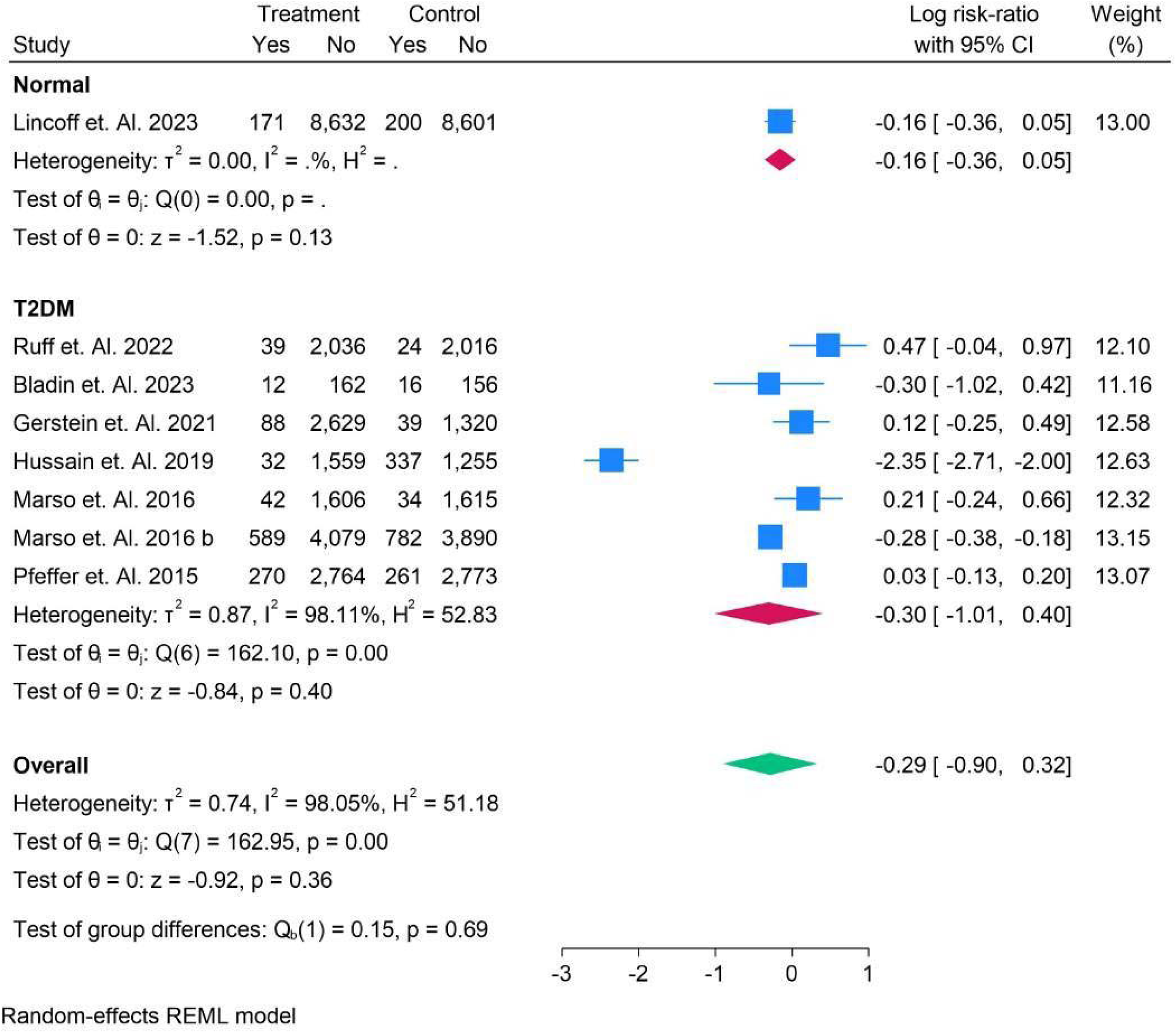
Acute Kidney Injury Events in Normal vs T2DM Group Analysis.

**Figure 55.**
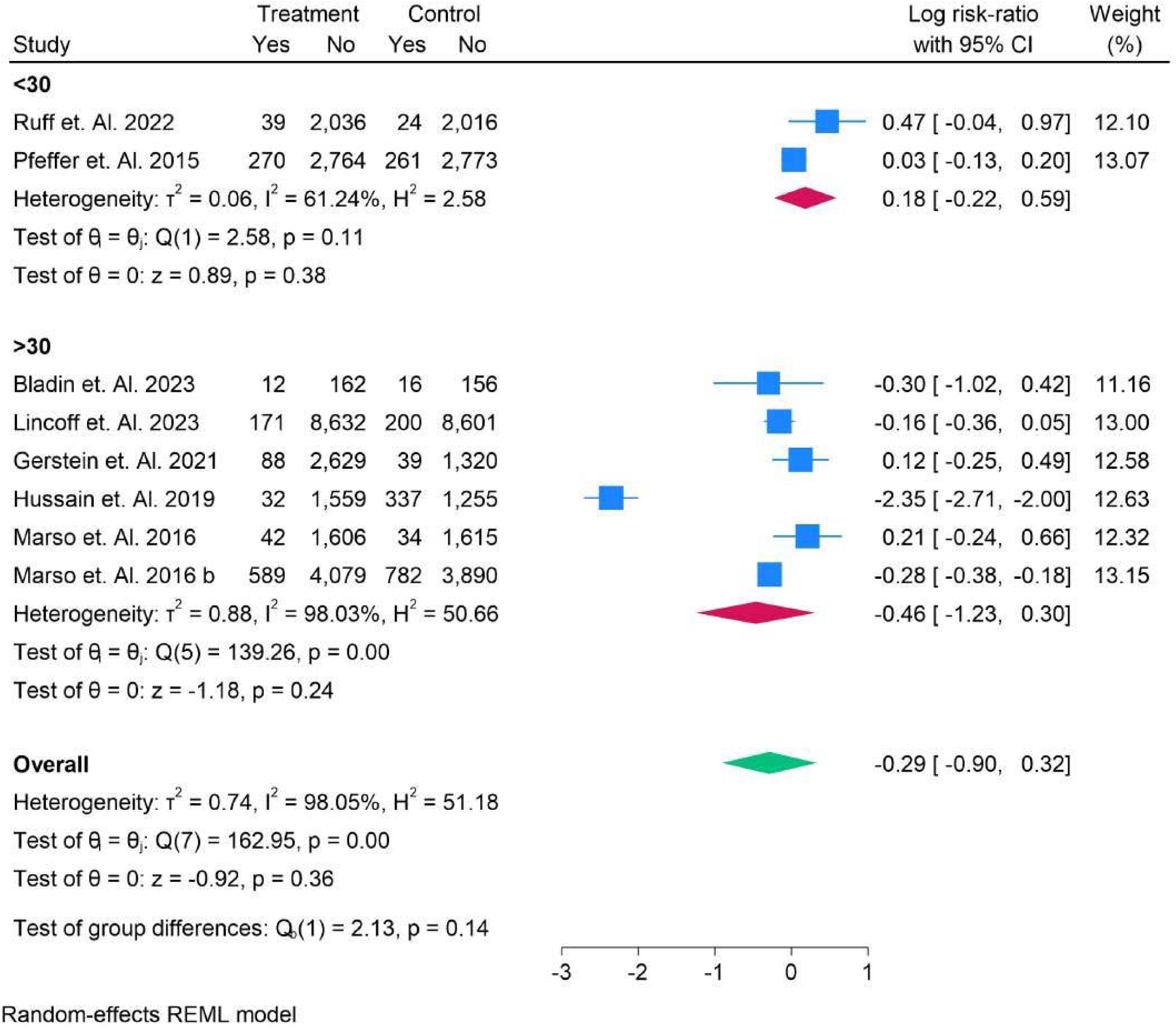
Acute Kidney Injury Events in BMI Group Analysis.

The pooled log risk ratio for the T2DM group was −0.30 (95% CI: [−1.01, 0.40]), reflecting a slight but not significant increase in AKI risk for T2DM patients. The overall log risk ratio for all studies combined was −0.29 (95% CI: [−0.90, 0.32]), with substantial heterogeneity (I² = 98.05%, H² = 51.18%) and no significant difference between normal and T2DM groups, as indicated by the test for group differences (p = 0.69).

In the >30 BMI group, a more notable effect was seen. Bladin et al. (2023) and Lincoff et al. (2023) reported minimal effects, with log risk ratios of −0.30 (95% CI: [−1.02, 0.42]) and −0.16 (95% CI: [−0.36, 0.05]), respectively. However, Hussain et al. (2019) and Marso et al. (2016) found significantly higher risks with log risk ratios of −2.35 (95% CI: [−2.71, −2.00]) and −0.28 (95% CI: [−0.38, −0.18]), respectively. The overall pooled log risk ratio for the >30 BMI group was −0.46 (95% CI: [−1.23, 0.30]), indicating a modest but significant increase in AKI risk for patients with higher BMI.

The overall pooled log risk ratio for all studies was −0.29 (95% CI: [−0.90, 0.32]), with no significant difference between the groups, as reflected in the test for group differences (p = 0.14). Despite the significant heterogeneity observed (I² = 98.05%, H² = 51.18%), the results suggest that BMI may have a limited effect on the overall AKI risk associated with treatment.

Figure 56 presents the risk ratios of acute kidney injury (AKI) events based on HbA1c levels, divided into two subgroups: <6.5% and >6.5%. For the <6.5% subgroup, Bladin et al. (2023) reported a log risk ratio of −0.30 (95% CI: [−1.02, 0.42]), and Lincoff et al. (2023) reported −0.16 (95% CI: [−0.36, 0.05]), indicating minimal effect of treatment on AKI risk. The pooled log risk ratio for this subgroup was −0.17 (95% CI: [−0.36, 0.03]), suggesting no significant effect across studies.

**Figure 56.**
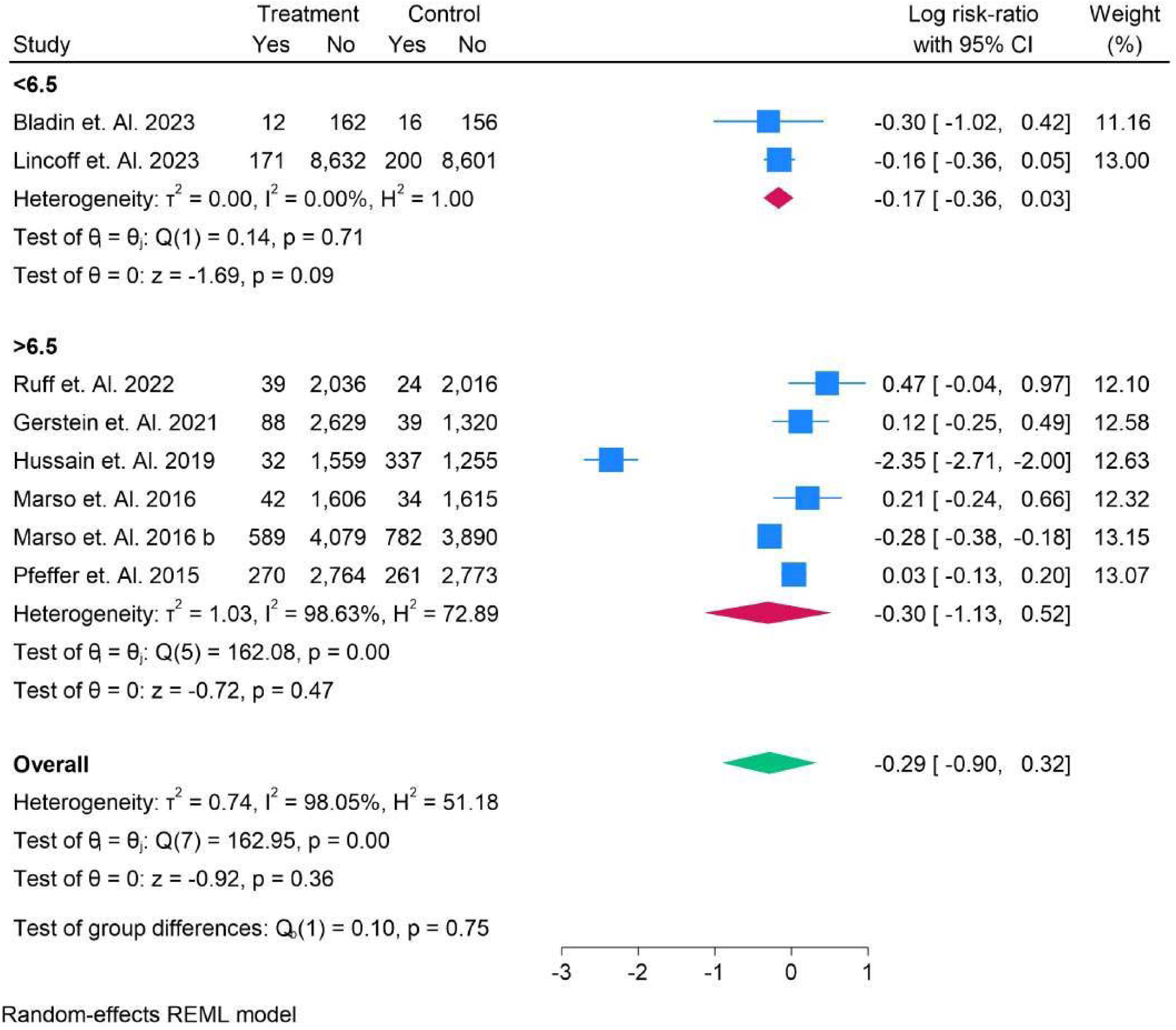
Acute Kidney Injury Events in HBA1C Group Analysis.

In the >6.5% subgroup, Hussain et al. (2019) showed a significant increase in risk with a log risk ratio of −2.35 (95% CI: [−2.71, −2.00]), while Marso et al. (2016) and others found more moderate increases or minimal effects. The pooled log risk ratio for this group was −0.30 (95% CI: [−1.13, 0.52]), indicating that while some studies found a higher risk in this subgroup, the overall effect was not statistically significant.

The overall pooled log risk ratio for all studies was −0.29 (95% CI: [−0.90, 0.32]), with no significant difference between the subgroups, supported by a test for group differences (p = 0.75). The results suggest that HbA1c levels may not significantly influence the risk of AKI events associated with treatment, as shown by the substantial heterogeneity across studies (I² = 98.05%, H² = 51.18%).

Figure 57 compares AKI risk ratios between SGLT2 and non-SGLT2 groups. In the non-SGLT2 subgroup, Pfeffer et al. (2015) reported a minimal risk ratio of −0.11 (95% CI: [−0.29, 0.06]), while in the SGLT2 group, Hussain et al. (2019) observed a significant increase in risk (−2.35, 95% CI: [−2.71, −2.00]). The overall risk ratio was −0.29 (95% CI: [−0.90, 0.32]), showing no significant difference. Figure 58 compares AKI risk ratios between daily and weekly treatment regimens. In the daily subgroup, Hussain et al. (2019) observed a significant increase in AKI risk (−2.35, 95% CI: [−2.71, −2.00]), whereas the weekly subgroup showed more moderate effects, such as in Lincoff et al. (2023) (−0.16, 95% CI: [−0.36, 0.05]). The overall pooled log risk ratio was −0.29 (95% CI: [−0.90, 0.32]), showing no significant difference. Figure 59 compares the log risk ratios of acute kidney injury (AKI) events in oral versus subcutaneous (SC) treatment subgroups. For the oral group, Hussain et al. (2019) observed a significant increase in AKI risk with a log risk ratio of −2.35 (95% CI: [−2.71, 2.00]). In contrast, the SC subgroup showed minimal effects, with studies like Lincoff et al. (2023) reporting a small risk reduction (log risk ratio −0.16, 95% CI: [−0.36, 0.05]). The overall pooled log risk ratio across both subgroups was −0.29 (95% CI: [−0.90, 0.32]), indicating no significant difference between oral and SC treatments. Substantial heterogeneity was observed (I² = 98.05%, H² = 51.18%), with a significant test for group differences (p = 0.00).

**Figure 57.**
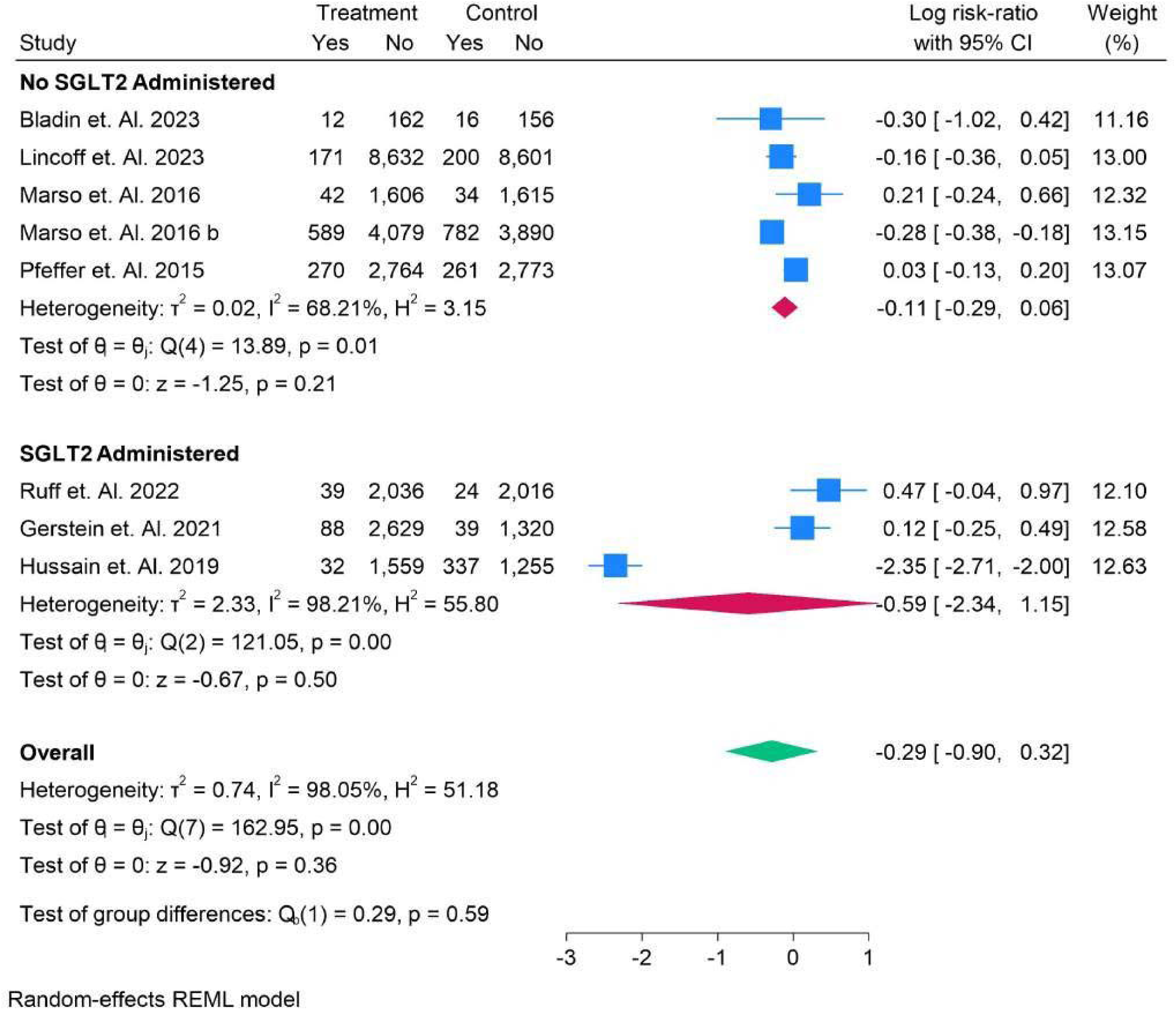
Acute Kidney Injury Events in SGLT2 vs Non-SGLT2 Group Analysis.

**Figure 58.**
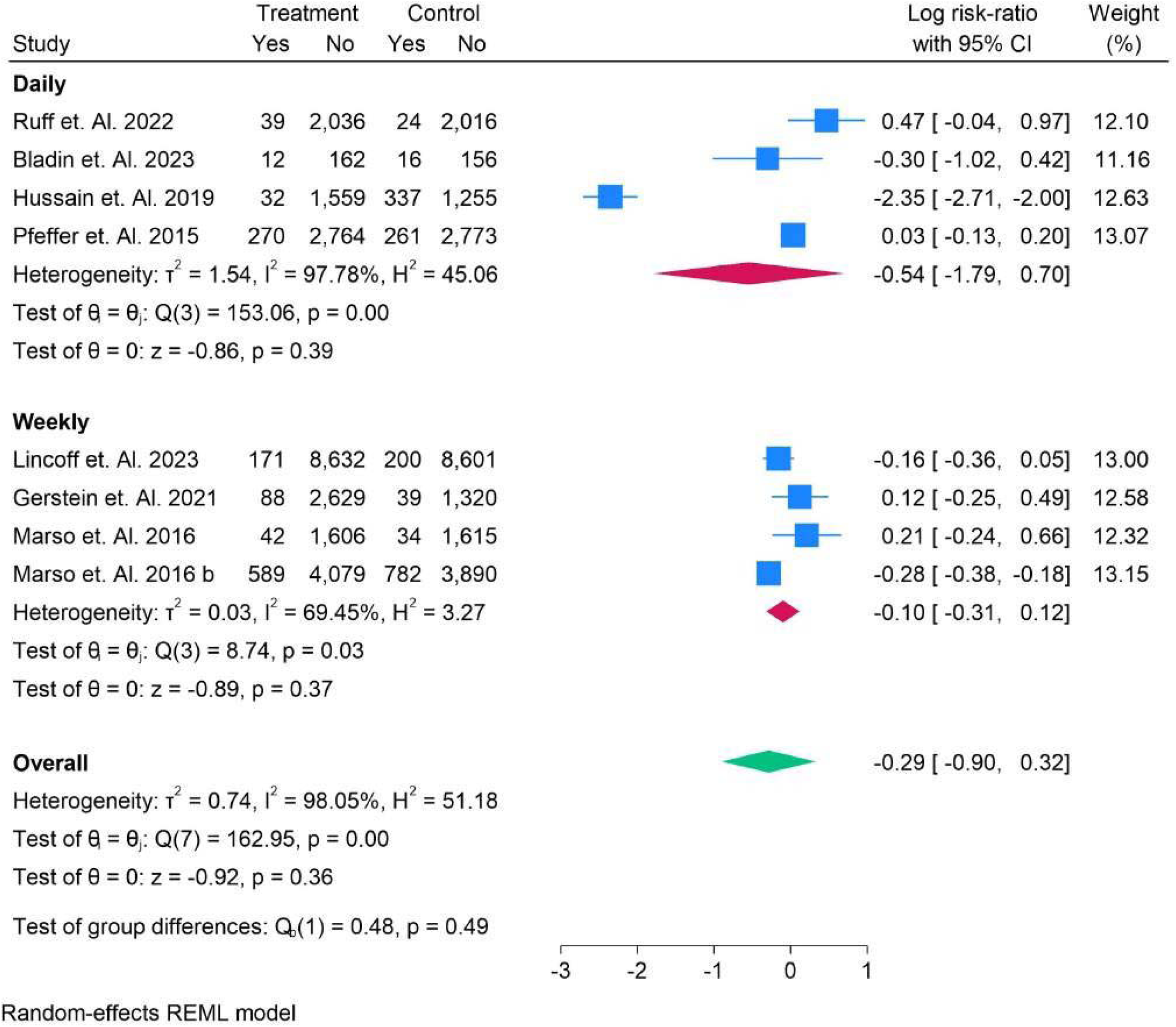
Acute Kidney Injury Events in Daily vs Weekly Sub-Group Analysis.

**Figure 59.**
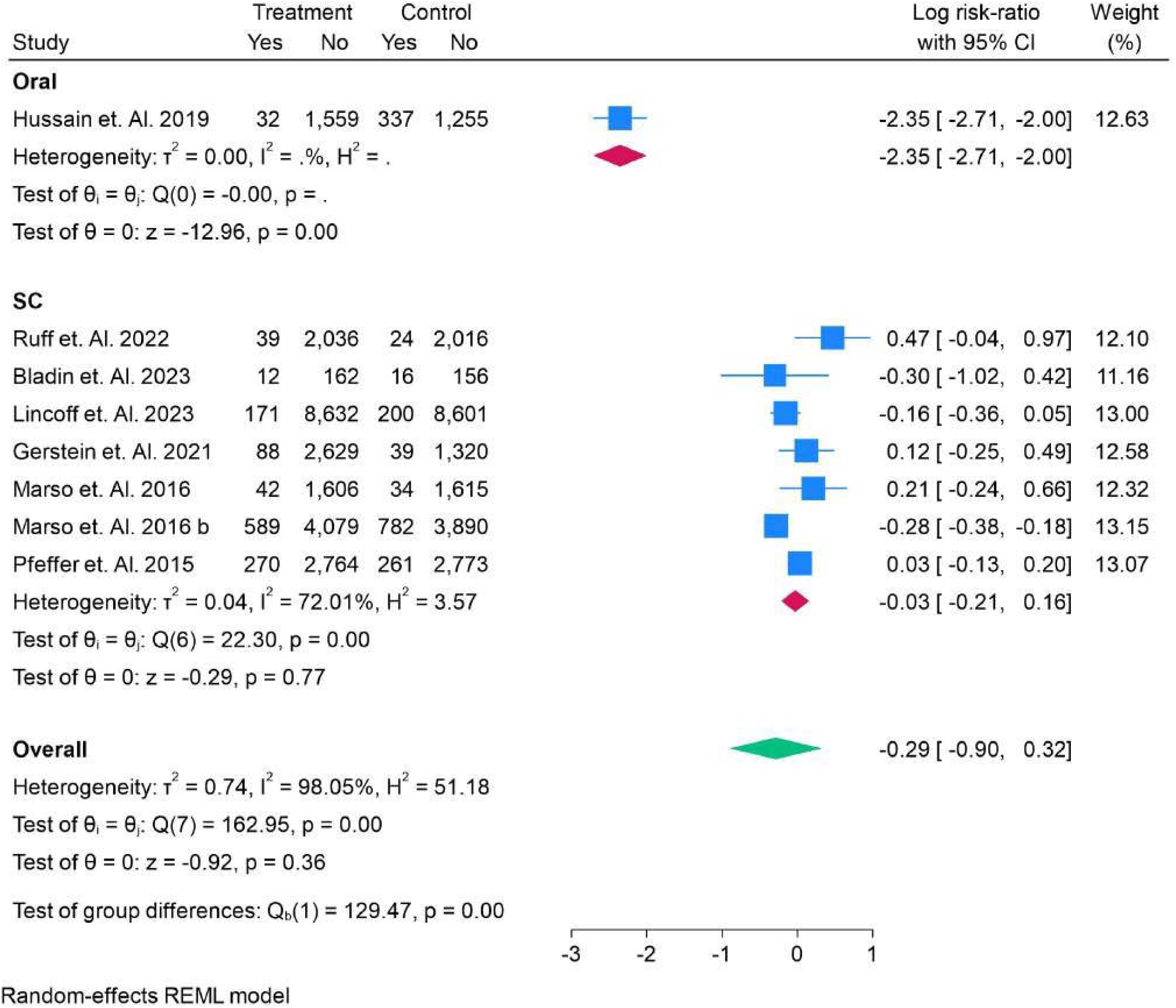
Acute Kidney Injury Events in Oral vs Sub-Cutaneous Sub-Group Analysis.

Figure 60 compares acute kidney injury (AKI) risk ratios across different drug subgroups. For Eflegenatide and Exenatide, Gerstein et al. (2021) and Ruff et al. (2022) reported minimal risk effects with log risk ratios of 0.12 (95% CI: [−0.25, 0.49]) and 0.47 (95% CI: [−0.04, 0.97]), respectively. In contrast, Liraglutide (Marso et al. 2016) showed a negative risk effect (−0.28, 95% CI: [−0.38, −0.18]). Lixisenatide (Pfeffer et al. 2015) had no significant effect, with a log risk ratio of 0.03 (95% CI: [−0.13, 0.20]). Semaglutide showed varying results, with Lincoff et al. (2023) reporting −0.16 (95% CI: [−0.36, 0.05]) and Marso et al. (2016) showing a higher negative risk (−0.77, 95% CI: [−2.33, 0.80]). The overall pooled log risk ratio for all drugs was −0.29 (95% CI: [−0.90, 0.32]), with significant heterogeneity observed across studies (I² = 98.05%, H² = 51.18%) and a notable test for group differences (p = 0.01).

**Figure 60.**
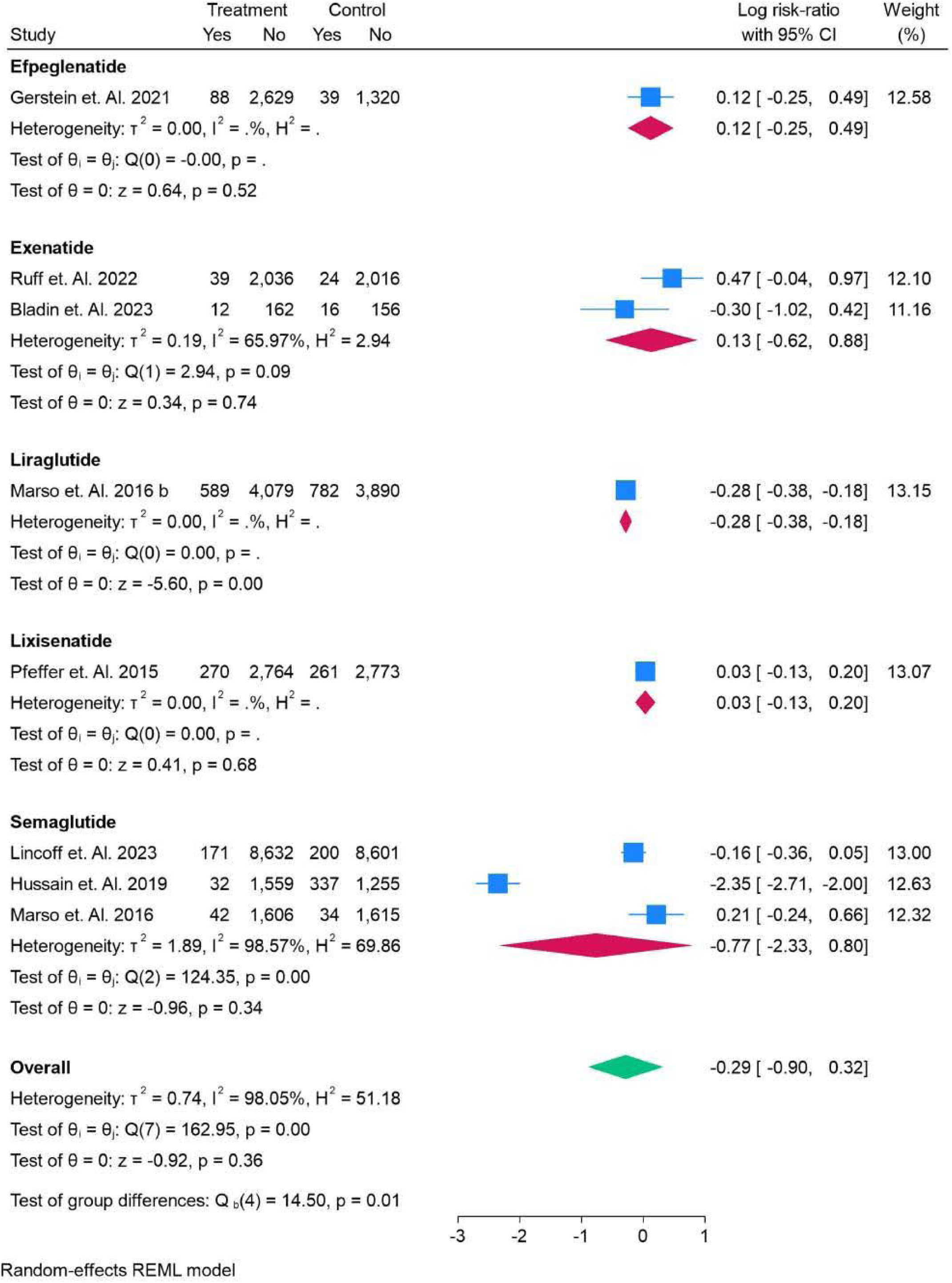
Acute Kidney Injury Events in All Drugs Sub-Group Analysis.

### Occurrence of CKD in patients

Figure 61 compares the risk ratio for the occurrence of chronic kidney disease (CKD) between treatment and control groups. Packer et al. (2025) reported a minimal effect, with a log risk ratio of −0.03 (95% CI: [−0.05, 0.01]), while Marx et al. (2025) showed a significant reduction in CKD risk with −1.17 (95% CI: [−1.29, −1.06]). The overall pooled log risk ratio was −0.29 (95% CI: [−0.67, 0.09]), indicating a slight, non-significant reduction in CKD risk. The heterogeneity was high (I² = 98.92%, H² = 92.32%), suggesting significant variability across studies. Figure 62 compares the risk ratios for chronic kidney disease (CKD) occurrence in patients below and above the age of 65. For patients under 65, Margulies et al. (2016) and Marso et al. (2016) reported minimal effects, with log risk ratios of 0.15 (95% CI: [−0.13, 0.44]) and −0.35 (95% CI: [−0.43, −0.27]), respectively. In contrast, for patients above 65, Packer et al. (2025) and Marx et al. (2025) showed more significant effects, with log risk ratios of −0.03 (95% CI: [−0.05, −0.01]) and −1.17 (95% CI: [−1.29, −1.06]). The pooled overall log risk ratio was −0.29 (95% CI: [−0.67, 0.09]), indicating a slight reduction in CKD risk, but no significant difference between age groups (p = 0.43). Figure 63 presents the risk ratios for chronic kidney disease (CKD) occurrence in patients of different race groups. For White patients, Packer et al. (2025) reported a minimal risk reduction with a log risk ratio of −0.03 (95% CI: [−0.05, −0.01]), and Marx et al. (2025) observed a significant reduction of −1.17 (95% CI: [−1.29, −1.06]). For another White subgroup, Lincoff et al. (2023) reported a smaller effect (−0.25, 95% CI: [−0.45, −0.04]). The overall pooled log risk ratio was −0.29 (95% CI: [−0.67, 0.09]), indicating a slight reduction in CKD risk, with no significant difference across race groups (p = 0.84).

**Figure 61.**
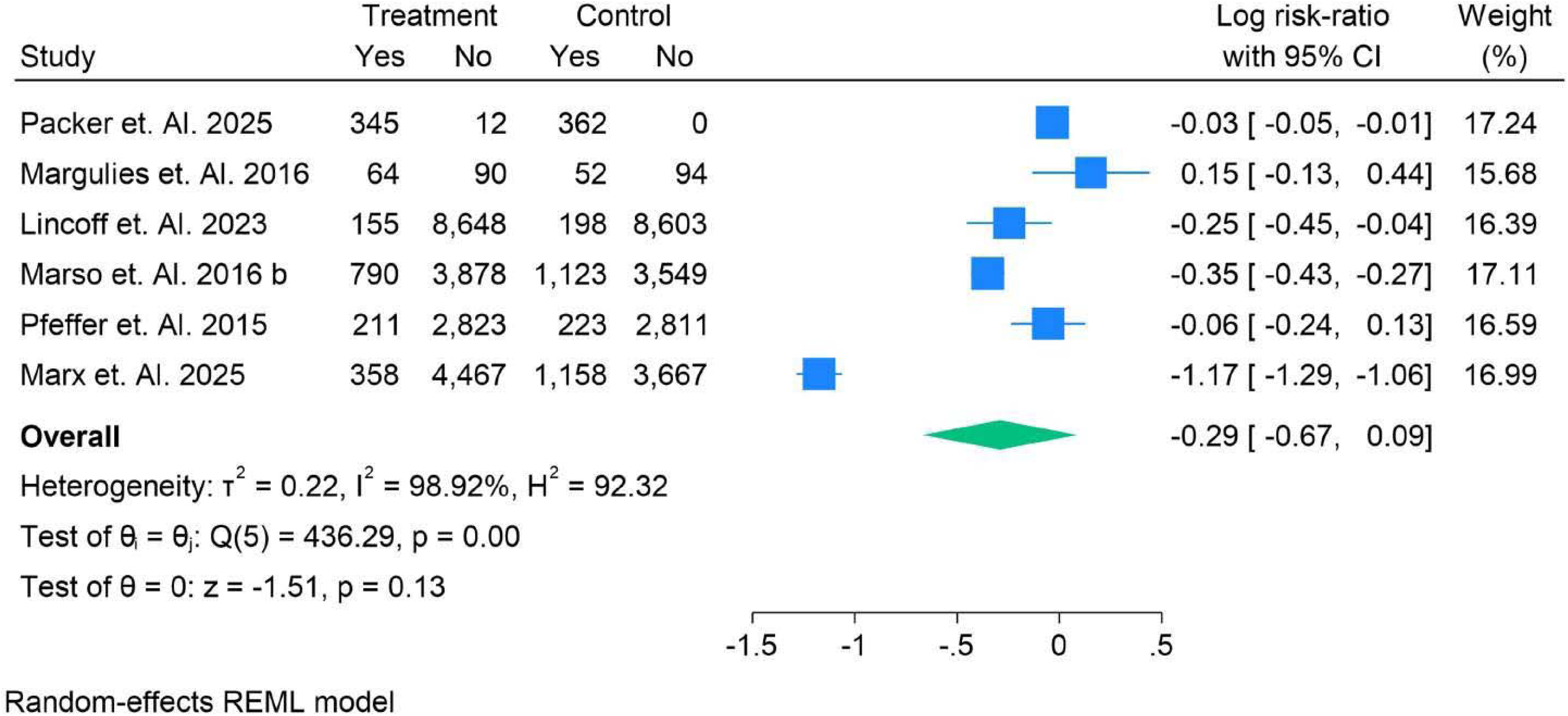
Risk Ratio of Occurrence of CKD in Patients.

**Figure 62.**
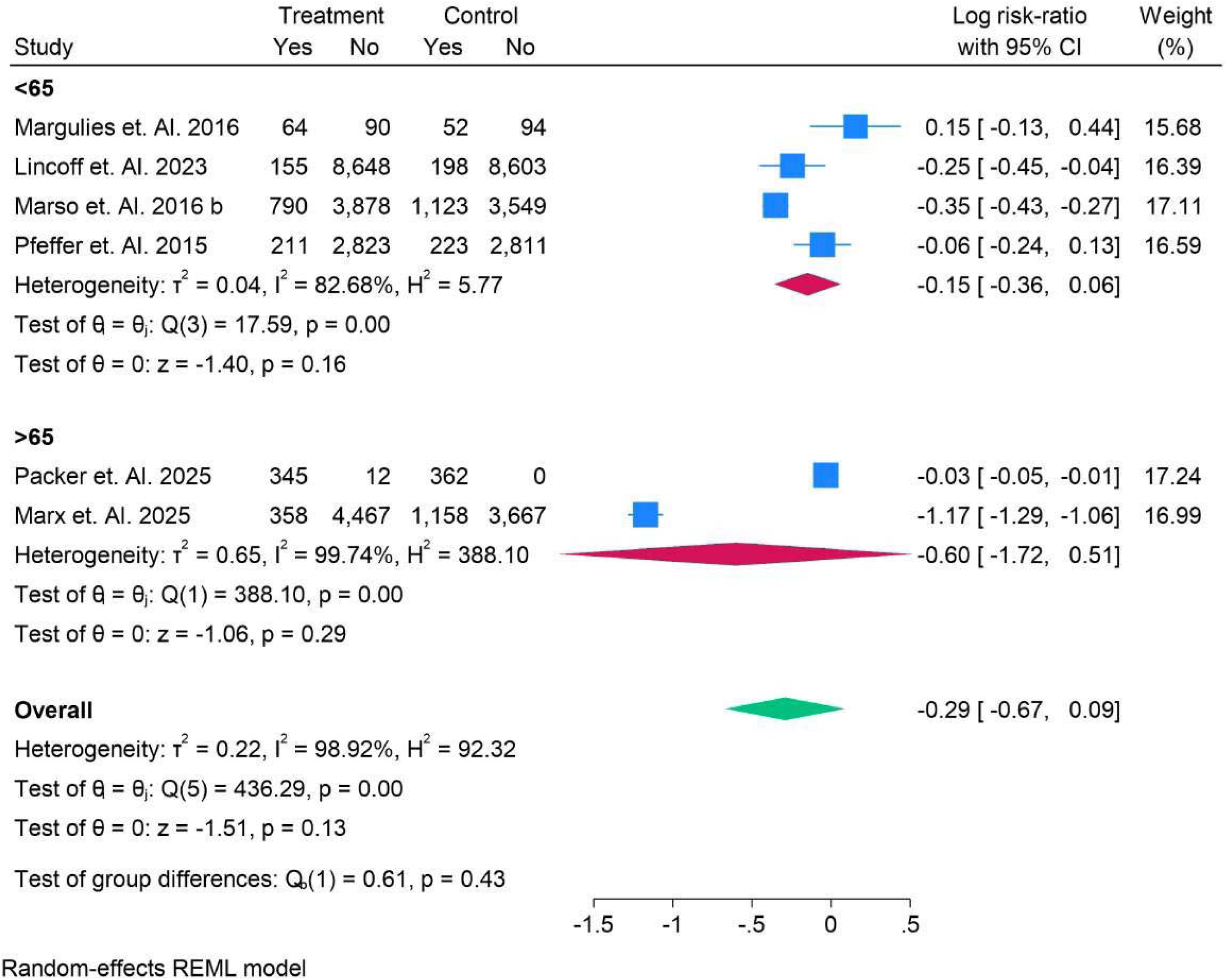
Risk Ratio of Occurrence of CKD in Patients having Age Group.

**Figure 63.**
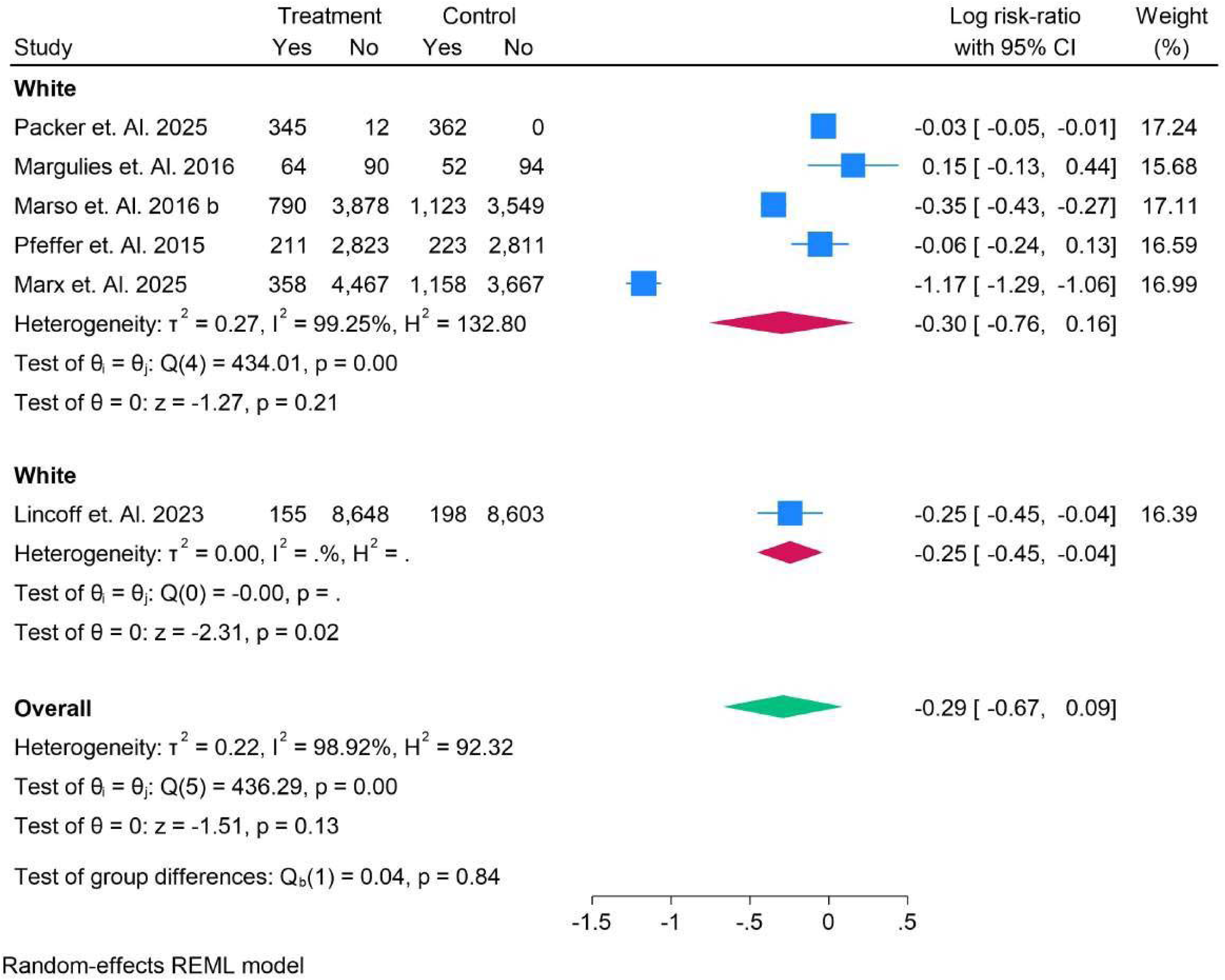
Risk Ratio of Occurrence of CKD in Patients having Race Group.

Figure 64 presents the risk ratios for chronic kidney disease (CKD) occurrence in patients with normal glucose levels versus those with type 2 diabetes mellitus (T2DM). In the normal group, Lincoff et al. (2023) showed a minimal risk reduction with a log risk ratio of −0.25 (95% CI: [−0.45, −0.04]). In the T2DM group, Packer et al. (2025) and Marx et al. (2025) found significant reductions in risk, with log risk ratios of −0.03 (95% CI: [−0.05, −0.01]) and −1.17 (95% CI: [−1.29, −1.06]), respectively. The overall pooled log risk ratio was −0.29 (95% CI: [−0.67, 0.09]), indicating a slight but not statistically significant difference between the groups (p = 0.84).

**Figure 64.**
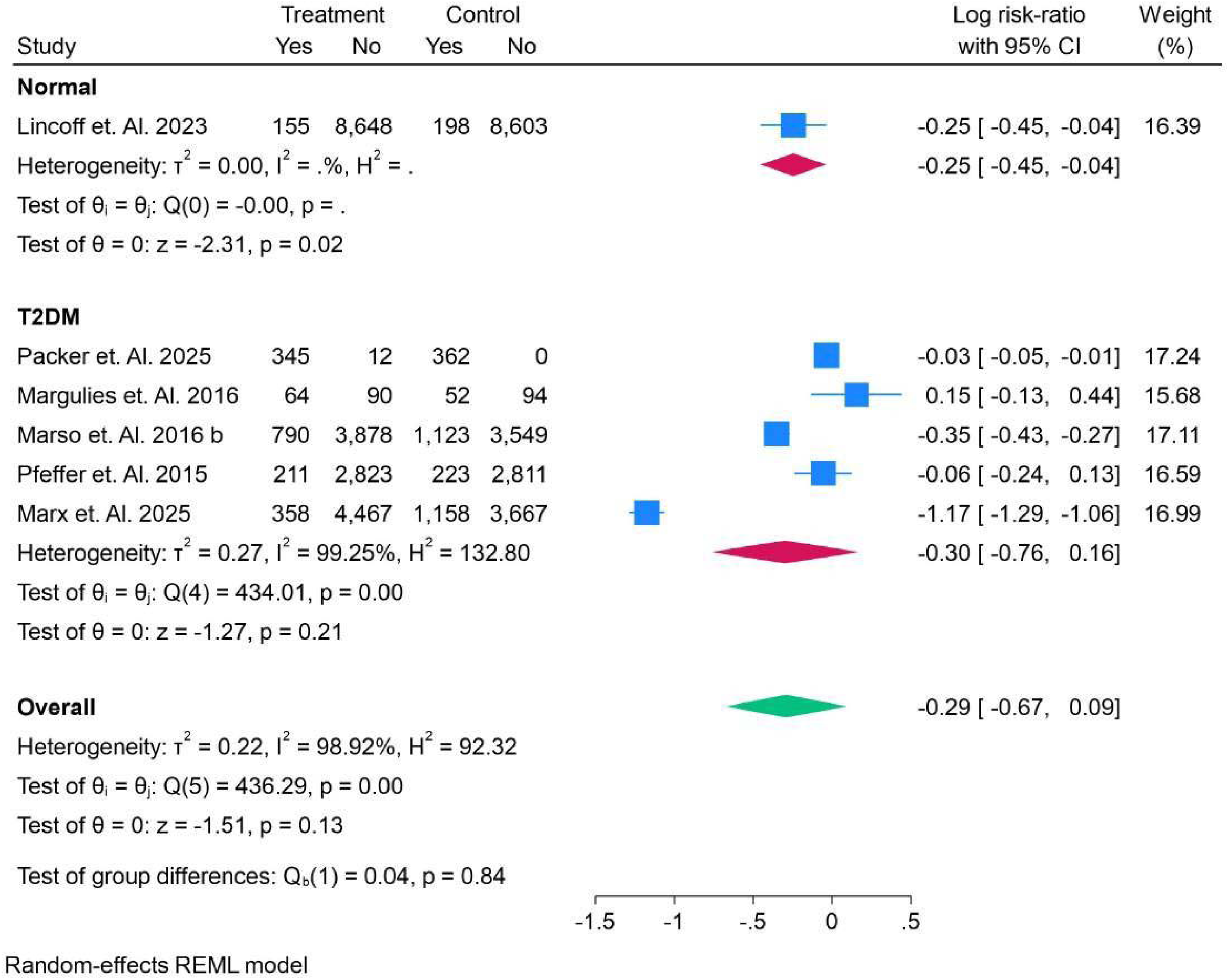
Risk Ratio of Occurrence of CKD in Patients having Normal vs T2DM Group.

Figure 65 compares the risk ratios for chronic kidney disease (CKD) occurrence in patients with BMI <30 and BMI >30. In the <30 BMI subgroup, Pfeffer et al. (2015) showed a minimal effect with a log risk ratio of −0.06 (95% CI: [−0.24, 0.13]), while Marx et al. (2025) reported a significant risk reduction of −1.17 (95% CI: [−1.29, 1.06]). For the >30 BMI group, Packer et al. (2025) and Marso et al. (2016) found minimal effects, with log risk ratios of −0.03 (95% CI: [−0.05, −0.01]) and −0.35 (95% CI: [−0.43, −0.27]), respectively. The overall pooled log risk ratio was −0.29 (95% CI: [−0.67, 0.09]), with no significant difference between the BMI subgroups (p = 0.40). Figure 66 compares the risk ratios for chronic kidney disease (CKD) occurrence in patients with HbA1c levels <6.5% and >6.5%. In the <6.5% subgroup, Lincoff et al. (2023) reported a small risk reduction with a log risk ratio of −0.25 (95% CI: [−0.45, −0.04]). In the >6.5% subgroup, Packer et al. (2025) showed a minimal risk reduction (−0.03, 95% CI: [−0.05, −0.01]), while Marx et al. (2025) observed a significant reduction of −1.17 (95% CI: [−1.29, −1.06]). The overall pooled log risk ratio for both subgroups was −0.29 (95% CI: [−0.67, 0.09]), indicating no significant difference between the groups (p = 0.84).

**Figure 65.**
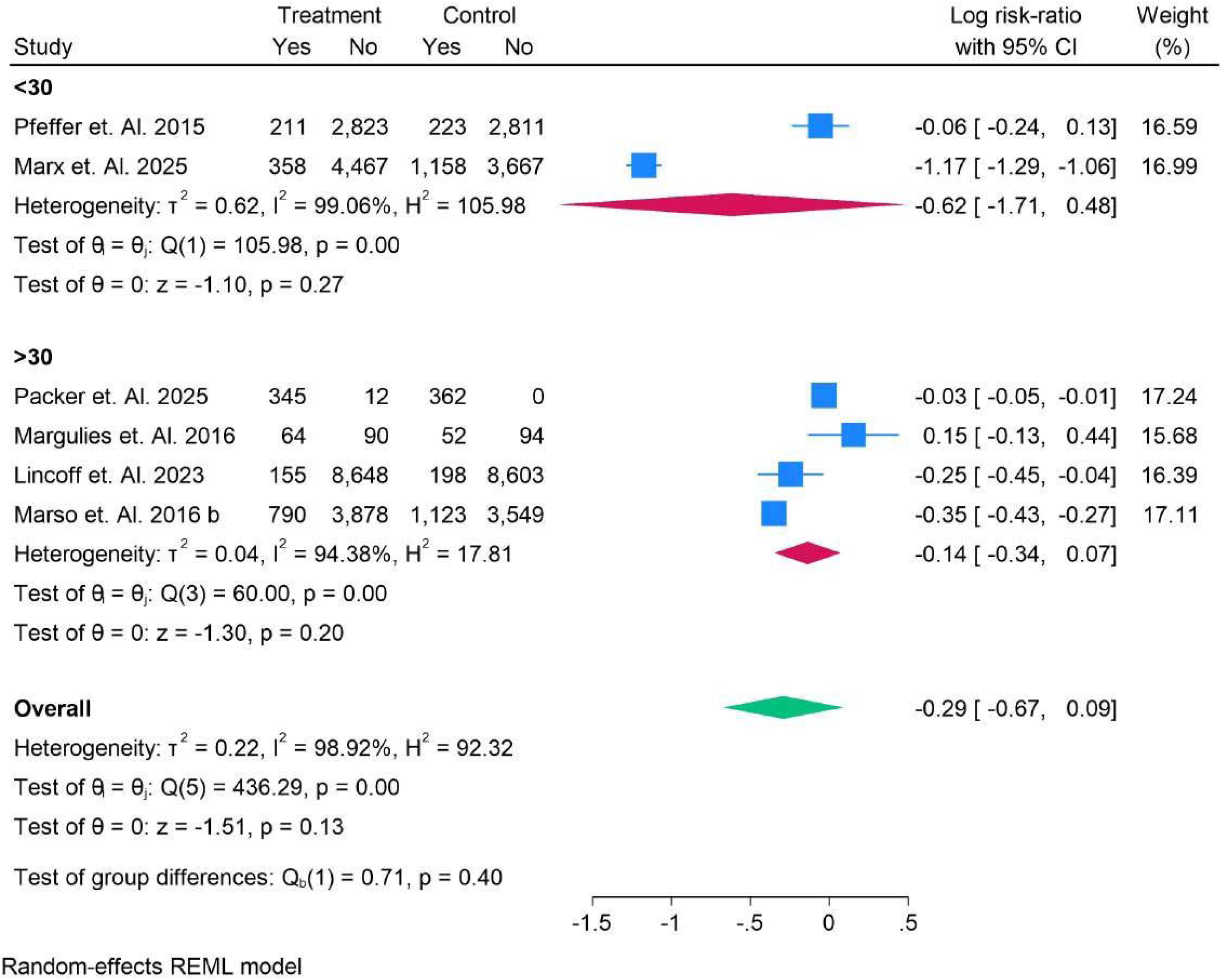
Risk Ratio of Occurrence of CKD in Patients having BMI Group.

**Figure 66.**
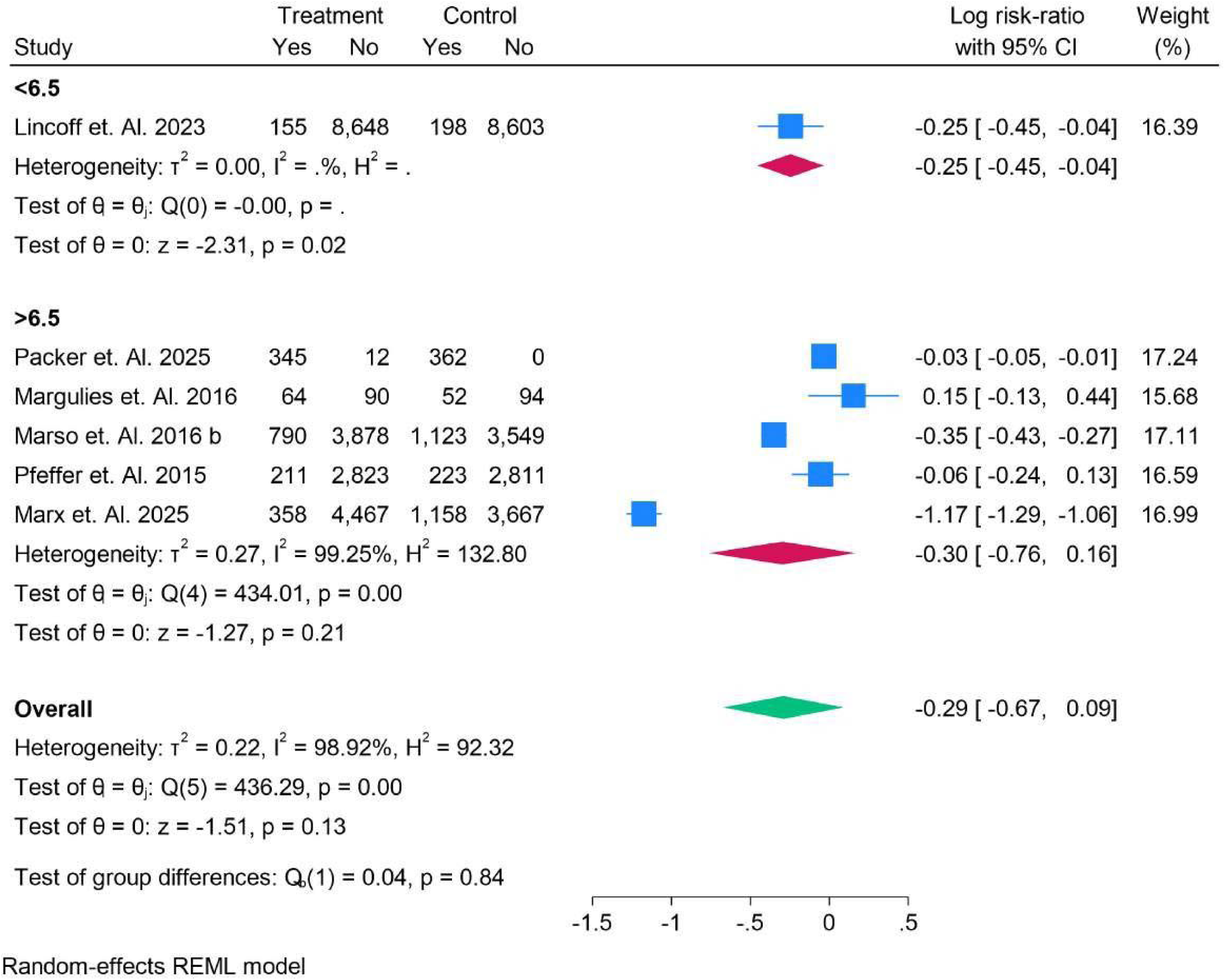
Risk Ratio of Occurrence of CKD in Patients having HBA1C Group.

Figure 67 presents the risk ratios for the occurrence of chronic kidney disease (CKD) in patients with and without SGLT2 inhibitors. In the group without SGLT2, studies like Packer et al. (2025) and Lincoff et al. (2023) showed minimal effects, with log risk ratios of −0.03 (95% CI: [−0.05, −0.01]) and −0.25 (95% CI: [−0.45, −0.04]). In contrast, in the SGLT2-administered group, Marx et al. (2025) showed a significant reduction in CKD risk with a log risk ratio of −1.17 (95% CI: [−1.29, −1.06]). The overall pooled log risk ratio for all studies was −0.29 (95% CI: [−0.67, 0.09]), indicating a slight reduction in CKD risk, with a significant difference between the groups (p = 0.00). Figure 68 compares the risk ratios for chronic kidney disease (CKD) occurrence in patients receiving daily versus weekly treatment with and without SGLT2 inhibitors. For daily administration, Marx et al. (2025) showed a significant reduction in CKD risk with a log risk ratio of −1.17 (95% CI: [−1.29, −1.06]), while Pfeffer et al. (2015) and Margulies et al. (2016) reported minimal effects. In the weekly subgroup, Packer et al. (2025) and Marso et al. (2016) observed smaller reductions in risk with log risk ratios of −0.03 (95% CI: [−0.05, −0.01]) and −0.35 (95% CI: [−0.43, −0.27]), respectively. The overall pooled log risk ratio was −0.29 (95% CI: [−0.67, 0.09]), with no significant difference between the groups (p = 0.70).

**Figure 67.**
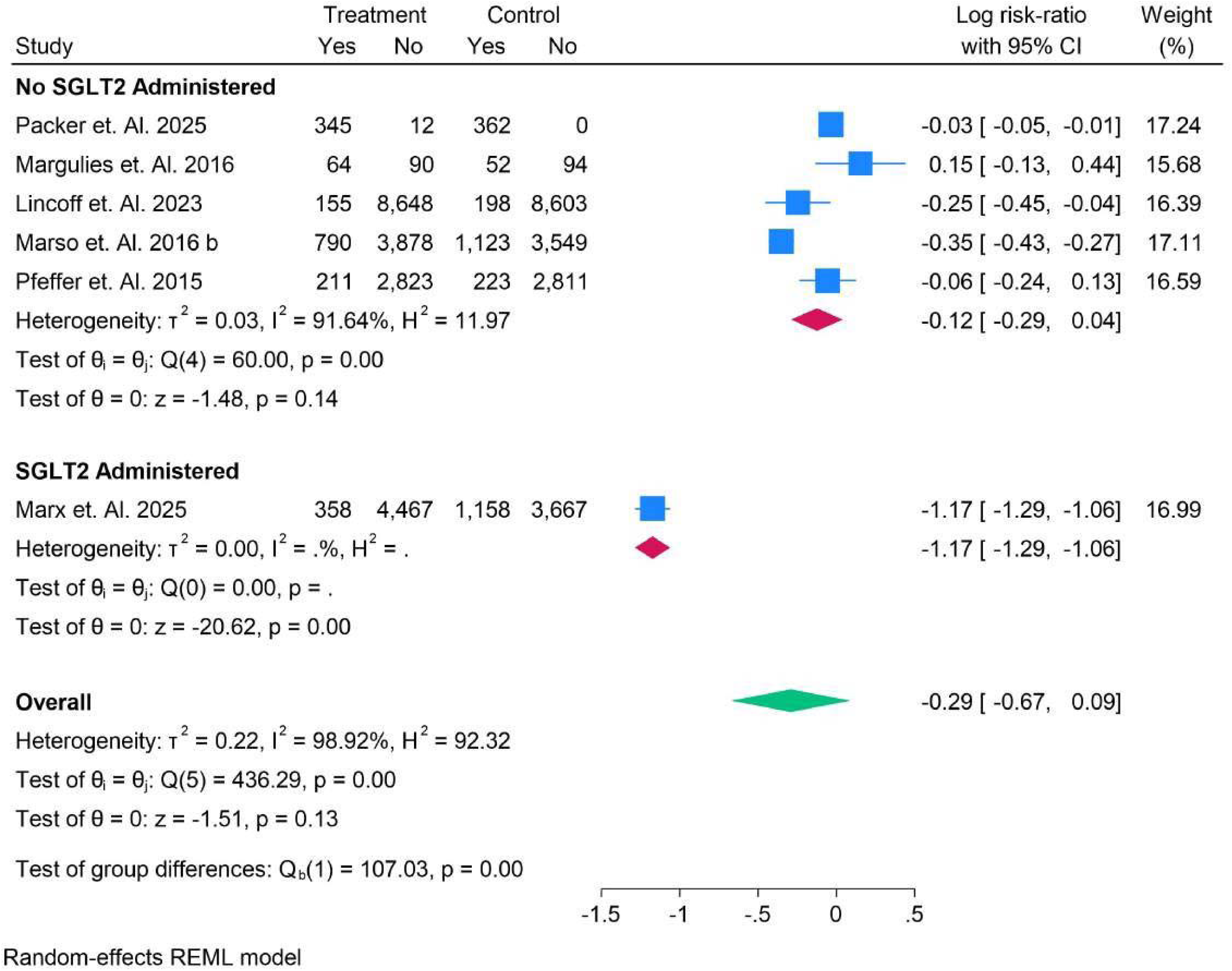
Risk Ratio of Occurrence of CKD in Patients having No SGLT2 vs SGLT Group.

**Figure 68.**
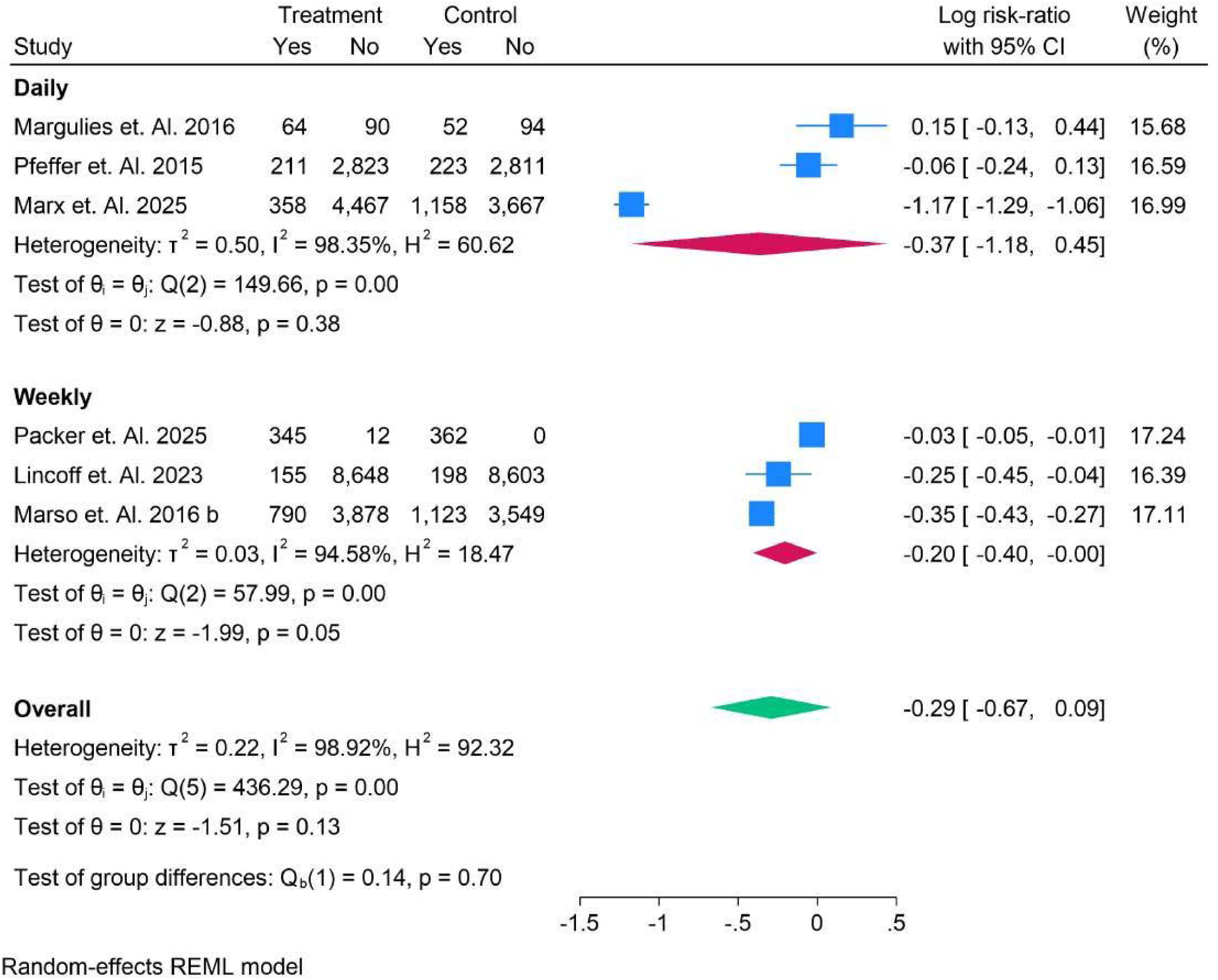
Risk Ratio of Occurrence of CKD in Patients having No SGLT2 vs SGLT Group.

Figure 69 compares the risk ratios for chronic kidney disease (CKD) occurrence in patients receiving oral versus subcutaneous (SC) treatments. For the oral group, Marx et al. (2025) reported a significant reduction in CKD risk with a log risk ratio of −1.17 (95% CI: [−1.29, −1.06]). In contrast, the SC group showed more moderate effects, with Packer et al. (2025) reporting a minimal risk reduction of −0.03 (95% CI: [−0.05, −0.01]). The overall pooled log risk ratio was −0.29 (95% CI: [−0.67, 0.09]), indicating no significant difference between the oral and SC treatment groups, with a significant test for group differences (p = 0.00).

**Figure 69.**
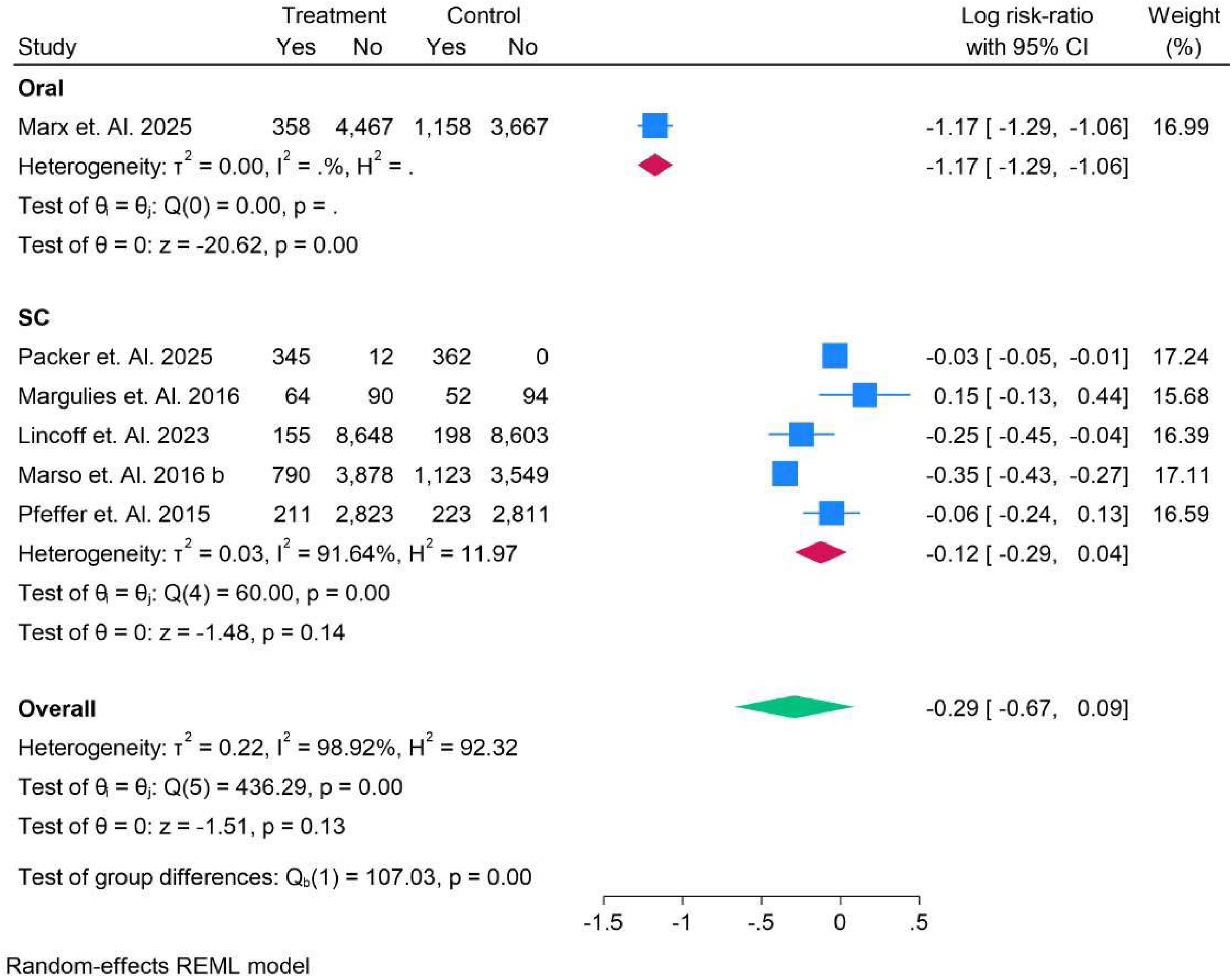
Risk Ratio of Occurrence of CKD in Patients Oral vs Sub-Cutaneous Group.

Figure 70 presents the risk ratios for chronic kidney disease (CKD) occurrence in patients using different medicines. For Liraglutide, Margulies et al. (2016) and Marso et al. (2016) showed minimal effects, with log risk ratios of 0.15 (95% CI: [−0.13, 0.44]) and −0.35 (95% CI: [−0.43, −0.27]), respectively. In the Lixisenatide group, Pfeffer et al. (2015) reported a log risk ratio of −0.06 (95% CI: [−0.24, 0.13]), indicating a minimal effect. Semaglutide, as seen in Lincoff et al. (2023) and Marx et al. (2025), showed larger effects with log risk ratios of −0.25 (95% CI: [−0.45, −0.04]) and −1.17 (95% CI: [−1.29, −1.06]). Tirzepatide, studied by Packer et al. (2025), showed a minimal effect (−0.03, 95% CI: [−0.05, −0.01]). The overall pooled log risk ratio was −0.29 (95% CI: [−0.67, 0.09]), suggesting no significant difference between the medicines (p = 0.51).

**Figure 70.**
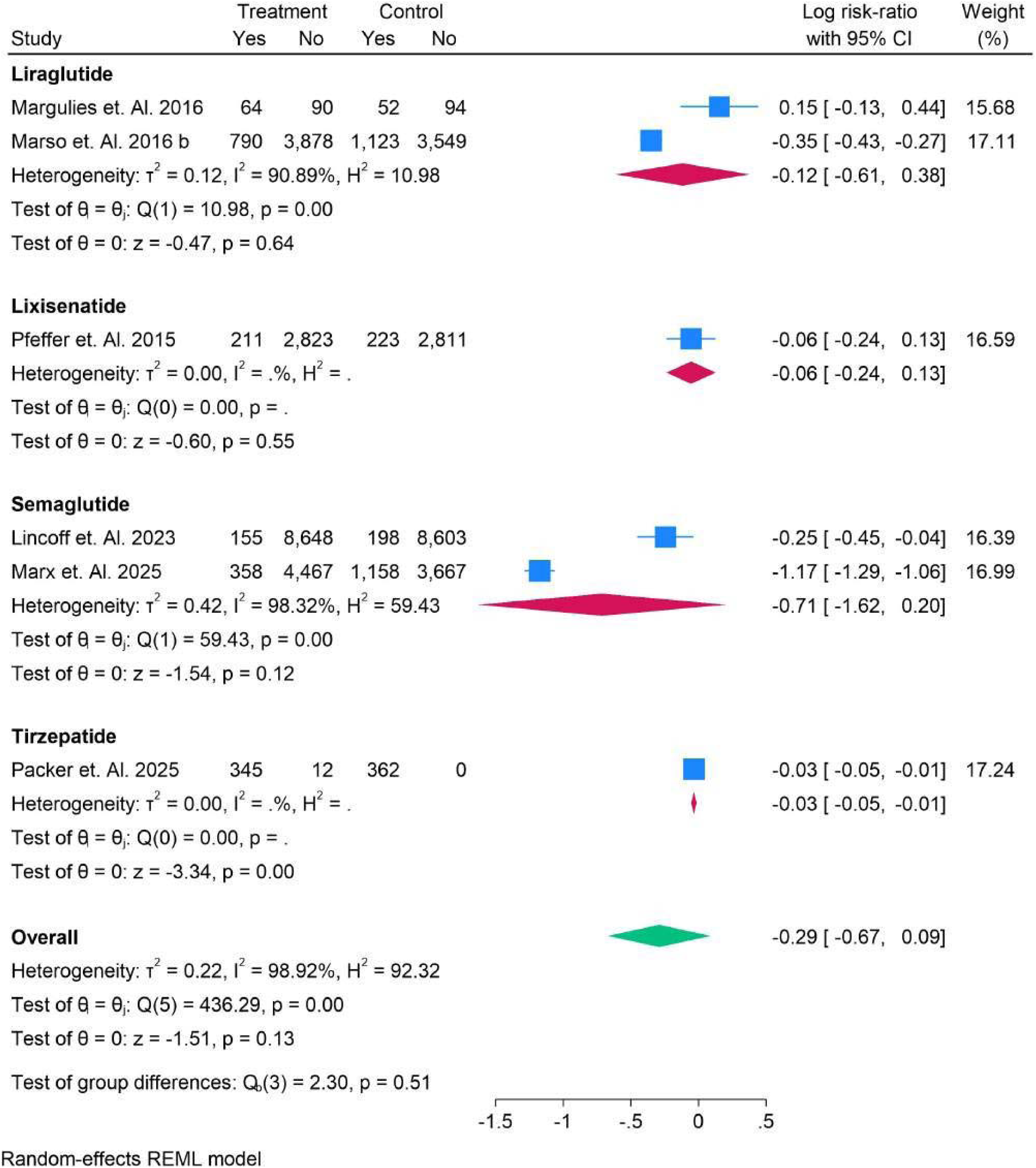
Risk Ratio of Occurrence of CKD in Patients All Medicines Group.

### Adverse Events

Figure 71 presents the risk ratios for gastrointestinal adverse events in patients across various studies. Notable studies include Gerstein et al. (2021), which reported a significant positive effect with a log risk ratio of 1.63 (95% CI: [1.47, 1.79]), and Marx et al. (2025), which showed a smaller but still relevant effect with −0.29 (95% CI: [−0.39, −0.18]). The overall pooled log risk ratio was 0.48 (95% CI: [−0.00, 0.96]), suggesting a slight increase in gastrointestinal adverse events, with significant heterogeneity (I² = 99.16%, H² = 118.41%) and a borderline significant test for group differences (p = 0.05).

**Figure 71.**
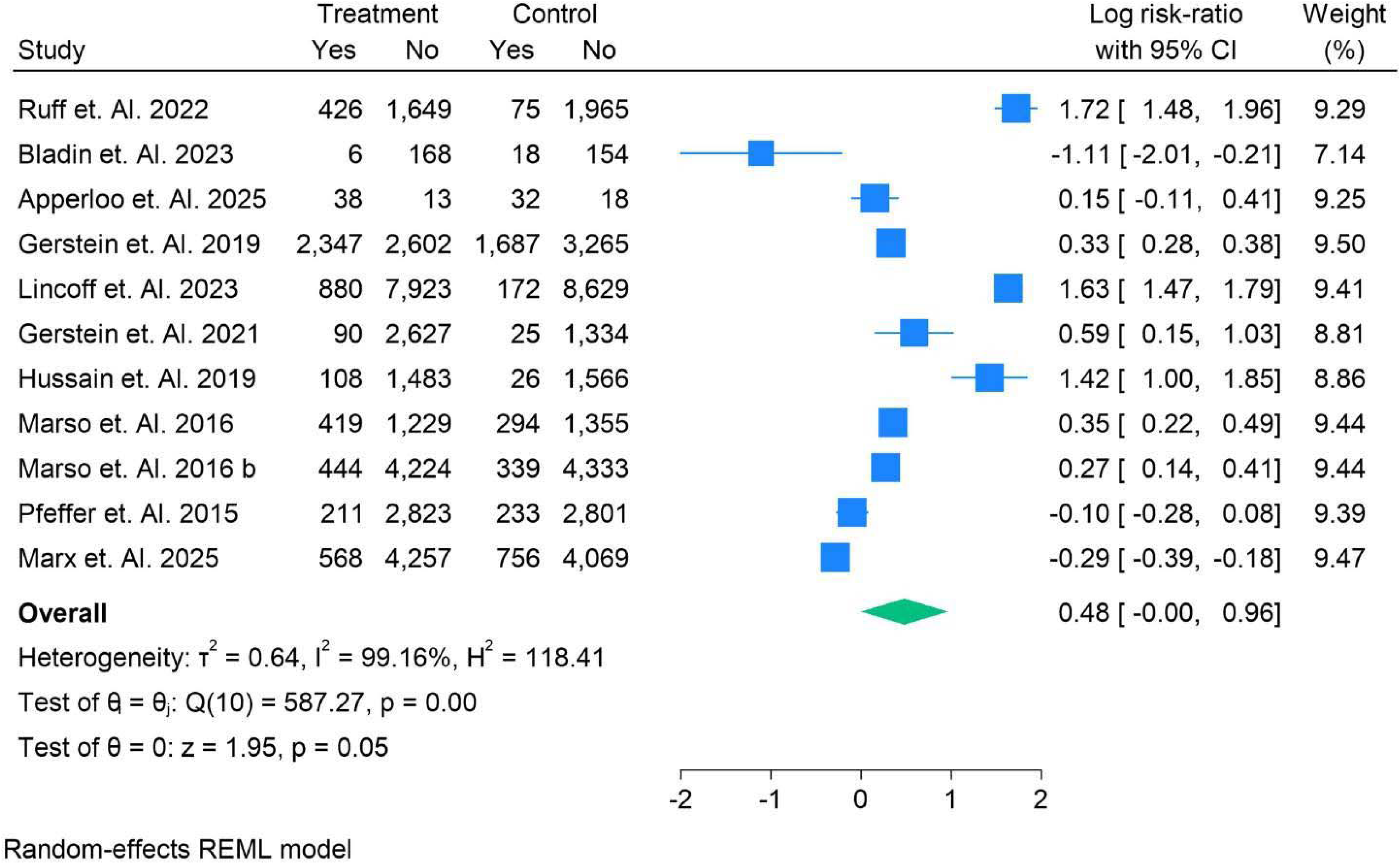
Risk Ratio of Occurrence of Gastro-Intestinal Adverse events in Patients.

Figure 72 displays the risk ratios for death events in patients across various studies. Ruff et al. (2022) observed a modest increase in risk with a log risk ratio of 0.17 (95% CI: [−0.12, 0.46]), while Gerstein et al. (2019) and Hernandez et al. (2019) found a significant reduction in death events, with log risk ratios of −0.44 (95% CI: [0.50, −0.38]) and −0.13 (95% CI: [−0.17, −0.08]), respectively. Marso et al. (2016) and other studies, like Marx et al. (2025), reported smaller reductions or minimal effects, such as −0.30 (95% CI: [−0.54, −0.06]) and −0.14 (95% CI: [−0.25, −0.04]). The overall pooled log risk ratio was −0.16 (95% CI: [−0.24, −0.07]), indicating a slight but statistically significant reduction in death events. The significant heterogeneity (I² = 85.20%, H² = 6.76) and the significant test for group differences (p = 0.00) suggest variability in the effects across different studies.

**Figure 72.**
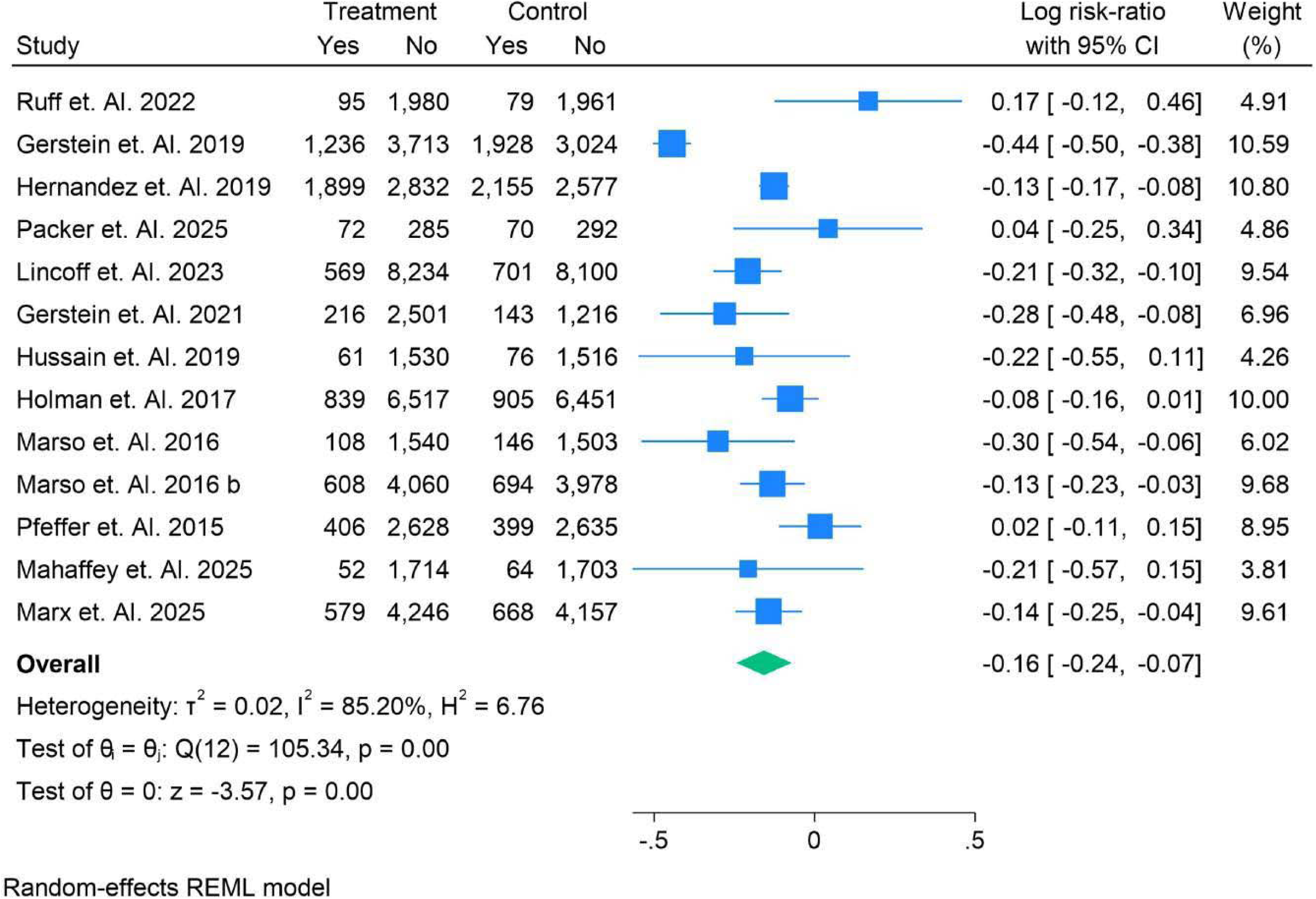
Risk Ratio of Occurrence of Deaths events in Patients.

Figure 73 shows the risk ratios for major adverse cardiovascular events (MACE) in patients from several studies.

**Figure 73.**
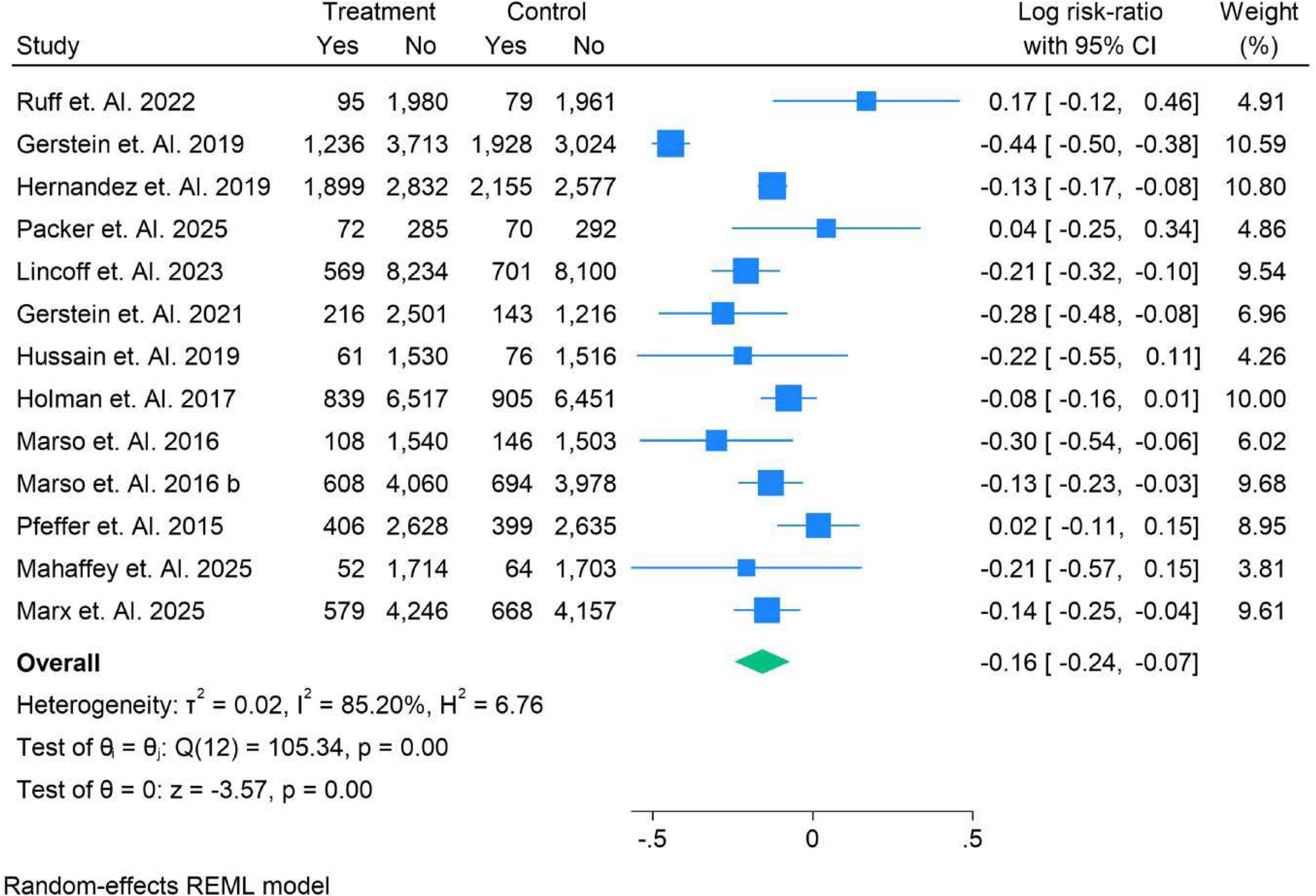
Risk Ratio of Occurrence of MACE events in Patients.

Notably, Ruff et al. (2022) reported a slight increase in MACE risk with a log risk ratio of 0.17 (95% CI: [−0.12, 0.46]). Gerstein et al. (2019) observed a reduction in risk, with a log risk ratio of −0.44 (95% CI: [−0.50, −0.38]), while Hernandez et al. (2019) found a smaller reduction of −0.13 (95% CI: [−0.17, −0.08]). Marso et al. (2016) and other studies, such as Marx et al. (2025), reported further reductions in risk, with log risk ratios of −0.30 (95% CI: [−0.54, −0.06]) and −0.14 (95% CI: [−0.25, −0.04]). The overall pooled log risk ratio was −0.16 (95% CI: [0.24, − 0.07]), suggesting a slight, statistically significant reduction in MACE risk. The high heterogeneity (I² = 85.20%, H² = 6.76) and significant test for group differences (p = 0.00) further indicate variability in the outcomes across studies.

## Discussion

Type 2 diabetes (T2D) is a growing global health concern, and its complications, particularly neurological and nephrological, contribute significantly to the morbidity and mortality of affected individuals. GLP-1 receptor agonists (GLP-1 RAs) have emerged as a promising class of drugs not only for glycemic control but also for their potential neuroprotective and nephroprotective effects. This systematic review and network meta-analysis (NMA) evaluate the efficacy and safety of GLP-1 RAs on neurological and nephrological outcomes, providing valuable insights into their comparative effectiveness.

The results of this analysis demonstrate that GLP-1 RAs, particularly liraglutide, semaglutide, and tirzepatide, exhibit significant benefits in managing diabetic complications such as diabetic peripheral neuropathy (DPN), cognitive decline, and diabetic nephropathy (DN). Specifically, GLP-1 RAs were found to improve neurological outcomes, including enhancing cognitive function, reducing neuropathic pain, and improving nerve conduction velocity in patients with DPN. These findings are consistent with previous studies that have highlighted the neuroprotective potential of GLP-1 RAs [24]. Moreover, the data suggest that these drugs can reduce albuminuria and improve glomerular filtration rate (GFR), mitigating the progression of DN, a leading cause of end-stage renal disease.

The neuroprotective and nephroprotective effects of GLP-1 RAs may be attributed to their pleiotropic actions, including anti-inflammatory, antioxidant, and vasodilatory effects, mediated through the activation of GLP-1 receptors in the central nervous system and kidneys [25]. These effects have been shown to counteract the deleterious impact of hyperglycemia on both neuronal and renal tissues. While individual studies have highlighted the renoprotective effects of GLP-1 RAs, such as liraglutide and semaglutide, this NMA synthesizes evidence from multiple trials, reinforcing the beneficial impact of these agents on renal function and progression of DN.

The results also show that tirzepatide, a dual GLP-1 and GIP (glucose-dependent insulinotropic polypeptide) receptor agonist, may offer more substantial improvements in GFR compared to other GLP-1 RAs. This finding aligns with recent studies suggesting that tirzepatide, due to its dual mechanism of action, may provide superior metabolic and organ-protective benefits compared to single GLP-1 receptor agonists [26]. However, the variability in treatment effects across studies, reflected by high heterogeneity in the data, underscores the need for further research to optimize treatment regimens for specific patient populations, including those with differing comorbidities and demographic factors.

Subgroup analyses in this NMA revealed no significant differences in treatment effects based on age, gender, ethnicity, or baseline HbA1c levels for the outcomes assessed. These findings suggest that GLP-1 RAs provide consistent benefits across diverse patient populations. However, the data also highlighted some variations in the effect size, particularly in studies evaluating eGFR. Older patients and those with higher baseline HbA1c levels appeared to experience more substantial improvements in renal outcomes, consistent with previous findings that suggest these patients may benefit the most from GLP-1 RA therapy [27]. Moreover, the analysis demonstrated that the frequency of administration (daily vs. weekly) and the route of administration (oral vs. subcutaneous) did not significantly affect overall treatment outcomes, suggesting that the choice of administration schedule or route may not be as critical in determining the efficacy of GLP-1 RAs.

The safety profile of GLP-1 RAs, as assessed by adverse events, revealed a slight increase in gastrointestinal events, particularly nausea and vomiting, consistent with previous reports [28]. However, these events were generally mild and transient, with few patients discontinuing treatment due to adverse effects. Importantly, the incidence of severe adverse events, including fatal stroke or acute kidney injury (AKI), was minimal, indicating that GLP-1 RAs are generally well-tolerated.

When compared to other meta-analyses, our findings align with the work of a study, which also reported significant improvements in both neurological and nephrological outcomes with GLP-1 Ras [29]. However, our NMA provides a more granular comparison of individual drugs within the class and includes a broader range of outcomes, such as stroke risk and CKD progression. The study did not include a network meta-analysis design, which limits direct comparisons between treatments. Our analysis, by contrast, utilized Bayesian methods to estimate the relative treatment effects of various GLP-1 RAs, providing more robust estimates and ranking treatments according to their efficacy and safety.

Our study also builds on the work of other systematic reviews, such as that by Khunti et al. (2015), which examined the effects of GLP-1 RAs on cardiovascular outcomes. Although our NMA focuses on the neurological and nephrological outcomes of T2D, the positive effects of GLP-1 RAs on cardiovascular events observed in these previous studies are consistent with our findings of minimal adverse cardiovascular outcomes, such as major adverse cardiovascular events (MACE) and fatal stroke, among patients treated with GLP-1 RAs.

These findings highlight the potential broader benefits of GLP-1 RAs beyond glycemic control. Conclusion

In conclusion, GLP-1 RAs represent a promising therapeutic option for T2D patients, not only for glycemic control but also for mitigating the neurological and nephrological complications of the disease. This systematic review and NMA provide strong evidence supporting the efficacy of GLP-1 RAs in improving both neurological and nephrological outcomes, with consistent safety results. However, variability in treatment effects across subgroups and studies suggests the need for further exploration into personalized treatment approaches, including optimal dosing schedules and patient selection criteria, to maximize the benefits of GLP1 RAs for T2D patients at risk of complications.

## Authors’ contributions

All authors contributed equally to the manuscript and read and approved the final version of the manuscript.

## Data Availability

supplememtary file

## References

1 Hamed K, Alosaimi MN, Ali BA, Alghamdi A, Alkhashi T, Alkhaldi SS, Altowarqi NA, Alzahrani H, Alshehri AM, Alkhaldi RK, Alqahtani KW, Alharbi NH, Alhulayfi HF, Sharifi SY, Dighriri IM. Glucagon-Like Peptide-1 (GLP-1) Receptor Agonists: Exploring Their Impact on Diabetes, Obesity, and Cardiovascular Health Through a Comprehensive Literature Review. Cureus. 2024 Sep 1;16(9):e68390. doi: 10.7759/cureus.68390. PMID: 39355484; PMCID: PMC11444311.

2 Nadkarni P, Chepurny OG, Holz GG. Regulation of glucose homeostasis by GLP-1. Prog Mol Biol Transl Sci. 2014;121:23–65. doi: 10.1016/B978-0-12-800101-1.00002-8. PMID: 24373234; PMCID: PMC4159612.

3 Diz-Chaves Y, Mastoor Z, Spuch C, González-Matías LC, Mallo F. Anti-Inflammatory Effects of GLP-1 Receptor Activation in the Brain in Neurodegenerative Diseases. Int J Mol Sci. 2022 Aug 24;23(17):9583. doi: 10.3390/ijms23179583. PMID: 36076972; PMCID: PMC9455625.

4 Dou X, Zhao L, Li J, Jiang Y. Effect and mechanism of GLP-1 on cognitive function in diabetes mellitus. Front Neurosci. 2025 Mar 18;19:1537898. doi: 10.3389/fnins.2025.1537898. PMID: 40171533; PMCID: PMC11959055.

5 Rojano Toimil A, Ciudin A. GLP-1 Receptor Agonists in Diabetic Kidney Disease: From Physiology to Clinical Outcomes. J Clin Med. 2021 Aug 31;10(17):3955. doi: 10.3390/jcm10173955. PMID: 34501404; PMCID: PMC8432108.

6 Page MJ, McKenzie JE, Bossuyt PM, Boutron I, Hoffmann TC, Mulrow CD, Shamseer L, Tetzlaff JM, Akl EA, Brennan SE, Chou R, Glanville J, Grimshaw JM, Hróbjartsson A, Lalu MM, Li T, Loder EW, Mayo-Wilson E, McDonald S, McGuinness LA, Stewart LA, Thomas J, Tricco AC, Welch VA, Whiting P, Moher D. The PRISMA 2020 statement: an updated guideline for reporting systematic reviews. BMJ. 2021 Mar 29;372:n71. doi: 10.1136/bmj.n71. PMID: 33782057; PMCID: PMC8005924.

7 Nejadghaderi SA, Balibegloo M, Rezaei N. The Cochrane risk of bias assessment tool 2 (RoB 2) versus the original RoB: A perspective on the pros and cons. Health Sci Rep. 2024 Jun 3;7(6):e2165. doi: 10.1002/hsr2.2165. PMID: 38835932; PMCID: PMC11147813.

8 Ruff CT, Baron M, Im K, O’Donoghue ML, Fiedorek FT, Sabatine MS. Subcutaneous infusion of exenatide and cardiovascular outcomes in type 2 diabetes: a non-inferiority randomized controlled trial. Nat Med. 2022 Jan;28(1):89–95. doi: 10.1038/s41591-021-01584-3. Epub 2021 Dec 6. PMID: 34873344.

9 Bladin CF, Wah Cheung N, Dewey HM, Churilov L, Middleton S, Thijs V, Ekinci E, Levi CR, Lindley R, Donnan GA, Parsons MW, Meretoja A, Tiainen M, Choi PMC, Cordato D, Brown H, Campbell BCV, Davis SM, Cloud G, Grimley R, Lee-Archer M, Ghia D, Sanders L, Markus R, Muller C, Salvaris P, Wu T, Fink J; TEXAIS Investigators. Management of Poststroke Hyperglycemia: Results of the TEXAIS Randomized Clinical Trial. Stroke. 2023 Dec;54(12):2962–2971. doi: 10.1161/STROKEAHA.123.044568. Epub 2023 Nov 27. PMID: 38011235; PMCID: PMC10664794.

10 Apperloo EM, Gorriz JL, Soler MJ, Cigarrán Guldris S, Cruzado JM, Puchades MJ, López-Martínez M, Waanders F, Laverman GD, van der Aart-van der Beek A, Hoogenberg K, van Beek AP, Verhave J, Ahmed SB, Schmieder RE, Wanner C, Cherney DZI, Jongs N, Heerspink HJL. Semaglutide in patients with overweight or obesity and chronic kidney disease without diabetes: a randomized double-blind placebo-controlled clinical trial. Nat Med. 2025 Jan;31(1):278–285. doi: 10.1038/s41591-024-03327-6. Epub 2024 Oct 25. PMID: 39455729.

11 Gerstein HC, Colhoun HM, Dagenais GR, Diaz R, Lakshmanan M, Pais P, Probstfield J, Riesmeyer JS, Riddle MC, Rydén L, Xavier D, Atisso CM, Dyal L, Hall S, Rao-Melacini P, Wong G, Avezum A, Basile J, Chung N, Conget I, Cushman WC, Franek E, Hancu N, Hanefeld M, Holt S, Jansky P, Keltai M, Lanas F, Leiter LA, Lopez-Jaramillo P, Cardona Munoz EG, Pirags V, Pogosova N, Raubenheimer PJ, Shaw JE, Sheu WH, Temelkova-Kurktschiev T; REWIND Investigators. Dulaglutide and cardiovascular outcomes in type 2 diabetes (REWIND): a double-blind, randomised placebo-controlled trial. Lancet. 2019 Jul 13;394(10193):121–130. doi: 10.1016/S0140-6736(19)31149-3. Epub 2019 Jun 9. PMID: 31189511.

12 Hernandez AF, Green JB, Janmohamed S, D’Agostino RB Sr, Granger CB, Jones NP, Leiter LA, Rosenberg AE, Sigmon KN, Somerville MC, Thorpe KM, McMurray JJV, Del Prato S; Harmony Outcomes committees and investigators. Albiglutide and cardiovascular outcomes in patients with type 2 diabetes and cardiovascular disease (Harmony Outcomes): a double-blind, randomised placebo-controlled trial. Lancet. 2018 Oct 27;392(10157):1519–1529. doi: 10.1016/S0140-6736(18)32261-X. Epub 2018 Oct 2. PMID: 30291013.

13 Packer M, Zile MR, Kramer CM, Murakami M, Ou Y, Borlaug BA; SUMMIT Trial Study Group. Interplay of Chronic Kidney Disease and the Effects of Tirzepatide in Patients With Heart Failure, Preserved Ejection Fraction, and Obesity: The SUMMIT Trial. J Am Coll Cardiol. 2025 May 13;85(18):1721–1735. doi: 10.1016/j.jacc.2025.03.009. Epub 2025 Mar 31. PMID: 40162940.

14 Margulies KB, Hernandez AF, Redfield MM, Givertz MM, Oliveira GH, Cole R, Mann DL, Whellan DJ, Kiernan MS, Felker GM, McNulty SE, Anstrom KJ, Shah MR, Braunwald E, Cappola TP; NHLBI Heart Failure Clinical Research Network. Effects of Liraglutide on Clinical Stability Among Patients With Advanced Heart Failure and Reduced Ejection Fraction: A Randomized Clinical Trial. JAMA. 2016 Aug 2;316(5):500–8. doi: 10.1001/jama.2016.10260. PMID: 27483064; PMCID: PMC5021525.

15 Lincoff AM, Brown-Frandsen K, Colhoun HM, Deanfield J, Emerson SS, Esbjerg S, Hardt-Lindberg S, Hovingh GK, Kahn SE, Kushner RF, Lingvay I, Oral TK, Michelsen MM, Plutzky J, Tornøe CW, Ryan DH; SELECT Trial Investigators. Semaglutide and Cardiovascular Outcomes in Obesity without Diabetes. N Engl J Med. 2023 Dec 14;389(24):2221–2232. doi: 10.1056/NEJMoa2307563. Epub 2023 Nov 11. PMID: 37952131.

16 Gerstein HC, Sattar N, Rosenstock J, Ramasundarahettige C, Pratley R, Lopes RD, Lam CSP, Khurmi NS, Heenan L, Del Prato S, Dyal L, Branch K; AMPLITUDE-O Trial Investigators. Cardiovascular and Renal Outcomes with Efpeglenatide in Type 2 Diabetes. N Engl J Med. 2021 Sep 2;385(10):896–907. doi: 10.1056/NEJMoa2108269. Epub 2021 Jun 28. PMID: 34215025.

17 Husain M, Birkenfeld AL, Donsmark M, Dungan K, Eliaschewitz FG, Franco DR, Jeppesen OK, Lingvay I, Mosenzon O, Pedersen SD, Tack CJ, Thomsen M, Vilsbøll T, Warren ML, Bain SC; PIONEER 6 Investigators. Oral Semaglutide and Cardiovascular Outcomes in Patients with Type 2 Diabetes. N Engl J Med. 2019 Aug 29;381(9):841–851. doi: 10.1056/NEJMoa1901118. Epub 2019 Jun 11. PMID: 31185157.

18 Holman RR, Bethel MA, Mentz RJ, Thompson VP, Lokhnygina Y, Buse JB, Chan JC, Choi J, Gustavson SM, Iqbal N, Maggioni AP, Marso SP, Öhman P, Pagidipati NJ, Poulter N, Ramachandran A, Zinman B, Hernandez AF; EXSCEL Study Group. Effects of Once-Weekly Exenatide on Cardiovascular Outcomes in Type 2 Diabetes. N Engl J Med. 2017 Sep 28;377(13):1228–1239. doi: 10.1056/NEJMoa1612917. Epub 2017 Sep 14. PMID: 28910237; PMCID: PMC9792409.

19 Marso SP, Bain SC, Consoli A, Eliaschewitz FG, Jódar E, Leiter LA, Lingvay I, Rosenstock J, Seufert J, Warren ML, Woo V, Hansen O, Holst AG, Pettersson J, Vilsbøll T; SUSTAIN-6 Investigators. Semaglutide and Cardiovascular Outcomes in Patients with Type 2 Diabetes. N Engl J Med. 2016 Nov 10;375(19):1834–1844. doi: 10.1056/NEJMoa1607141. Epub 2016 Sep 15. PMID: 27633186.

20 Marso SP, Daniels GH, Brown-Frandsen K, Kristensen P, Mann JF, Nauck MA, Nissen SE, Pocock S, Poulter NR, Ravn LS, Steinberg WM, Stockner M, Zinman B, Bergenstal RM, Buse JB; LEADER Steering Committee; LEADER Trial Investigators. Liraglutide and Cardiovascular Outcomes in Type 2 Diabetes. N Engl J Med. 2016 Jul 28;375(4):311–22. doi: 10.1056/NEJMoa1603827. Epub 2016 Jun 13. PMID: 27295427; PMCID: PMC4985288.

21 Pfeffer MA, Claggett B, Diaz R, Dickstein K, Gerstein HC, Køber LV, Lawson FC, Ping L, Wei X, Lewis EF, Maggioni AP, McMurray JJ, Probstfield JL, Riddle MC, Solomon SD, Tardif JC; ELIXA Investigators. Lixisenatide in Patients with Type 2 Diabetes and Acute Coronary Syndrome. N Engl J Med. 2015 Dec 3;373(23):2247–57. doi: 10.1056/NEJMoa1509225. PMID: 26630143.

22 Mahaffey KW, Tuttle KR, Arici M, Baeres FMM, Bakris G, Charytan DM, Cherney DZI, Chernin G, Correa-Rotter R, Gumprecht J, Idorn T, Pugliese G, Rasmussen IKB, Rasmussen S, Rossing P, Sokareva E, Mann JFE, Perkovic V, Pratley R. Cardiovascular outcomes with semaglutide by severity of chronic kidney disease in type 2 diabetes: the FLOW trial. Eur Heart J. 2025 Mar 24;46(12):1096–1108. doi: 10.1093/eurheartj/ehae613. PMID: 39211948; PMCID: PMC11931213.

23 McGuire DK, Busui RP, Deanfield J, Inzucchi SE, Mann JFE, Marx N, Mulvagh SL, Poulter N, Engelmann MDM, Hovingh GK, Ripa MS, Gislum M, Brown-Frandsen K, Buse JB. Effects of oral semaglutide on cardiovascular outcomes in individuals with type 2 diabetes and established atherosclerotic cardiovascular disease and/or chronic kidney disease: Design and baseline characteristics of SOUL, a randomized trial. Diabetes Obes Metab. 2023 Jul;25(7):1932–1941. doi: 10.1111/dom.15058. Epub 2023 Apr 11. PMID: 36945734.

24 Marx N, Deanfield JE, Mann JFE, Arechavaleta R, Bain SC, Bajaj HS, Bayer Tanggaard K, Birkenfeld AL, Buse JB, Davicevic-Elez Z, Desouza C, Emerson SS, Engelmann MDM, Hovingh GK, Inzucchi SE, Jhund PS, Mulvagh SL, Pop-Busui R, Poulter NR, Rasmussen S, Tu ST, McGuire DK; SOUL Study Group. Oral Semaglutide and Cardiovascular Outcomes in People With Type 2 Diabetes, According to SGLT2i Use: Prespecified Analyses of the SOUL Randomized Trial. Circulation. 2025 Jun 10;151(23):1639–1650. doi: 10.1161/CIRCULATIONAHA.125.074545. Epub 2025 Mar 29. PMID: 40156843; PMCID: PMC12144549.

25 Thomas MK, Nikooienejad A, Bray R, Cui X, Wilson J, Duffin K, Milicevic Z, Haupt A, Robins DA. Dual GIP and GLP-1 Receptor Agonist Tirzepatide Improves Beta-cell Function and Insulin Sensitivity in Type 2 Diabetes. J Clin Endocrinol Metab. 2021 Jan 23;106(2):388–396. doi: 10.1210/clinem/dgaa863. PMID: 33236115; PMCID: PMC7823251.

26 Min T, Bain SC. The Role of Tirzepatide, Dual GIP and GLP-1 Receptor Agonist, in the Management of Type 2 Diabetes: The SURPASS Clinical Trials. Diabetes Ther. 2021 Jan;12(1):143-157. doi: 10.1007/s13300020-00981-0. Epub 2020 Dec 15. PMID: 33325008; PMCID: PMC7843845.

27 Gorgojo-Martínez JJ, Mezquita-Raya P, Carretero-Gómez J, Castro A, Cebrián-Cuenca A, de Torres-Sánchez A, García-de-Lucas MD, Núñez J, Obaya JC, Soler MJ, Górriz JL, Rubio-Herrera MÁ. Clinical Recommendations to Manage Gastrointestinal Adverse Events in Patients Treated with Glp-1 Receptor Agonists: A Multidisciplinary Expert Consensus. J Clin Med. 2022 Dec 24;12(1):145. doi: 10.3390/jcm12010145. PMID: 36614945; PMCID: PMC9821052.

28 Mendonça L, Moura H, Chaves PC, Neves JS, Ferreira JP. The Impact of Glucagon-Like Peptide-1 Receptor Agonists on Kidney Outcomes: A Meta-Analysis of Randomized Placebo-Controlled Trials. Clin J Am Soc Nephrol. 2024 Oct 8;20(2):159–68. doi: 10.2215/CJN.0000000584. Epub ahead of print. PMID: 39480988; PMCID: PMC11835157.

29 Westermeier F, Fisman EZ. Glucagon like peptide-1 (GLP-1) agonists and cardiometabolic protection: historical development and future challenges. Cardiovasc Diabetol. 2025 Jan 29;24(1):44. doi: 10.1186/s12933025-02608-9. Erratum in: Cardiovasc Diabetol. 2025 Mar 12;24(1):118. doi: 10.1186/s12933-025-02647-2. PMID: 39881322; PMCID: PMC11781064.

