## supplementary file for "Comparative Efficacy and Safety of GLP-1 Receptor Agonists in Neurological and Nephrological Outcomes of Type 2 Diabetes: A Systematic Review and Network MetaAnalysis"

Search String

("GLP-1 receptor agonists" OR "Glucagon-Like Peptide-1 agonists" OR "GLP-1 analogs" OR "liraglutide" OR "semaglutide" OR "exenatide" OR "dulaglutide") AND

("Type 2 Diabetes" OR "T2DM" OR "Diabetes Mellitus, Type 2") AND

("neurological outcomes" OR "neuroprotection" OR "neuropathy" OR "cognitive function" OR "stroke" OR "dementia" OR "neurodegeneration") AND

("nephrological outcomes" OR "renal function" OR "kidney disease" OR "chronic kidney disease" OR "diabetic nephropathy" OR "glomerular filtration rate" OR "albuminuria") AND

("efficacy" OR "safety" OR "treatment outcomes" OR "clinical trial" OR "systematic review" OR "network meta-analysis") AND

("randomized controlled trial" OR "RCT" OR "observational study" OR "meta-analysis") AND

("English" OR "human studies")

Table S1. Demographics and Summary Table

| Ref no | Author Name | Country of Trial | Total Population | Male | Female | Mean Age | Race | Type of Diabetes | Rx Name | Rx Number | Cx Name | Cx number | GRADE | Main Finding |
| --- | --- | --- | --- | --- | --- | --- | --- | --- | --- | --- | --- | --- | --- | --- |
| 8 | Ruff et. Al. 2022 | Multicenter | 4156 | 2611 | 1545 | 63 | White, Black | T2DM | Exenatide | 2075 | Placebo | 2040 | High | **ITCA 650** was **non-inferior** to placebo in terms of cardiovascular safety in patients with type 2 diabetes and atherosclerotic cardiovascular disease (ASCVD). However, the study showed no statistically significant reduction in major adverse cardiovascular events (MACE) |
| 9 | Bladin et. Al. 2023 | Multicenter | 350 | 124 | 117 | 71 | White | T2DM | Exenatide | 174 | Placebo | 172 | High | Exenatide did not reduce neurological impairment at 7 days poststroke but reduced the frequency of hyperglycemic events |
| 10 | Apperloo et. Al. 2025 | Multicenter | 101 | 61 | 40 | 55.5 | White | T2DM | Semaglutide | 51 | Placebo | 50 | High | The main finding is that **semaglutide** reduced albuminuria and showed benefits in kidney function in patients with CKD and obesity, even without diabetes . |
| 11 | Gerstein et. Al. 2019 | Multicenter | 9901 | 4589 | 5312 | 66.2 | White | T2DM | Dulaglutide | 4949 | Placebo | 4952 | High | Dulaglutide reduces cardiovascular events compared to placebo in individuals with type 2 diabetes and either previous cardiovascular disease or cardiovascular risk factors . |
| 12 | Hernandez et. Al. 2019 | Multicenter | 9463 | 6569 | 2894 | 64.1 | Hispanic | T2DM | Albiglutide | 4731 | Placebo | 4732 | High | Albiglutide reduced the risk of major adverse cardiovascular events in patients with type 2 diabetes and cardiovascular disease . |
| 13 | Packer et. Al. 2025 | Multicenter | 731 | 281 | 450 | 70 | White | T2DM | Tirzepatide | 357 | Placebo | 362 | High | The main finding was that **tirzepatide** improved renal function and heart failure outcomes, especially in patients with **obesity and chronic kidney disease (CKD)** |
| 14 | Margulies et. Al. 2016 | USA | 300 | 154 | 146 | 61 | White | T2DM | Liraglutide | 154 | Placebo | 146 | High | The study found that liraglutide did not improve post-hospitalization clinical stability in patients with advanced heart failure and reduced ejection fraction |
| 15 | Lincoff et. Al. 2023 | Multicenter | 17604 | 12732 | 4872 | 61 | White | Normal | Semaglutide | 8803 | Placebo | 8801 | Moderate | Semaglutide at 2.4 mg weekly was superior to placebo in reducing the incidence of death from cardiovascular causes, nonfatal myocardial infarction, or nonfatal stroke in patients with obesity and cardiovascular disease but without diabetes |
| 16 | Gerstein et. Al. 2021 | Multicenter | 4076 | 2732 | 1344 | 64.5 | Asian | T2DM | Efpeglenatide | 2717 | Placebo | 1359 | High | Efpeglenatide reduced cardiovascular and renal risks in patients with type 2 diabetes and a history of cardiovascular disease or kidney disease . |
| 17 | Hussain et. Al. 2019 | Multicenter | 3138 | 2176 | 1007 | 66 | Hispanic | T2DM | Semaglutide | 1591 | Placebo | 1592 | High | The main finding was that **oral semaglutide** demonstrated **cardiovascular safety** similar to **placebo** and did not show an excess cardiovascular risk |
| 18 | Holman et. Al. 2017 | Multicenter | 14752 | 7376 | 7376 | 66 | White | T2DM | Exenatide | 7356 | Placebo | 7356 | Moderate | The main finding was that exenatide administered once-weekly was noninferior to placebo with respect to safety, but not superior in terms of efficacy regarding cardiovascular outcomes. |
| 19 | Marso et. Al. 2016 | Multicenter | 3297 | 2002 | 1295 | 64.5 | Caucasian | T2DM | Semaglutide | 1648 | Placebo | 1649 | High | Semaglutide significantly reduced the risk of cardiovascular death, nonfatal myocardial infarction, and nonfatal stroke compared to placebo in patients with type 2 diabetes at high cardiovascular risk. |
| 20 | Marso et. Al. 2016 b | Multicenter | 9340 | 6003 | 3337 | 57 | White | T2DM | Liraglutide | 4668 | Placebo | 4672 | Moderate | Liraglutide showed reduced cardiovascular events and overall mortality compared to placebo |
| 21 | Pfeffer et. Al. 2015 | Multicenter | 6068 | 4200 | 1861 | 60 | White | T2DM | Lixisenatide | 3034 | Placebo | 3034 | High | Lixisenatide did not reduce cardiovascular events compared to placebo but showed noninferiority (safe with respect to CV outcomes) |
| 22 | Mahaffey et. Al. 2025 | Multicenter | 3533 | 1720 | 1813 | 52 | White | T2DM | Semaglutide | 1766 | Placebo | 1767 | High | Semaglutide reduced major CV events and all-cause death in people with T2D and CKD across CKD severity levels |
| 23 | Marx et. Al. 2025 | Multicenter | 9650 | 6860 | 2790 | 66.1 | White | T2DM | Semaglutide | 4825 | Placebo | 4825 | High | Oral semaglutide reduced MACE by 14% compared to placebo in patients with T2D and ASCVD and/or CKD, with benefits consistent regardless of SGLT2i use |

Table S2. **Treatment effects for all studies: comparison of all treatment pairs for overall survival**

|  | Albiglutide | Dulaglutide | Efpeglenatide | Exenatide | Liraglutide | Lixisenatide | Placebo | Semaglutide | Tirzepatide |
| --- | --- | --- | --- | --- | --- | --- | --- | --- | --- |
| Albiglutide | Albiglutide | 1.14 (0.49, 2.68) | 2.99 (1.18, 7.63) | 1.04 (0.44, 1.97) | 1.58 (0.64, 3.1) | 1.08 (0.46, 2.55) | 1.02 (0.55, 1.88) | 1.22 (0.64, 2.43) | 1.67 (0.66, 4.28) |
| Dulaglutide | 0.88 (0.37, 2.05) | Dulaglutide | 2.62 (1.05, 6.57) | 0.92 (0.39, 1.69) | 1.39 (0.56, 2.66) | 0.95 (0.4, 2.19) | 0.89 (0.49, 1.61) | 1.07 (0.57, 2.11) | 1.47 (0.59, 3.69) |
| Efpeglenatide | 0.33 (0.13, 0.85) | 0.38 (0.15, 0.96) | Efpeglenatide | 0.35 (0.14, 0.73) | 0.52 (0.2, 1.12) | 0.36 (0.14, 0.91) | 0.34 (0.17, 0.69) | 0.41 (0.19, 0.89) | 0.56 (0.21, 1.53) |
| Exenatide | 0.96 (0.51, 2.27) | 1.09 (0.59, 2.57) | 2.9 (1.37, 7.31) | Exenatide | 1.51 (0.8, 2.86) | 1.04 (0.55, 2.44) | 0.98 (0.7, 1.68) | 1.17 (0.79, 2.24) | 1.62 (0.77, 4.09) |
| Liraglutide | 0.63 (0.32, 1.55) | 0.72 (0.38, 1.77) | 1.91 (0.89, 4.97) | 0.66 (0.35, 1.25) | Liraglutide | 0.68 (0.35, 1.68) | 0.64 (0.44, 1.19) | 0.77 (0.5, 1.57) | 1.07 (0.5, 2.8) |
| Lixisenatide | 0.93 (0.39, 2.2) | 1.06 (0.46, 2.49) | 2.76 (1.1, 7.07) | 0.96 (0.41, 1.83) | 1.46 (0.59, 2.87) | Lixisenatide | 0.94 (0.52, 1.74) | 1.13 (0.6, 2.25) | 1.55 (0.62, 3.94) |
| Placebo | 0.98 (0.53, 1.8) | 1.12 (0.62, 2.04) | 2.94 (1.45, 5.99) | 1.02 (0.59, 1.42) | 1.55 (0.84, 2.28) | 1.06 (0.58, 1.93) | Placebo | 1.2 (0.94, 1.61) | 1.64 (0.81, 3.37) |
| Semaglutide | 0.82 (0.41, 1.55) | 0.94 (0.47, 1.76) | 2.45 (1.12, 5.14) | 0.85 (0.45, 1.26) | 1.29 (0.64, 2) | 0.89 (0.44, 1.66) | 0.83 (0.62, 1.07) | Semaglutide | 1.37 (0.63, 2.9) |
| Tirzepatide | 0.6 (0.23, 1.53) | 0.68 (0.27, 1.7) | 1.79 (0.65, 4.86) | 0.62 (0.24, 1.31) | 0.93 (0.36, 2.01) | 0.65 (0.25, 1.62) | 0.61 (0.3, 1.24) | 0.73 (0.35, 1.59) | Tirzepatide |

Figure S1. SUCRA ranking for survival

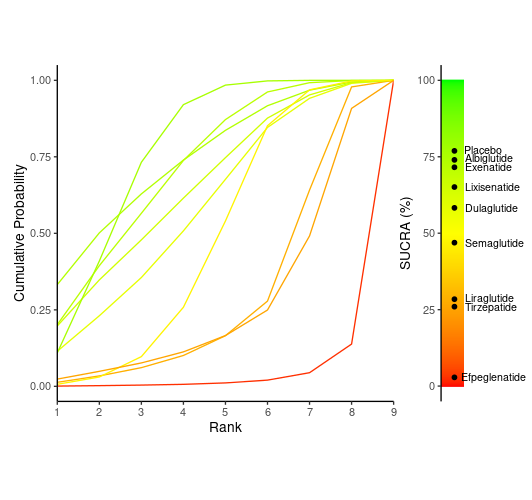

Figure S2. Rankogram for Survival

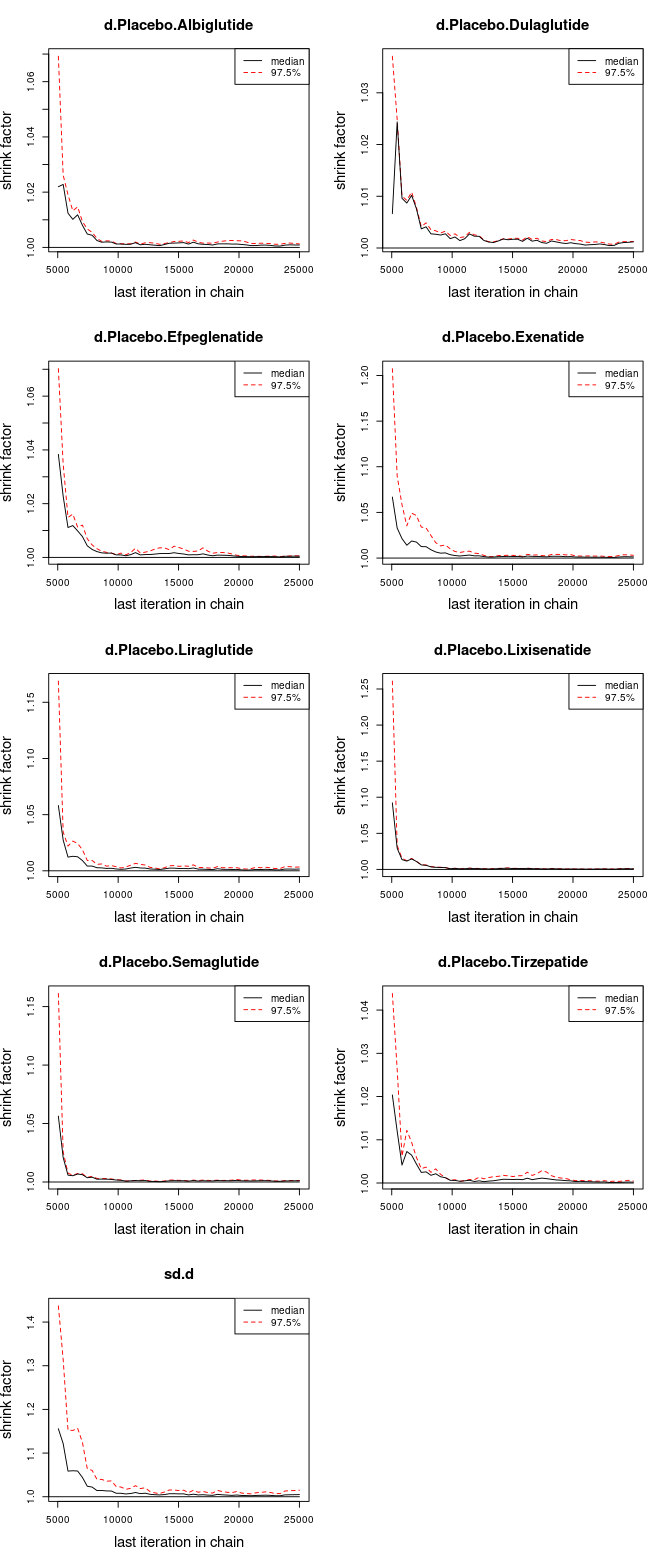

Figure S3. Overall Survival in Subgroup analysis – Funnel Plot.

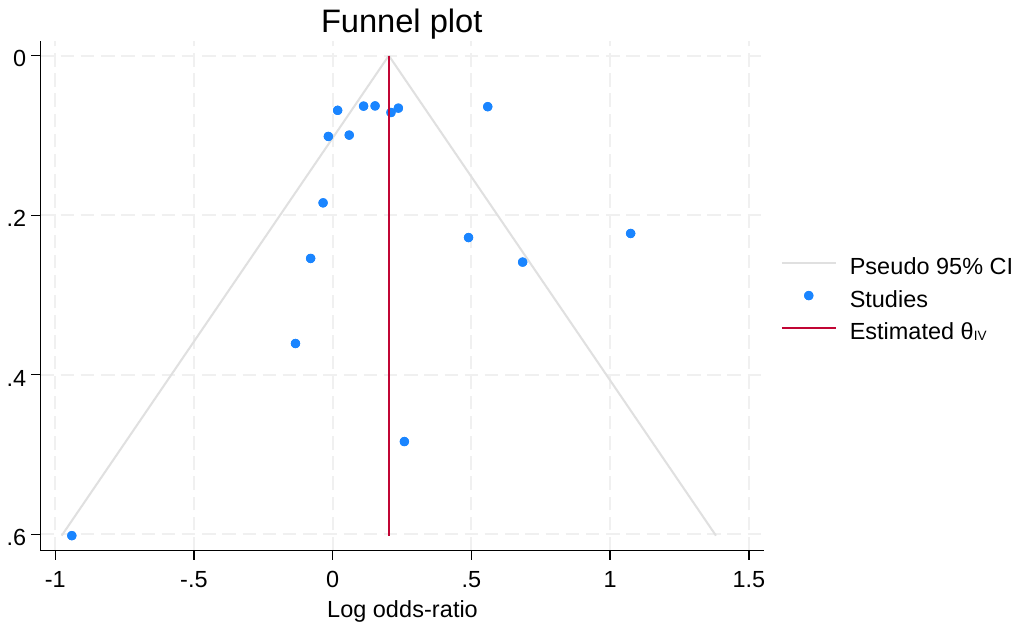

Figure S4. Overall Survival in Subgroup analysis of RACE Survivors – Funnel Plot

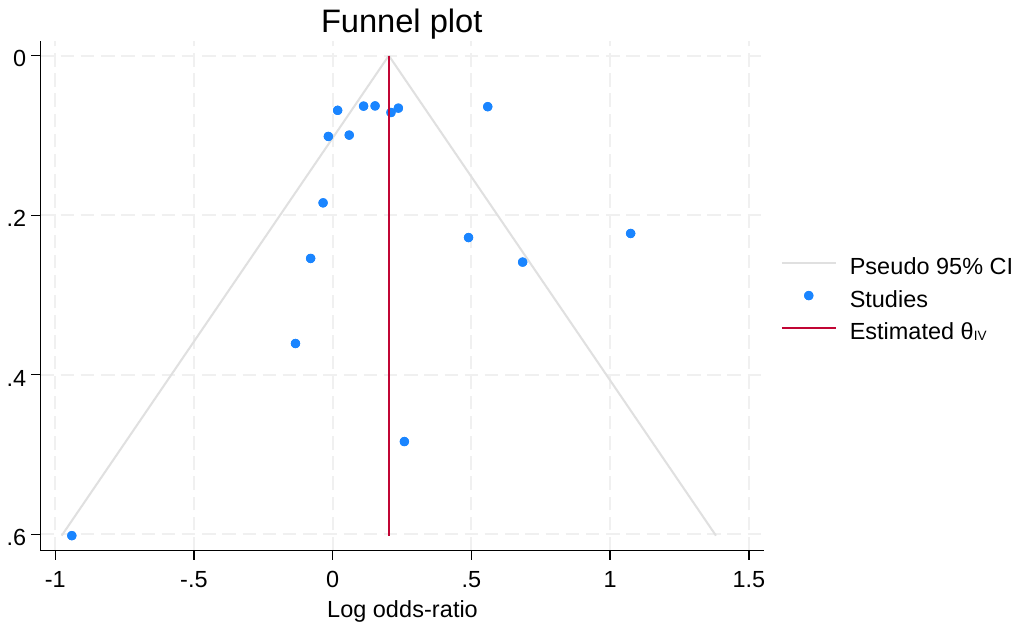

Figure S5. Overall Survival of Diabetic vs Non-Diabetic Patients Funnel Plot

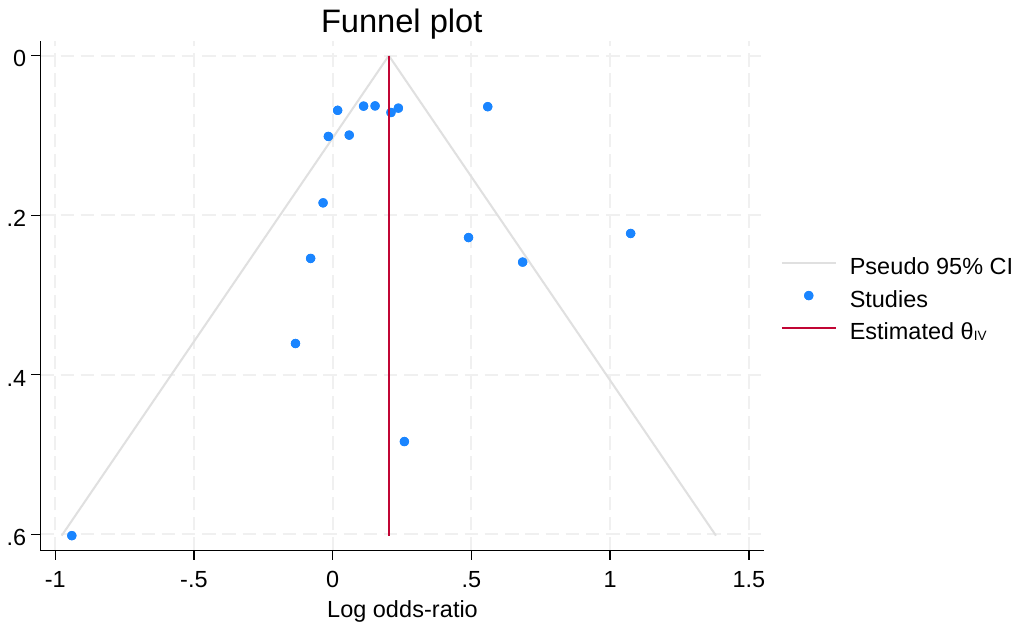

Figure S6. Overall Survival analysis – BMI Subgroup Analysis – Funnel Plot

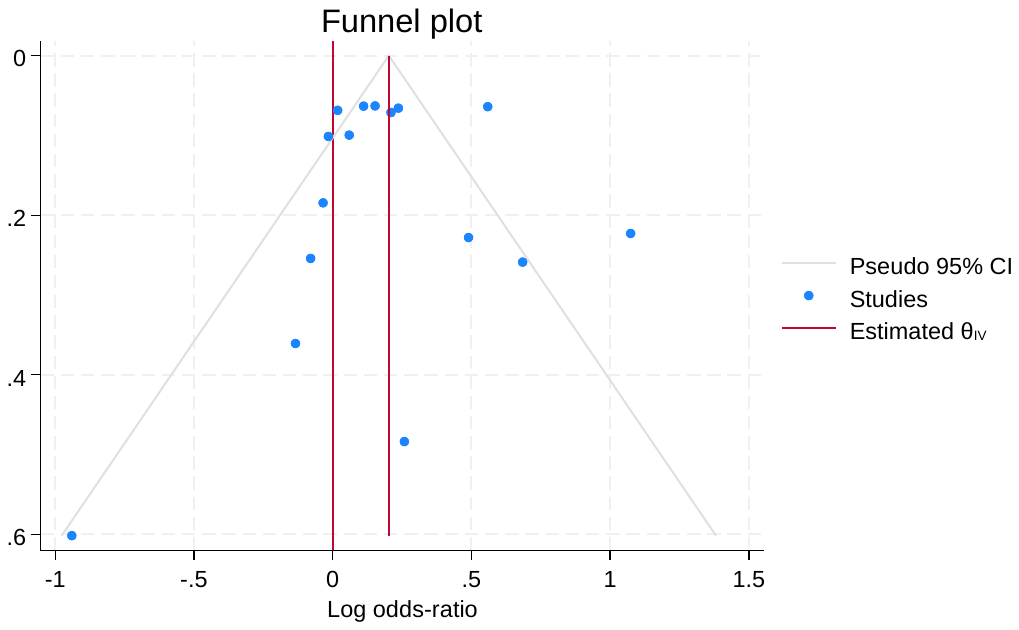

Figure S7. Overall Survival analysis – HBA1C Survivors – Funnel Plot

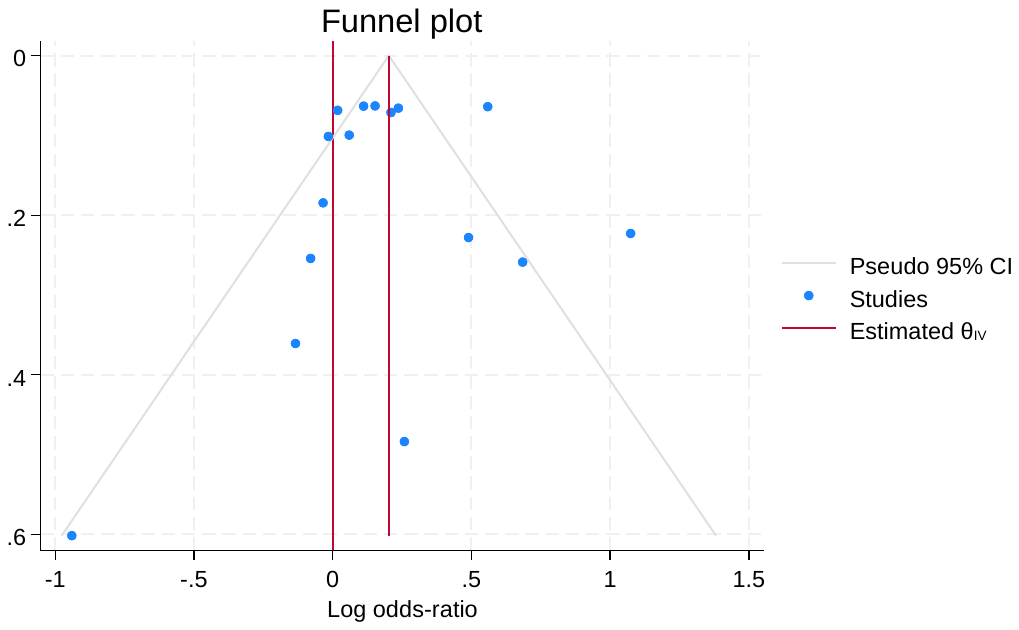

Figure S8. Overall Survival Analysis – SGLT2 Administration – Funnel Plot

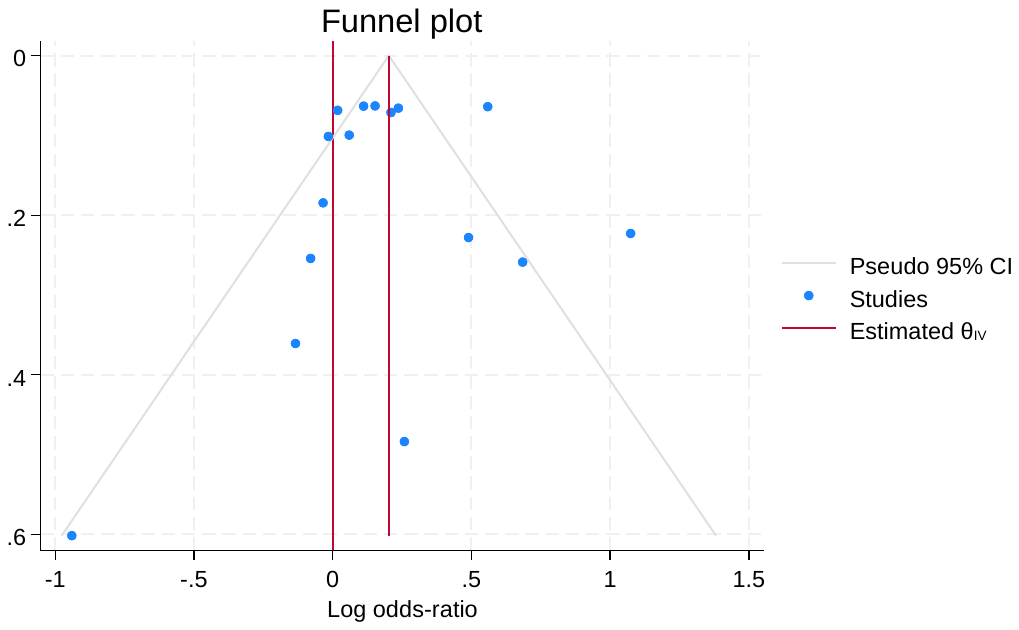

Figure S9. Daily vs Weekly administration funnel plot overall survivorship.

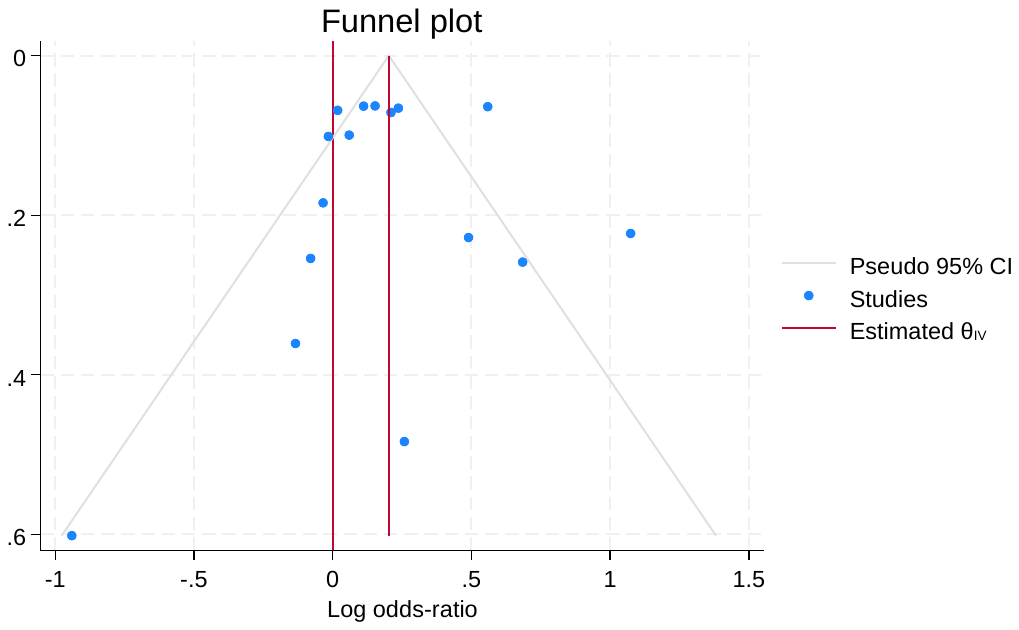

Figure S10. Overall Survivors for Route of Administration Funnel Plot

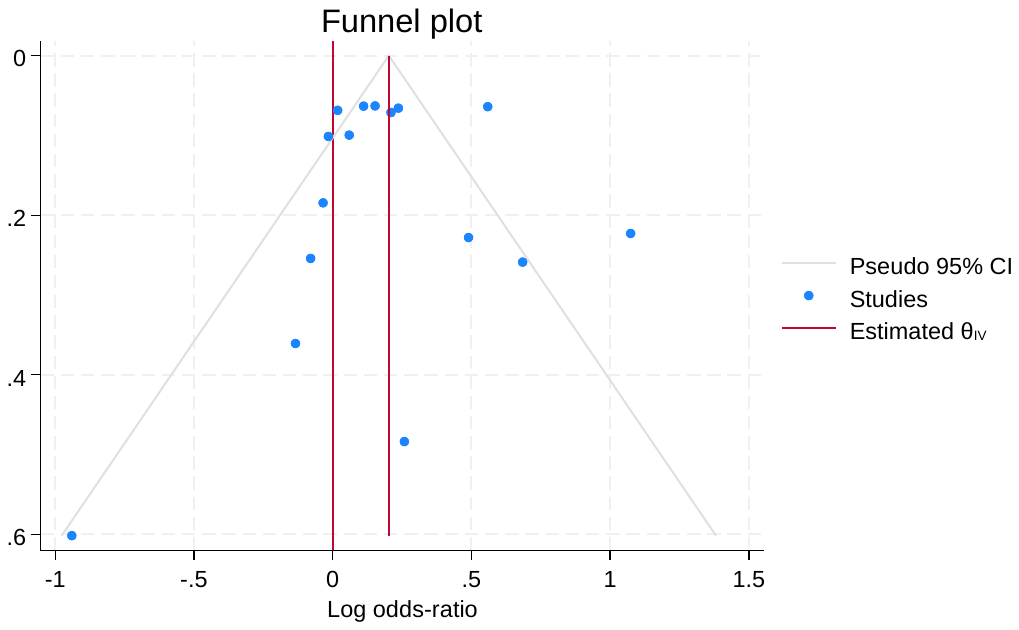

Figure S11. Overall Survival for Type of Drug Administered.

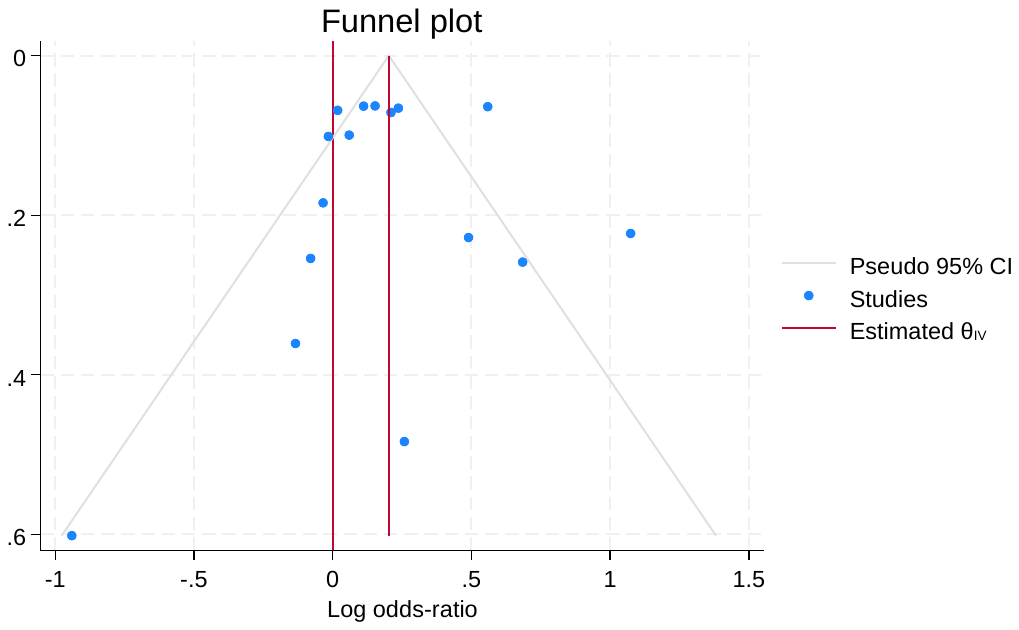

Figure S12. Fatal Stroke Figures Subgroup Analysis – Funnel Plot

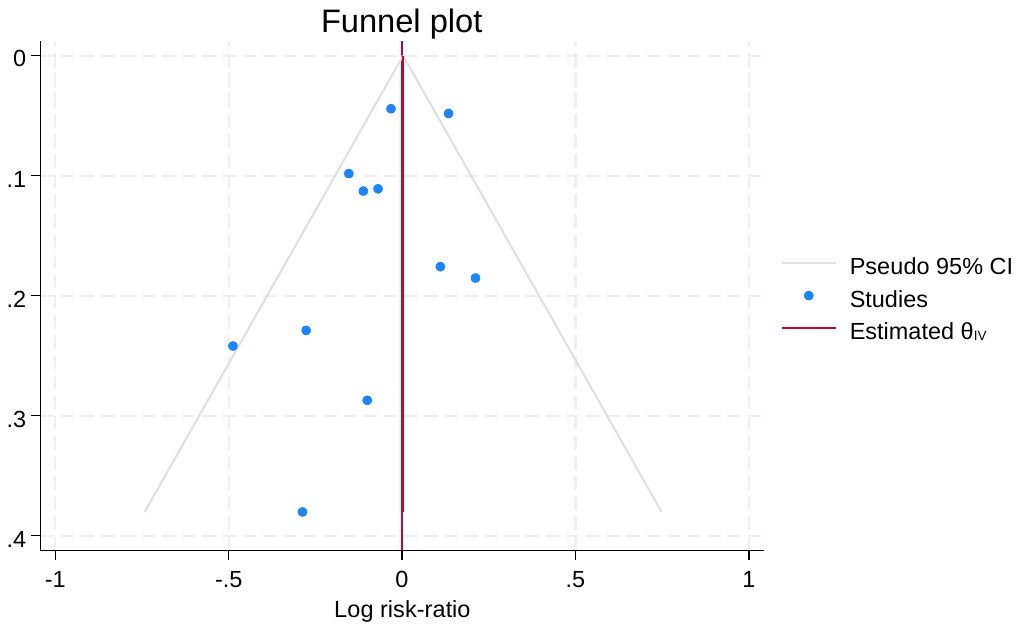

Figure S13. RACE Subgroup analysis for Fatal Stroke – Funnel Plot

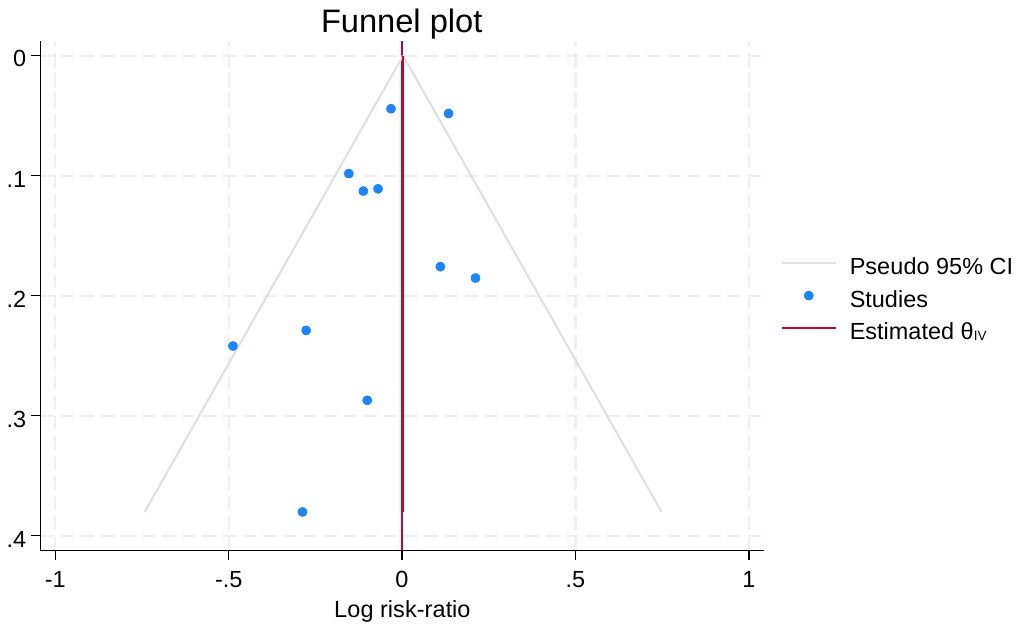

Figure S14. T2Dm vs Normal patients in Fatal Stroke – Funnel Plot

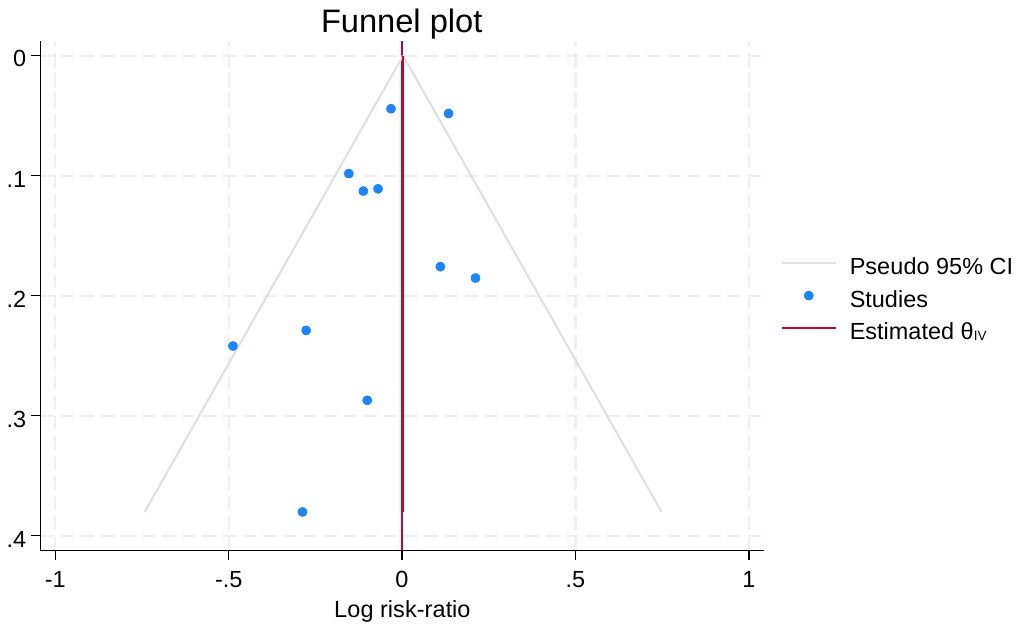

Figure S15. BMI patients in fatal stroke – funnel plot

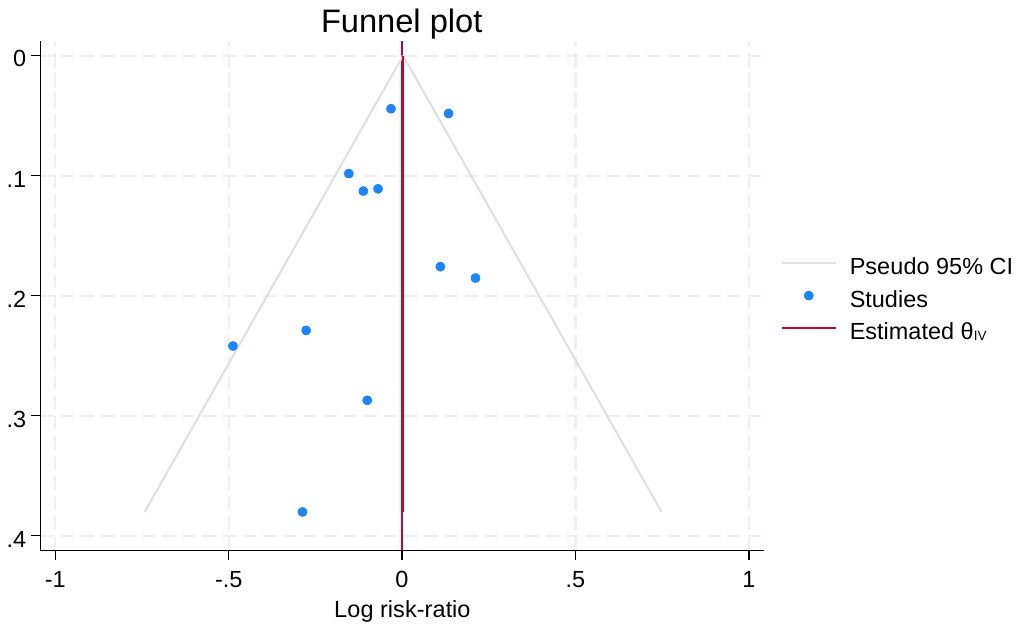

Figure S16. HBA1C Subgroup analysis in fatal stroke – funnel plot

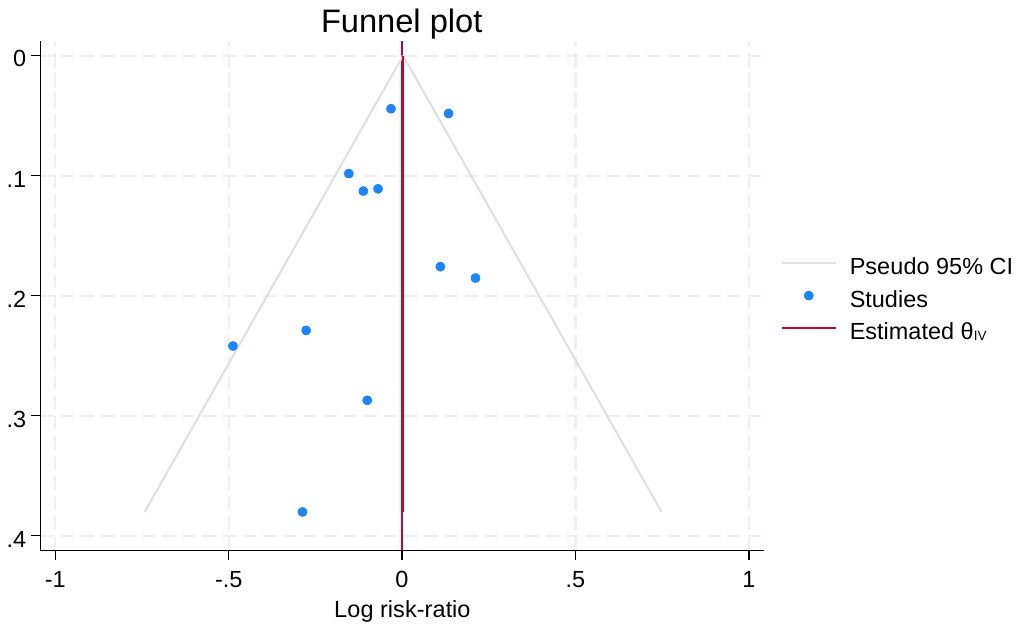

Figure S17. SGLT2 inhibitors Subgroup analysis in fatal stroke – funnel plot

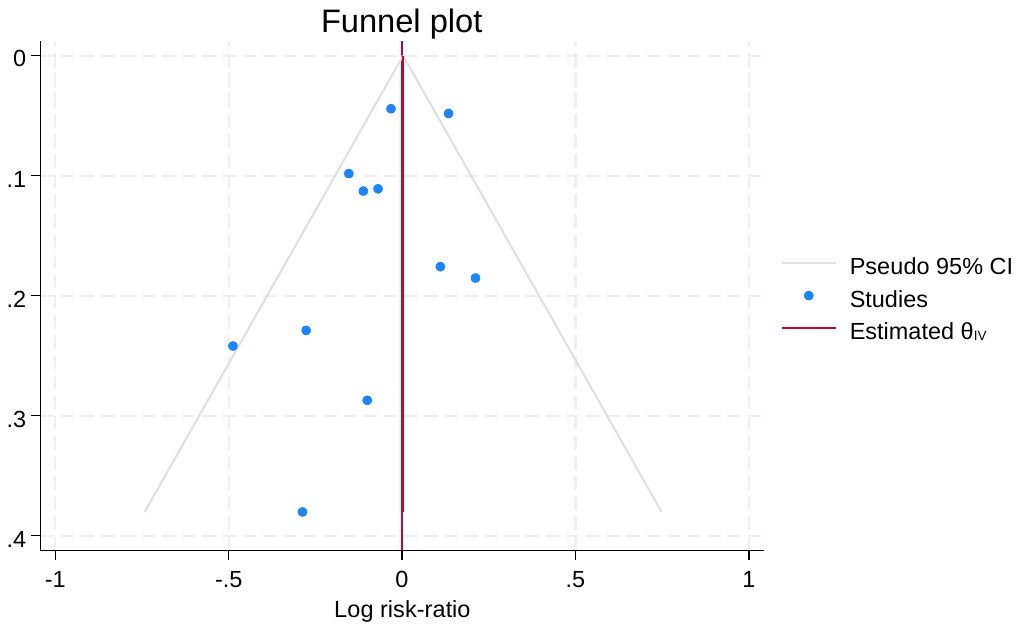

Figure S18. Daily Vs Weekly Subgroup analysis in fatal stroke – funnel plot

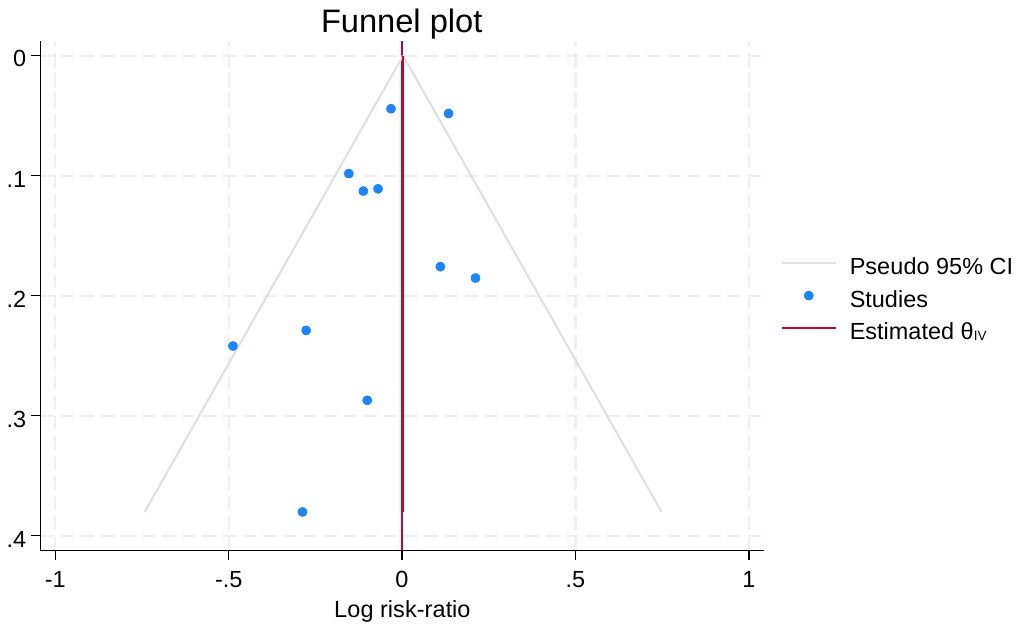

Figure S19. Oral vs Sub-cutaneous subgroup analysis in fatal stroke – funnel plot

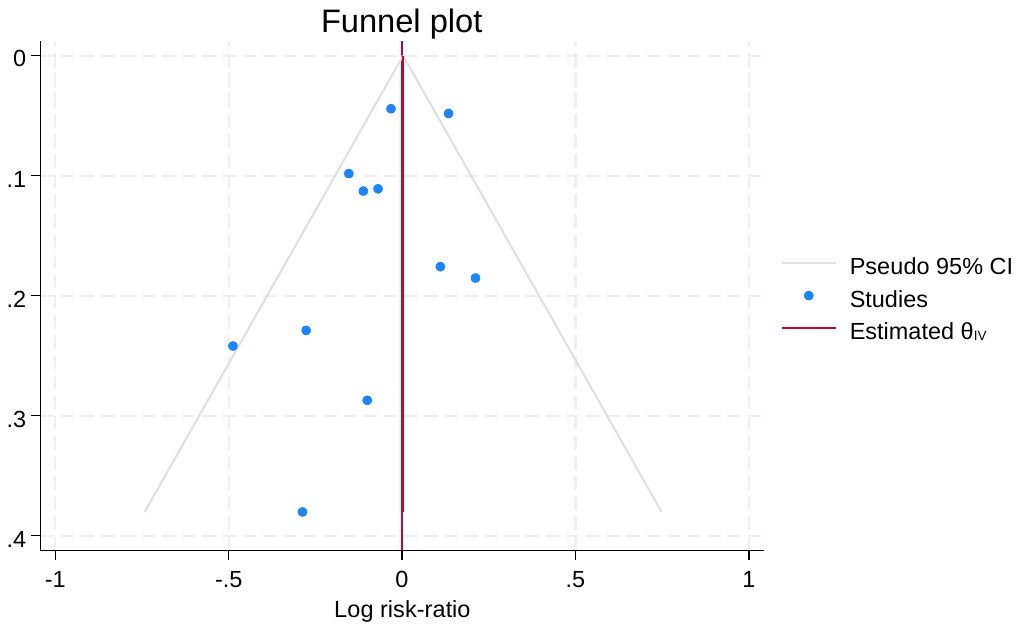

Figure S20. Fatal stroke in each medication subgroup analysis

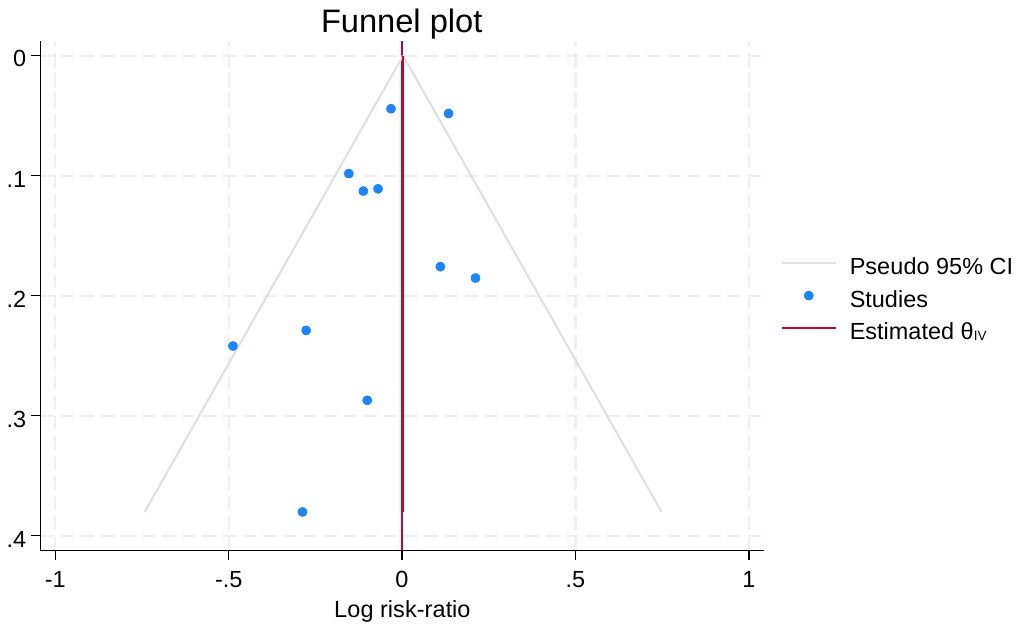

Figure S21. Non-Fatal Stroke in Age Group Overall Survival

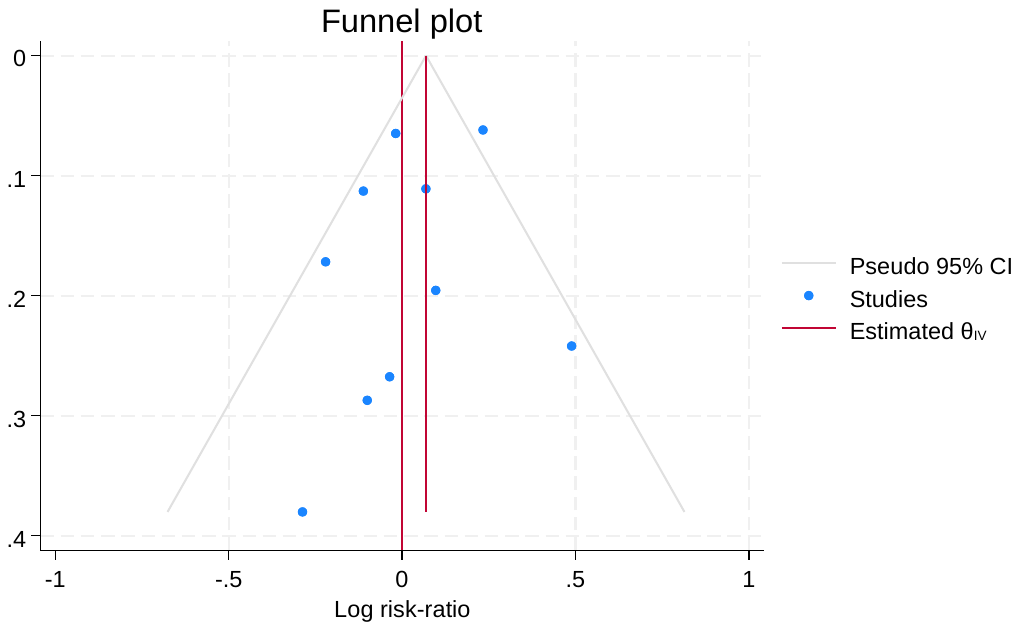

Figure S22. Non-Fatal Stroke in Race with overall survival

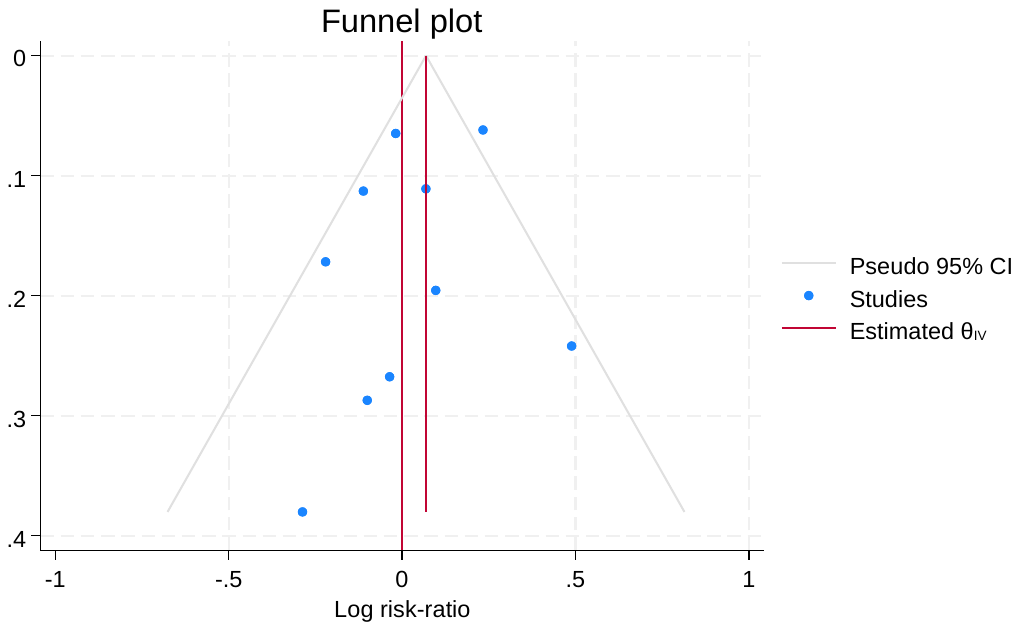

Figure S23. T2DM vs Normal Subgroup analysis for Non-Fatal Stroke

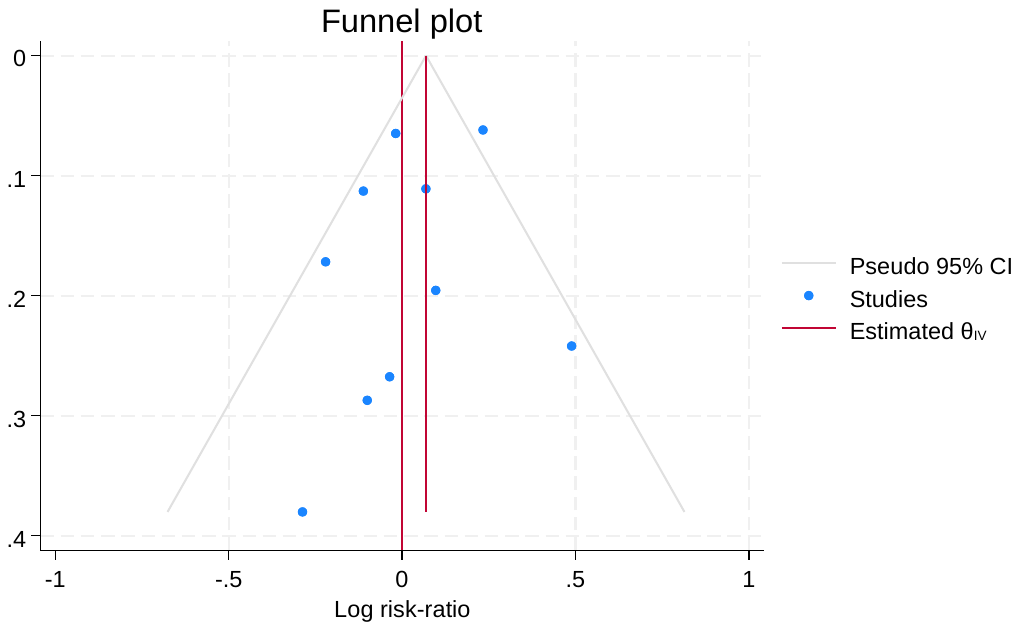

Figure S24. BMI Subgroup analysis for Non-Fatal Stroke

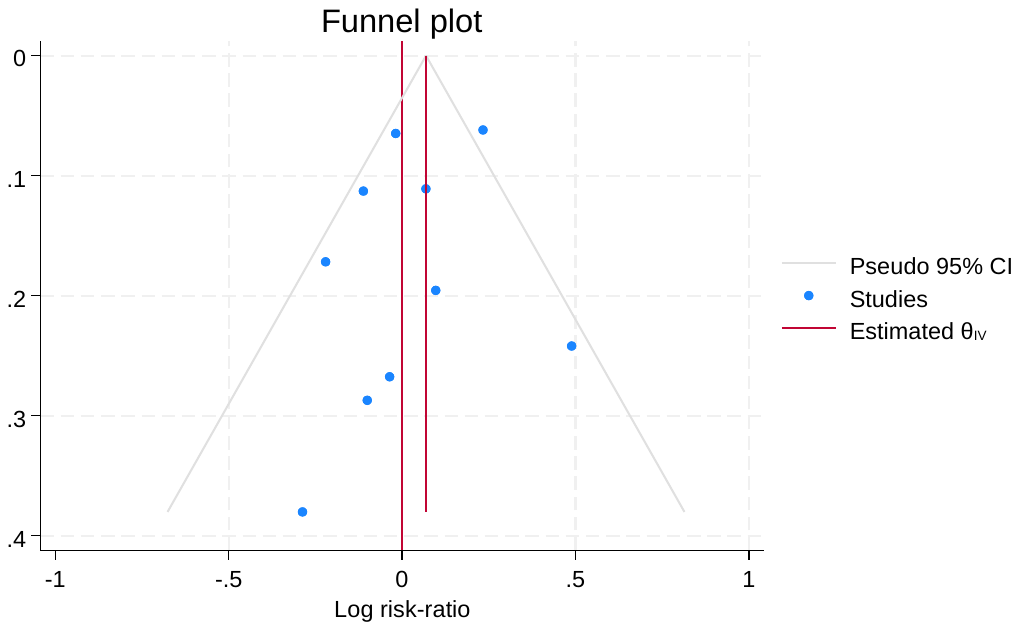

Figure S25. HBA1C Subgroup analysis for Non-Fatal Stroke

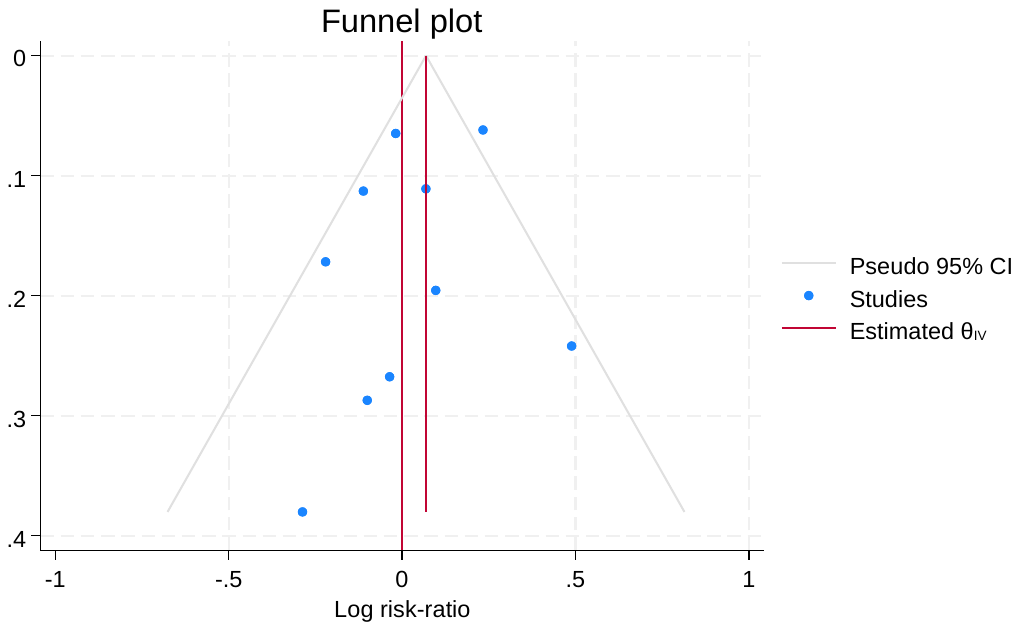

Figure S26. SGLT2 Administration Sub-Group analysis for Non-Fatal Stroke

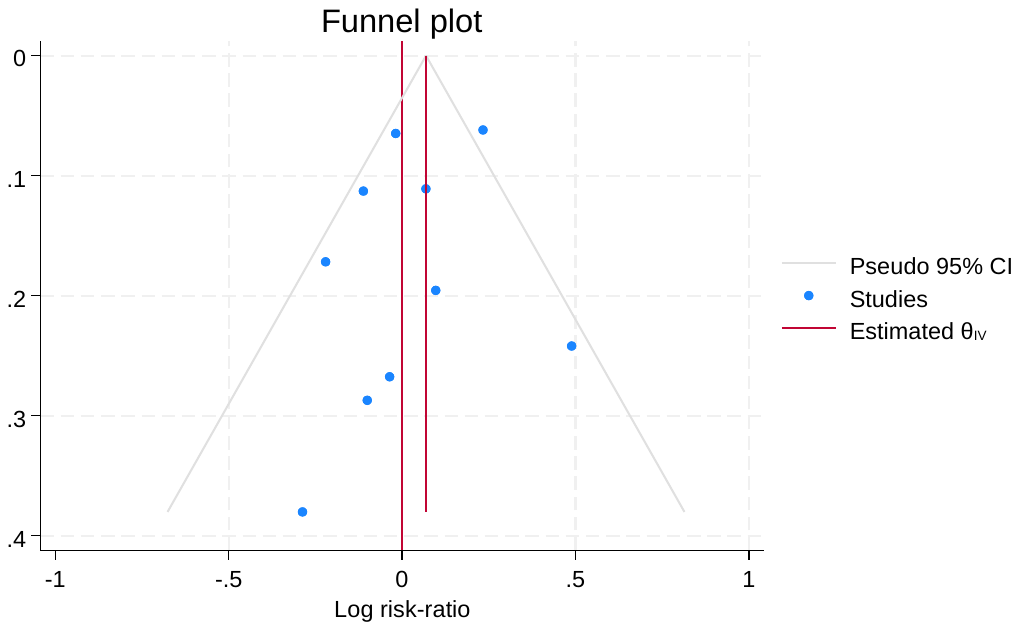

Figure S27. Weekly vs Daily Dosage Sub-group Analysis for Non-Fatal Stroke

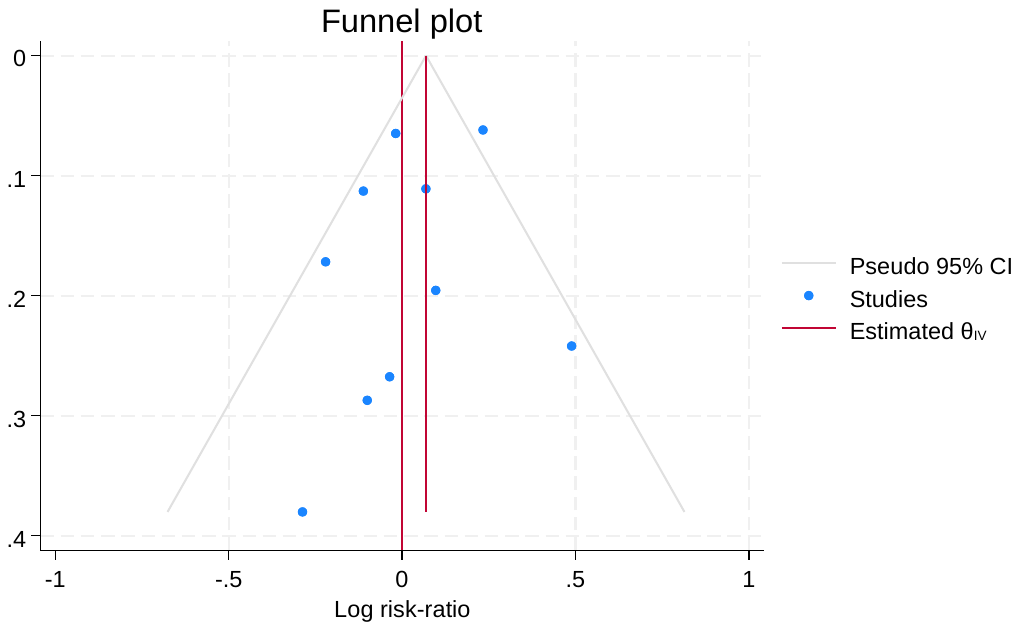

Figure S28. Oral vs Sub-cutaneous Sub-Group Analysis for Non-Fatal Stroke

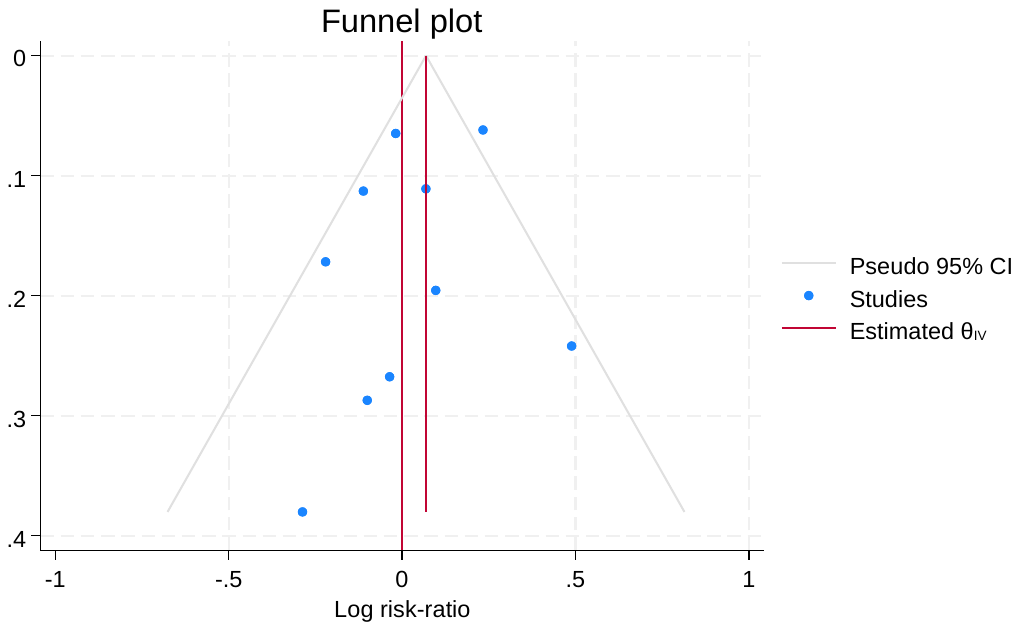

Figure S29. For each drug Subgroup analysis for Non-fatal Stroke

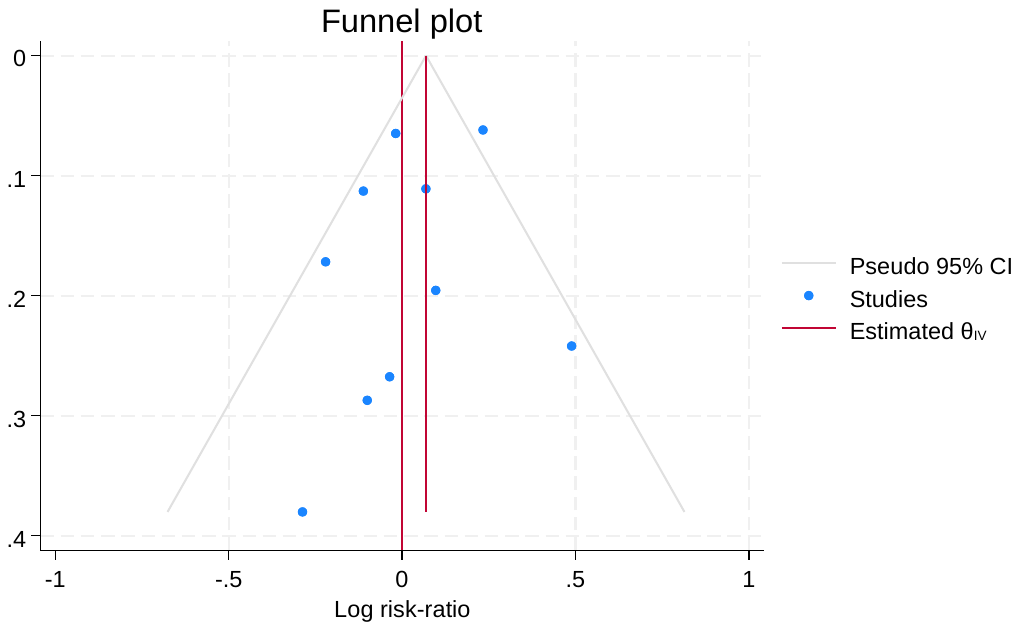

Figure S30. Death Due to fatal Stroke in age

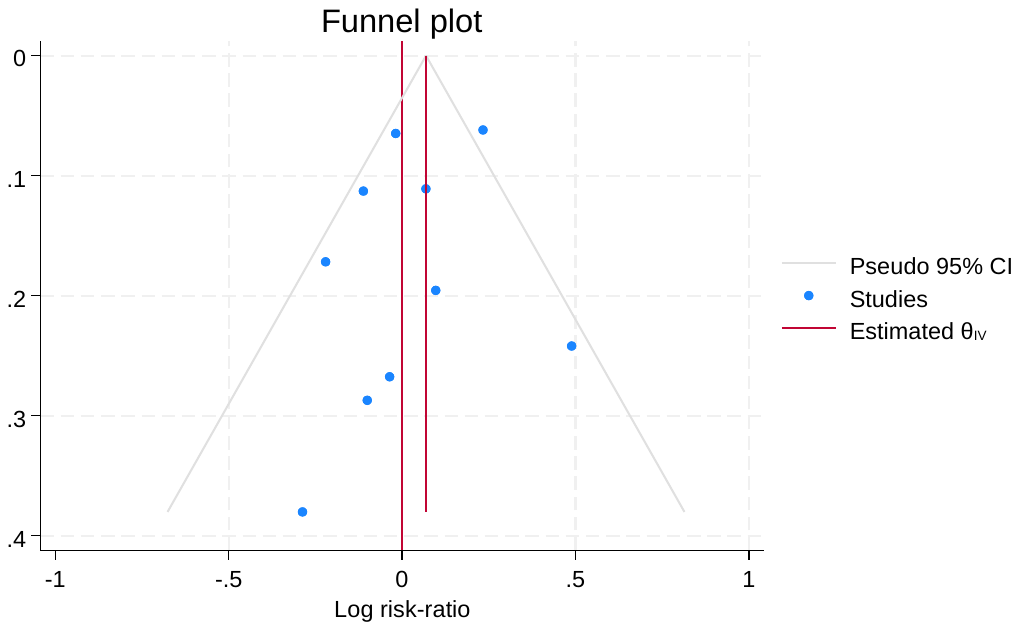

Figure S31. Death Subgroup Analysis in Race

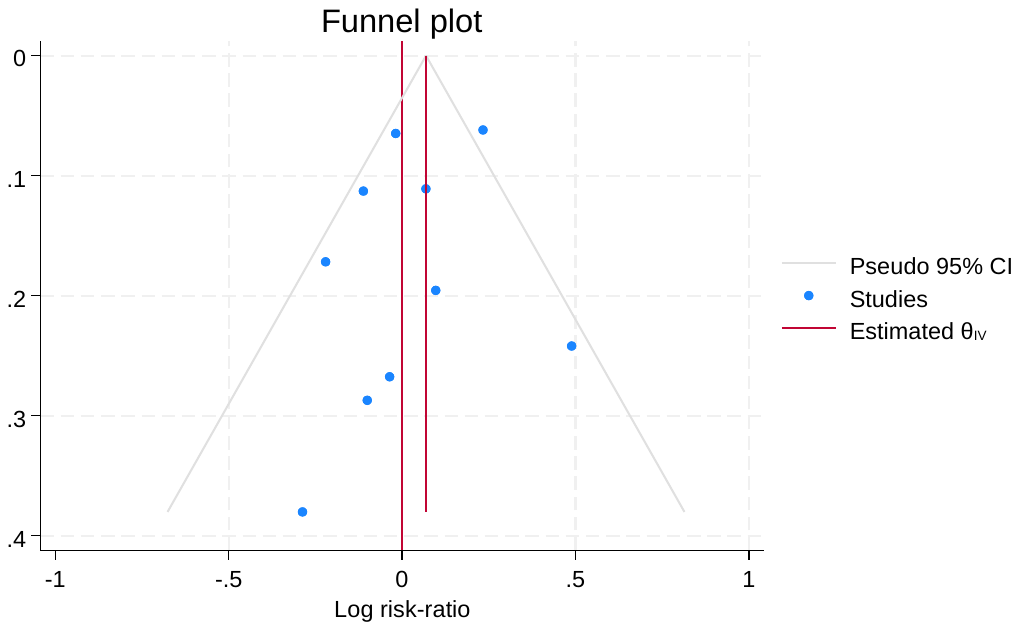

Figure S32. Death subgroup analysis in Normal vs T2DM

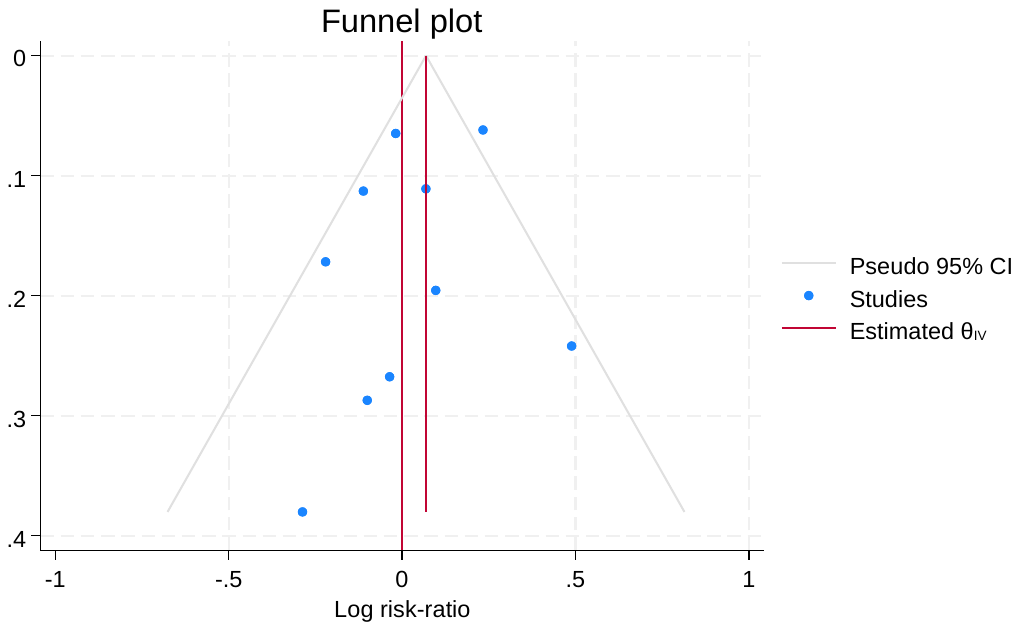

Figure S33. Death subgroup analysis in BMI

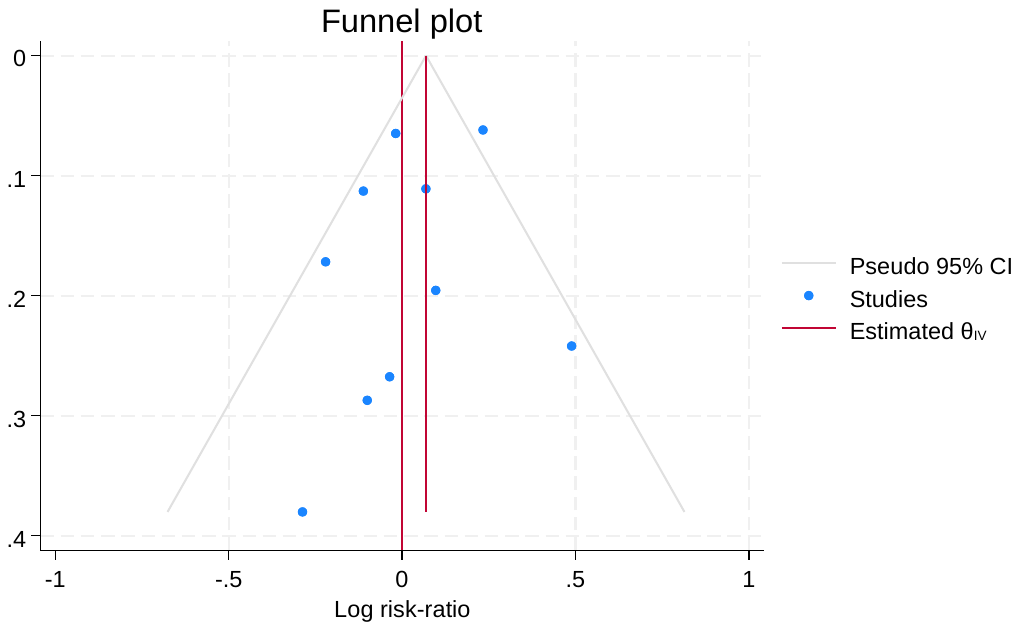

Figure S34. Death Subgroup analysis in HBA1C

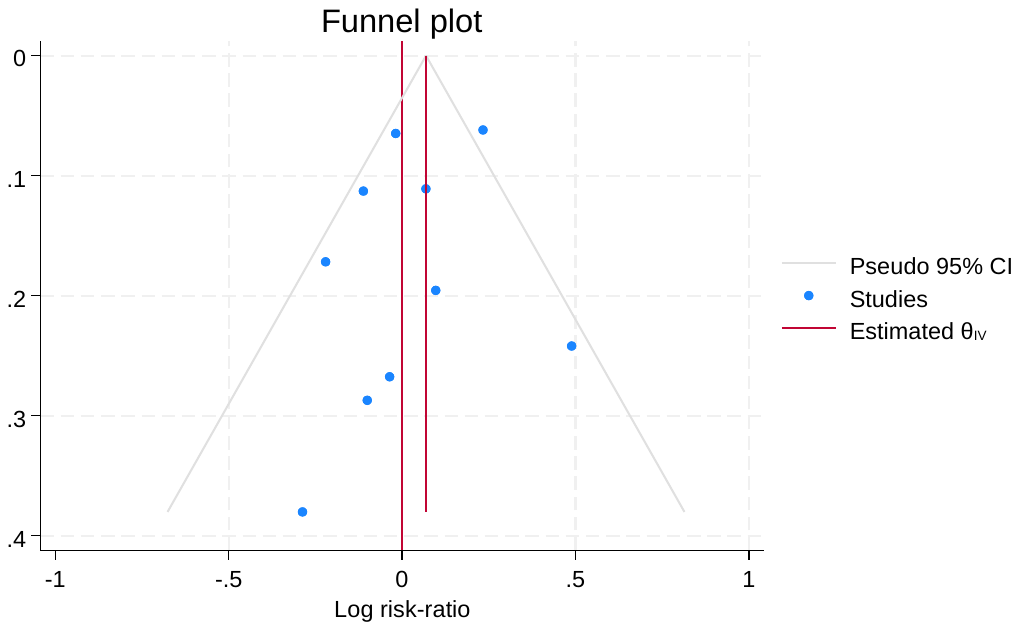

Figure S35. Death in SGLT2 Subgroup analysis.

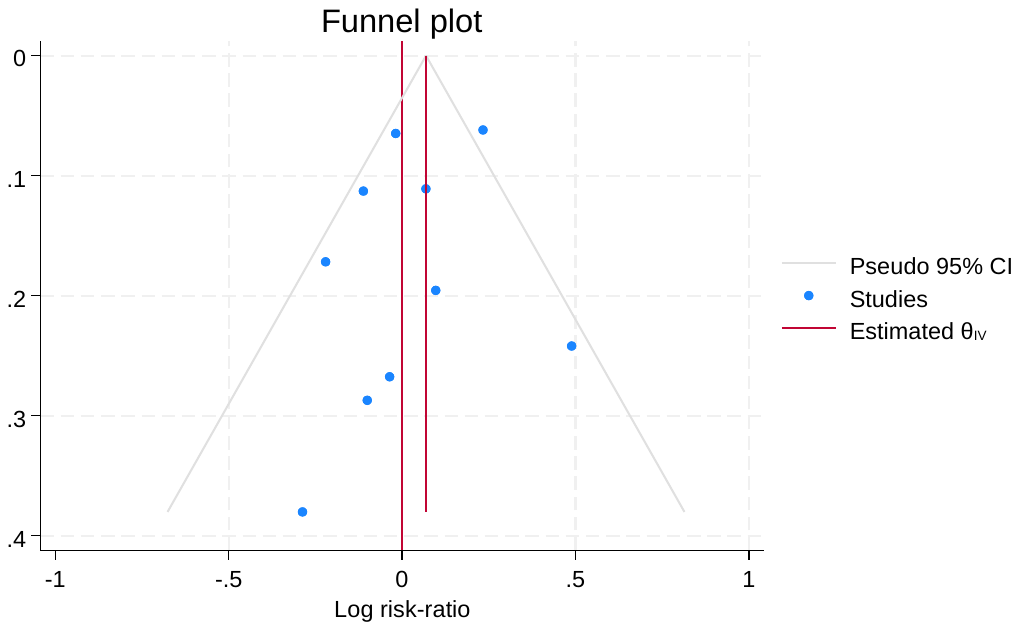

Figure S36. Death in Dosage Subgroup analysis

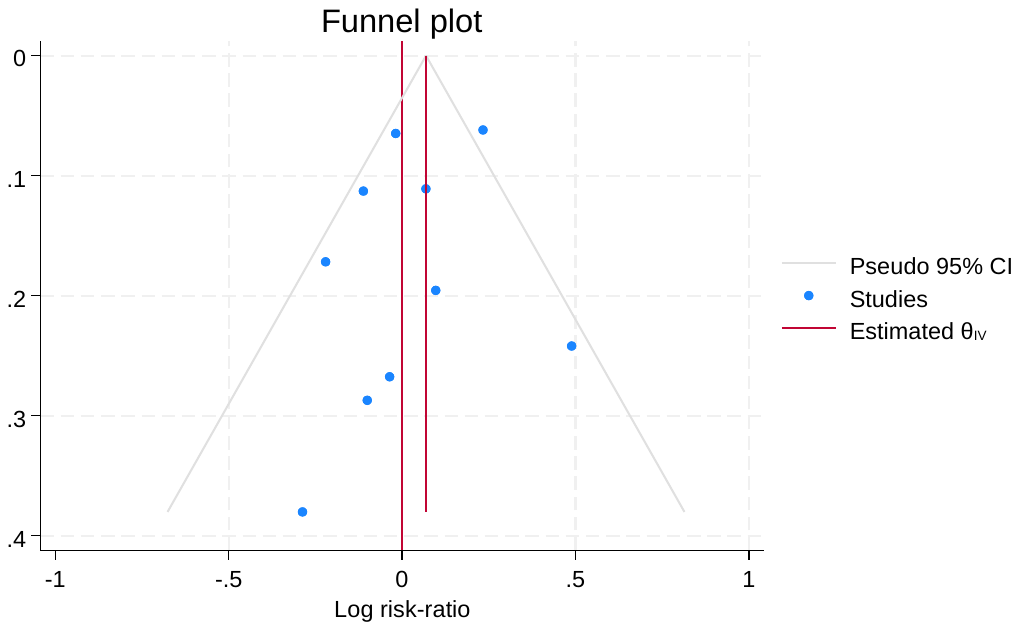

Figure S37. Death in Route of administration Sub Group Analysis

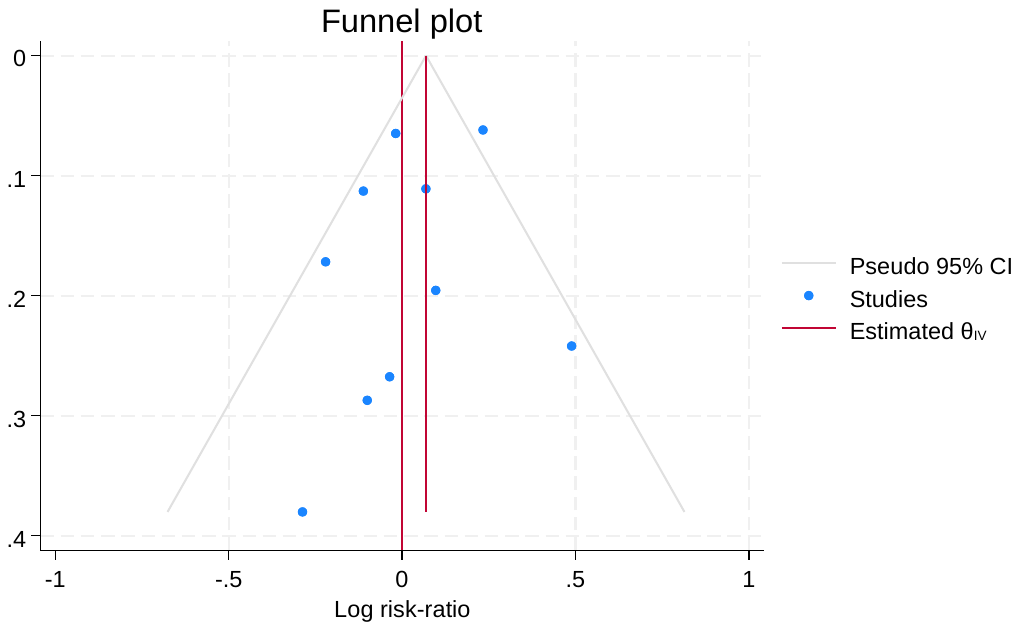

Figure S38. Death in Drug wise Subgroup Analysis

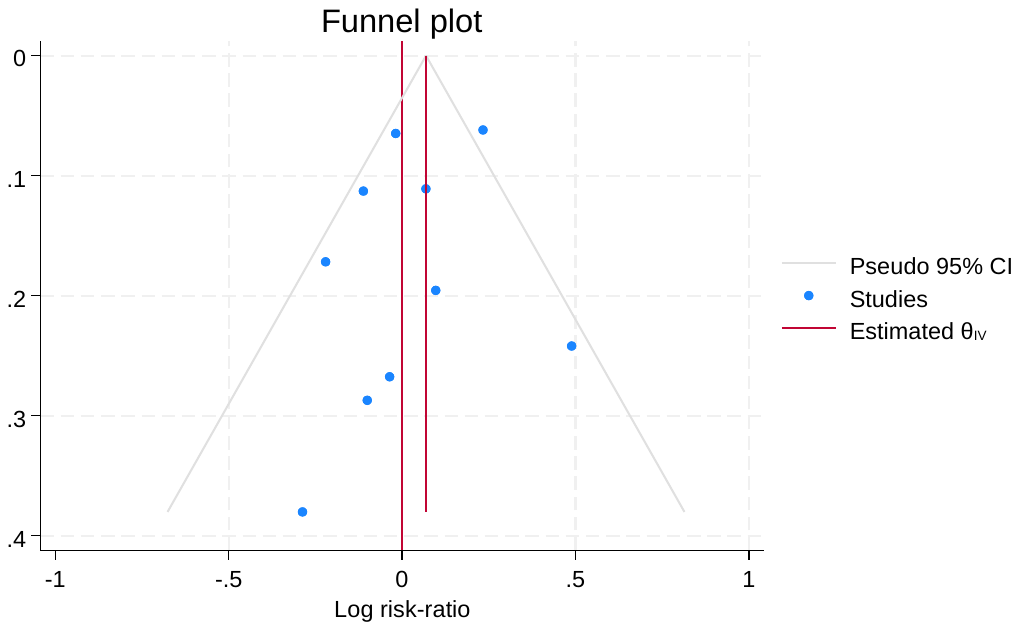

Table S3. Relative Treatment Effects in ranked order for all studies

|  | Placebo | Liraglutide | Semaglutide | Efpeglenatide | Exenatide | Tirzepatide |
| --- | --- | --- | --- | --- | --- | --- |
| Placebo | Placebo | -1.20 [-3.33; 0.93] | -1.26 [-2.51; 0.00] | -1.30 [-3.44; 0.84] | -2.00 [-4.14; 0.14] | -2.50 [-4.64; -0.36] |
| Liraglutide | -1.20 [-3.33; 0.93] | Liraglutide | . | . | . | . |
| Semaglutide | -1.26 [-2.51; 0.00] | -0.06 [-2.53; 2.42] | Semaglutide | . | . | . |
| Efpeglenatide | -1.30 [-3.44; 0.84] | -0.10 [-3.12; 2.92] | -0.04 [-2.52; 2.43] | Efpeglenatide | . | . |
| Exenatide | -2.00 [-4.14; 0.14] | -0.80 [-3.82; 2.22] | -0.74 [-3.22; 1.73] | -0.70 [-3.72; 2.32] | Exenatide | . |
| Tirzepatide | -2.50 [-4.64; -0.36] | -1.30 [-4.33; 1.73] | -1.24 [-3.73; 1.24] | -1.20 [-4.23; 1.83] | -0.50 [-3.53; 2.53] | Tirzepatide |

Table S4. Assessment of all inconsistence of studies for Renal Outcomes

| **Comparison** | **No.Studies** | **NMA** | **Direct** | **Indirect** | **Difference** | **Diff_95CI_lower** | **Diff_95CI_upper** | **pValue** |
| --- | --- | --- | --- | --- | --- | --- | --- | --- |
| Efpeglenatide:Exenatide | 0 | -0.70 | NA | -0.70 | NA | NA | NA | NA |
| Efpeglenatide:Liraglutide | 0 | 0.10 | NA | 0.10 | NA | NA | NA | NA |
| Efpeglenatide:Placebo | 1 | 1.30 | 1.30 | NA | NA | NA | NA | NA |
| Efpeglenatide:Semaglutide | 0 | 0.04 | NA | 0.04 | NA | NA | NA | NA |
| Efpeglenatide:Tirzepatide | 0 | -1.20 | NA | -1.20 | NA | NA | NA | NA |
| Exenatide:Liraglutide | 0 | 0.80 | NA | 0.80 | NA | NA | NA | NA |
| Exenatide:Placebo | 1 | 2.00 | 2.00 | NA | NA | NA | NA | NA |
| Exenatide:Semaglutide | 0 | 0.74 | NA | 0.74 | NA | NA | NA | NA |
| Exenatide:Tirzepatide | 0 | -0.50 | NA | -0.50 | NA | NA | NA | NA |
| Liraglutide:Placebo | 1 | 1.20 | 1.20 | NA | NA | NA | NA | NA |
| Liraglutide:Semaglutide | 0 | -0.06 | NA | -0.06 | NA | NA | NA | NA |
| Liraglutide:Tirzepatide | 0 | -1.30 | NA | -1.30 | NA | NA | NA | NA |
| Semaglutide:Placebo | 3 | 1.26 | 1.26 | NA | NA | NA | NA | NA |
| Tirzepatide:Placebo | 1 | 2.50 | 2.50 | NA | NA | NA | NA | NA |
| Semaglutide:Tirzepatide | 0 | -1.24 | NA | -1.24 | NA | NA | NA | NA |

Figure S39. SUCRA ranking of Renal Outcomes

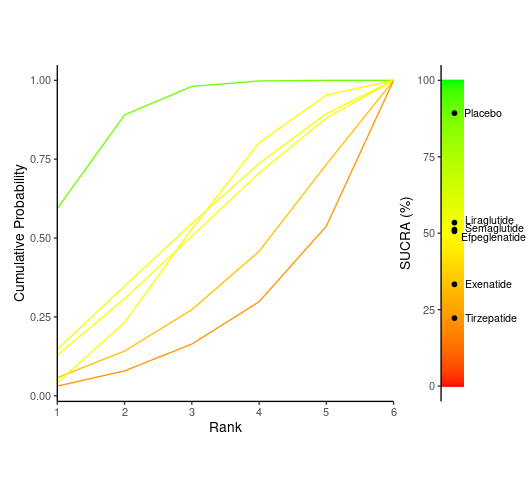

Figure S40. Risk of Bias ROBS 2.0

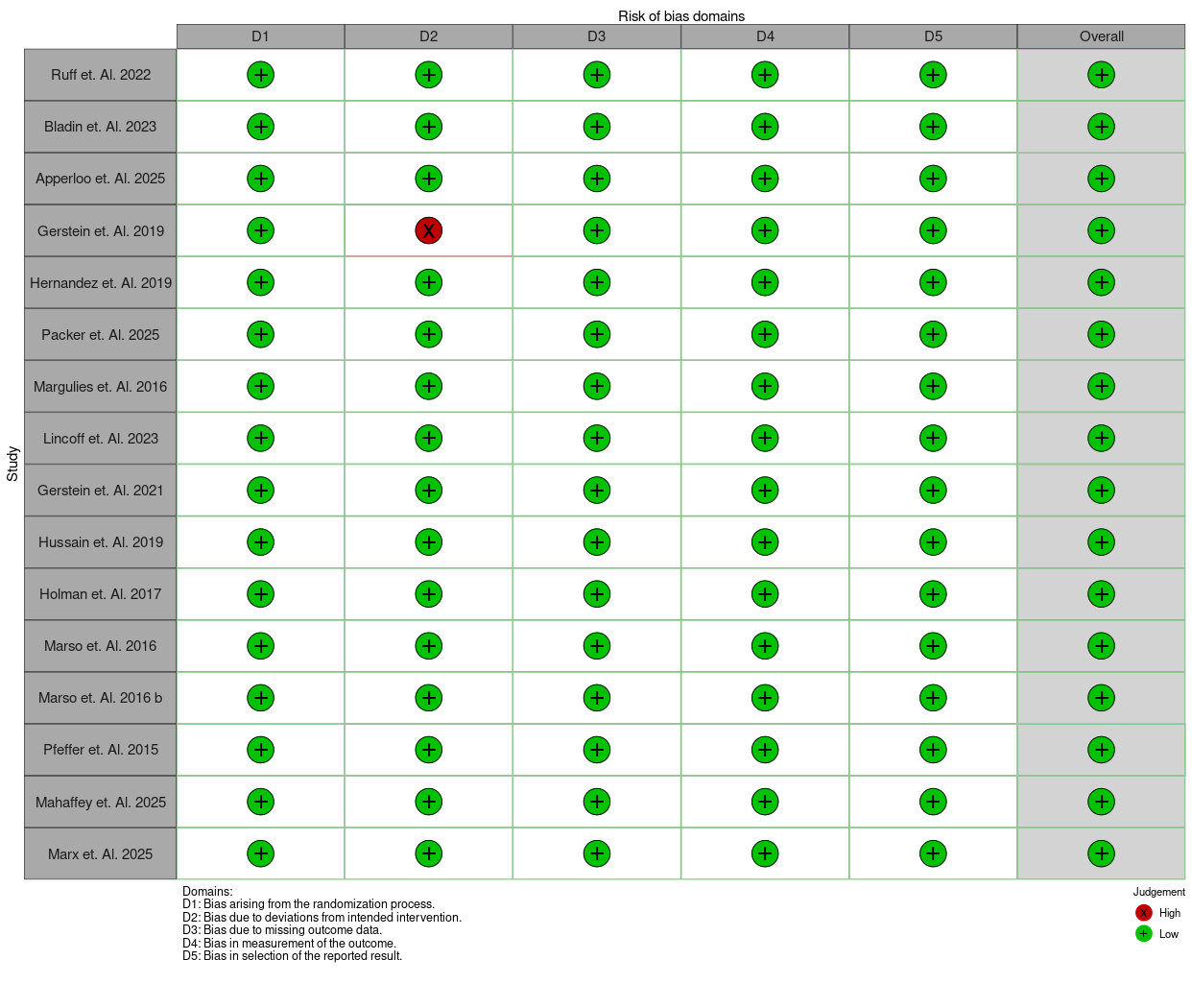
